# Integrating 730,947 exome sequences with clinical literature improves gene discovery

**DOI:** 10.64898/2026.03.23.26349081

**Authors:** Jeremy Guez, Julia K. Goodrich, Mikhail A. Moldovan, Katherine R. Chao, Prathitha Kar, Ruchit Panchal, Michael W. Wilson, Kristen M. Laricchia, Greg Rohlicek, Dmitry Biba, Daniel Marten, Qin He, Philip W. Darnowsky, Riley Grant, Ben Weisburd, Samantha M. Baxter, Joshua Nadeau, Wenhan Lu, Steve Jahl, Sophie Parsa, Abdallah Lamane, Stephanie DiTroia, Jack Fu, Xuefang Zhao, Elissa Alarmani, Charlotte Tolonen, Sam Novod, Sam Bryant, Christine Stevens, Sinéad B. Chapman, Caroline Cusick, Christopher Vittal, Laura D. Gauthier, Jacqueline I. Goldstein, Daniel Goldstein, Daniel King, Timothy Poterba, Grace Tiao, gnomAD Project Consortium, Matteo Tranchero, William Lotter, Daniel G. MacArthur, Harrison Brand, Vladimir Seplyarskiy, Evan Koch, Michael E. Talkowski, Matthew Solomonson, Benjamin M. Neale, Anne O’Donnell-Luria, Hilary K. Finucane, Shamil R. Sunyaev, Mark J. Daly, Heidi L. Rehm, Kaitlin E. Samocha, Konrad J. Karczewski

## Abstract

Accurate estimates of allele frequencies aid in genetic discovery, including rare disease diagnosis, common disease investigations, and population genetics. Here, we present the Genome Aggregation Database version 4 (gnomAD v4), including 730,947 with exome sequences, a fivefold increase over previous releases. We demonstrate that statistical power to detect strong selective constraint continues to increase with sample size. We develop a new loss-of-function annotation pipeline, which learns genomic features predictive of nonsense-mediated decay and splicing effects from selection signals, achieving 90% precision for distinguishing likely true versus false positive loss-of-function variants. This improved pipeline, along with incorporation of highly deleterious missense variants into measures of loss-of-function intolerance, improves disease gene detection, particularly for short genes and those with gain-of-function mechanisms. To improve disease gene prediction, we systematically extract gene–disease associations from biomedical literature, map these to gene-level biological features, and integrate both with refined constraint metrics within a Bayesian framework, yielding state-of-the-art prediction of gene–disease relevance. We highlight genes under strong constraint but with limited clinical characterization, which are enriched in embryonic lethal and fertility phenotypes, thus prioritizing previously under-characterized disease genes. Together, these advances establish a unified framework for accelerating gene discovery and improving rare disease diagnosis.

## Introduction

Population sequencing resources^1–6^ have become essential for rare disease diagnosis and gene discovery across rare and common diseases. In particular, observation of an allele above a certain frequency in a large sample (e.g., 0.01%) is an exclusionary criterion for severe disease-causing dominant alleles, and comprehensive characterization of such alleles across multiple ancestry groups is necessary for accurate diagnostic pipelines. In addition, quantifying depletion of gene-disrupting variation (e.g., predicted loss-of-function [pLoF] variants) in large datasets yields constraint metrics^1,6^ that have proven valuable for prioritizing candidate disease genes^7–9^ and refining clinical variant classification^10,11^. Despite these advances, nearly half of rare disease patients remain without a molecular diagnosis after genetic testing^12–14^, and thousands of Mendelian conditions are estimated to remain genetically uncharacterized^15^, due in part to undiscovered gene–disease associations.

Accurate characterization of variant effects improves our understanding of disease etiology, including loss-of-function (LoF)-based haploinsufficiency or gain-of-function (GoF) effects. For pLoF variants, we previously developed LOFTEE (Loss-Of-Function Transcript Effect Estimator)^1^ to predict which variants trigger nonsense-mediated decay (NMD), which proved instrumental for predicting variants causing loss-of-function, although false positives revealed by manual curation remain^16^. Furthermore, new annotation methods^17–19^ prioritize functional missense variants, which can include both LoF and GoF effects, but distinguishing between these effects, and determining appropriate thresholds across applications, remains a challenge. Improved variant-level prediction and annotation of pLoF and missense variants will provide deep insights into disease pathogenesis.

Beyond characterizing observed and unobserved variation, molecular diagnostics rely on existing clinical knowledge, including published literature and ClinVar. However, as of 2021, 65% of genes in the most constrained LOEUF decile lacked any OMIM disease association^20^, indicating that a large fraction of genes under strong purifying selection have no established clinical relevance. One plausible explanation is that loss-of-function variants in many of these genes cause prenatal or early embryonic lethality that escapes clinical ascertainment: while roughly 35–39% of mouse genes are essential for development^21,22^, only 3% of human protein-coding genes are currently associated with prenatal or infantile lethality^21^. This disparity suggests a large number of potentially lethal genes whose effects on human development remain uncharacterized.

Here, we present quality control and analysis of 730,947 exomes from the Genome Aggregation Database (gnomAD v4), together with a systematic gene discovery framework integrating evolutionary constraint and clinical knowledge. This dataset provides a near-complete catalog of variants with allele frequencies >0.01% across five genetic ancestry groups, enabling improved variant interpretation in clinical diagnostics. Using this resource, we refine pLoF annotation and develop a constraint framework with increased power to capture both LoF and GoF signals. We further introduce a gene-level clinical significance score derived from agentic literature curation. We highlight genes under strong constraint but limited clinical characterization, which are enriched for embryonic lethal and fertility phenotypes, consistent with their under-ascertainment in current disease gene catalogs. Together, these advances establish a framework for combining evolutionary constraint with clinical knowledge to accelerate gene discovery.

## Results

### Overview of the gnomAD v4 dataset

We aggregated exome or genome sequencing data from 1,108,389 individuals, including the previous exome and genome releases (gnomAD v2 and v3, respectively), UK Biobank, and additional large cohorts. As the vast majority of genome samples included in gnomAD v4 were previously released in the v3 dataset^2^, we focus here on the exome sequenced samples. All samples were uniformly processed and jointly called using a harmonized pipeline aligned to GRCh38 and analyzed using Hail’s VariantDataset (VDS) representation^23^. We applied stringent sample and variant quality control, removing low-quality samples, close relatives, individuals lacking appropriate consent for aggregate data release, and cohorts ascertained for severe pediatric disease. We inferred genetic ancestry using principal component analysis and a supervised classifier trained on reference panels and additional labeled samples representing diverse ancestries, adopting the ancestry group labels defined in those datasets.^5,24,25^ As we use a blend of reference panels including our own, we recommend a nomenclature of gnomAD-AFR-like to describe individuals with genetic similarity to the AFR genetic ancestry group in this manuscript, in accordance with recommendations^26^. This process, detailed in Supplementary Material, yields a high-quality set of 730,947 unrelated individuals with exome sequence data spanning multiple genetic ancestry groups, representing an approximately five-fold size increase over previous releases.

We discover 19,940,035 high-quality coding variants, the majority of which are rare: 96.5% (5,307,409/5,502,337) of synonymous, 97.8% (12,568,444/12,853,521) of missense, and 98.9% (1,566,943/1,584,177) of pLoF variants fall below an allele frequency (AF) of 0.01% (Extended Data Fig. 1a). At the individual level, however, most variants are common: a person carries on average ∼13,507 synonymous, ∼11,289 missense, and ∼204 pLoF variants, of which only ∼51, ∼98, and ∼7, respectively, have AF < 0.01% (Extended Data Fig. 1b).

The growth of the gnomAD dataset from v2 to v4 substantially increases the catalog of common variants (Fig. 1a), which are critical for filtering in diagnostic pipelines. The increase in observed missense variants from gnomAD v2 to v4 across genetic ancestry groups closely matches the expectation from a Poisson sampling model, confirming that variant accumulation between releases is driven by increased sample size.

**Figure 1.**
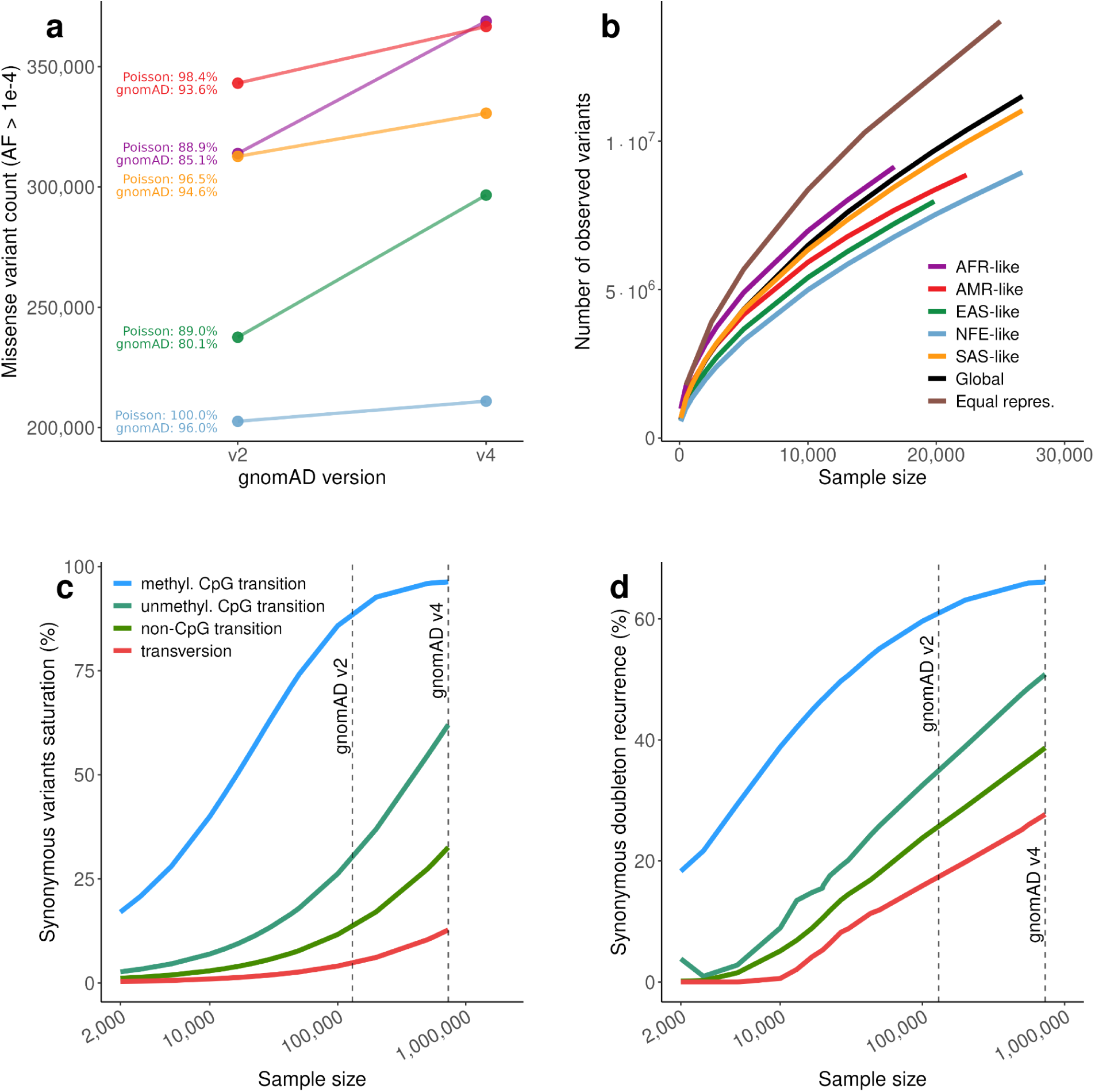
Observed variation, saturation, and mutational recurrence in gnomAD v4. **a**, Observed missense variant counts at allele frequency > 10⁻⁴ in gnomAD v2 and v4 across genetic ancestry groups. Annotations indicate the expected ratio of v2 to v4 counts under a Poisson model and the observed ratio in the data. **b**, Number of observed variants as a function of sample size across genetic ancestry groups (AFR = African, AMR = Admixed American, EAS = East Asian, NFE = Non-Finnish European, SAS = South Asian), globally, and under equal representation across genetic ancestry groups. **c**, Fraction of all possible synonymous variants that are polymorphic in gnomAD v4, by mutational class as a function of sample size. Dashed vertical lines indicate the sample sizes of gnomAD v2 and v4. **d**, Estimated fraction of recurrent doubletons by mutational class as a function of sample size. Dashed vertical lines indicate gnomAD v2 and v4 sample sizes. Colors are consistent in c and d.

Consistent with previous work, we observe that the accumulation of variants with increasing sample size is dependent on genetic ancestry. Individuals of AFR-like genetic ancestry show a larger number of detected variants than other genetic ancestry groups (Fig. 1b), consistent with demographic modeling of the out-of-Africa bottleneck^27,28^. However, an equitable sampling strategy across genetic ancestries identifies more variants overall than AFR-like-only sampling: at a sample size of 10,000 individuals, equal genetic ancestry representation yields 8,360,443 variants compared to 6,973,288 for AFR-like alone (Fig. 1b), highlighting how incorporating individuals across multiple inferred genetic ancestries can increase the number of variants identified.

### Mutational saturation and recurrence reshape variant discovery and constraint estimates

Given the substantial increase in sample size in gnomAD v4, we next quantified mutational saturation associated with this expansion, defined as the observation of all possible variants within a given category. We observe near-complete saturation for synonymous variants with high mutation rates. Specifically, 96.3% of all possible methyl-CpG transition variants are observed, compared with 62.0% of unmethyl-CpG transitions, 32.6% of non-CpG transitions, and 12.8% of transversions (Fig. 1c).

This high level of saturation is accompanied by extensive mutational recurrence in gnomAD v4, defined as the presence of variants arising from multiple independent mutational events. Recurrence alters the shape of the site frequency spectrum (SFS), which describes the distribution of variant frequencies in the sample. For methyl-CpG sites, the SFS shows an unusual pattern, with more variants observed in ten samples than as singletons, a hallmark of strong recurrence (Extended Data Fig. 2a). We estimate the proportion of recurrent mutations among doubletons for each mutational category via an infinite sites model (see Supplementary Material). This approach quantifies recurrence and reveals that, while for synonymous variants, recurrence is particularly strong at highly mutable sites (66.11% for methyl-CpG transitions), it is also present at low mutation rate sites, including transversions (27.7%) (Fig. 1d). Missense and predicted loss-of-function (pLoF) variants show lower levels of saturation and recurrence, consistent with their deleterious effects on fitness and the action of purifying selection (Extended Data Fig. 2b-d).

Given the central role of constraint metrics in clinical variant interpretation and gene prioritization, we next examined the impact of variant saturation on LOEUF (loss-of-function observed/expected upper bound fraction), a measure of purifying selection acting on a gene. LOEUF is defined as the upper bound of the ratio of observed pLoF variants to those expected under neutrality, with smaller values indicating stronger selection. We show analytically that while LOEUF measures selection, it is also affected by sample size and mutation rate (Supplementary Material). As sample size or mutation rate increases, saturation causes observed variants to approach expected numbers, increasing the observed/expected ratio and, consequently, LOEUF (Extended Data Fig. 3a,b). The absolute increases are strongest for genes under weak selection, while genes under no selection remain stable near 1 (Fig. 2a); however, relative increases are largest for strongly constrained genes (Extended Data Fig. 3d).

**Figure 2.**
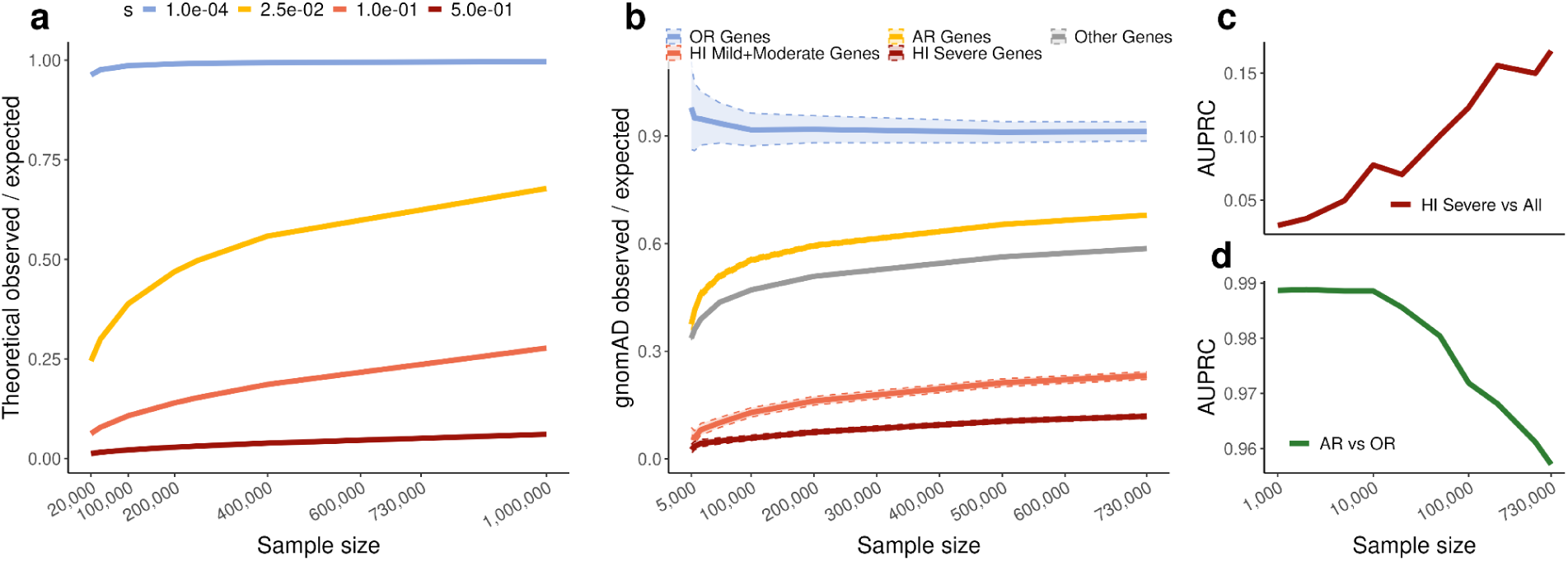
Impact of variant saturation on values and performance of LOEUF. **a**, Analytical predictions for the observed/expected ratio as a function of sample size under different strengths of purifying selection (selection coefficient *s*). **b**, Empirical observed/expected ratios as a function of sample size for different gene sets, showing qualitative agreement with the analytical predictions. Haploinsufficient (HI) genes associated with severe phenotypes represent strong selection, neurodevelopmental disorder-associated (NDD) genes intermediate selection, genes with autosomal recessive inheritance (AR) weak heterozygous selection, and olfactory receptor (OR) genes no selection. Shaded areas indicate upper and lower bound at 0.05. **c**, Area under the precision–recall curve (AUPRC) for distinguishing HI genes associated with severe phenotypes from all other genes as a function of sample size. **d**, AUPRC for distinguishing AR genes from OR genes as a function of sample size.

These patterns are supported by empirical data across gene classes spanning a range of selection regimes, including haploinsufficient (HI) genes associated with severe phenotypes (strong selection), HI genes associated with moderate and mild phenotypes (intermediate selection), autosomal recessive genes (AR; weak heterozygous selection), and olfactory receptor genes (OR; neutral) (Fig. 2b). We also observe the predicted effect of mutation rate on LOEUF in empirical data, with genes with high overall mutation rate showing higher LOEUF values on average, after controlling for expected and possible number of variants (Extended Data Fig. 3c).

These dynamics have important consequences for gene prioritization. As sample size increases in gnomAD v4, discrimination between genes under strong and weak selection increases, improving power to detect strongly constrained genes. Consistent with this, the area under the precision–recall curve (AUPRC) for distinguishing HI genes associated with severe phenotypes^1^ from other genes continues to increase with sample size (Fig. 2c). In contrast, saturation reduces discrimination between genes under weak selection and no selection, decreasing power to distinguish these categories and causing a slight performance decline for the AR versus OR benchmark at large sample sizes (Fig. 2d). More comprehensive mathematical analysis further characterizes interactions between selection coefficient, sample size, and mutation rate on LOEUF power (Extended Data Fig. 4a-c, Supplementary Material). We show that power remains to be gained from larger sample sizes, with gains continuing through 10 million samples for genes under strong purifying selection—the class most relevant to rare human disease (Extended Data Fig. 4b-c).

### Improving loss-of-function variant annotation using population genetics and NMD-informed features

To improve pLoF variant interpretation, and LOEUF by extension, it is critical to define a confident set of pLoF variants that trigger nonsense-mediated decay (NMD). LOFTEE previously classified stop-gained, splice donor, and splice acceptor variants into High- and Low-Confidence categories, but manual curation revealed substantial room for improvement¹⁵. In genes under strong selection, true LoF variants can be distinguished from false annotations by allele frequency, as they are expected to segregate at lower frequencies than misannotated variants^29,30^. This enables estimation, for each pLoF variant, of the probability that it does not belong to the distribution under selection^30^, which we term the probability of neutrality (p_neutral_).

However, this approach is limited to genes under strong purifying selection. We therefore developed a framework that infers p_neutral_ and leverages genomic properties of pLoF variants with low p_neutral_ to generalize this signal genome-wide.

We first constructed a Bayesian mixture model to estimate p_neutral_ for each variant. For each gene, pLoF variant frequencies are assumed to be drawn from one of two distributions: a distribution under selection^31^ and a neutral distribution learned from exome-wide synonymous variants (Supplementary Material). Given known mutation rates, this model jointly infers the heterozygous selection coefficient (s_het_) and p_neutral_. High p_neutral_ typically results from high-frequency variants in strongly constrained genes, which functionally reflects their propensity to trigger nonsense-mediated decay (NMD). For example, in *CACNA1A*, a gene implicated in severe neurological disorders, variants observed in ∼10 individuals show p_neutral_ near 1 (Fig. 3a) and are therefore unlikely to cause loss-of-function. Within this gene (Fig. 3b) and genome-wide (Extended Data Fig. 5a), p_neutral_ follows a bimodal distribution, with 0.52% of variants exceeding 80% genome-wide.

**Figure 3.**
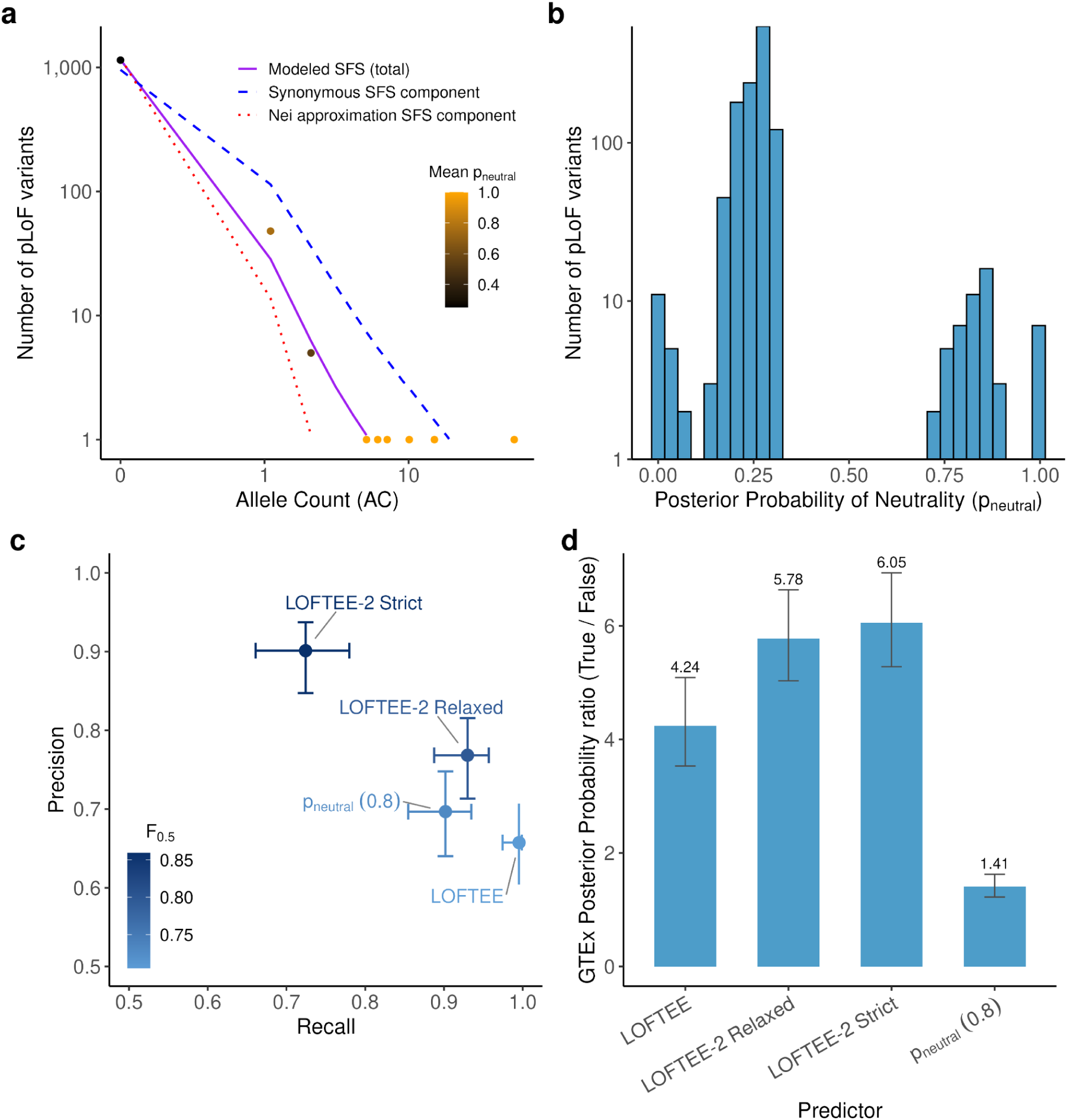
Estimation of loss-of-function misannotation and benchmarking of LOFTEE-2. **a,** Site frequency spectrum (SFS) of pLoF variants in a representative strongly constrained gene (*CACNA1A*), modeled as a mixture of a distribution under selection (red dotted line; Nei approximation of site frequency spectrum under selection) and a neutral distribution learned from synonymous variants (blue dashed line). The combined fitted model is shown in purple. Points represent observed pLoF variants, colored by their mean posterior probability of neutrality (p_neutral_). **b,** Distribution of p_neutral_ across all pLoF variants in *CACNA1A*, showing a bimodal pattern separating likely true loss-of-function (left) variants from likely neutral variants (right). **c,** Performance of LOFTEE v1, LOFTEE-2 relaxed, LOFTEE-2 strict, and a p_neutral_-based filter (threshold = 0.8) evaluated against a manually curated set of 329 variants. Points are plotted by recall (x axis) and precision (y axis), with point color indicating the F_0.5_ score. For the p_neutral_ predictor, variants above the p_neutral_ threshold (neutral-like) were excluded from the predicted pLoF set, and variants below threshold were retained as predicted pLoF. Results are insensitive to modest changes in the p_neutral_ threshold because the p_neutral_ distribution is strongly bimodal. Horizontal and vertical error bars indicate 95% confidence intervals for recall and precision, respectively. **d,** Benchmark using RNA expression data from GTEx (stop-gained variants with GTEx allelic-expression coverage). For each variant, we estimated a posterior probability of NMD-like depletion from a mixture model fit to allelic expression, using a synonymous-variant background with three components (balanced expression, monoallelic/extreme imbalance, and random imbalance) plus an additional NMD component. Bars show, for each method, the ratio of the geometric mean posterior NMD probability for variants classified as pLoF versus variants classified as non-pLoF (higher values indicate better separation by NMD-consistent RNA signal). Numeric values above bars denote mean ratios.

We next leveraged this signal in strongly constrained genes to derive genome-wide rules for pLoF interpretation, by testing how p_neutral_ aligns with known genomic determinants of NMD. For stop-gained variants, NMD is triggered when the premature termination codon lies more than 50–55 nucleotides upstream of the most distal 3′ exon–exon junction^32^. Consistent with this, comparing p_neutral_ to junction distance revealed a cliff at ∼50 nucleotides (Extended Data Fig. 6a), supporting the use of p_neutral_ to calibrate additional NMD-relevant features.

We applied the same procedure to distance from the CDS start site (Extended Data Fig. 6b,d) and from the canonical transcript end for single-exon genes (Extended Data Fig. 6c), yielding informative thresholds used to construct LOFTEE-2. For splice-affecting variants, we compared two splicing impact predictors: Pangolin^33^ (Extended Data Fig. 6e) and SpliceAI^34^ (Extended Data Fig. 6f). Pangolin showed superior variant separation by p_neutral_ and was selected for LOFTEE-2. For several features, we defined both relaxed and stringent thresholds, yielding two annotation modes: LOFTEE-2 relaxed and LOFTEE-2 strict.

We benchmarked LOFTEE-2 against LOFTEE using three independent approaches. Against 329 manually curated variants^16^, LOFTEE-2 improved precision (v1: 0.66; v2 relaxed: 0.77; v2 strict: 0.90; Fig. 3C), the key metric for loss-of-function curation. LOFTEE-2 strict gained precision at the expense of recall (0.72, vs 1 for LOFTEE), whereas LOFTEE-2 relaxed retained high recall (0.93). Because NMD depletes transcripts carrying the variant allele, true pLoF variants are expected to cause allelic imbalance in autosomal genes. We therefore used GTEx^35^ allelic expression to model signatures of balanced expression, monoallelic expression, and NMD-driven depletion, and to test whether pLoF classifications recapitulate these patterns (Supplementary Material). LOFTEE-2 showed higher true-to-false geometric-mean posterior NMD ratios (v1: 4.24; v2 relaxed: 5.78; v2 strict: 6.05; Fig. 3d). Finally, defining pLoF variants with each method to compute LOEUF, LOFTEE-2 improved AUPRC in a severe HI versus all-genes benchmark (v1: 0.19; v2 relaxed: 0.224; v2 strict: 0.253; Extended Data Fig. 5b). Filtering pLoF variants directly with p_neutral_ also yielded a high-performing LOEUF (0.277), consistent with this benchmark’s focus on genes under strong purifying selection, for which p_neutral_ is especially informative.

### Combining deleterious missense and loss-of-function constraint improves disease gene detection

We next investigated whether predicted deleterious missense variants provide additional constraint information. Three deep learning variant effect predictors—ESM1v^36^, AlphaMissense^17^ (AM), and PopEVE^19^—show that observed/expected ratios for missense variants in gnomAD v4 decline with increasing predicted deleteriousness (Fig. 4a). For weakly deleterious variants, AM and PopEVE show observed/expected ratios exceeding synonymous variants (synonymous ≈ 1; AM = 1.16; PopEVE = 1.05), whereas ESM1v does not (0.93), likely reflecting overfitting to human population presence/absence data used to train AM and PopEVE. At the most deleterious end (top 1%), missense variants are more constrained than both average pLoF variants (obs/exp: pLoF = 0.55; AM = 0.28; ESM1v = 0.39; PopEVE = 0.16) and gene-matched pLoF variants (obs/exp: AM = 0.31; ESM1v = 0.42; PopEVE = 0.19; p = 1.57 × 10^-12^, 6.81 × 10^-7^, 1.76 × 10^-9^, respectively; Supplementary Material). While overfitting could partly explain this for AM and PopEVE, it cannot explain ESM1v’s pattern given its behavior for weakly deleterious variants.

**Figure 4.**
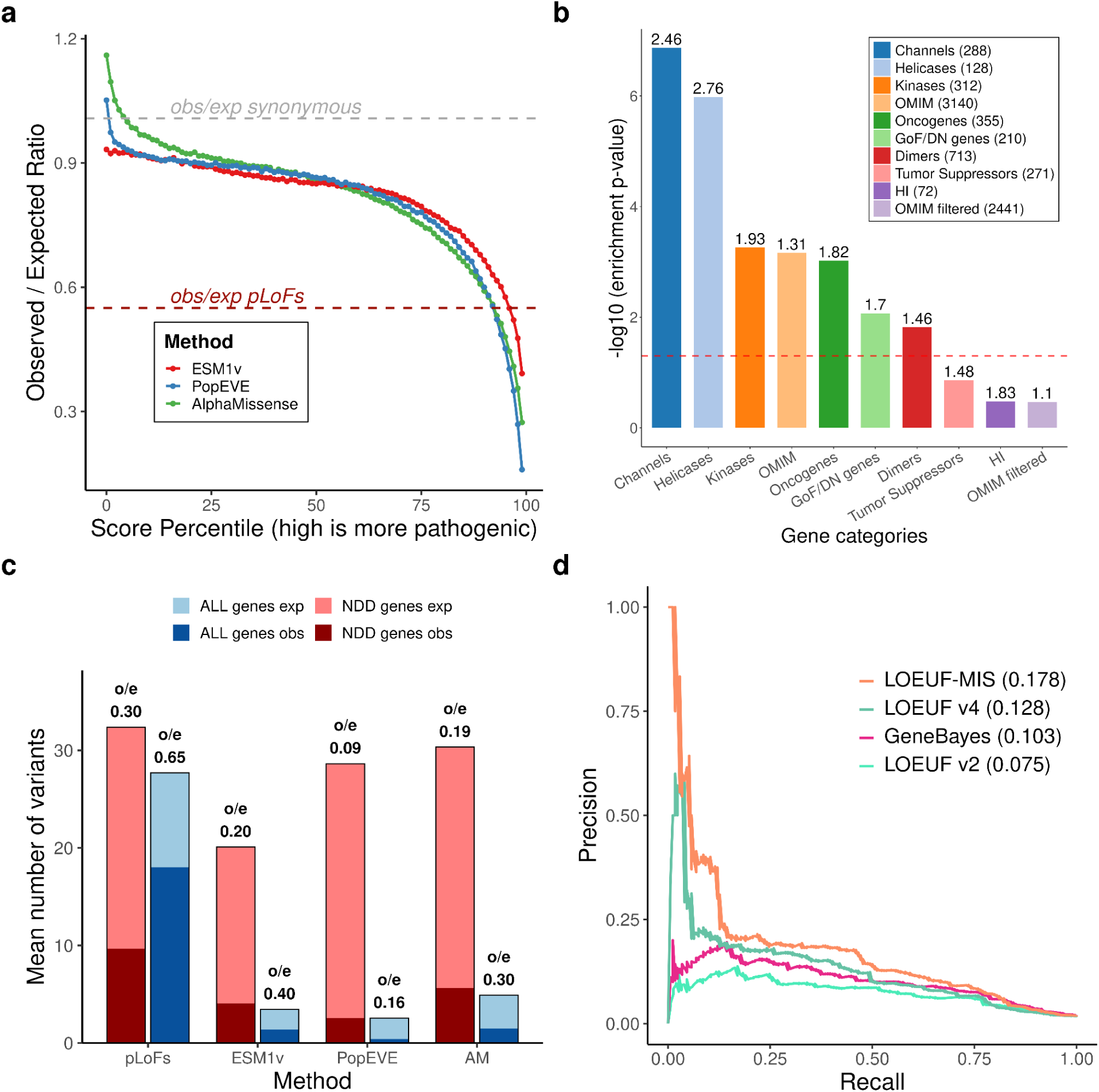
Constraint on highly deleterious missense variants and its contribution to disease gene detection. **a**, Observed/expected (obs/exp) ratios across score percentiles (99 = most pathogenic) for missense variants scored by three deep learning-based variant effect predictors: ESM1v, AlphaMissense (AM) and PopEVE. Dashed grey line denotes synonymous variants (obs/exp ≈ 1), and dashed red line denotes pLoF variants (obs/exp = 0.55). **b**, Enrichment of genes for which the most deleterious missense variants are more constrained than pLoF variants across gene categories, shown as −log10 enrichment P values. Filtered OMIM is the OMIM gene set from which all enriched categories have been removed. Enrichment values are displayed above each bar. **c**, Mean number of highly deleterious missense variants per gene in neurodevelopmental disorder (NDD) genes (left) and in all genes (right), compared with variants across methods (pLoF, and top 1% of each of ESM1v, PopEVE and AM). Bars show expected (light) and observed (dark) counts; obs/exp ratios are indicated above each bar. Only short genes are shown (expected < 50). **d**, Precision-recall curves for detection of NDD genes using constraint metrics based on pLoF variants alone (LOEUF v2 and v4) or combining pLoF and highly deleterious missense variants (LOEUF-MIS). Only short genes are used (expected pLoF < 50).

We hypothesized these variants may be enriched for gain-of-function (GoF) or dominant-negative (DN) mechanisms, for which heterozygotes can be more strongly selected against than LoF mechanisms. Consistent with this, when restricting analysis to HI genes, this effect disappears: the most deleterious missense variants are no longer more constrained than pLoF variants (Extended Data Fig. 7a), as expected in HI genes associated with severe phenotypes where LoF represents the most severe mechanism for disease.

By comparing missense and pLoF constraint, we can identify genes where GoF effects may be more deleterious than LoF, a property relevant for drug discovery, as illustrated by *PCSK9*^37,38^. Curated GoF/DN-prone genes^39^ are enriched for genes where missense variants are more constrained than pLoF variants compared with LOEUF-matched controls (enrichment = 1.7, p = 8.5 × 10^-3^), supporting our hypothesis (Fig. 4b). Several functional categories show similar enrichment: ion channels, which are known to be GoF-prone (enrichment = 2.47, p = 1.36 × 10^-7^); protein dimers, which are known to be DN-prone^40,41^ (enrichment = 1.46, p = 1.52 × 10^-2^); and oncogenes (enrichment = 1.82, p = 9.53 × 10^-4^), but not tumor suppressor genes (enrichment = 1.48, p = 0.14), consistent with their biology^42,43^. OMIM^44^ genes show enrichment (enrichment = 1.31, p = 6.92 × 10^-4^); however, this effect is attenuated after removing GoF/DN-prone categories (enrichment = 1.11, p = 0.34), indicating the OMIM enrichment is driven by genes with known GoF or DN mechanisms.

Constraint on the most deleterious missense variants can be combined with pLoF constraint to improve disease gene detection, particularly for short genes with few expected pLoF variants. Consistent with this, NDD genes are more constrained for deleterious missense variants in these genes (Fig. 4C). Incorporating the most deleterious 1% of missense variants according to AM, PopEve and ESM1v, into LOEUF improves NDD gene identification performance (AUPRC LOEUF: 0.126; LOEUF-MIS: 0.176; Fig. 4d and Extended Data Fig. 7b), many of which act through GoF mechanisms.

### Integrating literature and constraint improves disease gene prediction

Next, we leveraged the biomedical literature to derive a clinical impact score for each gene. We developed an agentic large language model (LLM) framework to extract disease associations, penetrance, inheritance patterns, clinical severity, age of onset, and molecular mechanisms from PubMed abstracts (Fig. 5a). For each gene, we compute a score we term Phenotype Evidence from Published Papers Extracted via Representation with Large Language Models (PEPPER_LLM_), of which we take a maximum across diseases and which serves two purposes: combination (this section) and contrasting (next section) with constraint metrics.

**Figure 5.**
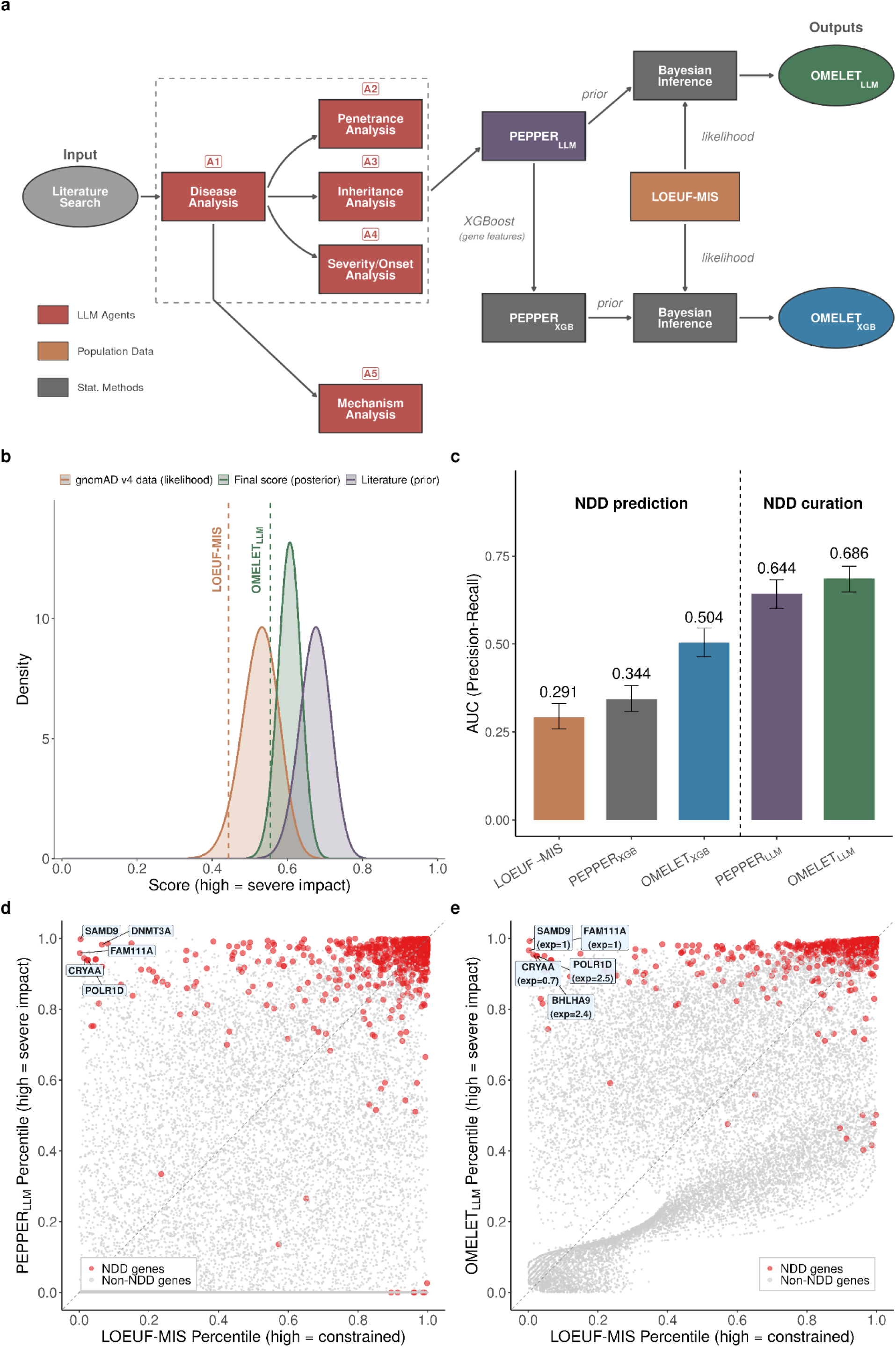
Literature-derived Clinical Impact Scores complement constraint-based metrics for disease gene prediction. **a**, Schematic of the agentic LLM framework and Bayesian integration approach. Five specialized LLM agents (A1-A5) extract clinical features from literature to compute PEPPER_LLM_. PEPPER serves as prior in Bayesian inference with LOEUF-MIS as likelihood, yielding Bayes(PEPPER_LLM_, LOEUF-MIS), which we term OMELET_LLM_. An XGBoost model trained on biological features enables literature-independent prediction, generating Bayes(PEPPER_XGB_, LOEUF-MIS), which is OMELET_XGB_. **b**, Distribution of literature-derived prior (purple), gnomAD v4 likelihood from observed and expected counts (orange), and Bayesian posterior (green) scores for an example gene (*ABCC9*). OMELET is the lower bound of the posterior distribution. **c**, Performance comparison on NDD gene prediction (585 NDD-positive and 16,582 negative). Error bars denote 95% confidence intervals from 2,000 stratified bootstrap resamples. **d**, Comparison of PEPPER percentile versus LOEUF percentile for all genes. NDD genes (red) cluster at high PEPPER values. Labeled genes show cases where literature information substantially improves prediction relative to constraint alone. **e**, Comparison of Bayesian score versus LOEUF percentile. NDD genes (red) cluster at high Bayes values. Labeled genes show cases where literature information substantially improves prediction relative to constraint alone.

To assess the accuracy of our LLM-based curation, we compared the 47,839 gene-disease associations identified by our pipeline with the GenCC database^45^. Our method recovered 95.7% of all GenCC associations (limited to definitive), with concordance exceeding 99% for well-established associations (Supplementary Material). Our pipeline also identified 1,961 highly penetrant associations not present in GenCC (Supplementary Material), demonstrating that LLM-based curation can reliably support manual curation.

PEPPER_LLM_ and LOEUF-MIS provide correlated but complementary measures of clinical impact: PEPPER_LLM_ reflects LLM-curated clinical evidence, whereas LOEUF-MIS quantifies depletion of variants in gnomAD population data. We integrated these orthogonal sources in a Bayesian framework, using literature-derived scores as priors and gnomAD variant counts as likelihood data to derive a posterior distribution. We termed the 95% lower bound of this posterior OMELET_LLM_ (Fig. 5a-b; Supplementary Material), which achieves an AUPRC of 0.686 for predicting NDD genes (Fig. 5c). Most NDD genes show higher predicted clinical impact when literature information is incorporated (Fig. 5d-e). The largest score increases occur for short genes with low variant counts and for GoF or DN disease mechanisms that PEPPER_LLM_ captures better than LOEUF-MIS (Fig. 5e).

Since NDD gene lists were curated using literature, comparing our LLM-based scores to this benchmark primarily tests curation quality rather than predictive power. To enable unbiased benchmarking, we trained an XGBoost model to predict PEPPER_LLM_ from gene-level features^30^, without using curated disease labels (Supplementary Material), yielding PEPPER_XGB_. Using fivefold cross-validation to ensure each gene’s prediction was independent of its literature-derived score, genes for which PEPPER_XGB_ most exceeds PEPPER_LLM_ are associated with significantly fewer published articles yet exhibit lower LOEUF values indicative of stronger selective constraint (Extended Data Fig. 8a-b), confirming that the model captures genuine biological signals in understudied genes rather than recapitulating literature biases. The resulting Bayesian score combining PEPPER_XGB_ with LOEUF-MIS, OMELET_XGB_ (Omnibus Mutation Effects with LOEUF and Embedded Texts), achieves a AUPRC of 0.504, outperforming either score alone (PEPPER_XGB_: 0.344; LOEUF-MIS: 0.291; bootstrap p-value < 5 × 10^-4^; Fig 5c). This approach effectively distills literature knowledge into a generalizable model based on biological features, complementing population-level constraint.

PEPPER_XGB_ enables prospective identification of disease genes with limited literature coverage, uncovering novel associations before sufficient literature accrues. For example, at the time of analysis, *DENND2B* had no GenCC or OMIM entry and only three PubMed-indexed publications, yielding a PEPPER_LLM_ in the 72th percentile. In contrast, PEPPER_XGB_ ranks *DENND2B* in the 96th percentile based on biological features, including high transcript diversity, strong missense constraint, and mouse development and brain expression. A recent study^46^ confirmed *DENND2B* as the cause of an autosomal dominant neurodevelopmental disorder with intellectual disability and vulnerability to psychosis, validating the model’s predictive capacity.

To systematically identify similar genes, we selected candidates with PEPPER_XGB_ above the 95th percentile, no established GenCC disease association, and strong discordance between PEPPER_XGB_ and PEPPER_LLM_ (Δ_PEPPER_, Supplementary Material). This yielded 220 candidate genes with significantly stronger constraint than background (median LOEUF 0.31 vs. 0.92; Wilcoxon rank-sum test, p = 2.4 × 10^-87^), indicating purifying selection despite lacking definitive disease associations (Supplementary Dataset 2). These genes represent high-priority targets for clinical sequencing and functional studies.

### Discordance between constraint and literature reveals high discovery potential genes

Beyond combining PEPPER_LLM_ and gnomAD constraint for improved prediction, direct comparison of these metrics enables identification of genes for which population constraint and literature-derived clinical evidence diverge. To this end, we defined a gene-level disagreement score that quantifies the distance between PEPPER_LLM-LoF_ (constructed from LoF disease associations from the agentic framework) and gnomAD loss-of-function constraint. This yields values close to zero when concordant, positive values when constraint exceeds literature-derived impact, and negative values when literature evidence exceeds constraint (Supplementary Material). We refer to this metric as the Discovery Potential (DisPo) score, as it highlights genes for which substantial disease-relevant biology likely remains to be discovered. As an illustrative example, the 220 candidate genes showing higher clinical significance by PEPPER_XGB_ than by PEPPER_LLM_ have a median DisPo score of 13.14, significantly higher than the median of 1.86 for other genes (Mann–Whitney p = 1.08 × 10^-103^), confirming that XGBoost generalizes literature knowledge using biological features to identify high discovery potential genes.

We examined DisPo scores among disease-associated genes submitted to GenCC and observed that DisPo increased with a more recent submission date, consistent with fewer publications for recently discovered genes (Spearman ρ = 0.96, p = 1.9 × 10^-6^; Fig. 6a), supporting the use of DisPo to prioritize genes whose disease associations are likely to be discovered or refined in the future. We further identified genes with high DisPo scores (top 1% DisPo = 182 genes, Extended Data Table 1, Supplementary Dataset 3), of which 26 also fall within the top 1% of Δ_PEPPER_ (Extended Data Table 1, Extended Data Figure 9).

**Figure 6.**
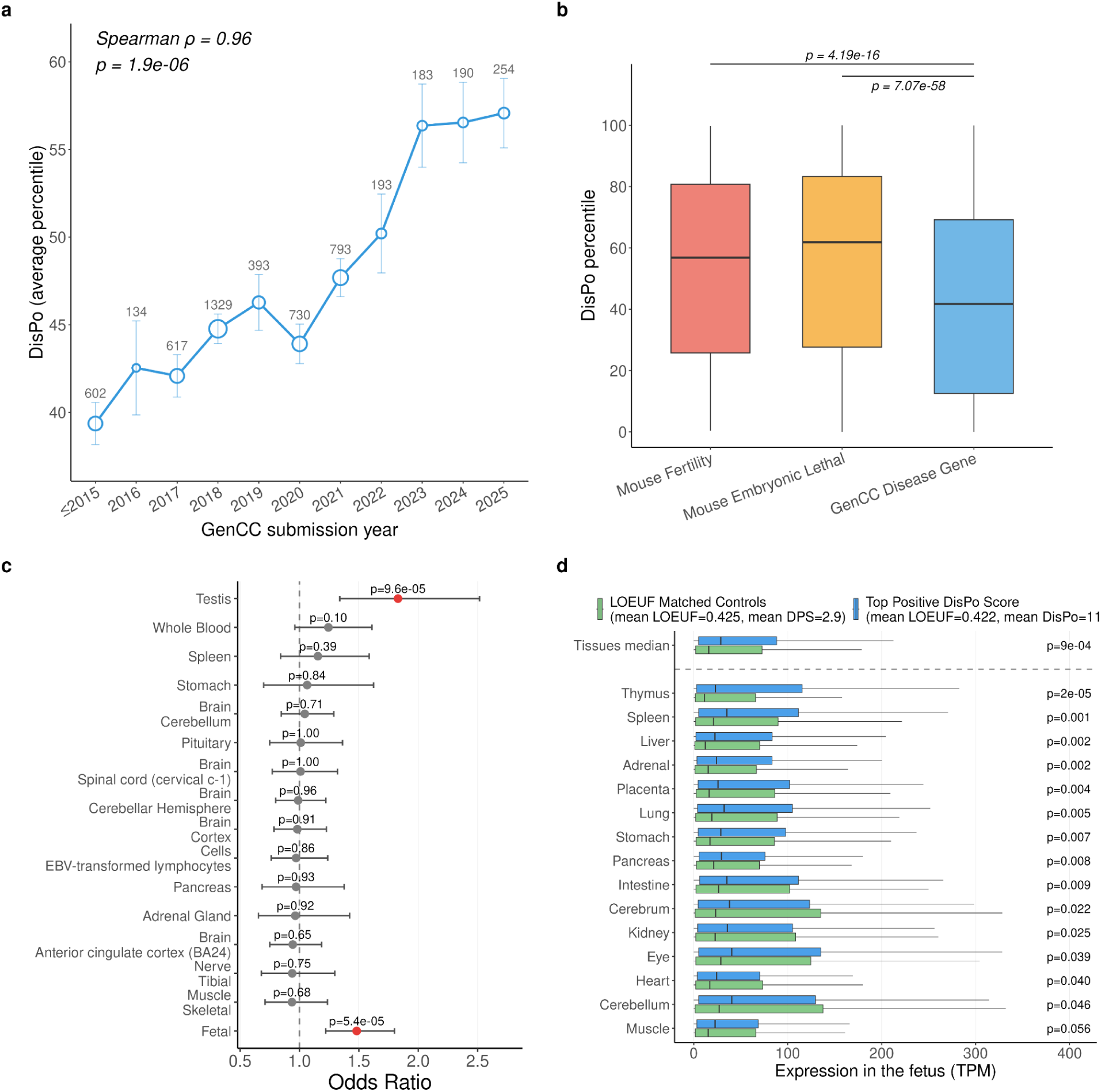
Discovery Potential (DP) across gene sets and tissues. **a**, Mean DisPo percentile of GenCC disease-associated genes by year of submission. Numbers above points indicate gene counts per year; error bars show s.e.m. **b**, DisPo score distributions for mouse infertility genes, mouse embryonic lethal genes, and GenCC disease-associated genes. Top 2 extreme DisPo genes are labeled in each category. **c,** Enrichment of tissue-specific expression among high-DisPo genes versus LOEUF-matched controls. Tissue specificity is defined as top-decile expression in a given GTEx tissue while below the 90th percentile of the median across all other tissues. The 15 tissues with the highest odds ratios (out of 52) are shown. "Fetal" denotes broad fetal expression after excluding genes with broadly elevated adult expression (median expression across all GTEx tissues ≥ 90th percentile) and high testis expression to remove fertility-related signals. Dots and bars show odds ratios with 95% confidence intervals; red, p < 0.05 (Fisher’s exact test). **d,** Fetal expression of high-DisPo genes versus LOEUF-matched controls (n = 901 pairs) after excluding genes with synonymous constraint depletion, high testis expression, or broadly elevated adult expression (see Supplementary Material). Box plots show transcript abundance (TPM) for each fetal tissue, sorted by significance. The top row shows the median TPM across all 15 tissues. P values are from one-sided Wilcoxon rank-sum tests (high-DisPo > controls).

DisPo can highlight genes with potential uncharacterized LoF disease even when a GoF disease is already established—a distinction that Δ_PEPPER_ does not capture, as it does not explicitly differentiate between LoF and GoF mechanisms. For example, *MTOR* (DisPo percentile=99.8, Δ_PEPPER_ = 61.4) is associated with a known gain-of-function disorder (Smith–Kingsmore syndrome), which does not explain the strong pLoF constraint observed in gnomAD. In mouse models, complete loss of *Mtor* results in early embryonic lethality^47^, yet no human disease attributable to *MTOR* loss-of-function was identified by our framework.

To generalize these results, we next assessed DisPo across three gene sets: disease-associated genes curated in GenCC (definitive and strong associations)^45^, mouse embryonic lethal genes^48^, and mouse infertility genes^48^ (Fig. 6b). Both mouse embryonic lethal and mouse infertility genes showed significantly higher DisPo scores than disease genes (median DisPo percentiles: mouse infertility = 56.8; mouse embryonic lethal = 61.9; disease genes = 41.7; Wilcoxon rank-sum test: mouse infertility vs. disease genes, p = 4.19 × 10^-16^; mouse embryonic lethal vs. disease genes, p = 7.07 × 10^-58^). This enrichment is consistent with their biology: pLoF variants affecting fertility or early embryonic development are under strong purifying selection but are often poorly captured in clinical datasets, resulting in high DisPo scores that reflect substantial unexplained constraint relative to documented clinical impact.

To explore the biological basis of high DisPo scores, we tested whether high-DisPo genes were enriched for tissue-specific expression across the 54 GTEx tissues. A gene was considered tissue-specific if its expression ranked above the 90th percentile for a given tissue but below the 90th percentile of the median across all other tissues, thereby excluding broadly expressed genes and focusing on effects more likely to produce viable, organ-specific phenotypes. Only testis showed significant enrichment (odds ratio = 1.83; Fisher’s exact test, p = 9.6 × 10^-5^; Fig. 6c). We next examined fetal-specific expression^49^ and found that high-DisPo genes were significantly enriched for broad fetal specific expression—defined as above-median expression across all 15 fetal tissues (odds ratio = 1.48; Fisher’s exact test, p = 5.4 × 10^-5^; Fig. 6c)—and exhibited elevated fetal expression both across tissues (median TPM; Wilcoxon rank-sum test, p = 9 × 10^-4^) and in 14 of 15 individual fetal tissues (p = 2 × 10^-5^ to 5.6 × 10^-2^; Fig. 6d).

## Discussion

With gnomAD v4, we observe high degrees of mutational saturation and recurrence, and continue to increase power for detecting strong selective constraint (the selection regime most relevant for rare human disease), with sustained gains expected beyond 10 million individuals. Alongside this enhanced resolution, we develop new methods for pLoF variant annotation (LOFTEE-2) and incorporate highly deleterious missense variants into LOEUF, improving disease gene prediction and enabling the detection of gain-of-function mechanisms. We systematically curate literature using large language models to extract gene–disease associations and distill this literature knowledge into a generalizable model trained on biological features (PEPPER_XGB_). By integrating this information with evolutionary constraint (OMELET_XGB_), we achieved improved prediction of gene-level clinical significance, outperforming existing methods. Finally, contrasting constraint and clinical literature evidence (DisPo) highlights genes with unexplained constraint and potentially undiscovered disease associations, such as those affecting fertility and embryonic development.

The saturation and recurrence found in gnomAD v4 will improve estimation of selection, mutation rates, linkage disequilibrium, and resolution for demographic events^50^. Furthermore, increased resolution of recurrence will enhance detection of genes involved in clonal expansion in sperm^51,52^ and blood^53^, and allow for quantification of expansion effects, directly informing prevalence estimates for several disorders^54^.

We identify genes in which deleterious missense variation is more constrained than LoF variation, a pattern that can highlight GoF/DN mechanisms, and show that this set is enriched for genes with established GoF/DN effects, such as oncogenes. Identifying genes where GoF/DN is more deleterious than LoF is therapeutically valuable: when GoF/DN is severe but LoF is mild, gene knockdown or inhibition may yield less deleterious outcomes. However, GoF/DN is not the only mechanism leading to stronger constraint against missense variants than pLoF variants. Clonal expansion genes may show inflated LoF tolerance because LoF can be advantageous in germline or blood cells while neutral or deleterious in the diploid organism. For example, *LZTR1* shows stronger missense than pLoF constraint (p = 0.0004), driven by a high pLoF observed/expected ratio (2.08) due to blood clonal expansion when lost^55^. A third explanation is transcriptional adaptation, whereby NMD induces compensatory upregulation of a paralog, reducing LoF but not missense variant impact^56,57^. Distinguishing among these three mechanisms will require future studies that explicitly account for all factors.

Our LLM-based curation of gene-disease associations can retrieve features such as penetrance, mode of inheritance, and mechanism of disease when they are annotated in the literature, offering a promising alternative or complement to traditional keyword-based text mining^58,59^. However, further work is needed to explore edge cases, build manually curated benchmarks, and compare different LLMs, including biomedical fine-tuned models versus general-purpose models. The clinical significance score PEPPER_LLM_ serves as a training target to generalize gene-level features associated with clinical impact (PEPPER_XGB_), enabling prospective gene discovery. Future extensions could predict specific disease attributes such as predominant molecular mechanism or mode of inheritance. Genes that are more constrained than expected based on the literature (high-DisPo) genes are enriched for expression in fetal tissues and embryonic lethality, biological contexts that strongly influence LOEUF while remaining underrepresented in the literature, as highlighted by the *MTOR* example. We envision integrating the DisPo score into gene prioritization pipelines for rare diseases, particularly those disorders with reduced reproductive fitness.

This study introduces a substantial number of complementary metrics, each addressing distinct analytical objectives. To assist users in selecting the appropriate metric for their specific application, we provide detailed practical guidance in the Supplementary Material. In brief, LOEUF should be used when the goal is to assess LoF constraint specifically, while LOEUF-MIS is preferable for a broader measure of intolerance to high-impact coding variants, particularly for short genes or those with GoF/DN mechanisms. For predicting clinically significant genes without literature bias, OMELET_XGB_ is the single best-performing metric that does not access the literature. To identify understudied disease gene candidates, users may consider DisPo for LoF-specific under-characterization or Δ_PEPPER_ for mechanism-agnostic discovery, with genes flagged by both approaches representing especially strong candidates for functional follow-up.

Our results are enabled by the continued growth of sequencing datasets and willingness of the data generators to contribute data toward this public allele frequency resource that benefits researchers worldwide. We show that the increase of observed mutations is better achieved through broader ancestry representation than further sampling within the same ancestry group. Accordingly, methods to aggregate data across federated sites while retaining high accuracy will enable the integration of restricted international datasets, thereby increasing ancestral representation and achieving continued growth and maximum utility of these resources.

## Data Availability

Aggregate allele frequencies, functional annotations, and constraint scores derived from the variant callset can be browsed and downloaded via the gnomAD browser at https://gnomad.broadinstitute.org/data#v4. We provide sites-level variant call format (VCF) files as well as Hail Tables for the exome and genome data. Other relevant data, such as all sites allele number, are available as Hail Tables or tab-separated TSV files.

In addition, we provide three Supplementary Datasets:

Supplementary Dataset 1: LOFTEE-2 and p_neutral_ for all putative pLoF sites (stop-gained and essential splice) in the exome

Supplementary Dataset 2: 220 candidate genes flagged by Δ_PEPPER_, as under characterized in the literature (PEPPER_XGB_ above the 95th percentile, no established GenCC disease association, and Δ_PEPPER_ > 25)

Supplementary Dataset 3: All presented scores, including LOEUF, LOEUF-MIS, PEPPER_XGB_, OMELET_XGB_, PEPPER_LLM_, OMELET_LLM_, DisPo, and associated statistics, for all genes

## Code Availability

Analysis scripts, pipelines, and supporting code for our quality control (QC) of gnomAD v4.1 are available at our GitHub repository: https://github.com/broadinstitute/gnomad_qc/releases/tag/v4.1

Code for recreating the analysis and plots in the main text can be found in the following Github repository: https://github.com/atgu/gnomAD_v4_flagship_paper

## Acknowledgements

We thank John Novembre, Luke Anderson-Trocmé, Bram Gorissen, Sanna Gudmundsson, and Trisha Karani for helpful discussions. The authors thank the individuals whose data is in gnomAD for their contributions to research. Research reported in this publication was supported by the National Institutes of Health, including the National Human Genome Research Institute (U24HG011450, U01HG011755, UG3HG014379, and R01HG012867), the National Institute Of General Medical Sciences under award number R35GM157035, and the National Institute of Diabetes and Digestive and Kidney Diseases under award number U54DK105566. The content is solely the responsibility of the authors and does not necessarily represent the official views of the National Institutes of Health. This work was also supported by the Novo Nordisk Foundation (NNF21SA0072102), the Simons Foundation (SFARI 1009802), a Manton Center for Orphan Disease Research Fellowship, and a Boston Children’s Hospital Pilot Grant.

## Extended Data Figures

**Extended Data Figure 1.**
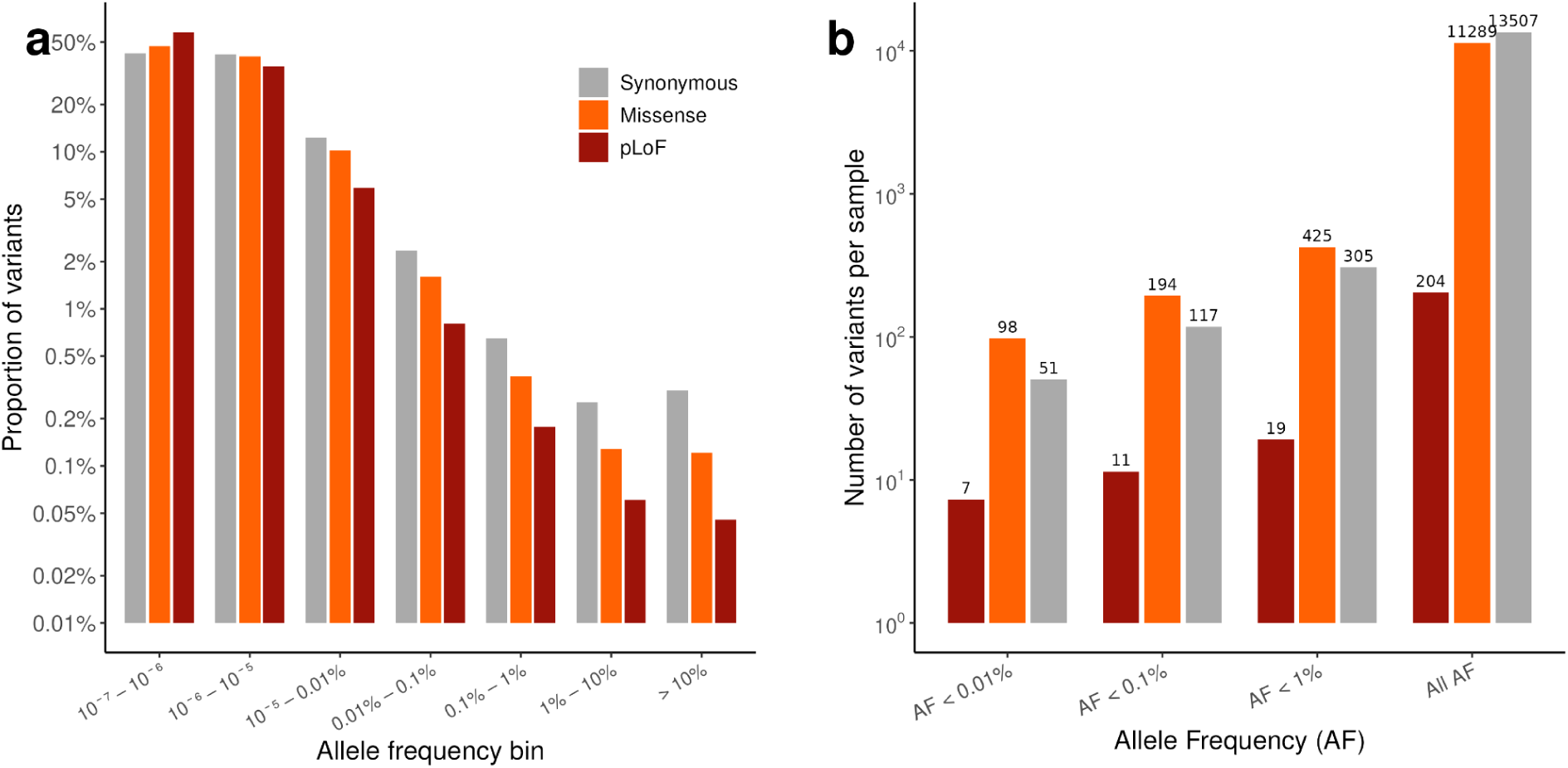
Variant distribution and per-sample variant counts in gnomAD v4. **a,** Proportion of variants in each allele frequency bin for synonymous, missense, and predicted loss-of-function (pLoF) variants in gnomAD v4 (global). The majority of variants across all consequence classes fall in the rarest frequency bins. **b,** Mean number of variants per sample at different allele frequency thresholds, stratified by consequence class (synonymous, missense, and pLoF). Note the log-scaled y-axes in both panels.

**Extended Data Figure 2.**
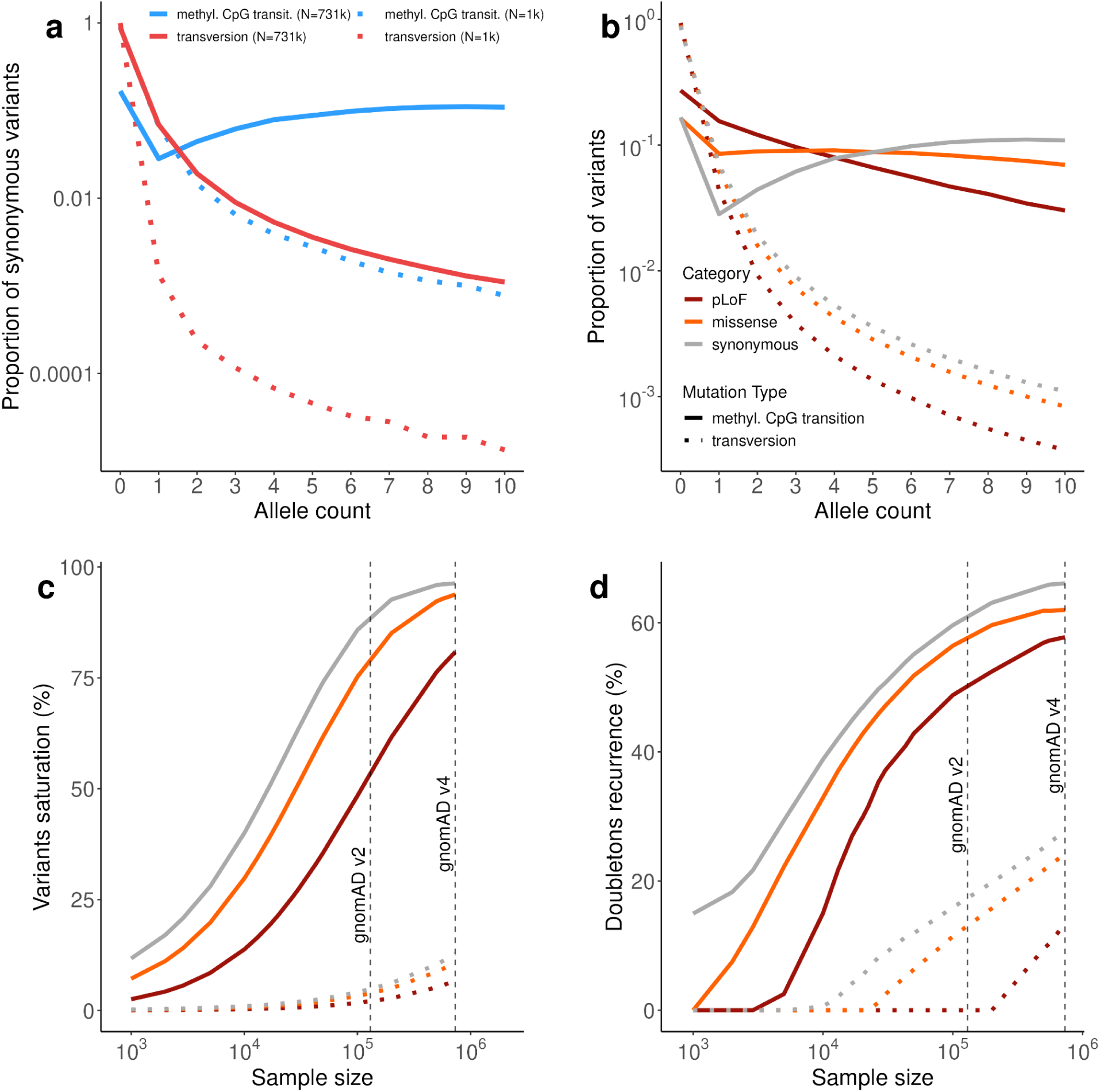
Observed variation, saturation, and mutational recurrence in gnomAD v4. **a**, Site frequency spectrum of synonymous variants comparing methylated CpG transitions (solid blue lines) and transversions (dotted red lines) at full sample size (N≈731k) and downsampled (N=1k), illustrating the effect of sample size on the observed allele frequency distribution. **b**, Proportion of observed variants as a function of allele count for predicted loss-of-function (pLoF), missense, and synonymous variants, stratified by mutation type (methylated CpG transitions shown as solid lines, transversions shown as dotted lines). **c**, Percentage of variants reaching saturation (observation of all possible variants) for pLoF, missense, and synonymous variants as a function of sample size. Solid lines represent methylated CpG transitions; dotted lines represent transversions. Dashed vertical lines indicate the sample sizes of gnomAD v2 and gnomAD v4. **d**, Percentage of doubletons showing recurrence (variants observed twice that arose from independent mutational events) for pLoF, missense, and synonymous variants, comparing methylated CpG transitions (solid lines) and transversions (dotted lines) as a function of sample size. Dashed vertical lines indicate gnomAD v2 and gnomAD v4 sample sizes.

**Extended Data Figure 3.**
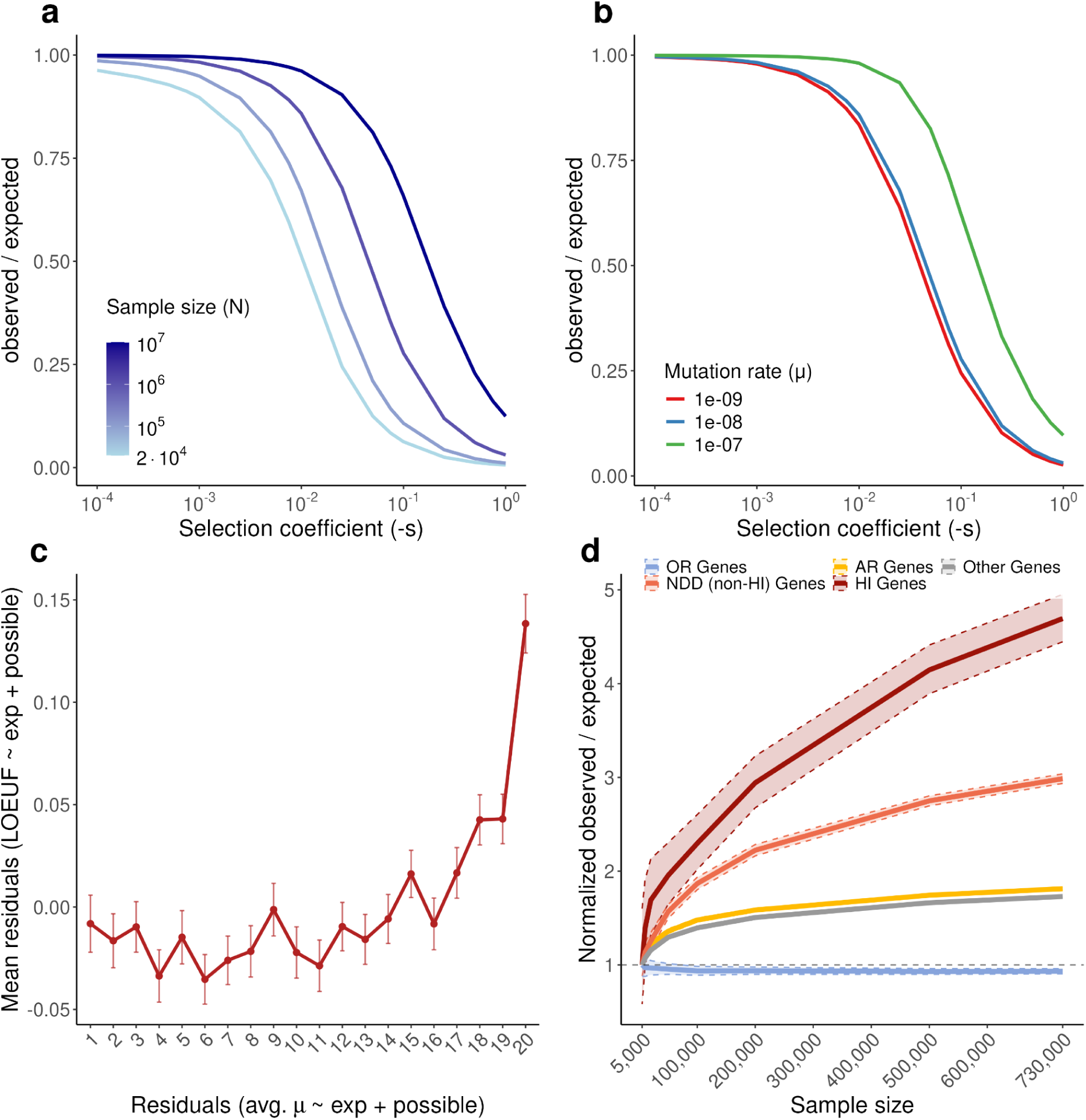
Analytical predictions and empirical validation of saturation effects on LOEUF. **a**, Analytical predictions showing how the observed/expected (obs/exp) ratio changes with sample size under different selection coefficients (s). As sample size increases, saturation causes the obs/exp ratio to increase toward 1, with the effect being more pronounced for genes under weaker selection. **b**, Analytical predictions demonstrating the effect of mutation rate on obs/exp ratios. Higher mutation rates lead to earlier saturation and faster increases in obs/exp ratios at smaller sample sizes. **c**, Relationship between per-gene mutation rate and LOEUF after double residualization. The average per-site LoF mutation rate (expected / possible variants) and LOEUF were each independently residualized against the expected number of LoF variants and the number of possible LoF variant sites using generalized additive models (GAMs). Genes were binned into vigintiles (5% quantiles) based on their mutation rate residuals (x-axis), and the mean LOEUF residual (y-axis) is shown for each bin. Error bars indicate standard error of the mean. The increase in LOEUF residuals in the highest vigintiles indicates that genes with higher-than-expected mutation rates tend to appear less constrained, consistent with the predicted effect of mutation rate on LOEUF. **d**, Empirical validation using gnomAD data showing obs/exp ratios as a function of sample size for different gene categories: haploinsufficient (HI) genes representing strong selection, neurodevelopmental disorder (NDD) genes representing intermediate selection, autosomal recessive (AR) genes representing weak heterozygous selection, and olfactory receptor (OR) genes representing neutral evolution. The empirical patterns qualitatively match the analytical predictions, with genes under strong selection (HI) showing smaller increases in obs/exp ratio compared to genes under weaker or no selection (AR, OR). Dashed vertical lines indicate the sample sizes of gnomAD v2 and gnomAD v4.

**Extended Data Figure 4.**
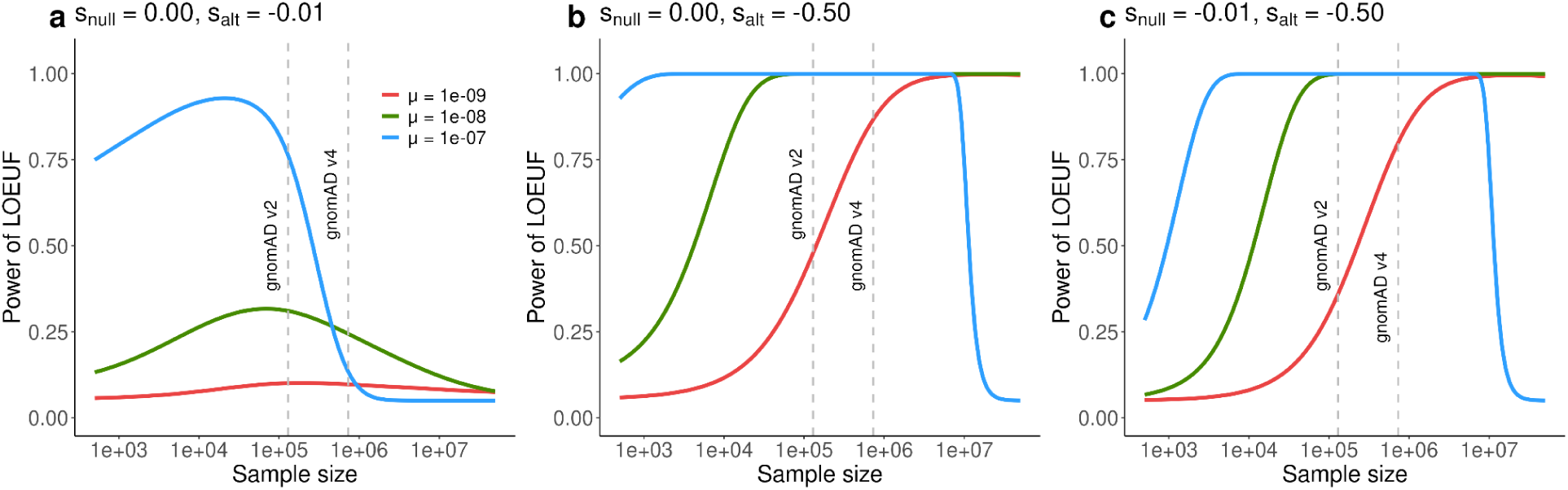
Power Analysis for LOEUF across different selection regimes and sample sizes. **a**, Power to detect loss-of-function constraint (measured as LOEUF) under neutral null hypothesis (s_null_ = 0.00) and weak alternative hypothesis (s_alt_ = -0.01) across three mutation rates (μ). The blue line (μ = 1e-07) shows high power at moderate sample sizes (∼10⁴-10⁵), while red (μ = 1e-09) and green (μ = 1e-08) lines show limited power even at large sample sizes. Vertical dashed lines indicate gnomAD v2 and v4 sample sizes. **b**, Power analysis under neutral null (s_null_ = 0.00) and strong alternative hypothesis (s_alt_ = -0.50) across mutation rates. All three mutation rates achieve near-complete power (approaching 1.0) at different sample size thresholds, with higher mutation rates reaching saturation at smaller sample sizes. The blue line (μ = 1e-07) reaches maximum power earliest, followed by green (μ = 1e-08) and red (μ = 1e-09). **c**, Power analysis under weakly selected null hypothesis (s_null_ = -0.01) and strong alternative hypothesis (s_alt_ = -0.50). This represents the realistic scenario of distinguishing genes under strong selection from those under weak selection. Power increases monotonically with sample size for all mutation rates, with substantial power gains continuing beyond gnomAD v4 sample sizes, particularly for lower mutation rates. These analyses demonstrate that substantial statistical power remains to be gained from further sample size increases, especially for genes under strong purifying selection (most relevant for rare disease), and that the rate of power gain depends on both selection coefficient and mutation rate.

**Extended Data Figure 5.**
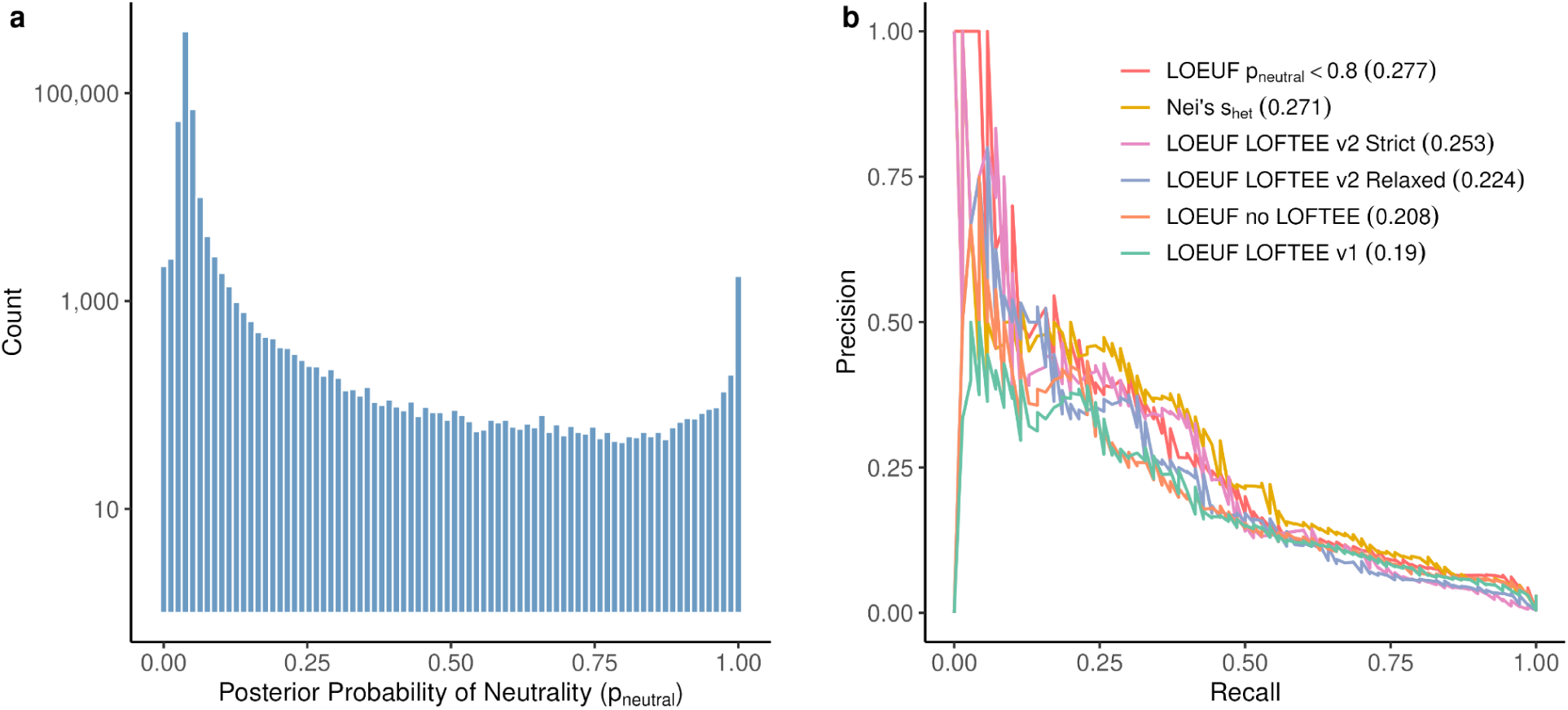
Distribution of probability of neutrality (p_neutral_) and benchmarking of constraint metrics for haploinsufficient gene detection. **a**, Genome-wide distribution of posterior probability of neutrality (p_neutral_) for pLoF variants, shown on a log10 scale. The distribution is strongly bimodal, with the vast majority of variants showing low p_neutral_ (left peak, p_neutral_ near 0) and a small subset showing high p_neutral_ (right peak, p_neutral_ near 1). Approximately 0.52% of variants exhibit p_neutral_ above 0.8, indicating likely false positive loss-of-function annotations. **b**, Precision-recall curves comparing the performance of different constraint metrics and loss-of-function annotation approaches for distinguishing haploinsufficient (HI) genes from all other genes. Area under the precision-recall curve (AUPRC) values are shown in parentheses in the legend. LOEUF with p_neutral_ filtering (p_neutral_ < 0.8, red line) shows the highest performance (AUPRC = 0.277), followed by Nei’s selection coefficient estimate (s_het_, yellow line, AUPRC = 0.271), LOFTEE-2 strict (pink line, AUPRC = 0.253), LOFTEE-2 relaxed (purple line, AUPRC = 0.224), unfiltered LOEUF (orange line, AUPRC = 0.208), and LOFTEE v1 (green line, AUPRC = 0.19).

**Extended Data Figure 6.**
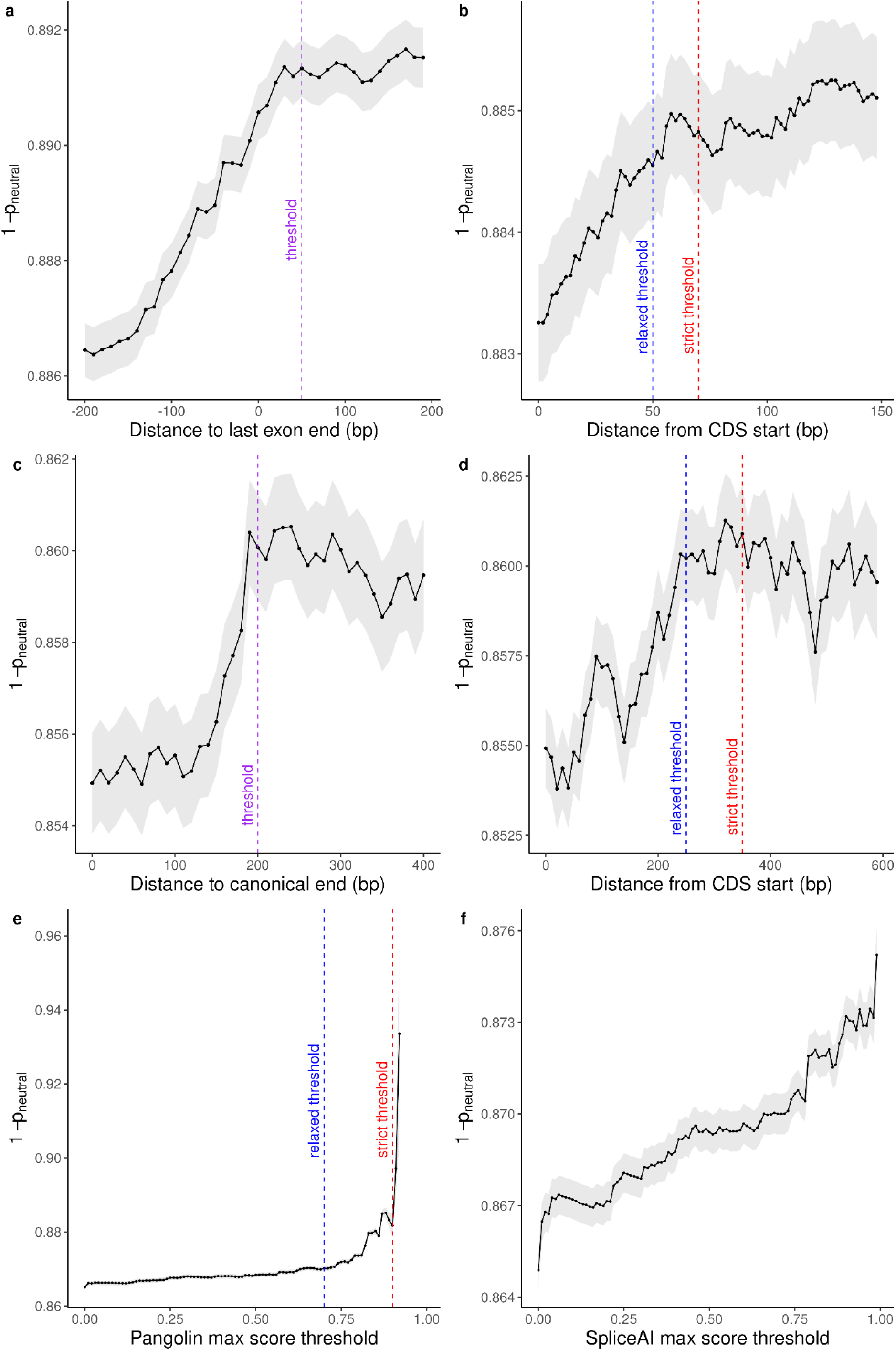
Calibration of genomic features for LOFTEE-2 using 1 - probability of neutrality (p_neutral_). **a**, Cumulative mean of 1 - p_neutral_ as a function of distance to the last exon-exon junction for stop-gained variants. The inflection point occurs at approximately 50 nucleotides (indicated by dashed purple line), consistent with the known 50-55 nucleotide rule for nonsense-mediated decay (NMD) triggering. Grey shading indicates 95% confidence intervals. **b**, Cumulative mean of 1 - p_neutral_ as a function of distance from the coding sequence (CDS) start site for stop-gained variants. Two thresholds are identified: a relaxed threshold (blue dashed line) and a strict threshold (red dashed line), used to define LOFTEE-2 relaxed and strict modes, respectively. Variants occurring very close to the start codon show elevated probability of neutrality. **c**, Cumulative mean of 1 - p_neutral_ as a function of distance to the canonical transcript end for single-exon genes. The purple dashed line indicates the threshold used to filter likely neutral variants in single-exon genes, where standard NMD rules may not apply. **d**, Cumulative mean of 1 - p_neutral_ as a function of distance from CDS start for single-exon genes, showing similar patterns to multi-exon genes but with distinct thresholds (blue and red dashed lines for relaxed and strict modes). **e**, Cumulative mean of 1 - p_neutral_ as a function of Pangolin splicing impact prediction scores. Pangolin scores range from 0 to 1, with higher scores indicating greater predicted splicing disruption. The inflection point (purple dashed line indicates relaxed threshold, red indicates strict threshold) demonstrates Pangolin’s ability to separate true splice-disrupting variants from neutral ones. **f**, Cumulative mean of 1 - p_neutral_ as a function of SpliceAI maximum delta score. While SpliceAI shows some separation ability, comparison with panel E reveals that Pangolin demonstrates superior variant separation by p_neutral_, leading to its selection for LOFTEE-2. Grey shading indicates 95% confidence intervals for all panels.

**Extended Data Figure 7.**
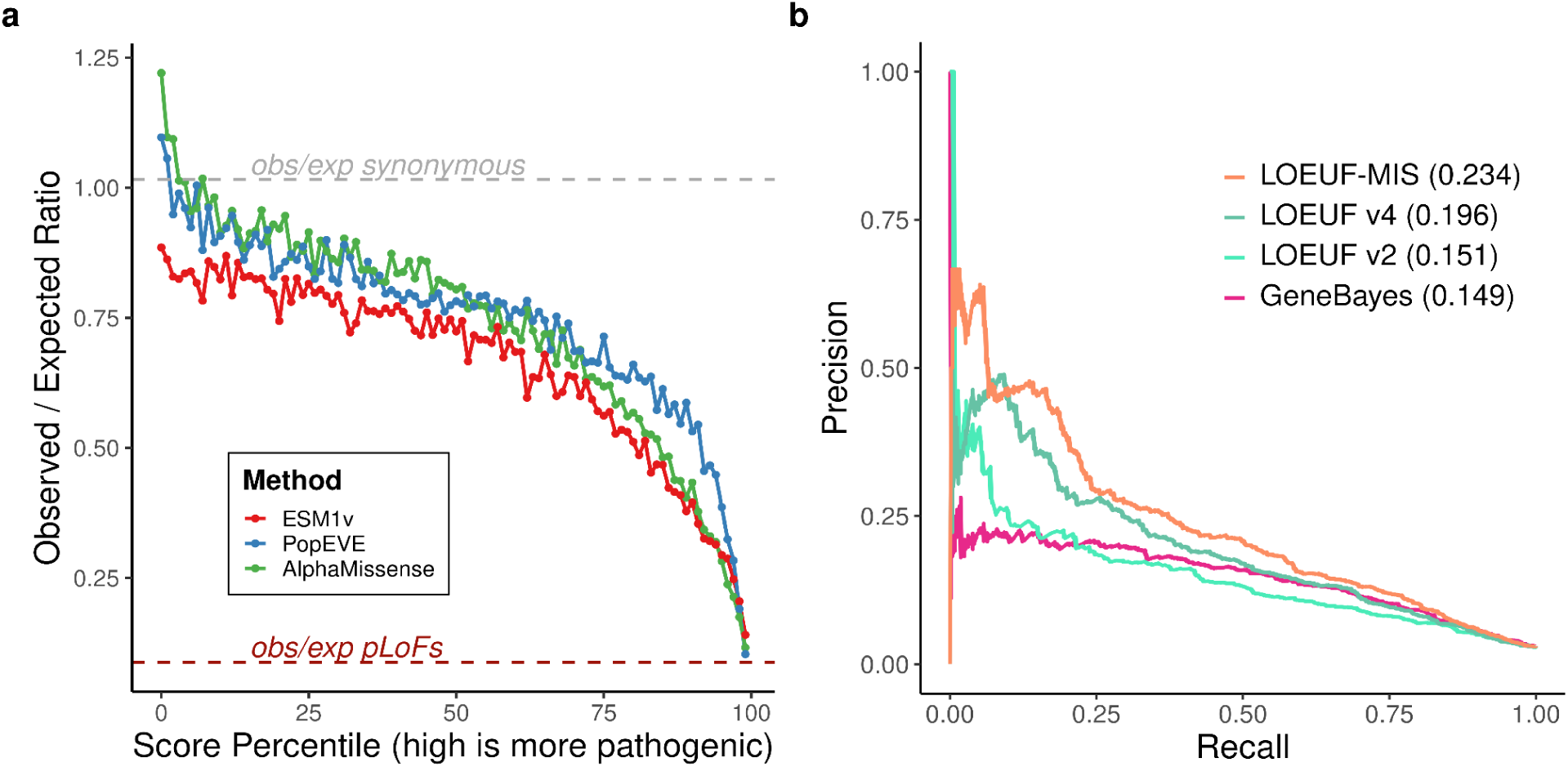
Missense constraint in haploinsufficient genes and performance of combined constraint metrics. **a**, observed/expected (obs/exp) ratios of missense variants across predicted pathogenicity percentiles in haploinsufficient (HI) genes only, using three variant effect predictors: ESM1v (red), PopEVE (blue), and AlphaMissense (green). The dashed grey line indicates synonymous variant obs/exp ratio (≈1.0), and the dashed brown line shows pLoF variant obs/exp ratio. In HI genes, the most deleterious missense variants do not show stronger constraint than pLoF variants, unlike the pattern observed across all genes. **b**, Precision-recall curves comparing constraint metrics for detecting neurodevelopmental disorder (NDD) genes. LOEUF-MIS (orange, AUPRC = 0.22) combines loss-of-function and highly deleterious missense variants. Other metrics shown are LOEUF v4 (dark green, AUPRC = 0.193), LOEUF v2 (light green, AUPRC = 0.15), and GeneBayes (pink, AUPRC = 0.148). AUPRC values are shown in parentheses.

**Extended Data Figure 8.**
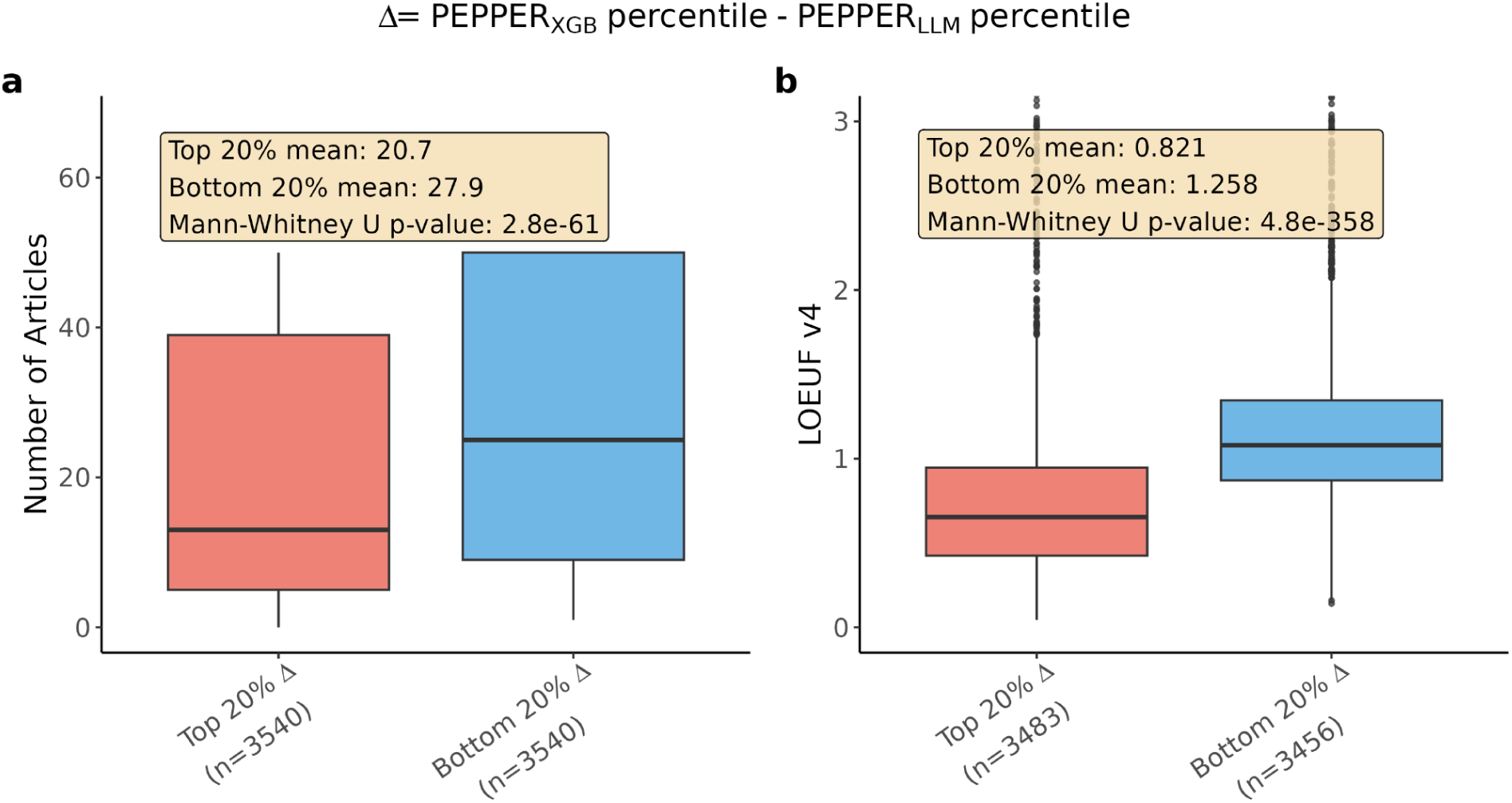
PEPPER_XGB_ captures constrained, understudied genes independently of literature coverage. Boxplots comparing the top 20% of genes most upranked by PEPPER_XGB_ relative to PEPPER_LLM_ (red) versus the bottom 20% most downranked (blue), ranked by the difference in percentile scores. **a**, Number of PubMed articles retrieved per gene. Genes upranked by PEPPER_XGB_ are associated with significantly fewer publications, indicating that the model does not simply recapitulate literature abundance. **b**, LOEUF v4. Genes upranked by PEPPER_XGB_ show significantly lower LOEUF values, reflecting greater intolerance to loss-of-function variants. Together, these results demonstrate that PEPPER_XGB_ identifies genes under strong selective constraint that are under-represented in the literature, supporting its utility as a literature-independent predictor of clinical significance. P-values are from one-sided Mann-Whitney U tests. Box plots show median (black line), interquartile range (box), and 1.5× IQR (whiskers).

**Extended Data Figure 9.**
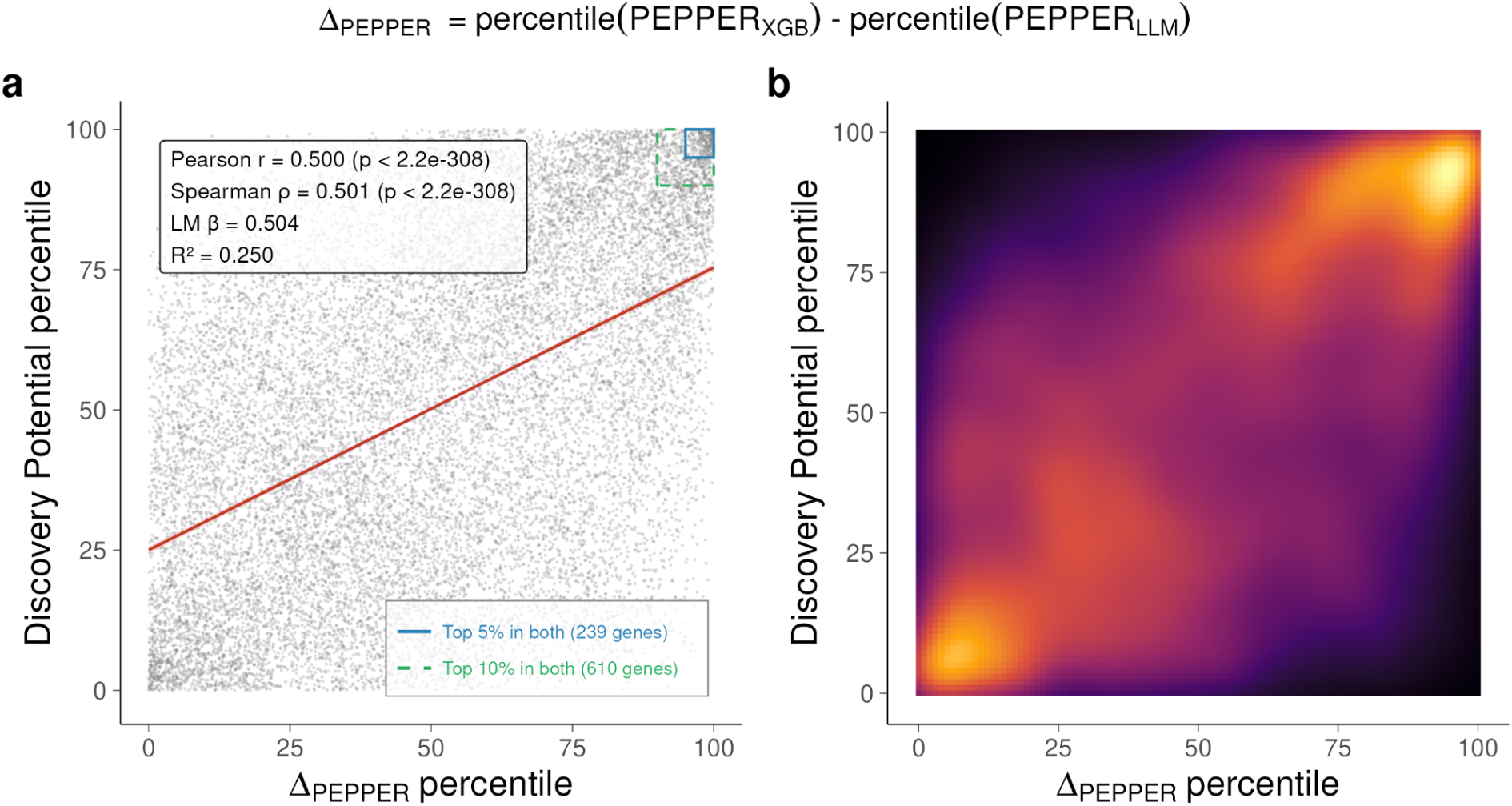
Relationship between Δ_PEPPER_ and Discovery Potential scores across 17,170 protein-coding genes. Δ_PEPPER_ is defined as the percentile rank of the difference between PEPPER_XGB_ and PEPPER_LLM_ percentile scores; high values identify genes for which the feature based model predicts substantially greater pathogenicity than literature evidence alone. (a) Scatter plot of Δ_PEPPER_ percentile versus Discovery Potential percentile. Red line, ordinary least-squares fit (shaded area, 95% CI). Dashed green and solid blue rectangles mark genes in the top 10% (n = 611) and top 5% (n = 239) for both metrics, respectively. Pearson r = 0.50, Spearman ρ = 0.50 (both P ≈ 0). (b) Two-dimensional kernel density estimate of the same data (inferno colour scale). For a more detailed analysis of this figure, see the last chapter of the Supplementary Material.

**Extended Data Table 1.** Number of genes in the top 1%, 5%, and 10% percentiles for each scoring metric individually and their intersection. Δ_PEPPER_ is defined as the percentile rank of the difference between PEPPER_XGBoost_ and PEPPER_LLM_ percentile scores. Discovery Potential (DisPo) is the distance between constraint and PEPPER_LLM_, with high positive values highlighting genes highly constrained despite lack of Mendelian disease association in the literature. "Both" denotes genes simultaneously exceeding the indicated threshold for both metrics. Gene counts are based on n = 17,170 protein-coding genes with non-missing values for all three scores.

| Top percentile | $\Delta_{\text{PEPPER}}$ | DisPo | In both |
| --- | --- | --- | --- |
| 1% | 172 | 182 | 26 |
| 5% | 859 | 897 | 239 |
| 10% | 1,717 | 1,780 | 611 |

## References

1. Karczewski, K. J. et al. The mutational constraint spectrum quantified from variation in 141,456 humans. Nature 581, 434–443 (2020).

2. Chen, S. et al. A genomic mutational constraint map using variation in 76,156 human genomes. Nature (2023) doi:10.1038/s41586-023-06045-0.

3. Bycroft, C. et al. The UK Biobank resource with deep phenotyping and genomic data. Nature 562, 203–209 (2018).

4. All of Us Research Program Genomics Investigators. Genomic data in the All of Us Research Program. Nature 627, 340–346 (2024).

5. 1000 Genomes Project Consortium et al. A global reference for human genetic variation. Nature 526, 68–74 (2015).

6. Lek, M. et al. Analysis of protein-coding genetic variation in 60,706 humans. Nature 536, 285–291 (2016).

7. Kosmicki, J. A. et al. Refining the role of de novo protein-truncating variants in neurodevelopmental disorders by using population reference samples. Nat. Genet. 49, 504–510 (2017).

8. Satterstrom, F. K. et al. Large-Scale Exome Sequencing Study Implicates Both Developmental and Functional Changes in the Neurobiology of Autism. Cell 180, 568–584.e23 (2020).

9. Kaplanis, J. et al. Evidence for 28 genetic disorders discovered by combining healthcare and research data. Nature 586, 757–762 (2020).

10. Abou Tayoun, A. N., et al. Recommendations for interpreting the loss of function PVS1 ACMG/AMP variant criterion. Hum. Mutat. 39, 1517–1524 (2018).

11. Seaby, E. G. et al. Targeting de novo loss-of-function variants in constrained disease genes improves diagnostic rates in the 100,000 Genomes Project. Hum. Genet. 142, 351–362 (2023).

12. Baxter, S. M. et al. Centers for Mendelian Genomics: A decade of facilitating gene discovery. Genet Med 24, 784–797 (2022).

13. Wright, C. F. et al. Genomic Diagnosis of Rare Pediatric Disease in the United Kingdom and Ireland. N Engl J Med 388, 1559–1571 (2023).

14. Chung, C. C. Y. et al. Meta-analysis of the diagnostic and clinical utility of exome and genome sequencing in pediatric and adult patients with rare diseases across diverse populations. Genet Med 25, 100896 (2023).

15. Bamshad, M. J., Nickerson, D. A. & Chong, J. X. Mendelian Gene Discovery: Fast and Furious with No End in Sight. Am. J. Hum. Genet. 105, 448–455 (2019).

16. Singer-Berk, M. et al. Advanced variant classification framework reduces the false positive rate of predicted loss-of-function variants in population sequencing data. Am J Hum Genet 110, 1496–1508 (2023).

17. Cheng, J. et al. Accurate proteome-wide missense variant effect prediction with AlphaMissense. Science 381, eadg7492 (2023).

18. Lin, Z. et al. Evolutionary-scale prediction of atomic-level protein structure with a language model. Science 379, 1123–1130 (2023).

19. Orenbuch, R. et al. Proteome-wide model for human disease genetics. Nat. Genet. 57, 3165–3174 (2025).

20. Seaby, E. G., Rehm, H. L. & O’Donnell-Luria, A. Strategies to uplift novel Mendelian gene discovery for improved clinical outcomes. Front. Genet. 12, 674295 (2021).

21. Dawes, R., Lek, M. & Cooper, S. T. Gene discovery informatics toolkit defines candidate genes for unexplained infertility and prenatal or infantile mortality. NPJ Genom. Med. 4, 8 (2019).

22. Dickinson, M. E. et al. High-throughput discovery of novel developmental phenotypes. Nature 537, 508–514 (2016).

23. Poterba, T. et al. The scalable variant call representation: enabling genetic analysis beyond one million genomes. Bioinformatics 41, (2024).

24. Koenig, Z. et al. A harmonized public resource of deeply sequenced diverse human genomes. Genome Res. 34, 796–809 (2024).

25. Bergström, A. et al. Insights into human genetic variation and population history from 929 diverse genomes. Science 367, (2020).

26. National Academies of Sciences, Engineering, and Medicine. Using Population Descriptors in Genetics and Genomics Research: A New Framework for an Evolving Field. (National Academies Press, 2023).

27. Gravel, S. et al. Demographic history and rare allele sharing among human populations. Proc. Natl. Acad. Sci. U. S. A. 108, 11983–11988 (2011).

28. Henn, B. M. et al. Distance from sub-Saharan Africa predicts mutational load in diverse human genomes. Proc. Natl. Acad. Sci. U. S. A. 113, E440–9 (2016).

29. MacArthur, D. G. & Tyler-Smith, C. Loss-of-function variants in the genomes of healthy humans. Hum. Mol. Genet. 19, R125–30 (2010).

30. Zeng, T., Spence, J. P., Mostafavi, H. & Pritchard, J. K. Bayesian estimation of gene constraint from an evolutionary model with gene features. Nat. Genet. 56, 1632–1643 (2024).

31. Nei, M. The frequency distribution of lethal chromosomes in finite populations. Proc. Natl. Acad. Sci. U. S. A. 60, 517–524 (1968).

32. Nagy, E. & Maquat, L. E. A rule for termination-codon position within intron-containing genes: when nonsense affects RNA abundance. Trends Biochem. Sci. 23, 198–199 (1998).

33. Zeng, T. & Li, Y. I. Predicting RNA splicing from DNA sequence using Pangolin. Genome Biol. 23, 103 (2022).

34. Jaganathan, K. et al. Predicting splicing from primary sequence with deep learning. Cell 176, 535–548.e24 (2019).

35. GTEx Consortium. The GTEx Consortium atlas of genetic regulatory effects across human tissues. Science 369, 1318–1330 (2020).

36. Meier, J. et al. Language models enable zero-shot prediction of the effects of mutations on protein function. bioRxiv 29287–29303 (2021) doi:10.1101/2021.07.09.450648.

37. Cohen, J. C., Boerwinkle, E., Mosley, T. H., Jr & Hobbs, H. H. Sequence variations in PCSK9, low LDL, and protection against coronary heart disease. N. Engl. J. Med. 354, 1264–1272 (2006).

38. Cohen, J. et al. Low LDL cholesterol in individuals of African descent resulting from frequent nonsense mutations in PCSK9. Nat. Genet. 37, 161–165 (2005).

39. Thormann, A. et al. Flexible and scalable diagnostic filtering of genomic variants using G2P with Ensembl VEP. Nat. Commun. 10, 2373 (2019).

40. Saadi, I., Kuburas, A., Engle, J. J. & Russo, A. F. Dominant negative dimerization of a mutant homeodomain protein in Axenfeld-Rieger syndrome. Mol. Cell. Biol. 23, 1968–1982 (2003).

41. Veitia, R. A. Exploring the molecular etiology of dominant-negative mutations. Plant Cell 19, 3843–3851 (2007).

42. Fearon, E. R. & Dang, C. V. Cancer genetics: tumor suppressor meets oncogene. Curr. Biol. 9, R62–5 (1999).

43. Dakal, T. C. et al. Oncogenes and tumor suppressor genes: functions and roles in cancers. MedComm 5, e582 (2024).

44. Amberger, J. S., Bocchini, C. A., Schiettecatte, F., Scott, A. F. & Hamosh, A. OMIM.org: Online Mendelian Inheritance in Man (OMIM®), an online catalog of human genes and genetic disorders. Nucleic Acids Res 43, D789–98 (2015).

45. DiStefano, M. T. et al. The Gene Curation Coalition: A global effort to harmonize gene-disease evidence resources. Genet Med 24, 1732–1742 (2022).

46. Murthy, H. et al. Variants in DENND2B are associated with vulnerability for neurodevelopmental impairment, psychosis and catatonia. Brain 149, 252–261 (2026).

47. Gangloff, Y.-G. et al. Disruption of the mouse mTOR gene leads to early postimplantation lethality and prohibits embryonic stem cell development. Mol. Cell. Biol. 24, 9508–9516 (2004).

48. Blake, J. A. et al. Mouse Genome Database (MGD): Knowledgebase for mouse-human comparative biology. Nucleic Acids Res. 49, D981–D987 (2021).

49. Cao, J. et al. A human cell atlas of fetal gene expression. Science 370, eaba7721 (2020).

50. Schraiber, J. G., Spence, J. P. & Edge, M. D. Estimation of demography and mutation rates from one million haploid genomes. Am. J. Hum. Genet. 112, 2152–2166 (2025).

51. Seplyarskiy, V. et al. Hotspots of human mutation point to clonal expansions in spermatogonia. Nature 647, 429–435 (2025).

52. Neville, M. D. C. et al. Sperm sequencing reveals extensive positive selection in the male germline. Nature 647, 421–428 (2025).

53. Bernstein, N. et al. Analysis of somatic mutations in whole blood from 200,618 individuals identifies pervasive positive selection and novel drivers of clonal hematopoiesis. Nat. Genet. 56, 1147–1155 (2024).

54. Goriely, A., McVean, G. A. T., Röjmyr, M., Ingemarsson, B. & Wilkie, A. O. M. Evidence for selective advantage of pathogenic FGFR2 mutations in the male germ line. Science 301, 643–646 (2003).

55. Chen, S. et al. Impaired proteolysis of noncanonical RAS proteins drives clonal hematopoietic transformation. Cancer Discov. 12, 2434–2453 (2022).

56. El-Brolosy, M. A. et al. Genetic compensation triggered by mutant mRNA degradation. Nature 568, 193–197 (2019).

57. El-Brolosy, M. A. et al. Mechanisms linking cytoplasmic decay of translation-defective mRNA to transcriptional adaptation. Science 391, eaea1272 (2026).

58. Badonyi, M. & Marsh, J. A. Buffering of genetic dominance by allele-specific protein complex assembly. Sci. Adv. 9, eadf9845 (2023).

59. Badonyi, M. & Marsh, J. A. Proteome-scale prediction of molecular mechanisms underlying dominant genetic diseases. PLoS One 19, e0307312 (2024).

