## Supplementary Material for "Integrating 730,947 exome sequences with clinical literature improves gene discovery"

### Table of Contents


**Quality Control for gnomAD v4 9**

Data Sources and Cohort Composition 9

Supplementary Table 1 | Study diseases in gnomAD. 10

Data Processing and Alignment 11

Sample Quality Control 11

High Quality Sites Definition 11

Supplementary Figure 1 | Workflow for defining the high-quality variant set used in sample QC and ancestry inference. 13

Hard Filters 13

Supplementary Figure 2 | Data quality and processing metrics used to exclude low quality samples. 14

Supplementary Figure 3 | Hail-derived sample quality control metrics used to exclude low quality samples. 15

Supplementary Table 2 | Counts of samples removed during hard filtering. 17

Platform Inference 18

Supplementary Figure 4 | Exome platform principal components. 18

Supplementary Table 3 | Exome platform assignments. 19

Interval QC 19

Supplementary Table 4 | Intervals included in interval QC. 20

Supplementary Figure 5 | Schematic of exome padding around exome capture intervals. 21

Sex Inference per Platform 21

Supplementary Figure 6 | Sex karyotype assignment for platform 11. 23

Supplementary Table 5 | Inferred sex karyotype counts per platform. 24

Relatedness Inference 25

Supplementary Figure 7 | Pairwise relatedness inference using cuKING. 26

Supplementary Table 6 | Counts of inferred relationships among sample pairs. 27

Genetic Ancestry Group Assignment 28

Supplementary Figure 8 | Principal component analysis of genetic ancestry groups. 29

Supplementary Table 7 | Counts of inferred genetic ancestry groups. 29

Outlier Filtering 30

Supplementary Figure 9 | Sample QC metric outlier detection. 31

Supplementary Table 8 | Counts of samples excluded by outlier filters. 32

Intermediate and Final Sample Counts 32

Supplementary Table 9 | Sample counts by filtering stage. 33

Supplementary Table 10 | Final number of individuals per genetic ancestry group in gnomAD v4. 33

Variant Quality Control 34

Supplementary Figure 10 | Precision–recall performance of variant filtering thresholds. 35
Supplementary Table 11 | Counts of variants during filtering steps. 35
Genotype Quality Control 36
Variant Annotation 36
Coverage information 36
Allele Number Estimation Across All Sites 36
Supplementary Figure 11 | Differentiation between sites with no possible genotype calls vs. no alternate genotype calls. 37
Data Availability 37
Release files 38
De novo variant files 38
Code availability 38
The gnomAD browser 39
Proportion expressed across transcripts (pext) 39
Supplementary Figure 12 | The proportion expressed across tissues (pext) and tissue expression tracks on the PCSK9 gene page. 39
Stats page 39
Supplementary Figure 13 | The gnomAD Stats page. 40
Copy number variant (CNV) view 40
**Modeling expected variant counts for constraint analysis 41**
Transcript annotation and possible variant space 41
Determining observed counts 41
Calculating mutation rates 42
Determining the expected number of mutations 42
Use of allele number (AN) as a coverage metric 42
Calibrating mutation rates to the exome (plateau model) 43
Supplementary Figure 14 | Plateau models for the per context mutation rates 43
Coverage correction for low-coverage sites 43
Per-variant expected count computation 44
Regional mutation rate correction (adj_r) 44
Creation of Z scores 45
Determination of Z score cutoffs 45
**Recurrence, LOEUF, and Power Calculations 47**
Recurrence calculations 47
Generating sample SFS using analytical solution 48
LOEUF calculation 48
Power calculation 48
LOEUF estimation 49
**A fully Bayesian metric of loss-of-function constraint in population data 51**
Introduction 51

The Model 51
Model of selection 51
Model of LoF misannotation 52
Inference 53
Likelihood 55
Priors 55
Data preprocessing 56
Inference procedure 57
Robustness of the inference 58
Robustness to perturbations of the prior 59
Robustness of the sampler 59
Per-site estimation of posterior pneutral 60
**LOFTEE-2: a framework for building predictors of loss-of-function 61**
Introduction 61
Construction of LOFTEE-2 62
Stop-gained variants in multi-exon genes 62
Stop-gained variants in single-exon genes 63
Splice-affecting variants 63
Supplementary Table 12 | Strict versus relaxed filters for LOFTEE-2 63
Equations: threshold selection criterion 63
Methods compared to LOFTEE-2 64
LOFTEE 64
pneutral thresholding 64
Benchmarks of LoF prediction 64
Curated LoF variant set 65
Expression-based benchmark 65
Data 66
Background model 66
Model of NMD 67
Inference 68
Application of the benchmark 68
Gene-level constraint benchmark (LOEUF) 69
**Definition of gene lists 70**
Haploinsufficient (HI) genes associated with severe phenotypes 70
Supplementary Table 13 | Haploinsufficient genes used in this study. 70
Haploinsufficient (HI) genes associated with moderate and mild phenotypes 70
Neurodevelopmental disorder (NDD) associated genes 70
Genes with autosomal recessive (AR) inheritance 70
Olfactory receptor (OR) genes 71
Human transcription factors (TFs) 71
Kinases 71

Dimer genes 71
Helicase genes 71
Channel genes 71
Oncogenes / Tumor suppressor genes (TSG) 72
Gain of function (GoF) and Dominant Negative (DN) external list 72
Online Mendelian Inheritance in Man (OMIM) genes 72
**Combining Deleterious Missense and Loss-of-Function Constraint (Figure 4) 73**
Overview 73
Data Sources 73
Observed/Expected Ratios by Predicted Pathogenicity Percentile 74
Data Preparation 74
Reference Lines 74
Interpretation 74
Analysis of other missense effect predictors 75
Supplementary Figure 15 | Observed-to-expected (obs/exp) ratios across score percentiles for missense variants scored by missense effect predictors. 76
Enrichment of Gene Categories for Missense-over-pLoF Constraint 77
Per-Gene p-Value Computation 77
Statistical Model 77
Missense and pLoF Posteriors 78
Hypothesis and p-value 78
Interpretation 79
Synonymous Variant Filter 79
LOF-Matched Controls 79
Enrichment Calculation 79
Background Cleaning 80
Gene Categories Tested 80
Enrichment analysis using individual variant effect predictors 80
Observed and Expected Variant Counts in NDD Genes Versus All Genes 82
Variant Classes 82
Gene Filtering 83
Computation 83
Precision-Recall Curves for NDD Gene Classification 83
Constraint Metrics Compared 83
Evaluation Procedure 84
Gene Set Definition 84
Complete-Case Analysis 84
Decomposition of LOEUF-MIS by individual variant effect predictor 85
Supplementary Figure 17 | Precision-recall analysis of LOEUF-MIS decomposed by individual variant effect predictor. 85
Software and Packages 86

**PEPPER: A clinical impact score derived from an LLM pipeline** **87**
Overview 87
Literature Retrieval 87
Base PubMed Search 87
Disease-Specific Searches 88
Agent Architecture 89
Disease Association Agent (A1) 89
Penetrance Agent (A2) — Probabilistic 91
Inheritance Agent (A3) — Probabilistic 92
Mechanism Agent (A4) 93
Onset/Severity Agent — Probabilistic 95
PEPPER Calculation 98
Continuous Score Framework (v2) 98
Monte Carlo Simulation 100
PEPPER Interpretation 101
Gene-Level Aggregation 101
Gene-level PEPPER 101
Variance and Bayesian Prior 101
Mechanism-Stratified Scores 102
Implementation Details 102
Software and Dependencies 102
Reproducibility Parameters 102
Output Format 102
Citation Validation (Hallucination rate) 103
Methodology 103
Results 103
Supplementary Table 14 | Hallucination rate for agentic agents. 103
Interpretation 104
External Validation Against GenCC 104
GenCC Database Overview 104
Disease Matching Methodology 104
GenCC Disease Grouping 104
Pairwise Disease Comparison 105
Match Classification 105
Validation Metrics 105
Interpretation Considerations 106
Results Summary 107
Supplementary Figure 18 | Matching rate per GenCC confidence category. 107
Supplementary Figure 19 | Number of disease associations for each level of confidence in the agentic LLM framework. 108
Agentic Framework Results 108

Gene Statistics 108
Supplementary Figure 20 | Classification of genes by maximum evidence level. 109
Supplementary Figure 21 | Distribution of genes across disease mechanism combinations, stratified by mutation origin. 110
Supplementary Table 15 | Representative examples of genes exhibiting single and multiple disease mechanisms. 111
Gene-disease associations-level statistics 112
Distribution of gene-disease associations 112
Supplementary Figure 22 | Distribution of inheritance modes among high-confidence gene-disease associations (Definitive and Strong). 112
Supplementary Figure 23 | Distribution of penetrance categories among high-confidence gene–disease associations (Definitive and Strong). 113
Supplementary Figure 24 | Distribution of age-of-onset categories among high-confidence gene–disease associations (Definitive and Strong). 114
Supplementary Figure 25 | Distribution of disease severity among high-confidence gene–disease associations (Definitive and Strong). 115
Joint analysis of gene-diseases association categories 115
Supplementary Figure 26 | Contingency matrix of inheritance mode and molecular mechanism among high-confidence gene–disease associations. 116
Supplementary Figure 27 | Contingency matrix of age of onset and disease severity among high-confidence gene–disease associations. 117
Supplementary Figure 28 | Contingency matrix of penetrance level and age of onset among high-confidence gene–disease associations. 118
**XGBoost Feature-Based Prediction of Clinical Impact 119**
Rationale for XGBoost-Based PEPPER Prediction 119
XGBoost Model Architecture and Training 120
Feature Set 120
Supplementary Table 16 | Categories for gene-level features used in the XGBoost model 120
Model Configuration 121
Supplementary Table 17 | Hyperparameters used in the XGBoost regression. 121
Cross-Validation Strategy 121
Feature Importance Analysis 121
Global Category Importance 121
Supplementary Table 18 | XGB features SHAP analysis. 122
Top Individual Features 122
Supplementary Table 19 | Predictive features from the XGBoost. 123
Integration with Bayesian Scoring 123
Supplementary Figure 29 | OMELETXGB percentile vs LOEUF-MIS percentile. 124
Case Study: ACVR1 124
Clinical Context 124

Not Detected by Constraint Metrics 125
Supplementary Table 20 | Constraint scores for ACVR1. 125
XGBoost Feature Attribution 125
Supplementary Table 21 | Top contributing XGBoost features for ACVR1. 125
Mechanistic Insights 126
Case Study: DENND2B — Prospective Gene Discovery 126
Clinical Context 126
Pipeline Performance 127
Supplementary Table 22 | PEPPER scores for DENND2B. 127
XGBoost Feature Attribution 127
Supplementary Table 23 | XGBoost Features of DENND2B. 127
Implications for Gene Discovery 128
Candidate Disease Gene List 128
Rationale 128
Selection Criteria 128
Supplementary Table 24 | Selection criteria to define candidate disease genes using GenCC and PEPPER scores. 129
Candidate Gene Characteristics 129
Supplementary Table 25 | 221 Candidate disease genes 129
Supplementary Table 25 | LOEUF scores for candidate genes versus other genes. 129
**OMELET: A Bayesian Integration of Constraint and Clinical Significance Scores 130**
Overview 130
Model Formulation 130
Prior: Beta distribution from PEPPER 130
Adaptive kappa from Monte Carlo variance 131
Likelihood: Poisson model from LOEUF-MIS 131
Posterior 132
Summary statistic 132
Agreement and Signed Disagreement 133
Inputs and Data Sources 133
PEPPERLLM (Clinical impact score from LLM agents) 133
PEPPERXGB (Clinical Impact Score from XGBoost) 133
LOEUF-MIS 134
Implementation Details 134
**Discovery Potential (DisPo) Score: Definition, Computation, and Validation Analyses 135**
Overview 135
Mathematical Definition of the DisPo Score 135
Prior and Likelihood as Distributions over Theta 135
Centers of Mass 136
Variances 136

Signed Disagreement (DisPo) 136
DisPo Percentile 137
Relationship to OMELET 137
Validation Analyses 138
Temporal Trend in GenCC Submissions 138
Mouse Phenotype Validation 138
Tissue-Specific Expression Enrichment 139
LOEUF-Matched Design 139
GTEx Tissue Enrichment 140
Fetal Expression Enrichment 140
Fetal Tissue Expression Boxplots 141
Hyperparameters and Implementation 141
**Definitions and Applications of Key Metrics 143**
Score Summaries 143
1. LOEUF (Loss-of-Function Observed/Expected Upper Bound Fraction) 143
2. LOEUF-MIS (Loss-of-Function Observed/Expected Upper Bound Fraction incorporating Missense variants) 144
3. PEPPERLLM (Phenotype Evidence from Published Papers Extracted via Representation with Large Language Models) 144
4. PEPPERXGB (XGBoost-predicted PEPPER) 144
5. OMELETLLM (Omnibus Mutation Effects with LOEUF and Embedded Texts, literature-based) 144
6. OMELETXGB (Omnibus Mutation Effects with LOEUF and Embedded Texts, XGBoost-based) 145
7. DisPo (Discovery Potential) 145
Summary of Metrics 145
Practical Guidance: Selecting the Appropriate Metric 146
Assessing loss-of-function constraint 146
Assessing broad constraint against high-impact coding variants 146
Assessing the clinical characterization of a gene 147
Identifying understudied disease gene candidates 147
Ranking genes by overall disease relevance 149

### Quality Control for gnomAD v4

Julia Goodrich, Michael Wilson, Kristen Laricchia, Daniel Marten, Qin He, Wenhan Lu, Chris Vittal, Ben Weisburd, Charlotte Tolonen, Sam Bryant, Sam Novod, Christine Stevens, Sinead Chapman, Caroline Cusick, Laura Gauthier, Sam Lee, Jackie Goldstein, Daniel Goldstein, Tim Poterba, Dan King, Grace Tiao, Samantha Baxter, Katherine Chao, Kaitlin Samocha, Konrad Karczewski

The version of the Genome Aggregation Database (gnomAD; v4) described in this manuscript consists of 909,084,110 single nucleotide variants and insertions/deletions (indels) after variant quality control (QC) from 807,172 individuals (730,947 exomes and 76,215 genomes after QC of an initial total of 1,108,389 individuals) aligned to the GRCh38 build of the human reference genome. gnomAD v4 introduces over 600k individuals from multiple studies, including large population biobanks such as the UK Biobank[1], and increases the ancestral representation within the dataset. This increase in scale and global diversity supports more accurate allele frequency estimates and improved power to detect selective constraint.

gnomAD v4 builds upon methods used to create earlier releases of the resource, including v2[2] and v3[3]. This section of the supplement will discuss updates and improvements to the exome data processing, quality control, and analysis introduced in gnomAD v4.

#### Data Sources and Cohort Composition

A full list of contributing studies and consortia is available on the gnomAD browser (https://gnomad.broadinstitute.org/about). All samples included in gnomAD v4 were collected with appropriate institutional approvals and informed consent permitting aggregate data sharing and public release. Samples without such consent were excluded from the dataset.

Cohorts that were recruited for pediatric disease were excluded from the dataset, except for a small number of diverse cohorts where we have included unaffected relatives. Samples from biobanks were not filtered beyond removing samples that withdrew consent or technical outliers.

During sample aggregation, we collected information on the disease of interest and case/control status for a subset of the studies included in v4, allowing us to generate a high-level phenotype breakdown in gnomAD (**Supplementary Table 1**). In the past, the gnomAD project has supported disease-specific subsets (e.g., non-cancer subset). However, we have removed these subsets for two reasons: sample metadata and sample size. While we are provided high level study phenotype and case/control status for some samples, we do not have comprehensive phenotype metadata for gnomAD samples, and many samples are now derived from large biobanks, which can include individuals with disease. As such, we cannot ensure that samples in a non-disease subset do not have the specified disease. Additionally, as the dataset has grown, concerns about enrichment of any particular phenotype decreases. And finally, we continue to remove cohorts recruited for severe pediatric disease and have also removed the TCGA cancer samples due to data quality. The only subset we have released for the single nucleotide variants and indels in gnomAD v4.1 is a non-UK Biobank subset, which only impacts the gnomAD exomes and does not impact allele frequencies of the gnomAD genomes.

| Phenotypes | Case | Control | Unknown | Total | % of cases out of all v4 exomes |
|---|---|---|---|---|---|
| Alzheimer's disease | 2,594 | 665 | 1,632 | 4,890 | 0.35% |
| Atrial Fibrillation | 4,398 | 3,546 | 38,289 | 46,233 | 0.60% |
| Biobank or control dataset* | - | 24,016 | 447,750 | 471,766 | N/A |
| Bipolar disorder | 19,284 | 16,383 | 80 | 35,747 | 2.64% |
| Cardiac arrhythmia | 458 | - | - | 458 | 0.06% |
| Coronary heart disease | 1,557 | - | - | 1,557 | 0.21% |
| Inflammatory bowel disease spectrum and related disorders^ | 35,008 | 11,928 | 280 | 47,217 | 4.79% |
| Myocardial infarction | 11,900 | 369 | - | 12,269 | 1.63% |
| Neurodevelopmental** | - | 132 | - | 143 | N/A |
| Non-specific cardiovascular disease | 1,888 | 11,376 | 15,000 | 28,264 | 0.26% |
| Schizophrenia spectrum and related disorders | 30,278 | 17,689 | 39 | 47,994 | 4.14% |
| Type 2 Diabetes | 17,506 | 13,096 | 3,807 | 34,409 | 2.39% |
| **Grand Total** | **124,871** | **99,200** | **506,877** | **730,947** | **17.08%** |

* This category includes: GTEx, 1KG, UKBB, and the Qatar Genome Project, as well as the FinnGen and MGB biobank samples when no phenotype was specified
^ includes diseases like Crohn's disease, irritable bowel syndrome, interstitial cystitis, ulcerative colitis
** Neurodevelopmental controls are unaffected parents of children with confirmed or suspected de novo cause of their neurodevelopmental disorder

**Supplementary Table 1 | Study diseases in gnomAD.**
Project-provided high-level phenotype and case/control status for the gnomAD v4 exomes. "Inflammatory bowel disease spectrum and related disorders" includes diseases such as Crohn's disease, irritable bowel syndrome, interstitial cystitis, and ulcerative colitis, and the "Neurodevelopmental" controls are unaffected parents of children with a confirmed or suspected *de novo* cause of their neurodevelopmental disorder. Note that we do not have comprehensive phenotype metadata for gnomAD samples, including samples derived from large population biobanks.

### Data Processing and Alignment

Reads were aligned and processed using established pipelines, including BWA-MEM with default scoring parameters and the `-M` flag, Picard, and GATK (4.0.10.1, 4.1.4.1, or 4.1.8.0). Base quality score recalibration (BQSR) and variant calling were performed with GATK using standard parameters. The GRCh38 reference FASTA and known sites VCFs (dbSNP build 138, Mills, and 1000 Genomes) were used for alignment and recalibration.

To improve storage and computational efficiency, single-sample gVCFs were reblocked prior to merging. For non-UK Biobank samples, reblocking reduced the resolution of homozygous reference genotype quality by binning into four bands (genotype quality [GQ]20, GQ30, GQ40, GQ60) and omitting Phred-scaled Likelihoods (PLs) at homozygous reference sites, while for UK Biobank samples, GQ values were binned into seven bands (GQ0, GQ10, GQ20, GQ30, GQ40, GQ50, and GQ60). In both cases, essential annotations—including `QUALapprox`, `AS_QUALapprox`, and `RAW_GT_COUNT`—were retained to support downstream filtering and variant quality score recalibration (VQSR).

Variants were jointly combined using the Hail Variant Dataset (VDS) format, an evolution of the sparse MatrixTable introduced in gnomAD v3[3,4]. The VDS maintains a row for every locus with non-reference genotypes or reference block starts, preserves reference and non-reference genotypes separately, and enables incremental incorporation of new samples without reprocessing the entire dataset. Reference block information can be densified dynamically during analysis to reconstruct per-sample coverage and genotype confidence metrics. This combination of reblocked gVCFs and the VDS allowed efficient scaling to over 730,000 exomes while maintaining a fully lossless representation of input gVCFs and enabling accurate joint allele number estimation across all callable sites (see *Allele Number Estimation*).

### Sample Quality Control

#### High Quality Sites Definition

For specific downstream sample QC steps—including relatedness estimation and genetic ancestry inference—we defined a consistent set of high-quality autosomal single nucleotide variants (SNVs) based on existing datasets (**Supplementary Figure 1**). We restricted to SNVs located in autosomal regions present in both NHGRI Centers for Common Disease Genomics (CCDG) and UK Biobank high confidence exome intervals. High-confidence exome intervals were defined as follows: for UK Biobank exomes, intervals where at least 80% of samples achieved >20X coverage across each interval; for CCDG exomes, intervals where at least 80% of samples exceeded 10X coverage across each interval. Variants overlapping low-complexity regions (LCR) or segmental duplications (segdup) were excluded. Variants additionally required a call rate greater than 95% in the combined gnomAD v3.1.2 and CCDG genome datasets, greater than 99% in CCDG exomes, and greater than 99% in UK Biobank exomes. Variants were further filtered to remove sites with excess heterozygosity (inbreeding coefficient > -0.8) and to require a minimum allele frequency exceeding 0.01% in the combined genome datasets, as well as an allele count greater than 10 in gnomAD v3.

The v4 exome Hail VariantDataset and the v3 genome VariantDataset were each filtered to this predetermined set of high-quality sites,reformatted as dense Hail MatrixTables, and unioned to enable efficient analysis across both datasets. Callset-specific filters—including call rate > 99%, Hardy-Weinberg equilibrium $p > 10^{-8}$, and minimum allele frequency > 0.01% in the v4 joint exome/genome call set—were applied to remove low-quality sites and genotypes prior to population genetic analyses. Finally, linkage disequilibrium pruning was performed using a window size of 1,000,000 base pairs and an $r^2$ threshold of 0.1 to retain approximately independent variants. This process produced the joint QC MatrixTable containing 175,043 variants used for relatedness and genetic ancestry inference.

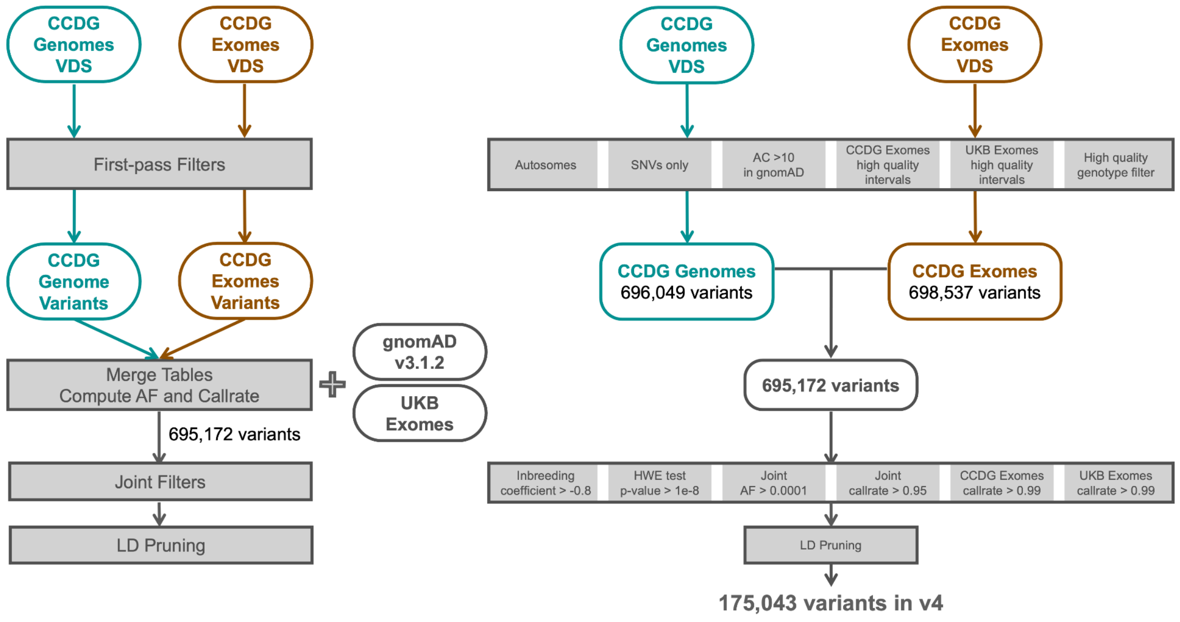


**Supplementary Figure 1 | Workflow for defining the high-quality variant set used in sample QC and ancestry inference.**
Variants were first filtered separately within the CCDG genomes and CCDG exomes Variant Datasets (VDS). For genomes, filters included restriction to autosomal, single nucleotide variants (SNVs) with allele count > 10 in gnomAD v3.1.2. For exomes, variants were required to be within high-confidence intervals in both the CCDG and UK Biobank exome datasets and to pass high-quality genotype filters. After initial filtering, 696,049 CCDG genome variants and 698,537 CCDG exome variants were retained. These were merged, and joint allele frequency and call rate metrics were computed across CCDG genomes, CCDG exomes, gnomAD v3.1.2 genomes, and UK Biobank exomes. Additional joint filters were applied, requiring inbreeding coefficient > -0.8, Hardy-Weinberg equilibrium p-value > $10^{-8}$, minimum allele frequency > 0.01% in the joint dataset, and call rate thresholds > 95% in genomes and > 99% in exomes. Finally, variants were linkage disequilibrium pruned ($r^2$ < 0.1, 1 Mb window), yielding a final set of 175,043 high-quality variants used for downstream genetic ancestry principal component analysis and relatedness estimation.

#### Hard Filters

As an initial sample identity check, we performed genotype fingerprinting using Picard CrosscheckFingerprints, which evaluates concordance using SNP haplotype blocks and LOD score-based comparisons. Fingerprints were computed from gVCF genotypes at sites defined in the GRCh38 haplotype map (*Homo_sapiens_assembly38.haplotype_database.txt*). Samples with mismatched or ambiguous fingerprint results were excluded from further processing. Thirty-one samples failed fingerprinting and were removed; most had no variant observations at fingerprinting sites in their input gVCFs and would also have been excluded by other sample QC metrics.

Samples were then evaluated for several core metrics reflecting data quality and processing artifacts. We calculated coverage across all coding intervals, restricted to sites falling within the intersection of the Broad and UK Biobank interval sets padded by 50 base pairs. Samples with mean chromosome 20 coverage below 10X were flagged for exclusion (**Supplementary Figure 2A**). We estimated contamination using CHARR[5], which calculates the mean allele balance at biallelic SNVs, and samples exceeding 1.5% estimated contamination were removed (**Supplementary Figure 2B**). Per-sample call rate was computed on the QC MatrixTable (described above), which was filtered to high-quality genotypes (GQ ≥ 20, DP ≥ 10, and allele balance ≥ 0.2 for heterozygotes; **Supplementary Figure 2C**). Samples with a high-quality genotype call rate below 80% were excluded. Chimeric read fraction was computed, and samples exceeding 5% chimeras were removed (**Supplementary Figure 2D**). No insert size filtering was applied in this release.

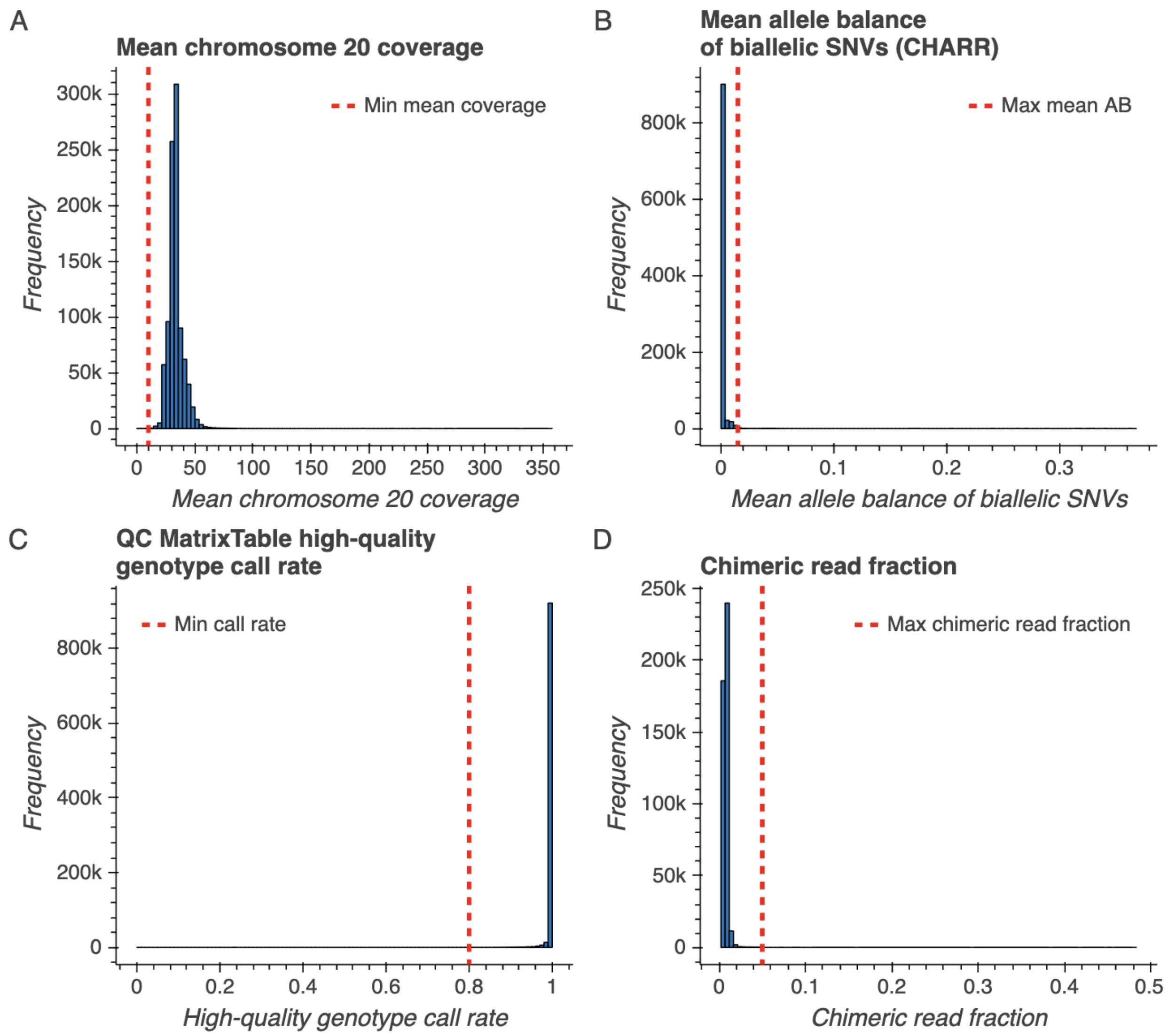


**Supplementary Figure 2 | Data quality and processing metrics used to exclude low quality samples.**

**A**, Per-sample mean coverage for chromosome 20. Coverage was computed only on chromosome 20 for cost efficiency. Samples with mean coverage less than 10X were filtered. **B**, Contamination estimates using CHARR[5]. Samples with mean allele balance larger than 0.015 for homozygous alternate genotype calls were filtered. **C**, Sample call rate calculated on high quality genotypes (GQ ≥ 20, DP ≥ 10, and allele balance ≥ 0.2 for heterozygotes) across a curated set of high quality sites (Supplementary Figure 1). Samples with call rates below 80% were filtered. **D**, Sample chimeric read fractions. Samples with chimeric read fractions larger than 5% were filtered. Red dotted lines indicate filtering thresholds in **A-D**.

Sample QC metrics were calculated on autosomal bi-allelic variants using Hail's `sample_qc` module. Samples were excluded if they were clear outliers for any of the following metrics:

- Number of singletons (> 5,000; **Supplementary Figure 3A**)
- Ratio of heterozygous to alternate homozygous genotypes (> 10; **Supplementary Figure 3B**)
- Number of bases with DP > 1X (< 5x$10^7$; **Supplementary Figure 3C**)
- Number of bases with DP > 20X (< 4x$10^7$; **Supplementary Figure 3D**)

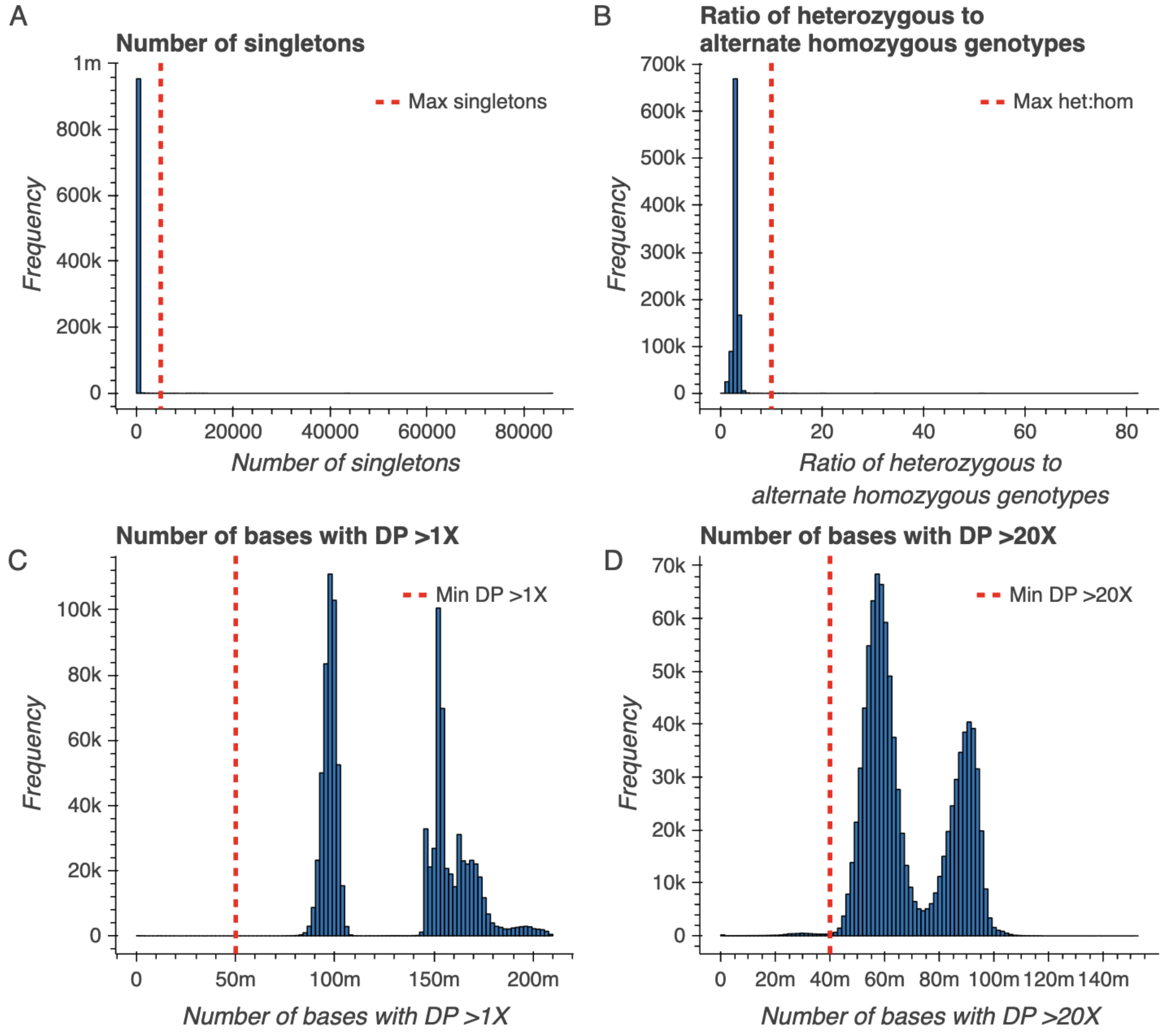


**Supplementary Figure 3 | Hail-derived sample quality control metrics used to exclude low quality samples.**

Metrics were computed using Hail's `sample_qc` module on a Variant Dataset, Hail's implementation of a scalable variant call representation[4]. **A**, The number of singleton variants per sample. Samples with over 5,000 singletons were filtered. **B**, The ratio of heterozygous to homozygous alternate (het:hom) genotype calls per sample. Samples with a het:hom ratio larger than 10 were filtered. **C**, The number of bases covered at a depth of at least 1X per sample. Samples with fewer than $5x10^7$ bases at or above a depth of 1X were filtered. **D**, The number of bases covered at a depth of at least 20X per sample. Samples with fewer than $4x10^7$ bases at or above a depth of 20X were filtered. Red dotted lines indicate filtering thresholds in **A-D**.

Hard filtering was applied in two stages. The first round, performed before platform inference, included contamination, call rate, chimeric reads, and coverage filters, but did not exclude any samples based on sex inference to avoid biasing platform clustering. After platform inference, sex chromosome ploidy was estimated separately within each platform, and samples classified as ambiguous or aneuploid karyotype (i.e., other than XX or XY) were excluded. This two-stage filtering process ensured that low-quality samples were removed while preserving robust platform and genetic ancestry inference. Counts of samples removed during hard filtering are displayed in **Supplementary Table 2**.

**Supplementary Table 2 | Counts of samples removed during hard filtering.**

The number of samples excluded due to the filters described in Supplementary Figure 1 and Supplementary Figure 2. Counts for samples consented for aggregate data release ("Releasable") are separate from counts for samples not consented for data release ("Non-releasable"). Samples excluded using Hail-derived sample QC metrics (number of singletons, ratio of heterozygous to alternate homozygous genotypes, and number of bases > 1X or 20X; Supplementary Figure 2) are collapsed into a single column ("Extreme sample QC metric outlier").

| | High contamination (6,691) | Extreme sample QC metric outlier (5,284) | Low QC MT call rate (1,079) | Low coverage (303) | Failed fingerprinting (31) | High chimeric read fraction (20) | Number of samples (955,213) |
|---|---|---|---|---|---|---|---|
| Releasable (821,465) | | | | | | | 812,624 |
| | √ | | | | | | 4,404 |
| | | √ | | | | | 3,945 |
| | | √ | √ | | | | 239 |
| | | √ | √ | √ | | | 147 |
| | | | √ | | | | 51 |
| | | √ | √ | √ | √ | | 29 |
| | √ | √ | | | | | 9 |
| | | | | | | √ | 5 |
| | | | √ | | | √ | 5 |
| | | √ | √ | | | √ | 2 |
| | √ | √ | √ | | | | 2 |
| | | √ | √ | √ | | √ | 1 |
| | √ | | √ | | | | 1 |
| | √ | √ | √ | √ | | | 1 |
| Non-releasable (133,748) | | | | | | | 130,475 |
| | √ | | | | | | 2,232 |
| | | √ | | | | | 421 |
| | | √ | √ | | | | 339 |
| | | | √ | | | | 125 |
| | | √ | √ | √ | | | 105 |
| | √ | √ | √ | √ | | | 17 |
| | √ | √ | | | | | 17 |
| | | √ | √ | | | √ | 5 |
| | √ | | √ | | | | 4 |
| | √ | √ | √ | | | | 3 |
| | | √ | √ | √ | √ | | 2 |
| | | | | | | √ | 1 |
| | | | √ | √ | | | 1 |
| | √ | | | | | √ | 1 |

#### Platform Inference

Principal component analysis (PCA) was performed on the per-sample interval call rate matrix to detect clusters corresponding to sequencing platforms and capture kits (**Supplementary Figure 4**; **Supplementary Table 3**). Interval call rates were normalized per sample and restricted to autosomal sites within the intersection of the Broad and UK Biobank interval lists with 50 base pairs of padding. The first 9 principal components were retained for clustering. Clustering was performed using HDBSCAN[6] with min_cluster_size set to 150 and min_samples set to 100. All samples were assigned to clusters based on this unsupervised clustering, which identified 20 distinct platform clusters. To validate cluster assignments, concordance rates were computed between the assigned clusters and known metadata labels for samples with available information, and assignments were reviewed for consistency with expected per-interval coverage profiles and call rate distributions. This approach allowed consistent identification of platform-specific coverage artifacts and ensured robust downstream sample QC.

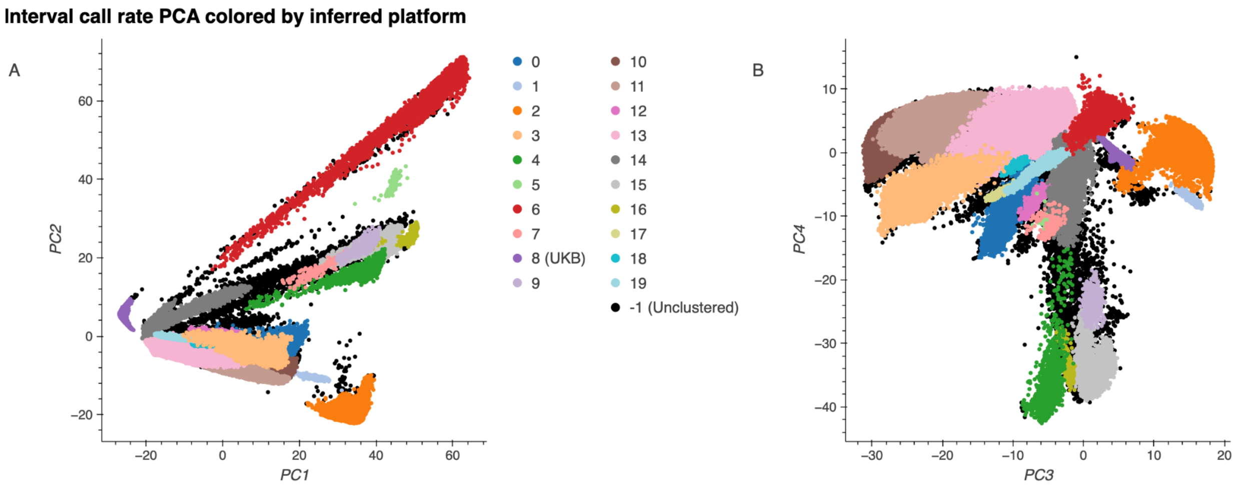


**Supplementary Figure 4 | Exome platform principal components.**
The top four platform principal components (**A,** PC1 vs. PC2 and **B,** PC3 vs. PC4) are shown. Each dot represents a single sample colored by its inferred exome platform.

**Supplementary Table 3 | Exome platform assignments.**
Sample count by platform. Samples consented for aggregate data release ("Releasable") are separate from counts for samples not consented for data release ("Non-releasable").

| Platform | Number of samples (943,099) | |
|---|---|---|
| | Releasable (812,624) | Non-releasable (130,475) |
| -1 (Unclustered) | 3,383 | 3,604 |
| -1 (Unclustered UKB) | 12 | 0 |
| 0 | 264 | 3,778 |
| 1 | 447 | 1 |
| 2 | 133,871 | 52,704 |
| 3 | 43,500 | 7,799 |
| 4 | 9,254 | 9 |
| 5 | 0 | 172 |
| 6 | 38,814 | 9,284 |
| 7 | 9 | 334 |
| 8 (UKB) | 454,469 | 0 |
| 9 | 0 | 953 |
| 10 | 29,681 | 1,756 |
| 11 | 77,531 | 39,381 |
| 12 | 183 | 469 |
| 13 | 14,602 | 181 |
| 14 | 537 | 4,146 |
| 15 | 2 | 4,448 |
| 16 | 84 | 895 |
| 17 | 409 | 0 |
| 18 | 0 | 298 |
| 19 | 5,572 | 263 |

#### Interval QC

Interval-level quality control was performed to ensure that targeted regions had adequate coverage and reliability across samples. Because the v4 callset included 416,555 UK Biobank exomes and 314,392 non-UK Biobank exomes, two distinct capture target interval lists were used: the UK Biobank target list and the Broad exome calling intervals (**Supplementary Table**

**4**). Non-UK Biobank samples were called using the Broad intervals padded by 150 base pairs, while UK Biobank samples were called using the UK Biobank intervals with the same padding. For the purpose of interval QC and consistent assessment across cohorts, intervals were defined as the intersection of these two capture sets, padded by an additional 50 base pairs (**Supplementary Figure 5**).

We empirically evaluated the impact of larger padding and the inclusion of intervals unique to one capture design, finding that these approaches reduced call rate and mean coverage, consistent with prior UK Biobank analyses[7]. For each interval, we computed per-interval metrics including call rate, mean depth, and the proportion of samples achieving mean coverage thresholds of 5X, 10X, 15X, 20X, and 25X. Intervals where at least 85% of samples achieved a mean coverage of 20X were considered high-coverage intervals and retained for some of the downstream analyses.

**Supplementary Table 4 | Intervals included in interval QC.**
Variant counts within the exome intervals from Broad and UK Biobank divided by region type (inside vs. outside exome capture or calling regions).

| | | | UKB Intervals | | | Totals |
|---|---|---|---|---|---|---|
| | | | Inside calling | | Outside calling | |
| | | | Inside capture | Outside capture | | |
| Broad Intervals | Inside calling | Inside capture | 21718832 | 10767326 | 9260124 | 41746282 |
| | | Outside capture | 169986 | 22626798 | 3979059 | 26775843 |
| | Outside calling | | 331764 | 298034 | NA | 629798 |
| Totals | | | 22220582 | 33692158 | 13239183 | 69151923 |

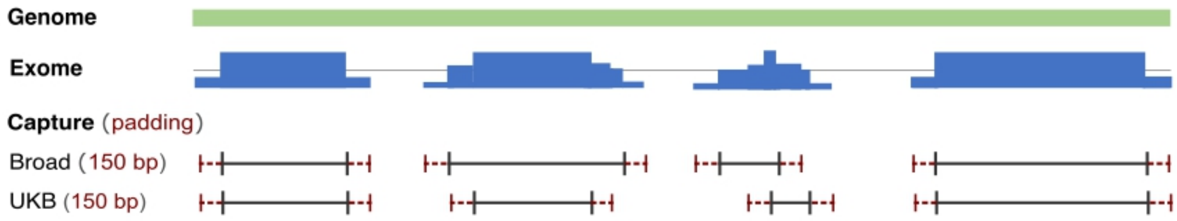


**Supplementary Figure 5 | Schematic of exome padding around exome capture intervals.** Visualization of the overlap and discrepancies between probe target regions across different capture technologies and the impact on exome coverage versus the genome. While shared target regions yield maximum sample coverage, regions uniquely captured by a single platform create localized gaps for non-target samples.

#### Sex Inference per Platform

Mean coverage was calculated across non-pseudoautosomal regions of chromosomes X and Y for each sample and normalized to the mean coverage on chromosome 20 to account for variability in sequencing depth within each exome. Chromosome X ploidy estimates were derived from mean depth across non-pseudoautosomal (PAR) variants, excluding sites within segmental duplications and low-complexity regions. Chromosome Y ploidy estimates were computed using the mean depth across all reference blocks in non-PAR regions, leveraging the completeness of reference block annotations to capture evidence of Y presence or absence even when few variant sites were observed.

To assign karyotypes, we used a two-step approach combining Gaussian mixture modeling and empirical thresholding (**Supplementary Figure 6**). Because substantial shifts in ploidy distributions persisted between sequencing platforms—even after per-sample normalization to chromosome 20—this process was performed separately within each platform to account for differences in capture design and read depth uniformity.

1. Initial clustering: For each platform, we fit Gaussian mixture models with two components to the distributions of normalized X and Y ploidy estimates. Samples were provisionally assigned to XX or XY clusters based on their component memberships, identifying the clusters with higher mean X ploidy and lower Y ploidy (XX) or lower X ploidy and higher Y ploidy (XY). For Y ploidy, this clustering distinguished samples with no detectable Y coverage from those with a single copy of Y.

2. Threshold refinement: Within each platform, we computed the mean and standard deviation of the normalized ploidy estimates for the assigned XX and XY groups. Sex chromosome ploidy cutoffs were then defined as follows:
    - Typical karyotypes: 5 standard deviations from the cluster mean
    - Aneuploidy detection: 6 standard deviations beyond the typical thresholds

These cutoffs were visually inspected and adjusted if necessary to account for batch-specific artifacts, non-normal distributions, or mosaic chromosome loss (e.g., mosaic loss of chromosome Y).

Samples falling below the upper cutoff of the single X distribution were assigned one X chromosome, while those exceeding the lower cutoff of the double X distribution were assigned two or more X chromosomes. Analogous thresholds were applied to the Y ploidy distribution to distinguish the presence or absence of Y and detect likely duplications.

For samples inferred to have a single X chromosome, we further computed the fraction of homozygous alternate genotypes across chrX (excluding low-complexity and segmental duplication regions) to detect likely XO karyotypes, which typically show elevated homozygosity due to monosomy.

Final karyotype assignments included XO, XX, XXX, XY, XXY, XXXY, XYY, XXYY, or ambiguous if ploidy estimates were inconsistent or intermediate between categories (**Supplementary Table 5**).

Unlike earlier releases, the gnomAD v4 approach did not use the Hail `impute_sex` method to derive F-statistics for primary classification but instead relied exclusively on normalized coverage-based ploidy estimation combined with per-platform Gaussian mixture modeling. This strategy enabled consistent inference across sequencing platforms and robust detection of sex chromosome aneuploidies in this heterogeneous dataset.

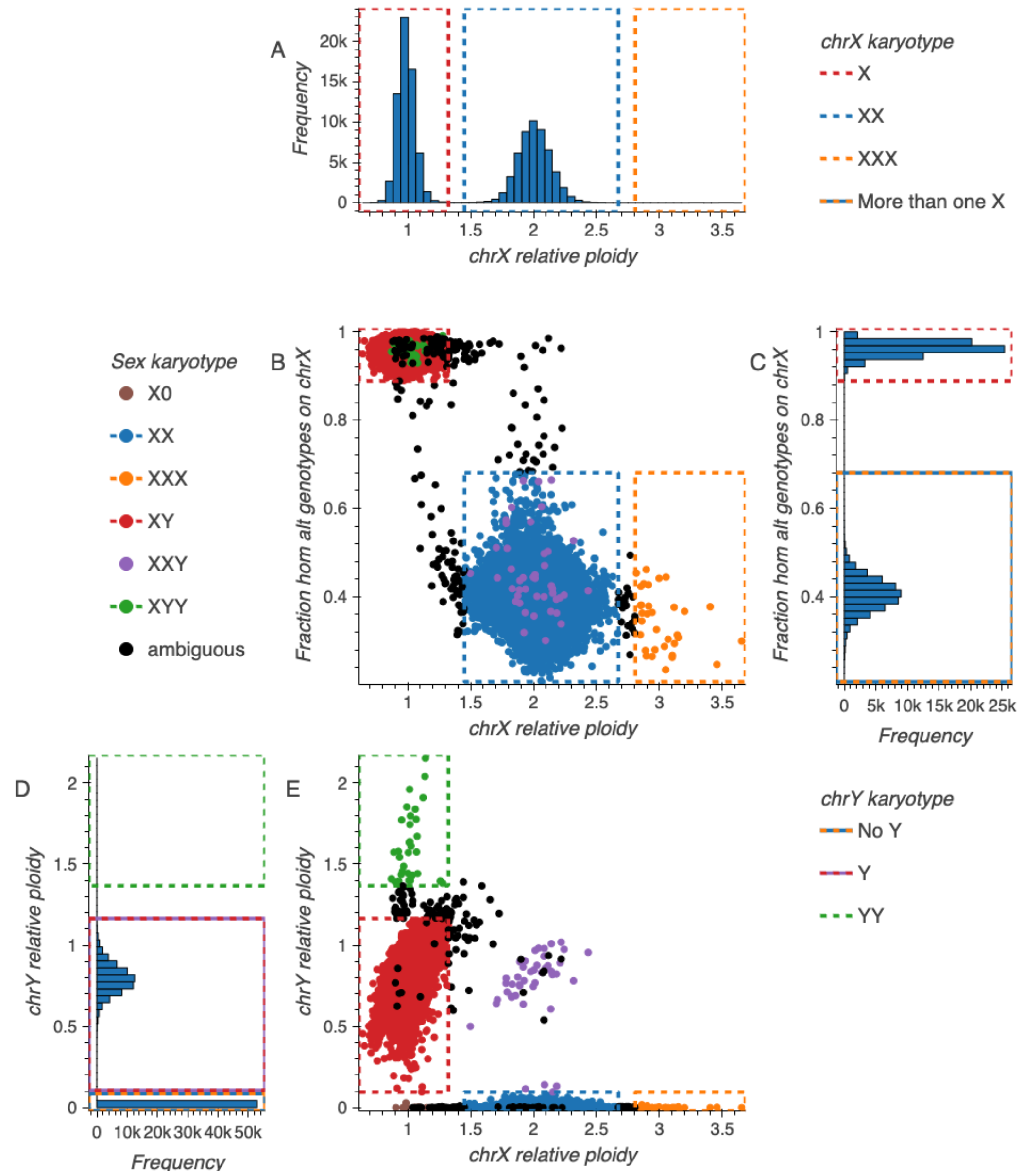


**Supplementary Figure 6 | Sex karyotype assignment for platform 11.**
**A,** Distribution of normalized chromosome X ploidy estimates across all samples from platform 11. Vertical dashed lines indicate platform-specific cutoffs delineating X (red), XX (blue), and XXX (orange) karyotype assignments. **B,** Normalized chromosome X ploidy versus fraction of homozygous alternate genotypes on chromosome X for each sample, colored by inferred sex karyotype. Dashed boxes indicate the expected regions for single X karyotypes (red for XY and green for XYY, upper region) and multi-X karyotypes (blue for XX, orange for XXX). **C,** Distribution of the fraction of homozygous alternate genotypes on chromosome X, displayed as a rotated histogram aligned with the y-axis of panel B. Horizontal dashed lines indicate platform-specific cutoffs separating single X samples (with high homozygosity) from samples carrying more than one X chromosome. **D,** Distribution of normalized chromosome Y ploidy

estimates, displayed as a rotated histogram aligned with the y-axis of panel E. Dashed boxes indicate expected ranges for no Y (blue/orange), single Y (purple/red), and double Y (green) assignments. **E,** Normalized chromosome X ploidy versus normalized chromosome Y ploidy for each sample, colored by inferred sex karyotype. Dashed boxes delineate the expected regions for XX (blue, low Y), XY (red, intermediate Y), XXX (orange, low Y/high X), and XYY (green, high Y/low X) karyotypes. Samples not falling within defined boundaries were labeled ambiguous.

**Supplementary Table 5 | Inferred sex karyotype counts per platform.**

| Platform | Number of samples | | | | | | | | |
|---|---|---|---|---|---|---|---|---|---|
| | Total | XY | XX | X0 | XXX | XXY | XYY | XXXY | Ambiguous |
| **Total** | 943,099 | 470,845 | 469,738 | 96 | 564 | 336 | 47 | 4 | 1,469 |
| **-1 (Unclassified)** | 6,999 | 3,852 | 3,044 | 0 | 0 | 3 | 1 | 0 | 99 |
| **0** | 4,042 | 2,177 | 1,854 | 0 | 1 | 3 | 1 | 0 | 6 |
| **1** | 448 | 214 | 234 | 0 | 0 | 0 | 0 | 0 | 0 |
| **2** | 186,575 | 96,482 | 89,310 | 33 | 353 | 98 | 1 | 4 | 294 |
| **3** | 51,299 | 29,084 | 22,044 | 7 | 12 | 16 | 0 | 0 | 136 |
| **4** | 9,263 | 4,361 | 4,865 | 1 | 2 | 3 | 0 | 0 | 31 |
| **5** | 172 | 107 | 63 | 0 | 0 | 0 | 0 | 0 | 2 |
| **6** | 48,098 | 27,709 | 20,237 | 3 | 13 | 26 | 0 | 0 | 110 |
| **7** | 343 | 162 | 172 | 0 | 0 | 1 | 0 | 0 | 8 |
| **8 (UKB)** | 454,469 | 207,737 | 246,142 | 34 | 115 | 101 | 0 | 0 | 340 |
| **9** | 953 | 514 | 428 | 0 | 0 | 7 | 0 | 0 | 4 |
| **10** | 31,437 | 15,578 | 15,708 | 3 | 31 | 21 | 11 | 0 | 85 |
| **11** | 116,912 | 63,750 | 52,828 | 10 | 34 | 44 | 33 | 0 | 213 |
| **12** | 652 | 314 | 332 | 2 | 0 | 0 | 0 | 0 | 4 |
| **13** | 14,783 | 11,007 | 3,733 | 2 | 2 | 3 | 0 | 0 | 36 |
| **14** | 4,683 | 2,708 | 1,929 | 0 | 0 | 2 | 0 | 0 | 44 |
| **15** | 4,450 | 2,561 | 1,861 | 1 | 0 | 1 | 0 | 0 | 26 |
| **16** | 979 | 591 | 369 | 0 | 0 | 1 | 0 | 0 | 18 |
| **17** | 409 | 183 | 223 | 0 | 0 | 1 | 0 | 0 | 2 |
| **18** | 298 | 166 | 131 | 0 | 0 | 0 | 0 | 0 | 1 |
| **19** | 5,835 | 1,588 | 4,231 | 0 | 1 | 5 | 0 | 0 | 10 |

#### Relatedness Inference

Pairwise relatedness among samples was assessed using CUDA-based KING (cuKING), a GPU-accelerated implementation of the KING algorithm that was necessary to compute relatedness metrics across more than one million exome and genome samples within practical time and memory constraints. Variants for this analysis were drawn from the joint QC MatrixTable described above, which contained a curated set of high-quality, LD-pruned autosomal SNVs selected to ensure robust estimation of genetic sharing.

Kinship coefficients were computed for all sample pairs using cuKING, and pairs exceeding a predefined threshold corresponding to first- or second-degree relationships (kinship coefficient > 0.0883) were flagged as related (**Supplementary Figure 7**). To validate these estimates and ensure consistency with prior methods, we also performed PC-Relate analysis on the same variant set, incorporating the top ancestry principal components as covariates to control for population structure. The comparison between cuKING and PC-Relate demonstrated high concordance in inferred relatedness, supporting the accuracy of the GPU-accelerated approach for this scale of data.

To identify a maximal set of unrelated individuals for downstream analyses, we applied the Hail maximal independent set algorithm. In cases where multiple samples within a related group could be retained, we prioritized (1) genome samples included in the gnomAD v3.1 release, (2) genome samples over exome samples, (3) samples with consent for the release of aggregate statistics, and (4) samples with higher mean coverage. This procedure ensured that the retained cohort represented the largest possible set of unrelated, high-quality samples for variant frequency estimation, constraint modeling, and other population genetic analyses (**Supplementary Table 6**).

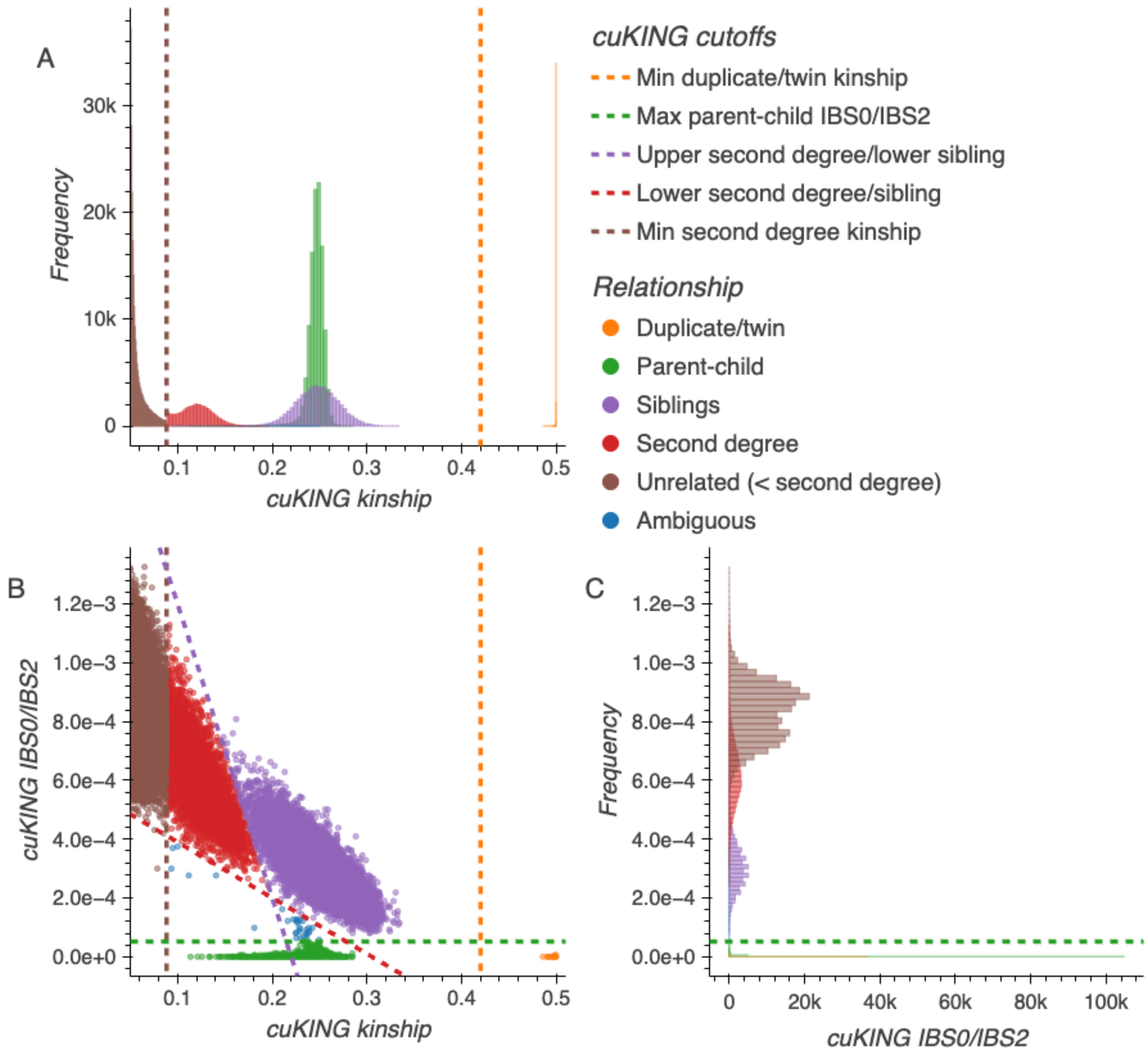


**Supplementary Figure 7 | Pairwise relatedness inference using cuKING.**
**A**, Distribution of pairwise kinship coefficients generated using cuKING with bars colored according to the inferred relationship class. The brown vertical dashed line denotes the threshold above which a pair is considered to be related, either by a first or second degree relationship, and the orange dashed line indicates the threshold above which a pair is considered to be duplicates or twins. **B,** Scatterplot of IBS0/IBS2 vs kinship coefficient, where IBS0/IBS2 is the ratio of sites with zero shared alleles to sites with two shared alleles identical by state. The dashed red and purple lines denote additional classification boundaries for determining if the pair is related by second degree or as siblings, while the dashed green line marks the maximum IBS0/IBS2 ratio consistent with parent–child relationships. **C,** Distribution of IBS0/IBS2 ratios, with parent-child and duplicate/twin pairs clustering near zero and more distant relationships showing higher ratios.

**Supplementary Table 6 | Counts of inferred relationships among sample pairs.**
Counts of pairwise relationships for releasable pairs, in which both samples are consented for aggregate data release, and non-releasable pairs, in which one or both samples are not consented for use in the aggregate release. Counts are further stratified by pair type, where "exomes" indicates both samples of the pair derive from exome data, "genomes" indicates both samples originate from genome data, and "genome-exome" indicates one sample comes from exome data and one from genome data.

| | Relationship | Pair data type | Number of pairs |
|---|---|---|---|
| **Releasable pair** | duplicate/twins (19,363) | exomes | 9,647 |
| | | genomes | 869 |
| | | genome-exome | 8,847 (7,078 in v3 release) |
| | parent-child (23,358) | exomes | 14,436 |
| | | genomes | 6,106 |
| | | genome-exome | 2,816 |
| | siblings (36,462) | exomes | 28,426 |
| | | genomes | 5,987 |
| | | genome-exome | 2,049 |
| | second degree relatives (33,493) | exomes | 20,712 |
| | | genomes | 9,281 |
| | | genome-exome | 3,500 |
| | ambiguous | exomes | 7 |
| **Non-releasable pair (at least one sample is not releasable)** | duplicate/twins (17,226) | exomes | 4,264 |
| | | genomes | 1,784 |
| | | genome-exome | 11,178 |
| | parent-child (87,255) | exomes | 39,689 |
| | | genomes | 27,913 |
| | | genome-exome | 19,653 |
| | siblings (18,804) | exomes | 5,289 |
| | | genomes | 8,757 |
| | | genome-exome | 4,758 |
| | second degree relatives (8,298) | exomes | 3,058 |
| | | genomes | 3,828 |
| | | genome-exome | 1,412 |
| | ambiguous (28) | exomes | 14 |
| | | genomes | 10 |
| | | genome-exome | 4 |

#### Genetic Ancestry Group Assignment

Genetic ancestry group was assigned using a multi-step approach combining PCA (**Supplementary Figure 8**) and supervised classification. To enable consistent genetic ancestry inference across the combined dataset, we first computed 30 principal components using unrelated samples from the gnomAD v4 joint QC MatrixTable, which included both exomes and genomes. After examining their variance explained and clustering performance, we retained the first 20 PCs for projection and genetic ancestry group classification. Related samples were then projected onto these principal components in Hail to capture the major axes of genetic variation.

We trained a random forest classifier using a diverse set of labeled samples assembled from multiple sources. Training data included:

- Human Genome Diversity Project (HGDP)[8] and 1000 Genomes Project[9] samples in gnomAD v3.1 genomes.
- All gnomAD v2 samples with genetic ancestry group labels harmonized from project-provided metadata.
- A subset of gnomAD v3.1 genomes from other projects representing African, Admixed American, Ashkenazi Jewish, East Asian, Finnish, Non-Finnish European, South Asian, and Amish groups. Cohort inclusion for training was determined by analyzing per-sample Euclidean distances to reference samples in each genetic ancestry group to ensure consistency with established genetic clusters.
- Additional gnomAD v4 samples labeled as Persian and Arab to better capture Middle Eastern ancestral diversity.

The classifier was trained using the first 20 PCs as features. To determine genetic ancestry group-specific probability thresholds for assignment, we held out 20% of labeled samples for evaluation and computed precision-recall curves. Minimum probability thresholds were inferred automatically to balance sensitivity and specificity for each genetic ancestry group, resulting in thresholds of AFR ≥ 0.93, AMR ≥ 0.86, ASJ ≥ 0.88, EAS ≥ 0.96, FIN ≥ 0.91, MID ≥ 0.56, NFE ≥ 0.78, and SAS ≥ 0.96. Samples exceeding the relevant threshold were assigned the corresponding ancestry label. Any sample that could not be confidently assigned to a single genetic ancestry group was labeled as “Remaining individuals” (**Supplementary Table 7**).

Note that the genetic ancestry groups in gnomAD were **artificially created** and are **not** naturally occurring. These group assignments are intended to summarize genetic similarity for quality control and allele frequency analyses and are **distinct** from an individual's self reported ancestry or sociocultural identity. Furthermore, the creation of these discrete genetic ancestry groups is highly dependent on the samples present in the dataset and on project-provided metadata, which is variable in availability, specificity, and collection methodology. For additional context, including a definition of genetic ancestry, a detailed discussion of why gnomAD creates genetic ancestry groups, and their limitations, see the gnomAD blog post on genetic ancestry.

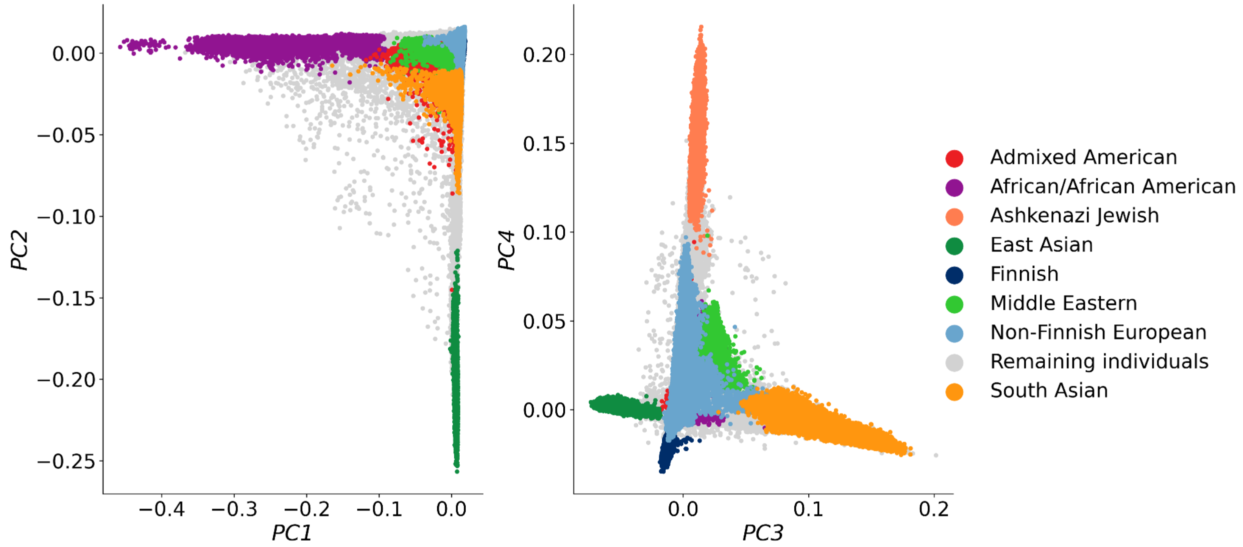


**Supplementary Figure 8 | Principal component analysis of genetic ancestry groups.**
The top four principal components are shown, with each point representing a sample colored by its inferred genetic ancestry group.

**Supplementary Table 7 | Counts of inferred genetic ancestry groups.**
All samples included in the analysis are displayed.

| Genetic Ancestry | Number of Samples |
|---|---|
| Admixed American | 29,786 |
| African/African American | 28,834 |
| Ashkenazi Jewish | 18,349 |
| East Asian | 33,574 |
| Finnish | 37,117 |
| Middle Eastern | 5,721 |
| Non-Finnish European | 690,488 |
| Remaining individuals | 41,629 |
| South Asian | 55,085 |

#### Outlier Filtering

Outlier filtering was performed using an ensemble approach designed to balance sensitivity for detecting low-quality samples with robustness to genetic ancestry- and platform-driven variation. This strategy combined two complementary methods applied to key per-sample quality control metrics: transition-to-transversion ratio, insertion-to-deletion ratio, heterozygous-to-homozygous variant ratio, and singleton transition-to-transversion (TiTv) ratio. For outlier filtering, we recomputed these metrics on all variants at sites containing two or fewer alternate alleles (i.e., biallelic or triallelic) rather than only biallelic sites. This was necessary because, at the scale of the v4 callset, the greatly increased sample size resulted in many true positive sites becoming multiallelic due to the accumulation of additional rare alleles. Restricting to only biallelic sites would have excluded a substantial fraction of real variation and reduced the sensitivity of per-sample QC metrics.

First, for each metric, QC values were stratified within each inferred exome platform cluster to control for systematic differences in sequencing performance across capture kits. Within each platform, we then fit a regression model using the top genetic ancestry principal components as covariates to capture continuous ancestry effects, rather than relying on discrete genetic ancestry strata. This sequential platform stratification and genetic ancestry adjustment allowed us to robustly model expected variation in QC metrics attributable to both technical and ancestral factors. Residuals from these regressions were standardized and used to detect samples with outlier values (**Supplementary Figure 9A**).

Second, we applied a nearest neighbors approach that compared each sample's QC metrics to the distribution of its 50 nearest neighbors in principal component space, defined using Euclidean distances computed from the same genetic ancestry PCs and restricted to samples within the same inferred exome platform cluster. This complementary method ensured that local variation in QC metrics—including subtle genetic ancestry substructure, batch-specific effects, and platform-specific differences—was accounted for in the filtering process.

Additionally, the TiTv ratio among singletons was examined as a complementary measure to assess the quality of singleton variants specifically. This metric effectively acted as a targeted proxy for excessive or low-quality singleton calls. Samples were only assessed for this metric if their singleton counts exceeded the median number of singletons in their 50 nearest neighbors or if their residual singleton counts were above the platform-specific median. Restricting its calculation to samples with sufficient singleton counts ensured that the metric was evaluated only where enough data were available to produce a stable estimate, avoiding spurious outlier calls driven by small sample size effects or the expected decline in singleton TiTv ratios as sample sizes grow.

Samples were flagged as outliers for a given metric if their value or standardized residual exceeded 4 median absolute deviations (MADs) from the median of the metric, except for the heterozygous:homozygous ratio, where only an upper threshold (> 4 MADs) was used. Importantly, a sample was only removed if it was identified as an outlier for the same metric by both the regression-based platform stratification and the nearest neighbors comparison (**Supplementary Figure 9B**). Counts of individuals removed by the outlier filters can be found in **Supplementary Table 8**.

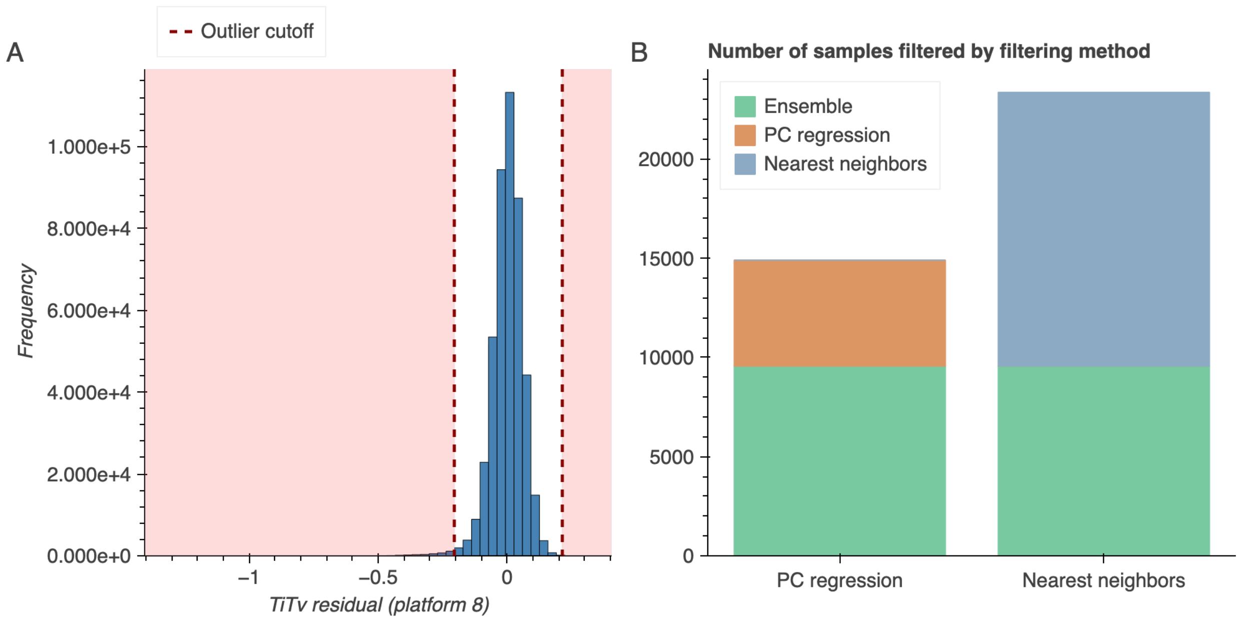


**Supplementary Figure 9 | Sample QC metric outlier detection.**
**A**, The residual of the ratio of transitions to transversions generated during platform-stratified genetic ancestry PC regression outlier filtering for inferred exome platform 8. Residuals for each metric were calculated for all platforms, but only platform 8 is shown here for clarity. Red dotted line indicates filtering thresholds (4 median absolute deviations [MADs] over or under the median). Samples with metrics falling in areas shaded in pink were considered outliers for this metric. **B**, Number of samples filtered by each sample QC metric outlier detection method. Samples filtered by platform-stratified genetic ancestry PC regression detection (PC regression) are shown in orange, by 50 nearest neighbors detection (Nearest neighbors) are shown in blue, and by both methods (Ensemble) are shown in green.

**Supplementary Table 8 | Counts of samples excluded by outlier filters.**
Counts only include samples consented for aggregate data release. Samples were excluded if their insertion to deletion, TiTv, or singleton TiTv ratios were over or under 4 MADs from the median value. Samples were also excluded if their heterozygous to homozygous alternate genotype ratio was over 4 MADs from the median value. Singleton TiTv was only calculated for samples that were identified as having more singleton variants than the median in either the platform-stratified regression or the nearest neighbor approach.

| Transition to transversion ratio (5,653) | Singleton transition to transversion ratio (3,262) | Insertion to deletion ratio (878) | Heterozygous to homozygous alternate genotype ratio (183) | Number of samples (821,465) |
|---:|---:|---:|---:|---:|
| | | | | 801,012 |
| √ | | | | 5,259 |
| | √ | | | 3,194 |
| | | √ | | 635 |
| | √ | √ | | 229 |
| √ | | | √ | 103 |
| | | | √ | 68 |
| √ | √ | | | 54 |
| | √ | √ | | 7 |
| √ | √ | | √ | 5 |
| | | √ | √ | 4 |
| √ | | √ | √ | 2 |
| | √ | | √ | 1 |
| √ | √ | √ | | 1 |

#### Intermediate and Final Sample Counts

Sample counts removed during filtering are presented in **Supplementary Table 9** and the final counts of samples, split by exome vs genome, inferred genetic ancestry group, and sex karyotype are in **Supplementary Table 10**.

**Supplementary Table 9 | Sample counts by filtering stage.**
Relatedness filters were recalculated after sample QC metric outlier detection to maximize the number of samples included in the dataset. Control samples in the dataset and additional samples missing GATK annotations required for variant quality control were removed after sample QC (Additional Release Filters).

| Filter type | Number of samples filtered (%) | Not Filtered |
|---|---:|---:|
| **Releasable** | 133,748 (14.0%) | 821,465 |
| **Hard Filters** | 10891 (1.33%) | 810,574 |
| **Outlier Filters** | 9,562 (1.18%) | 801,012 |
| **Relatedness Filters** | 66,023 (8.24%) | 734,989 |
| **All Filters** | 220,224 (23.05%) | 734,989 |
| **Additional Release Filters** | 4,042 (0.55%) | **730,947** |

**Supplementary Table 10 | Final number of individuals per genetic ancestry group in gnomAD v4.**

| | | **v4 Exomes** | | | **v4 Genomes** | | | **Combined** | | |
|---|---|---:|---:|---:|---:|---:|---:|---:|---:|---:|
| **Genetic Ancestry** | | Sample Count | XX | XY | Sample Count | XX | XY | Sample Count | XX | XY |
| Admixed American | AMR | 22,362 | 12,845 | 9,517 | 7,657 | 3,399 | 4,258 | 30,019 | 16,244 | 13,775 |
| African/African American | AFR | 16,740 | 9,663 | 7,077 | 20,805 | 11,094 | 9,711 | 37,545 | 20,757 | 16,788 |
| Ashkenazi Jewish | ASJ | 13,068 | 6,318 | 6,750 | 1,736 | 934 | 802 | 14,804 | 7,252 | 7,552 |
| Amish | AMI | - | - | - | 456 | 235 | 221 | 456 | 235 | 221 |
| East Asian | EAS | 19,850 | 10,356 | 9,494 | 2,598 | 1,136 | 1,462 | 22,448 | 11,492 | 10,956 |
| European (Finnish) | FIN | 26,710 | 13,824 | 12,886 | 5,316 | 1,287 | 4,029 | 32,026 | 15,111 | 16,915 |
| Middle Eastern | MID | 2,884 | 1,253 | 1,631 | 147 | 72 | 75 | 3,031 | 1,325 | 1,706 |
| European (non-Finnish) | NFE | 556,006 | 286,144 | 269,862 | 34,025 | 19,683 | 14,342 | 590,031 | 305,827 | 284,204 |
| Remaining | Remaining | 30,198 | 15,900 | 14,298 | 1,058 | 525 | 533 | 31,256 | 16,425 | 14,831 |
| South Asian | SAS | 43,129 | 11,020 | 32,109 | 2,417 | 577 | 1,840 | 45,546 | 11,597 | 33,949 |
| Total | | **730,947** | 367,323 | 363,624 | **76,215** | 35,543 | 33,015 | **807,162** | 406,265 | 400,897 |

#### Variant Quality Control

Variant quality control was conducted using allele-specific GATK Variant Quality Score Recalibration (AS VQSR) to robustly distinguish true variants from sequencing artifacts. SNVs and indels were modeled separately to optimize calibration across variant classes. The following variant annotations were used as features in the VQSR model:

- Quality by Depth (QD)
- Strand Odds Ratio (SOR)
- Fisher Strand Bias (FS)
- Mapping Quality (MQ, for SNVs only)
- Mapping Quality Rank Sum (MQRankSum)
- Read Position Rank Sum (ReadPosRankSum)

Positive training examples included standard resources such as HapMap, Omni, Mills, and 1000 Genomes datasets. To improve modeling of rare and singleton variation, we additionally incorporated a large set of empirical training variants derived from the gnomAD v4 callset itself, including transmitted singletons (variants observed exactly twice in the dataset and transmitted in 12,731 trios) and sibling singletons (variants present in only two siblings, derived from 26,738 siblings).

Thresholds were selected by evaluating a range of validation metrics:

- Transition-to-transversion (TiTv) ratios
- Proportions of singletons and biallelic variants
- Singleton rates and Mendelian error rates in trios
    - High-quality *de novo* SNVs per trio (see *Genotype Quality Control* for details on high-quality genotype filtering)
- Precision-recall curves from NA12878 and a synthetic pseudo-diploid sample
- Representation of ClinVar pathogenic and likely pathogenic variants

Final thresholds were chosen to retain approximately 90% of SNVs and 80% of indels, balancing sensitivity and specificity to ensure high-quality variant calls across the allele frequency spectrum (**Supplementary Figure 10**). Results were benchmarked against random forest and isolation forest filtering approaches to confirm performance.

Variants failing the selected thresholds or not meeting hard filter criteria were excluded from the final callset (**Supplementary Table 11**). Specifically, variants were removed if they showed evidence of excess heterozygosity (Inbreeding Coefficient < -0.3) or if no sample carried a high-quality genotype (AC0; see *Genotype Quality Control*).

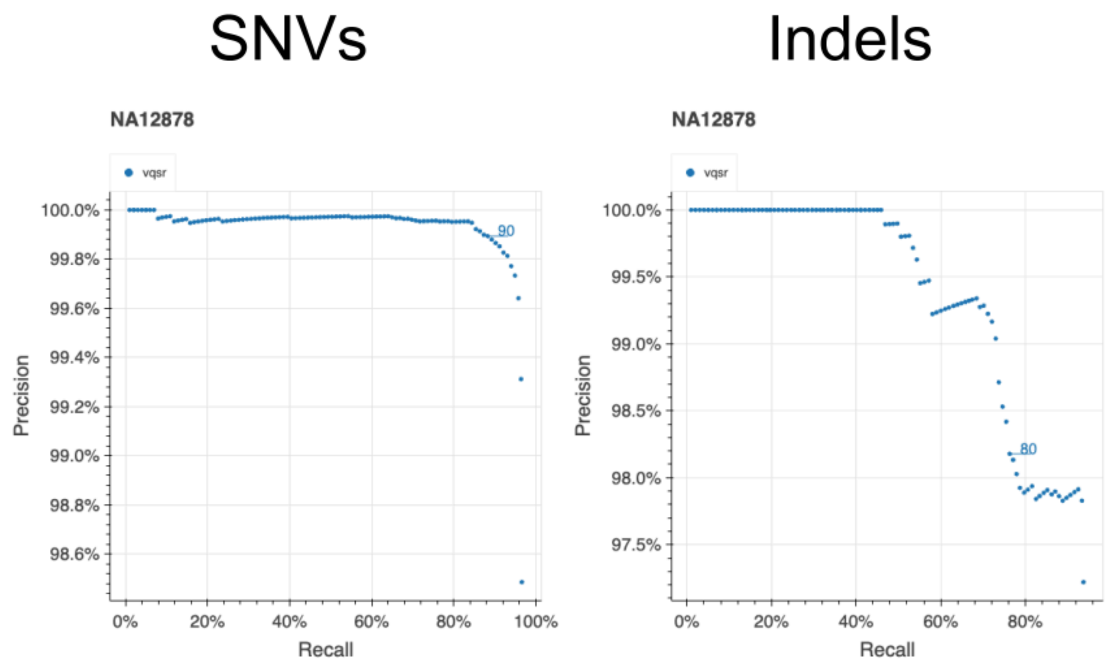


**Supplementary Figure 10 | Precision–recall performance of variant filtering thresholds.** Precision–recall curves used to evaluate variant quality score recalibration (AS-VQSR) thresholds for SNVs (left) and indels (right). Precision and recall were assessed using benchmark truth data (NA12878) across a range of filtering thresholds to evaluate the tradeoff between sensitivity and specificity. Final AS-VQSR cutoffs were selected to balance precision and recall while retaining approximately 90% of SNVs and 80% of indels in the final callset.

**Supplementary Table 11 | Counts of variants during filtering steps.**

| Filters | SNVs | Indels | Total |
|---|---:|---:|---:|
| **No Filter** | 62957594 | 6194329 | 69151923 |
| **AC0** | 79528497 | 4937154 | 84465651 |
| **AC0, AS_VQSR** | 19778677 | 3289938 | 23068615 |
| **AS_VQSR** | 5764333 | 1248137 | 7012470 |
| **InbreedingCoeff** | 4203 | 3332 | 7535 |
| **AC0, InbreedingCoeff** | 6728 | 244 | 6972 |
| **AS_VQSR, InbreedingCoeff** | 2179 | 425 | 2604 |
| **AC0, AS_VQSR, InbreedingCoeff** | 1153 | 338 | 1491 |
| **Total** | 105085770 | 9479568 | 114565338 |

#### Genotype Quality Control

Genotype quality control was applied using a standardized set of criteria (referred to as "adj" within this manuscript) to ensure that only high-confidence genotypes were retained. For diploid genotypes, calls were retained if they satisfied all of the following thresholds: sequencing depth (DP) ≥ 10, genotype quality (GQ) ≥ 20, and allele balance ≥ 0.2 for heterozygous genotypes. For haploid genotypes—specifically, non-pseudoautosomal (non-PAR) regions of chromosomes X and Y in XY individuals—a relaxed depth threshold of DP ≥ 5 was applied, and allele balance filters were not used, reflecting the expected ploidy.

#### Variant Annotation

Variant annotation was performed using Ensembl VEP version 105 configured with default parameters for the GRCh38 reference assembly. The LOFTEE plugin was applied in strict mode to flag and remove low-confidence predicted loss-of-function (pLoF) variants. Annotations were assigned relative to all available transcripts; however, unless otherwise specified, downstream analyses focused on Mane Select transcripts (version 0.95) as defined by Ensembl.

#### Coverage information

Coverage information was computed for all callable bases for the gnomAD v4 exome and genome samples using information from their gVCFs due to unavailability of read data. The summary metrics calculated were: total depth, mean, approximate median, and the percent of samples above 1X, 5X, 10X, 15X, 20X, 25X, 30X, 50X, and 100X.

#### Allele Number Estimation Across All Sites

Sample coverage information calculated from gVCFs is not as granular as information calculated from read data. In order to supplement this gVCF-derived coverage information, sample allele numbers (AN) were calculated across all callable sites in the exome and genome datasets. Unlike traditional callset processing—where allele number (AN) information is recorded only at sites with observed variation—this conversion of sample gVCFs to the Hail VariantDataset (VDS) format retains AN estimates even for loci where all samples are homozygous reference. The resulting per-site allele numbers provide precise denominators reflecting the number of samples with confidently called genotypes at each position and allows for differentiation between sites where no samples were able to be genotyped (**Supplementary Figure 11A)** vs. where zero samples had an alternate genotype call (**Supplementary Figure 11B)**.

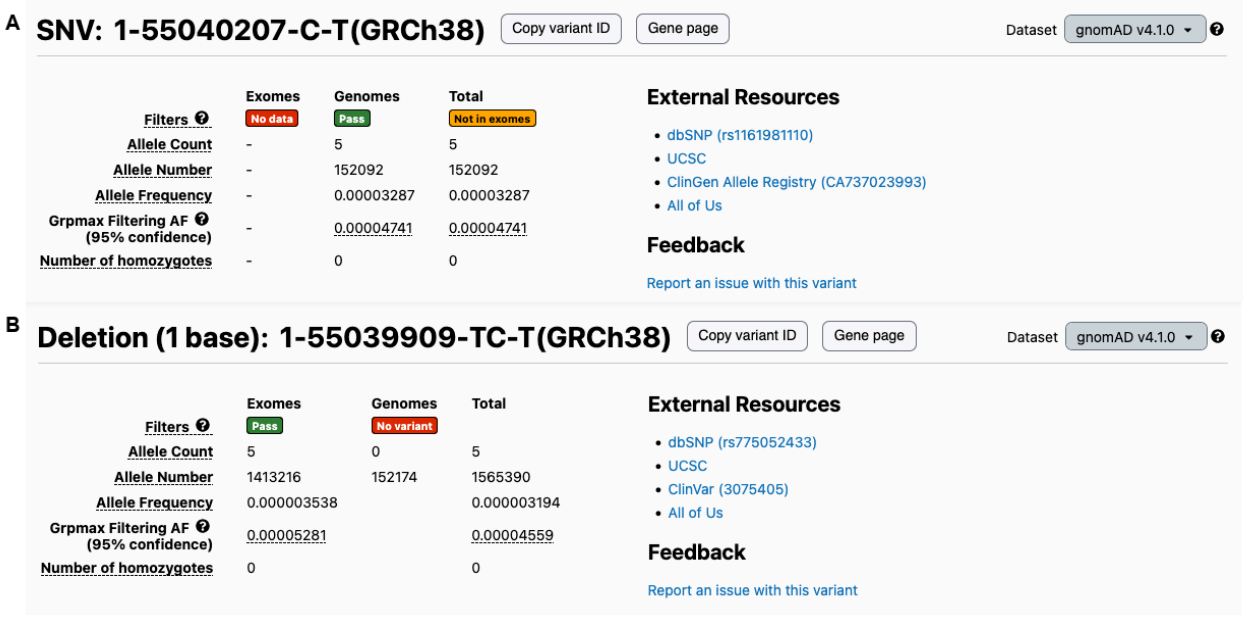


**Supplementary Figure 11 | Differentiation between sites with no possible genotype calls vs. no alternate genotype calls.**

**A**, Example variant in gnomAD v4 (https://gnomad.broadinstitute.org/variant/1-55040207-C-T?dataset=gnomad_r4) with five alleles in the genomes and zero possible alleles in the exomes (allele number is not defined in the exomes at this site as indicated with dashed line). **B**, Example variant in gnomAD v4 (https://gnomad.broadinstitute.org/variant/1-55039909-TC-T?dataset=gnomad_r4) with five alleles in the exomes and zero alternate alleles in the genomes. None of the genome samples genotyped at this site had a non-reference genotype call, as indicated by an allele count of zero and a defined allele number (AN=152,174).

These comprehensive allele counts were used as a proxy for coverage in constraint modeling, allowing more accurate estimation of the expected number of variants by accounting for variability in sequencing completeness across regions.

### Data Availability

#### Release files

Aggregate allele frequencies, functional annotations, and constraint scores derived from the variant callset can be browsed and downloaded via the gnomAD browser at https://gnomad.broadinstitute.org. All files can be accessed under the gnomAD v4 dataset page (https://gnomad.broadinstitute.org/data#v4), with detailed instructions for bulk downloads and programmatic queries provided in the browser documentation. The exome and genome datasets are available as sites-level variant call format (VCF) files as well as Hail Tables. Per-base coverage, all sites allele number, and constraint metrics are available as Hail Tables or tab-separated TSV files. Data underlying the gnomAD browser display are also available for download in Hail Table format under the gnomAD v4 dataset page (https://gnomad.broadinstitute.org/data#v4-browser-tables).

##### *De novo* variant files

In addition to the gnomAD v4 release files described above, we release 1,953 high quality coding *de novo* variants called from 1,517 trios in the gnomAD v4 exomes. We generated these calls by adapting Hail's `hl.de_novo` method (https://hail.is/docs/0.2/methods/genetics.html#hail.methods.de_novo) and Kaitlin Samocha's *de novo* caller (https://github.com/ksamocha/de_novo_scripts/tree/master). Briefly, we filtered to include only variants that were outside low-confidence regions, did not have a `*` alt allele, passed variant QC, had coding consequences, and passed gnomAD v4 exomes allele frequency and callset allele count filters. We additionally filtered to keep only variants with high and medium confidence of being true *de novos*. The observed *de novo* mutation rate per proband (~1.29 per exome) aligns with expected rates[10].

#### Code availability

All analysis scripts, pipelines, and supporting code for our quality control (QC), constraint calculation, and *de novo* variant calling pipelines are available at our GitHub repositories:
https://github.com/broadinstitute/gnomad_qc/releases/tag/v4.1
https://github.com/broadinstitute/gnomad-constraint
https://github.com/broadinstitute/gnomad_methods/releases/tag/v0.8.0

### The gnomAD browser

#### Proportion expressed across transcripts (pext)

We updated the tissue expression and proportion expressed across transcripts (pext) track on the gene and transcript pages to reflect data aligned to human genome reference GRCh38 (**Supplementary Figure 12**). The updated tissue expression track and pext metric (recalculated following the methods from Cummings *et al.*[11]) uses data from GTEx v10[12].

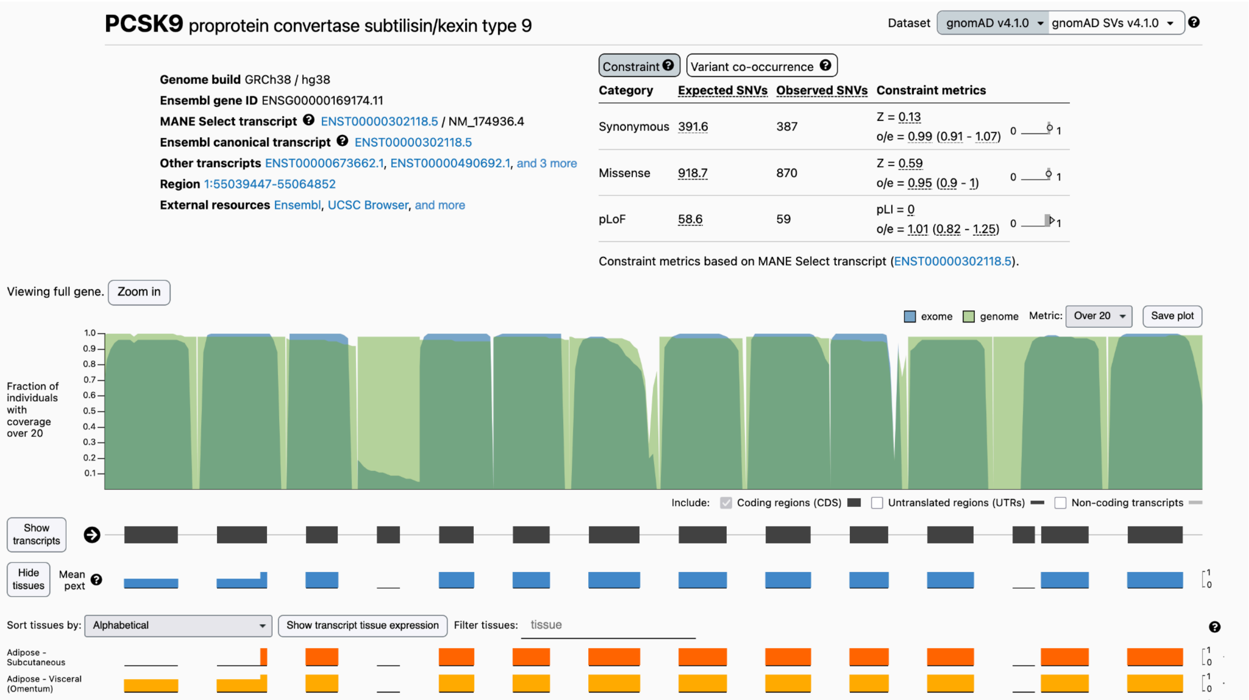


**Supplementary Figure 12 | The proportion expressed across tissues (pext) and tissue expression tracks on the *PCSK9* gene page.**

Screenshot of the *PCSK9* gene page, including the updated pext and tissue expression tracks (truncated for space) for gnomAD v4.

#### Stats page

In order to help our users better interpret the data in gnomAD v4, we added a new Stats page to the gnomAD browser (https://gnomad.broadinstitute.org/stats; **Supplementary Figure 13**). This page contains useful summary statistics on the v4 dataset (e.g., the number of samples included in the release, the total number of short and structural variants, variant type counts, etc.) and aggregate project metadata, including study-provided ancestry metadata labels and an overview of known study diseases in gnomAD (**Supplementary Table 1**).

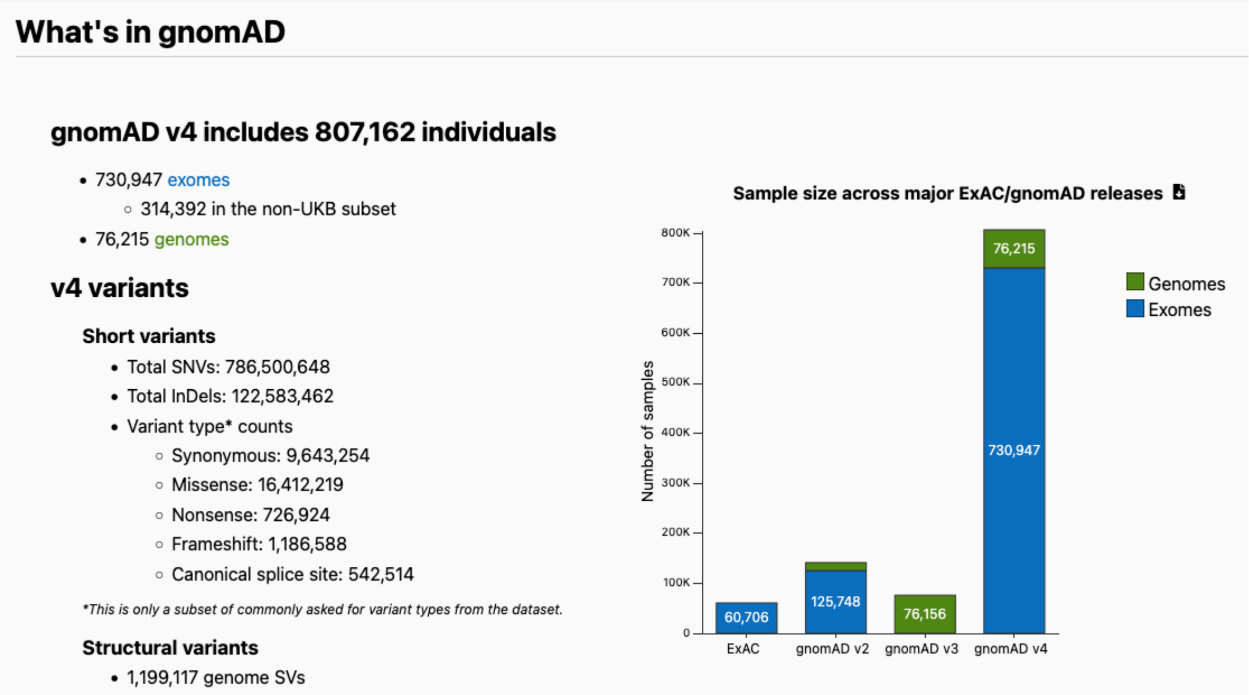


**Supplementary Figure 13 | The gnomAD Stats page.**

#### Copy number variant (CNV) view

We introduced a new browser view in gnomAD v4 for exome copy number variants, as described in our companion paper (Auwerx*, Fu* et al). This view allows users to browse rare deletions and duplications on gene and region pages in an interactive table and as tracks that are synchronized for navigation, or view details for individual CNVs on dedicated variant pages. A coverage track displays regions where CNVs can be confidently called from exome data. The table displays CNV class, size, genomic coordinate, and site frequency, with options to filter variants by class or quality. Individual variant pages display allele frequency stratified by genetic ancestry group, with sex-specific subpopulations, as well as a list of affected genes.

### Modeling expected variant counts for constraint analysis

Julia Goodrich, Ruchit Panchal, Jeremy Guez, Kristen Laricchia, Hilary Finucane, Katherine Chao, Konrad Karczewski, Kaitlin Samocha

#### Transcript annotation and possible variant space

Extending our previous methods[2], we created a dataset with every possible SNV in the human genome using the GRCh38 reference. We annotated this dataset with transcript information from Ensembl VEP version 105 (GENCODE v39) and used it as input for constraint metrics calculations. We restricted to coding sites, defining the possible variant space as every position where a coding SNV could occur, regardless of whether one was observed. A site is considered possible if it has defined exome coverage (a non-missing allele number). Sites where a variant was called but failed variant QC filters are still counted as possible, as are sites where no variant was observed. Only sites with no exome coverage or with a common variant (AF > 0.1%) are excluded from the possible count.

We generated constraint metrics for all transcripts but focused on a set of 18,411 Mane Select transcripts in this manuscript. We currently flag 1,348 MANE Select transcripts that have (1) no expected variants, (2) far too many synonymous, missense, or pLoF variants as determined by a Z-score, or (3) far too few synonymous variants as determined by a Z-score. After all outliers are removed, there are 17,063 MANE Select transcripts left for analyses.

#### Determining observed counts

The observed variant count is the number of unique single nucleotide variants observed in the gnomAD exomes with alternate allele frequency (AF) < 0.1% that pass variant quality control. Variants with AFs over 0.1% were not included; the rationale behind this choice is that, for pLoF variants where truly deleterious variants should not be common in the population, the total number of false positives far outweighs the number of true common variants.

For pLoF counts, only nonsense, splice donor, and acceptor site variants caused by single nucleotide changes and called as high confidence by LOFTEE were counted (see LOFTEE section below). This is because the mutation model does not account for insertions and deletions that underlie frameshift variants.

#### Calculating mutation rates

As before[2], we estimated baseline mutation rates from genome data to avoid coverage biases present in exome sequencing. We restricted to non-coding, putatively neutral autosomal sites by requiring the most severe VEP annotation to be “intron_variant” or “intergenic_variant”, GERP scores between the 5th and 95th genome-wide percentiles (−3.99 to 2.66), and mean gnomAD genome coverage between 15X and 60X. To avoid the saturation of methylated CpG variants that occurs at large sample sizes, we used a downsampling of 1,000 genomes. We additionally excluded sites that failed variant QC, or where the variant was found in more than 5 copies in the downsampled set.

Within this filtered set, we grouped sites by trinucleotide context, reference allele, alternate allele, and CpG methylation level (obtained from [3]). For autosomes, methylation levels ranged from 0–15, and were categorized as follows: high methylation (> 5), medium methylation (> 0 and <= 5), and low methylation (0). For chromosome X, methylation scores ranged from 0–12, and the levels were defined as follows: high methylation (> 3), medium methylation (> 0 and <= 3), and low methylation (0). For each group, we calculated the proportion of possible sites at which a variant was observed, representing the relative mutability of each substitution class. We then normalized these proportions so that the weighted genome-wide average equals the per-base, per-generation mutation rate of $1.2 \times 10^{-8}$, yielding an absolute per-context SNP mutation rate (μ_snp) for each substitution class.

#### Determining the expected number of mutations

Broadly, we followed similar methods to those in our previous work[2] and used synonymous variation to calibrate the mutation rates to scale from the gnomAD genomes (n = 76,215) to the exomes (n = 730,947).

##### Use of allele number (AN) as a coverage metric

Coverage calculated from sample genomic VCFs (gVCFs) is not as granular as coverage information from read data due to the reference block structure within gVCFs. We therefore switched from exome median read depth (used previously) to allele number (AN) as a proxy for exome coverage. Specifically, we used AN percent (the proportion of exome samples with a non-missing genotype at each position, expressed as a percentage) which provides per-base coverage resolution.

#### Calibrating mutation rates to the exome (plateau model)

We calibrated the genome-derived mutation rates to the exome dataset using synonymous variation at high-coverage sites (AN percent ≥ 90%). For each substitution, context, and methylation level, we computed the proportion of possible synonymous sites on MANE Select transcripts at which a variant was observed (using the observed and possible definitions described above).

We then fit a linear regression ("plateau model") relating the per-context mutation rate (μ_snp) to this proportion observed. Two separate regressions were fit: one for CpG transitions and one for all other sites (transversions and non-CpG transitions), as CpG sites have fundamentally different mutability characteristics (**Supplementary Figure 14**). Separate models were also fit for each genomic region (autosomes/PAR, chrX non-PAR, and chrY non-PAR).

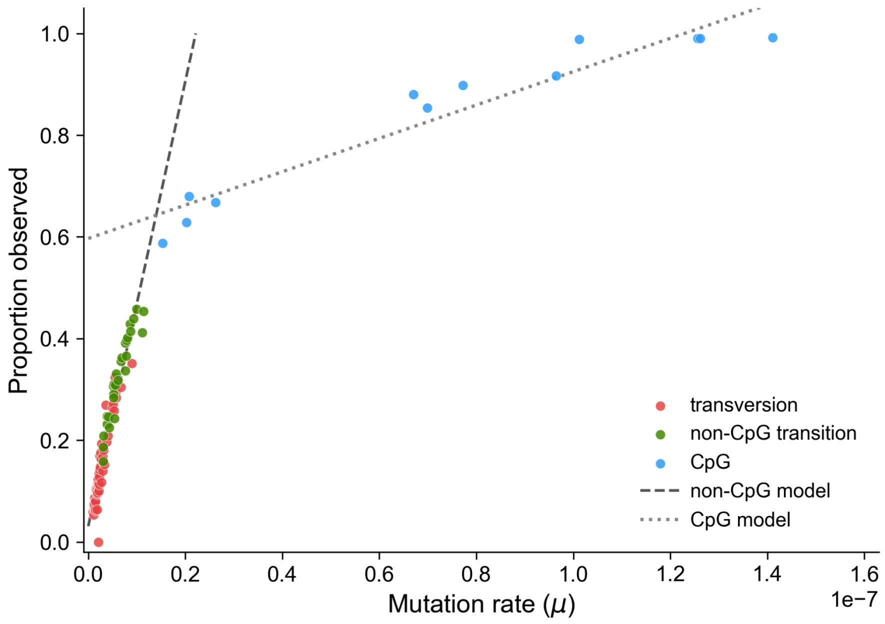


**Supplementary Figure 14 | Plateau models for the per context mutation rates**

#### Coverage correction for low-coverage sites

For sites with lower coverage (AN percent between 20% and 90%), we applied an additional coverage correction. We computed the observed/expected ratio of synonymous variation at each AN percent level, normalized by the ratio at high-coverage sites (AN percent ≥ 90%), and fit a linear regression of AN percent to this scaled ratio. This coverage correction factor was applied as a scaling factor to adjust expected variant counts at low-coverage sites. Sites with AN percent < 20% (0.58% of possible synonymous sites) were excluded from analysis entirely.

#### Per-variant expected count computation

Unlike in previous work, where the plateau and coverage models were applied to variant counts pre-aggregated by transcript, context, methylation level, and coverage bin, we applied both models at per-variant resolution. For each possible SNV in the coding genome, we looked up its μ_snp from the mutation rate table, determined its coverage correction from its site-level AN percent (as described above), and computed the per-variant expected count as:

expected_variant = plateau_model(μ_snp) × possible_variants × coverage_correction

where `plateau_model(μ_snp)` is the predicted proportion observed from the appropriate CpG or non-CpG regression for the variant's genomic region, `possible_variants` is 0 or 1, and `coverage_correction` is a scaling factor: 1.0 for high-coverage sites (AN percent ≥ 90%) and the linear model output (slope × AN_percent + intercept) for low-coverage sites (AN percent between 20% and 90%). These per-variant expected counts were then aggregated by transcript and functional consequence class (synonymous, missense, pLoF, etc.) to obtain transcript-level expected variant counts. Because both the plateau model and the coverage correction are linear, summing per-variant expected counts is mathematically equivalent to applying the models to pre-aggregated group counts, which we verified empirically (e.g., for *PCSK9* synonymous sites, both approaches yield identical expected counts). The per-variant approach has the advantage of enabling constraint assessment against any arbitrary set of variants after the fact, rather than being restricted to predefined groupings.

#### Regional mutation rate correction (adj_r)

We additionally corrected the expected number of mutations based on regional influences on mutation rate, such as replication timing, recombination rates, and known *de novo* mutation clusters, using an approach created previously[3]. Using 413,304 noncoding *de novo* mutations from two studies and 13 genomic features that influence mutation rate, we used logistic regression to identify those features that were most associated with observing a *de novo* mutation. We then used those selected features in a second regression model to determine the probability of observing a *de novo* mutation in a 1kb region versus the genome-wide average.

This ratio (adj_r) was used to scale the expected counts in each 1kb region, adjusting for the regional effects on mutation rate:

$$r_{1kb} = \frac{Predicted\ Probability\ (1kb)}{Predicted\ Probability\ (genome)} \approx \frac{exp(\beta . x_{1kb})}{exp(\beta . \bar{x})}$$

#### Creation of Z scores

As before, for each transcript and functional class (synonymous, missense, pLoF), we computed a z-score measuring the degree of depletion or enrichment of observed variants relative to expectation. We first computed a raw z-score as:

$$z\_raw = sign(obs - exp) \times \sqrt{(obs - exp)^2 / exp}$$

where obs and exp are the observed and expected variant counts, respectively. By convention, positive z-scores indicate depletion (fewer variants than expected, i.e., constraint), while negative z-scores indicate enrichment (more variants than expected).

To convert raw z-scores to standardized z-scores, we estimated the standard deviation of the raw z-score distribution separately for each functional class. For pLoF and missense variants, which are expected to be depleted relative to neutrality, we estimated the standard deviation using only the negative raw z-scores (transcripts with more variants than expected) and their mirror (the same values multiplied by −1), creating a symmetric distribution centered at zero. This mirroring approach avoids the influence of genuinely constrained genes on the standard deviation estimate. For synonymous variants, which are expected to be approximately neutral, we used all raw z-scores (without mirroring) to estimate the standard deviation.

The final z-score was computed as:

$$z = z\_raw / sd(z\_raw)$$

Transcripts with any constraint flags (z-score outliers or no expected variants; see below), undefined raw z-scores, or no observed variants across all functional classes were excluded from the standard deviation calculation.

#### Determination of Z score cutoffs

We flagged transcripts as z-score outliers using raw z-score thresholds of −8 and 8. For pLoF and missense variants, transcripts with a raw z-score below −8 were flagged (indicating far more variants than expected, suggesting mapping or annotation artifacts). For synonymous variants, transcripts were flagged if the raw z-score was below −8 or above 8 (both excess and depletion of synonymous variants can indicate artifacts). Transcripts with no expected variants were also flagged.

### Recurrence, LOEUF, and Power Calculations

Prathitha Kar, Jeremy Guez, Dmitry Biba, Misha Moldovan

#### Recurrence calculations

Under the infinite-sites model, each new mutation occurs at a unique site. The infinite sites model fits the site frequency spectrum (SFS) in population genomic data at smaller sample sizes[9]. But for current large sample sizes in gnomAD v4, mutations observed in the population could have arisen multiple times in the ancestry of the sample (recurrent mutations). Here, we describe a method to find the proportion of doubleton sites that are recurrent in the gnomAD v4 dataset (**Figure 1D**).

To avoid confounding due to selection, we restrict to synonymous sites, which may be assumed to be selectively neutral. Let the number of singletons be $C_1$ and doubletons be $C_{2,1}$ in the infinite sites limit. Let $O_1$ and $O_2$ be the observed number of singletons and doubletons.

We define $r = \frac{O_2}{O_1}$, ratio of proportion of doubletons to singletons. $r$ remains constant in the infinite sites limit regardless of sample size and mutation rate. Thus, at small sample sizes where the infinite sites limit holds, $r_0 = \frac{C_{2,1}}{C_1} = constant$. However, recurrence leads to a surplus of doubletons, thus, making $r > r_0$. A recurrent doubleton arises when two independent mutations occur at the same genomic position (number of such sites = $C_{2,2}$). If these mutational events had occurred at different sites, each would have been observed as a singleton. Thus, recurrence effectively converts two potential singletons into a single doubleton. Formally, $O_1 = C_1 - 2C_{2,2}$, and $O_2 = C_{2,1} + C_{2,2}$. The desired quantity which is the level of recurrence is defined to be $\frac{C_{2,2}}{O_2}$. We rewrite $O_2$ in terms of $r_0$, $C_1$ and $C_{2,2}$ as $O_2 = r_0 C_1 + C_{2,2}$. Using $r = \frac{O_2}{O_1}$, we find the $\frac{C_{2,2}}{C_1} = \frac{r - r_0}{1+2r}$. Thus, the empirical level of recurrence in doubletons is,

$$Recurrence = \frac{C_{2,2}}{O_2} = \frac{r - r_o}{r(1+2r_0)}$$

We calculate $r^*$ at sample size = 1000.

### Generating sample SFS using analytical solution

To generate the probability of allele count = $k$ from a sample of size $n$, we use an analytical solution of the sample site frequency spectrum (SFS) for rare variants[13]. The solution is obtained for a Binomial sample from a population following the Wright-Fisher model. The model has three parameters corresponding to the three different evolutionary forces – mutation rate $\mu$, selection coefficient $s$, and demography $N(t)$. We fix the demography to be for Non-Finnish Europeans (NFE) using previous studies[13]. The NFE population goes through a bottleneck with recent rapid expansion in effective population size. We allow for recurrent mutations to arise in the population. Dominant selection is assumed, thus, in the rare variant limit, selection is against heterozygous variants.

### LOEUF calculation

For any combination of mutation rate ($\mu$), selection ($s$), and sample size ($n$), we can calculate the probability of monomorphic site ($p(k = 0|\mu,\ s,\ n)$) using the analytical solution of the sample SFS. For a fixed target site and mutation rate, LOEUF as a function of $s$ is equivalent to ratio of probability of segregation at selection coefficient $s$ and probability of segregation at $s = 0$. To estimate LOEUF as a function of $s$ in Figure S2, we calculate $\frac{1-p(k=0|\mu, s, n)}{1-p(k=0|\mu,\ s=0,n)}$.

### Power calculation

We estimate the power of LOEUF to distinguish between two selection coefficients $s_{null}$ and $s_{alt}$. Power is the probability to reject the null hypothesis that selection is $s_{null}$ at a given significance level ($\alpha = 0.05$) provided the alternate hypothesis that selection $s_{alt}$ is true. For fixed target sites and mutation rate, LOEUF depends on the number of observed variants in those sites and can be determined, equivalently, from the number of monomorphic sites (= number of possible variants - number of observed variants). Thus, to obtain the power of LOEUF to distinguish $s_{null}$ from $s_{alt}$, we compare the distribution of number of monomorphic sites ($K$) under the null and alternate models.

We fix the number of target sites to be $n_t$. Let $p_{null}$ and $p_{alt}$ be the probability of non-segregation for selection $s_{null}$ and $s_{alt}$, respectively, obtained using the analytical solution of sample SFS. Under this formulation, the number of monomorphic sites follows a binomial distribution with success probability $p_{null}$ and $p_{alt}$ for the null and alternate hypothesis, respectively. To account for the discreteness of the number of variants, we approximate the binomial distribution by a continuous Beta distribution, which is the conjugate prior of the binomial distribution. The random variable of the Beta distribution $\hat{p} = \frac{K}{n_t}$ and $\hat{p} \sim Beta(n_t p + 1,\ n_t(1 - p) + 1)$, where $p$ is the probability of non-segregation. We find the critical threshold $(c_{1-\alpha})$ under $s_{null}$ at $\alpha = 0.05$ using the $Beta(\hat{p}_{null}|n_t p_{null} + 1,\ n_t\left(1 - p_{null}\right) + 1)$ distribution. Power is calculated to be the probability that $\hat{p}_{alt} > c_{1-\alpha}$ where $\hat{p}_{alt} \sim Beta(n_t p_{alt} + 1,\ n_t\left(1 - p_{alt}\right) + 1)$. Thus, Power = $1 - F_{Beta(\hat{p}_{alt}|n_t p_{alt} + 1,\, n_t\left(1 - p_{alt}\right) + 1)}(c_{1-\alpha})$, where $F_{Beta(\alpha,\,\beta)}(x)$ is the cumulative distribution of $Beta(X|\alpha, \beta)$ evaluated at $x$.

#### LOEUF estimation

We use LOEUF as a measure of the gene-level constraint against loss-of-function variants. We assume that all loss-of-function variants within a gene have the same effect. LOEUF is obtained by comparing the observed number of segregating loss-of-function variants within a gene against the expected from synonymous sites. If the number of observed variants is much lower than expected, the gene is under higher selective constraint. Below we describe the procedure to calculate LOEUF.

Assume a gene with a fixed number of $n$ target sites. Each site $i$ has a certain probability of segregation $p_i$ determined by the mutation rate $(\mu_i)$ and selection intensity $(s_i)$. For loss of function variants, we assume that all variants within the gene have same $s_i = s$. Additionally, we neglect errors in mutation rate. The number of observed variants in the gene $(O)$ is,

$$O = \sum_{i=1}^{n} \xi_i, \qquad 1$$

where $\xi_i$ is Bernoulli distributed with probability of success $p_i$. For strong selection (small $p_i$), we approximate the sum of Bernoulli random variables with Poisson distribution, thus, $O \sim Poisson(\lambda = \sum_{i=1}^{n} p_i)$. We aim to obtain the distribution of $\sum_{i=1}^{n} p_i$ i.e. the expected number of observed variants given we observe $O$ loss-of-function variants segregating in the gene.

More formally, we want to calculate the posterior distribution of $P(\sum_{i=1}^{n} p_i | O) = \frac{P(O| \sum_{i=1}^{n} p_i) P(\sum_{i=1}^{n} p_i)}{P(O)}$. Assuming prior $P\left(\sum_{i=1}^{n} p_i\right) \propto constant$ and using $O \sim Poisson\left(\lambda = \sum_{i=1}^{n} p_i\right)$, we obtain $P\left(\sum_{i=1}^{n} p_i | O\right) \sim Gamma(shape = O + 1, scale = 1)$. LOEUF is defined as a summary of this posterior distribution and equals the 95% upper bound of this distribution divided by the expected number of segregating variants assuming the sites are neutral.

### A fully Bayesian metric of loss-of-function constraint in population data

Mikhail A. Moldovan, Jeremy Guez, Julia Goodrich, Prathitha Kar, Kaitlin Samocha, Konrad Karczewski, Evan Koch

#### Introduction

Population frequencies of loss-of-function (LoF) variants in protein-coding genes reflect selective constraint against gene inactivation and, consequently, inform our understanding of gene function[2,14]. The availability of large-scale human variation resources has enabled population-genetic metrics of LoF intolerance (e.g., pLI, LOEUF) and selection against heterozygous LoF variants ($s_{het}$), which are widely used to summarize the phenotypic impact of LoF mutations.

Here we extend prior work on LoF constraint[15] by formulating a fully Bayesian model for estimating $s_{het}$ from population data. The framework provides coherent uncertainty quantification for all parameters and explicitly accounts for potential LoF misannotation. By modeling misannotation as a function of allele frequency, the approach yields a frequency-dependent measure of annotation accuracy, which we use to derive predictors of LoF variant pathogenicity.

#### The Model

##### Model of selection

Let us assume:

1. A genetic locus with two possible alleles, where $A_1$ denotes the ancestral, and $A_2$ – the derived allele. We let *x* be the frequency of $A_2$;
2. A diploid Wright-Fisher population of size $N_e$;
3. $A_1$ mutates to $A_2$ with rate μ;
4. Fitness of the $A_1A_1$ individuals to be 1, $A_1A_2$ individuals $1 - s_{het}$ and $A_2A_2$ individuals – $1 - s_{hom}$, where $s_{het}$ denotes selection against heterozygotes and $s_{hom}$ – selection against homozygotes;

5. $s_{het} > 0$, and, moreover, $s_{het} \gg 1/N_e$ (strong negative selection).

Under these assumptions, it generally holds that $x \ll 1$ and $x^2 \approx 0$, rendering the contribution of selection against $A_2A_2$ homozygotes and $A_2$-to-$A_1$ mutations to the distribution $\varphi(x)$ negligible. Thus, the Nei approximation to variant frequency distribution $\varphi(x)$ applies[16]. Specifically, $x \sim \text{Gamma}(x \mid k, \theta)$, where the shape parameter $k = 4N_e\mu$ and the scale parameter $\theta = 1/(4N_e s_{het})$. If a sample of size $D < N_e$ is taken from the population, an estimate of the probability density of the distribution of derived allele ($A_2$) counts $n$ may be obtained as:

$$\begin{aligned}\hat{f}_s(n|D, N_e, \mu, \mathrm{s_{het}}) &= \int_0^{\infty} \mathrm{Pois}(n|Dx)\mathrm{Gamma}\left(x|4N_e\mu, \frac{1}{4N_e\,\mathrm{s_{het}}}\right)\, dx \\ &= \mathrm{NegBinom}\left(n|k = 4N_e\mu, \theta = \frac{D}{4N_e\,\mathrm{s_{het}}}\right), \end{aligned} \tag{1}$$

where Pois(·|·), Gamma(·|·) and NegBinom(·|·) denote probability densities of Poisson, Gamma and Negative Binomial distributions. The Negative Binomial distribution is defined with respect to the shape and scale parameters.

Note that this approximation assumes a finite-sites model as an approximation to the full Wright equation, and hence may be applied to the gnomAD v4 dataset, where a certain degree of recurrence is expected, especially in highly mutable contexts.

#### Model of LoF misannotation

Prior studies have shown that a substantial (25–30%) proportion of variants with VEP consequences suggestive of loss-of-function (stop gained, splice donor variant and splice acceptor variant) do not result in an effective loss of a gene copy, due to various reasons including the absence of triggering of nonsense-mediated decay, technical issues with annotation of functional exons, annotation of splice variants and others[17]. From the standpoint of $\varphi(x)$, it means that any incoming $x$ is by necessity sampled either from the distribution of “true” LoF frequencies, $\varphi_s(x)$, or from the “contaminating” distribution $\varphi_0(x)$. Let us denote the fraction of frequencies sampled the contaminating distribution as $p_{neutral}$ and note that, due to the linear properties of integration, the same applies to the distribution of counts of $A_2$ sampled from the population (Eq. 1):

$$\begin{aligned}\phi(x) &= (1 - \mathrm{p_{neutral}})\phi_s(x) + \mathrm{p_{neutral}}\,\phi_0(x), \\ f(n) &= (1 - \mathrm{p_{neutral}})f_s(n) + \mathrm{p_{neutral}}\,f_0(n),\end{aligned} \tag{2}$$

where $f_s(n)$ and $f_0(n)$ denote the true and contaminating distributions of counts n.

Following Zeng *et al.*, 2024[15], we assume the contaminating mutations to be neutral. And, rather than assuming $\varphi_0(x)$, we directly estimate the genome-wide distribution of counts $f_0(n)$ from the frequencies of synonymous variants observed in gnomAD v4. And, because $f_0(n)$ depends on the mutation rate, we estimate an array of $f_0(n|\hat{\mu})$ with respect to the estimates of μ given by the Roulette mutation rate model[18].

Specifically, we:

1. Calculate the values of $f_0(n|\hat{\mu})$ for each Roulette mutation rate bin $\hat{\mu}$ from the counts of observed values of n conditional on $\hat{\mu}$ at synonymous sites.
2. To stabilize sparse bins, merge low-count bins. In practice (with 100 ordered bins), we merge the following ranges: 1–2, 30–34, 35–39, 40–44, 45–49, 50–54, 55–59, 60–64, 65–69, 70–74, 75–79, and 98–100.
3. Apply sliding-window smoothing to the resulting estimates of $\hat{f}_0(n|\hat{\mu})$. To both capture the fine properties of $f_0(n)$ at low values of n and account for the sparsity of large n-s, we make window sizes dependent on n as $\lfloor n/(20\times\log_{10}(n+1)) + 1\rfloor$.

Finally, following Zeng *et al.*, we account for the possibility of dependence of $p_{neutral}$ on specific annotation by introducing another condition a ∈ {“stop gained”, “splice donor variant”, “splice acceptor variant”}.

Thus, for a single count n, our estimate of f(n) in Eq. 2 may be written as:

$$\hat{f}(n|D, N_e, \hat{\mu}, \mathrm{s_{het}}, a) = (1 - \mathrm{p_{neutral}}_a)\hat{f}_s(n|D, N_e, \hat{\mu}, \mathrm{s_{het}}) + \mathrm{p_{neutral}}_a\, \hat{f}_0(n|\hat{\mu}). \quad (3)$$

### Inference

We assume that:

1. All of the possible “true” LoF variants within the gene incur the same heterozygote disadvantage $s_{het}$;
2. The frequencies of misannotated variants, conditional on $\hat{\mu}$, are sampled from the same distribution $\hat{f}_0$;
3. The values of $p_{neutral,a}$ do not vary along the genome;

4. Effective population size $N_e$ as well as the effective sample size $D$ do not vary along the genome.

We also note that, since the Roulette estimates $\hat{\mu}$ are defined up to a fixed constant $C_R$, we need to account for it in our parameter inference. Let $\hat{\mu} \approx C_R\mu$, where μ is the true mutation rate. Then, the parameters in Eq. (1) are:

$$k = \frac{4N_e\hat{\mu}}{C_R}, \qquad (4a)$$

$$\theta = \frac{D}{4N_e\,\mathrm{s_{het}}}. \qquad (4b)$$

Note that, from the standpoint of inference in the real population, there are issues with the identifiability of scaling parameters: scaling of $s_{het}$ is determined by the ratio $D/N_e$ and scaling of $\hat{\mu}$ – by the ratio $N_e/C_R$. To account for this, we re-parametrize the Negative Binomial distribution in Eq. (1) with the mean factor (M, mean of the Negative Binomial) and the dispersion factor (*k*, shape). From Eq. 4 and the properties of the Negative Binomial distribution, we have:

$$\begin{aligned} k &= \frac{4N_e\hat{\mu}}{C_R} \\ &:= \mathrm{N_{fac}}\,\hat{\mu}, \qquad (5a) \\ M &= k\theta \\ &= \frac{4N_e\hat{\mu}}{C_R}\frac{D}{4N_e\,\mathrm{s_{het}}} \qquad (5b) \\ &:= \frac{\hat{\mu}}{\mathrm{S_{fac}}}, \end{aligned}$$

where the N-factor $N_{fac}$ and the s-factor $S_{fac}$ absorb the four unknown constants (mutation $N_e$, selection $N_e$, effective sample size $D$ and Roulette scaling $C_R$). Also note that, since we do not model μ explicitly, inaccuracies of $\hat{\mu}$ would propagate into the inferences of both $N_{fac}$ and $S_{fac}$. We obtain the $s_{het}$ values by scaling the raw $S_{fac}$ in a two-step inference procedure (see below).

Let the total count of admitted genes be G, then the total vector of parameters estimated genome-wide is: $\Theta = \{S_{fac1}, \ldots, S_{facG}, p_{neutral,1}, p_{neutral,2}, p_{neutral,3}, N_{fac}\}$, where $p_{neutral\alpha}$, $\alpha \in [1, 2, 3]$ are the $p_{neutral}$ values for the three LoF annotations.

#### Likelihood

Let *i* be the index of individual pLoF variants in the genome, $g_i$ – the index of gene containing *i*-th variant. Then, with the assumption that all sites are independent conditional on parameters, the full likelihood for parameters Θ with respect to the incoming vector of derived allele frequencies $\{n_i\}$ over all the admitted pLoF variants within the genome is:

$$\begin{aligned}\mathcal{L}(\Theta|\{n_i\}) &= \prod_i \hat{f}(n_i|\hat{\mu}_i, a_i, \mathrm{S_{fac}}_{g_i}\,\mathrm{N_{fac}}) \\ &= \prod_i \left[(1-\mathrm{p_{neutral}}_{a_i})\mathrm{NegBinom}\left(n_i|k=\mathrm{N_{fac}}\,\hat{\mu}_i, M=\frac{\hat{\mu}_i}{\mathrm{S_{fac}}_{g_i}}\right)+\mathrm{p_{neutral}}_{a_i}\,\hat{f}_0(n_i|\hat{\mu}_i)\right].\end{aligned} \tag{6}$$

#### Priors

- **$N_{fac}$**. Because the Roulette $\hat{\mu}$ values aim to capture the population–scaled mutation rate $2N_e\mu$, we expect $N_{fac} \approx 2$. To account for uncertainty around this value, we assume $N_{fac}$ is generally of order 1 and place a base-10 log-Normal prior:

$$\log_{10}\mathrm{N_{fac}} \sim \mathcal{N}(0,\, 0.5^2).$$

This prior peaks at 1 on the original scale and spans one decimal order of magnitude between the inflection points of the density. Since $N_{fac}$ is a genome-wide parameter, its inference is largely data-dominated.

- **$p_{neutral}$**. Previous studies indicate that roughly 15–30% of pLoF variants are misannotated[17]. To capture uncertainty around these values, we use the same Beta prior for all three inferred $p_{neutral}$ parameters:

$$\mathrm{p_{neutral}} \sim \mathrm{Beta}(10,\, 40).$$

This prior choice provides robustness of the inference (see below) while allowing the data to shift the estimates substantially. As with $N_{fac}$, $p_{neutral}$ is genome-wide, so posterior inference is dominated by the likelihood under the assumed model.

- **$S_{fac}$**. Our priors for $S_{fac}$ values draw information from the mean prior estimates of the selection coefficient against heterozygotes, produced by Zeng *et al.*. These estimates are obtained using a set of global gene features that should be minimally impacted by variation in any one gene, providing a source of information orthogonal to the segregation in gnomAD v4. However, the $S_{fac}$ values are computed on a different scale (Eq. 5). To take this systematic bias into account, we performed two runs of the procedure: (i) the seed run with a global weakly informative prior for $S_{fac}$ values to determine the scaling coefficient $\hat{\alpha}$ and (ii) the run informed by priors drawing from Zeng *et al.* on $\hat{\alpha}\, S_{fac}$ values:

1. **Seed run on chromosome 21.** We assume extremely non-informative priors:

$$\mathrm{S_{fac}} \sim \mathrm{Cauchy}_{+}(0,\, 5),$$

where $\mathrm{Cauchy}_{+}$ denotes the Cauchy distribution restricted to $\mathbb{R}^{+}$.

2. **Genome-wide run.** After the seed run, we obtain an estimate $\hat{\alpha}$ of the scaling of $S_{fac}$ values relative to the mean prior $s_{het}$ estimates $s^0_g$ from Zeng *et al.*. For each gene g, we then use:

$$\log_{10}(\alpha\, \mathrm{S_{fac}}_g) \sim \left(1 - \Pr[\alpha\, \mathrm{S_{fac}}_g > 1]\right) \mathcal{N}\!\left(s^0_g,\, 0.5^2\right) \;+\; \Pr[\alpha\, \mathrm{S_{fac}}_g > 1]\; \delta_1, \quad (7)$$

i.e., a Log-Normal-on-base-10 prior centered at $s^0_g$ that is truncated with a Kronecker delta at the boundary to approximate the tail beyond $\hat{s}_{het} = \alpha S_{fac,g} > 1$. As with the $N_{fac}$ prior, the variance corresponds to the spread of one decimal order of magnitude between the inflection points.

We do not use the logit-Normal prior form adopted by Zeng et al. for two reasons. First, the logit-Normal has extremely light tails near 0 and 1, so prior–likelihood conflicts in cases of very strong or very weak selection should tend to be resolved in favor of the prior, which is the behavior we seek to avoid. Second, unlike the base-10 Log-Normal, the moments and mode of the logit-Normal are less tractable, which limits flexibility in specifying prior summaries. Our choice ensures that prior–data conflicts are resolved in favor of the data unless the amount of information is extremely small, while (i) maintaining proper tail behavior of all posteriors and (ii) propagating information from Zeng et al.'s function-based approach into our posterior estimates.

#### Data preprocessing

To ensure robust inference, we subsampled gnomAD v4 variants according to the following criteria:

- **Allele number (AN) filter:** AN > 0.9 × max(AN) within the dataset.
- **Roulette quality filter:** quality category "high" or "TFBS" (as specified by the Roulette model).

Synonymous variants were defined as those with "synonymous variant" as the most severe VEP consequence. Putative loss-of-function (pLoF) variants were defined as those whose most severe VEP consequence is one of "stop gained", "splice donor variant", or "splice acceptor variant".

#### Inference procedure

With priors and likelihoods defined, we can now define the full posterior for our vector of parameters Θ with respect to the assumed model and to the observed derived allele frequencies $\{n_i\}$:

$$P(\Theta|\{n_i\}) = P(\Theta)\frac{\mathcal{L}(\Theta|\{n_i\})}{\int P(\Theta)\,\mathcal{L}(\Theta|\{n_i\})\;d\Theta}, \quad (8)$$

where $L(\Theta|\{n_i\})$ is defined as in Eq. 6 and

$$P(\Theta) = P(\mathrm{N_{fac}})P(\mathrm{p_{neutral}}_1)P(\mathrm{p_{neutral}}_2)P(\mathrm{p_{neutral}}_3)\prod_{i=1}^{G} P(\mathrm{S_{fac}}_i),$$

$$d\Theta = d\,\mathrm{N_{fac}}\;\; d\,\mathrm{p_{neutral}}_1\;\; d\,\mathrm{p_{neutral}}_2\;\; d\,\mathrm{p_{neutral}}_3 \prod_{i=1}^{G} d\,\mathrm{S_{fac}}_i\,.$$

Because $S_{fac}$ values for different genes are independent conditional on $N_{fac}$ and $p_{neutral}$ values, the evidence in Eq. (8) may be simplified to:

$$\int P(\Theta)\,\mathcal{L}(\Theta|\{n_i\})\;d\Theta = \int\!\!\int\!\!\int\!\!\int \prod_g \prod_{i\in g}\left[\int P(\mathrm{S_{fac}}_g)\,\mathcal{L}(\mathrm{S_{fac}}_g, \mathrm{N_{fac}}, \mathrm{p_{neutral}}_i\,|n_i)\;d\,\mathrm{S_{fac}}_g\right]$$
$$\times\, P(\mathrm{N_{fac}})P(\mathrm{p_{neutral}}_1)P(\mathrm{p_{neutral}}_2)P(\mathrm{p_{neutral}}_3)\;d\,\mathrm{p_{neutral}}_1\,d\,\mathrm{p_{neutral}}_2\,d\,\mathrm{p_{neutral}}_3\,d\,\mathrm{N_{fac}}\,.$$

However, even with this simplification, the integral is not computationally tractable. To address this issue, we performed Bayesian inference in Stan (https://mc-stan.org/), which implements gradient-based Markov Chain Monte Carlo (MCMC)-based approach that scales to high dimensions. Specifically, Stan implements Hamiltonian Monte Carlo (HMC) with the No-U-Turn Sampler (NUTS), which provides a computationally fast way to generate samples from the full posterior in Eq. 8. And, because the computation of our likelihood does not rely on simulations or precomputed tabulated values (i.e. parameter space is not discrete), the gradient-based approaches are applicable.

We fit the model with this method, running 4 parallel chains, each with 1,000 warm-up (burn-in) iterations used for the adaptation and stabilization of the sampler, followed by 1,000 post–warm-up iterations saved for inference. The 4,000 posterior draws for each parameter (1,000 per chain with no thinning) were combined to estimate the summaries of interest. Convergence and sampling efficiency were assessed using rank-normalized $\hat{R}$ and by checking for divergent transitions (see below).

We conducted two rounds of Θ inference: the seed round on just the variants on chromosome 21 (≈ 1% of the admitted pLoF variants), where we used the Cauchy priors on raw S-factors. Next, to avoid the systematic prior-likelihood conflict with the priors on $s_{het}$ values in subsequent inference, we computed the scaling factor as $\hat{\alpha} = \Sigma_g s^0_g / \Sigma_g \hat{S}_{fac,g} \approx 0.02$, where $s^0_g$ are the mean prior values of Zeng *et al.*, $\hat{S}_{fac,g}$ are the estimates of S-factors obtained with Cauchy priors and g indexes genes on chromosome 21. The scaling factor α ensures that, firstly, there is no systematic shift of posterior means with respect to prior means and, secondly, that the $S_{fac}$ values are computed on the scale of selection coefficients. After the seed run, α was treated as a fixed effect. Finally, the inference was performed on all the admitted pLoF variants with priors on $\alpha\ S_{fac}$ values given by Eq. 7.

#### Robustness of the inference

We employed several techniques to ensure that the posteriors of the parameters are estimated in a reliable way.

**Genome-wide parameters agree with prior expectations.** First, we assessed the concordance of the genome-wide parameters ($N_{fac}$ and $\{p_{neutral,i}\}$) with expectations. As noted above, we expect $N_{fac} \approx 2$. The mean posterior estimate of 2.16 (95% credible interval (2.13, 2.18)) is consistent with that expectation. In addition, based on previous work[17], we expect that ~ 27.3% of variants designated as pLoF by VEP-based predictors are annotated incorrectly, and splice variants are expected to be misannotated more often than stop-gained variants. Our posterior estimates (stop-gained $p_{neutral}$ mean 0.279, 95% credible interval (0.276, 0.282); splice-donor $p_{neutral}$ mean 0.295, 95% credible interval (0.290, 0.300); splice-acceptor $p_{neutral}$ mean 0.320, 95% credible interval (0.314, 0.326)) agree with these expectations.

**Posterior shape.** Second, we confirmed by visual inspection the unimodality of posteriors for all four genome-wide parameters as well as for $S_{fac}$ values of 100 randomly chosen genes.

#### Robustness to perturbations of the prior

Third, we assessed sensitivity of posterior estimates to minor perturbations in the prior. Specifically, for the four genome-wide parameters we evaluated the sensitivity of posterior means to changes in the prior mean and variance, and for the $S_{fac}$ estimates we evaluated sensitivity of posterior geometric means to perturbations in prior variance.

- **$N_{fac}$ estimate.** Decreasing the prior variance tenfold changed the posterior mean by less than 1%, with the estimate under the more peaked prior lying within the 95% credible interval of the estimate obtained under the less peaked prior. Shifting the prior mean by 50% in either direction (the same shifts on the log scale) also changed the posterior mean by less than 1%.
- **$p_{neutral}$ estimates.** For all three $p_{neutral}$ posteriors, decreasing prior variance tenfold and shifting the prior mean by 50% in either direction produced changes < 1% in posterior means.
- **$S_{fac}$ estimates.** As expected, per-gene $S_{fac}$ values (especially for shorter genes) are somewhat more sensitive to prior perturbations due to smaller data volumes. Nevertheless, 50% changes in prior variance produced > 5% changes in posterior geometric means in 35% of genes, > 10% in 15% of genes, and > 20% in 4.7% of genes, with > 50% changes (suggestive of severe prior–posterior conflict) observed in only 62 genes (0.34%).

#### Robustness of the sampler

Fourth, because we did not evaluate the evidence in Eq. (8) by direct integration and instead used an MCMC-based approach, we additionally assessed the robustness of the posterior draws using both Stan's diagnostics and supplementary checks.

- **Stan diagnostics.** Default diagnostics implemented in Stan (checks for divergent MCMC transitions) reported no issues.
- **Convergence ($\hat{R}$).** For the four chains of parameter draws, we computed $\hat{R}$ values using:

$$\hat{R} = \sqrt{\frac{\frac{n-1}{n}W + \frac{B}{n}}{W}}$$

where $n$ is the chain length (number of post–warm-up iterations per chain), $W$ is the average within-chain variance, and $B$ is the between-chain variance. All parameters in all runs had $\hat{R} < 1.05$, indicating good convergence of the sampler. In addition to reporting summaries of posterior $s_{het}$ for each gene, we report $\hat{R}$ for $\hat{\alpha}$ $S_{fac}$ values.

- **Warm-up length.** We assessed robustness to the number of warm-up (burn-in) iterations. For variants on chromosome 21, we ran samplers with 250, 500, 1,000, and 10,000 warm-up iterations and computed $\hat{R}$ values after each setting. As $\hat{R} < 1.05$ for all parameters across all settings, the analysis is effectively insensitive to warm-up lengths around 1,000.
- **Sampling profiles.** We inspected sampling profiles (sequences of draws) for the genome-wide parameters and, by computing $\hat{R}$ on profiles split into chunks of 100 consecutive draws for all parameters, confirmed the absence of trends, supporting robustness of the sampling.

### Per-site estimation of posterior $p_{neutral}$

Having obtained posterior estimates of $N_{fac}$, $\{p_{neutral,\alpha}\}$, and $s_{het,g}$ for each gene, we apply the two–component decomposition (Eq. 3) at each pLoF site to estimate per–site $p_{neutral}$ posteriors. Let a focal site have derived allele count $n \in \{0, 1, 2, \dots\}$ and Roulette mutation–rate bin $\hat{\mu}$, with VEP annotation a and gene g. The site-level mixture is:

$$\hat{f}\big(n \mid \mathrm{N_{fac}}, \hat{\mu}, \mathrm{s_{het}}_g, \mathrm{p_{neutral}}_a\big) = \; (1-\mathrm{p_{neutral}}_a)\,\hat{f}_s\big(n \mid \mathrm{N_{fac}}, \hat{\mu}, \mathrm{s_{het}}_g\big) \; + \; \mathrm{p_{neutral}}_a\;\hat{f}_0(n \mid \hat{\mu})\,.$$

Writing LoF for a “true” loss-of-function site and $\text{LoF}^c$ for misannotation, Bayes’ rule gives:

$$\begin{aligned} P(\mathrm{LoF} \mid n, \hat{\mu}, a, g) &= \frac{P(\mathrm{LoF} \mid a)\,P(n \mid \mathrm{LoF}, \hat{\mu}, g)}{P(\mathrm{LoF} \mid a)\,P(n \mid \mathrm{LoF}, \hat{\mu}, g) + P(\mathrm{LoF}^c \mid a)\,P(n \mid \mathrm{LoF}^c, \hat{\mu})} \\ &= \frac{(1-\mathrm{p_{neutral}}_a)\,\hat{f}_s\big(n \mid \mathrm{N_{fac}}, \hat{\mu}, \mathrm{s_{het}}_g\big)}{(1-\mathrm{p_{neutral}}_a)\,\hat{f}_s\big(n \mid \mathrm{N_{fac}}, \hat{\mu}, \mathrm{s_{het}}_g\big) + \mathrm{p_{neutral}}_a\;\hat{f}_0(n \mid \hat{\mu})}. \end{aligned}$$

To propagate uncertainty in $N_{fac}$, $s_{het□}$, and $p_{neutral,\alpha}$, we average this quantity over the posterior:

$$\widehat{P}(\mathrm{LoF} \mid n, \hat{\mu}, a, g) = \mathbb{E}_{\mathrm{N_{fac}},\,\mathrm{s_{het}}_g,\,\mathrm{p_{neutral}}_a \mid \mathrm{data}}\left[\frac{(1-\mathrm{p_{neutral}}_a)\,\hat{f}_s\big(n \mid \mathrm{N_{fac}}, \hat{\mu}, \mathrm{s_{het}}_g\big)}{(1-\mathrm{p_{neutral}}_a)\,\hat{f}_s\big(n \mid \mathrm{N_{fac}}, \hat{\mu}, \mathrm{s_{het}}_g\big) + \mathrm{p_{neutral}}_a\;\hat{f}_0(n \mid \hat{\mu})}\right]$$

In practice, for each variant we approximate the expectation by Monte Carlo, averaging over 4,000 posterior draw triplets ($N_{fac}^{(m)}$, $s_{het,g}^{(m)}$, $p_{neutral,\alpha}^{(m)}$), where $s_{het,g}^{(m)}$ is drawn from the posterior for the gene g, and $p_{neutral,\alpha}^{(m)}$ from the posterior for its annotation class α:

$$\widehat{P}(\mathrm{LoF} \mid n, \hat{\mu}, a, g) \;\approx\; \frac{1}{4000}\sum_{m=1}^{4000} \frac{\big(1-{\mathrm{p_{neutral}}}_a^{(m)}\big)\,\hat{f}_s\Big(n \mid {\mathrm{N_{fac}}}^{(m)}, \hat{\mu}, {\mathrm{s_{het}}}_g^{(m)}\Big)}{\big(1-{\mathrm{p_{neutral}}}_a^{(m)}\big)\,\hat{f}_s\Big(n \mid {\mathrm{N_{fac}}}^{(m)}, \hat{\mu}, {\mathrm{s_{het}}}_g^{(m)}\Big) + {\mathrm{p_{neutral}}}_a^{(m)}\;\hat{f}_0(n \mid \hat{\mu})}.$$

### LOFTEE-2: a framework for building predictors of loss-of-function

Mikhail A. Moldovan, Jeremy Guez, Julia Goodrich, Greg Rohlicek, Evan Koch, Vladimir Seplyarskiy, Kaitlin Samocha, Konrad Karczewski

#### Introduction

Loss-of-function (LoF) variation reduces or abolishes the function of a gene product and can perturb the biological processes the gene influences. Several molecular mechanisms can lead to LoF in protein-coding genes:

- **nonsense-mediated decay (NMD)**, a post-transcriptional surveillance pathway that degrades mRNAs harboring premature termination codons (most efficiently when the stop lies ~50 nt upstream of the last exon-exon junction[19]), thereby decreasing the quantities of both the transcript and the protein;
- **splice-disrupting changes**[20] that alter donor/acceptor sites or regulatory motifs and yield exon skipping, intron retention, or frameshifts, often resulting in NMD or a nonfunctional isoform;
- **protein instability or misfolding**, which can reduce functional protein abundance via chaperone-mediated quality control and the ubiquitin-proteasome/ER-associated degradation pathways[21]; and
- **direct disruption of protein activity**, for example by truncating catalytic domains or essential interaction motifs. Notably, some pathogenic mechanisms (e.g., dominant-negative effects in TP53[22] or type I collagen genes[23]) are distinct from canonical LoF and should be treated separately from predicted loss of function (pLoF).

Despite extensive progress, predictors that classify sequence variants as LoF across mechanisms remain imperfect and difficult to compare systematically. Existing tools address complementary aspects. For example, splicing models (e.g., splice-site predictors) estimate splice disruption, and NMD propensity models estimate transcript degradation. In addition, benchmarking remains limited and largely ad hoc with no systematic strategies developed.

Here we develop a set of objective benchmarks for constructing and comparing predictors of LoF activity for SNVs, and we present version 2 of the Loss-Of-Function Transcript Effect

Estimator (LOFTEE-2). This method is restricted to canonical pLoF SNVs (specifically stop-gained, splice acceptor, and splice donor variants) and does not consider missense variants.

We introduce evaluation panels spanning curated high-confidence pLoF and benign sets coupled with clinical annotations; segregation-based inferences of LoF effect, and expression-based references. We show that LOFTEE-2 improves discrimination relative to the previous version under these benchmarks.

### Construction of LOFTEE-2

LOFTEE-2 is a set of rule-based filters designed to distinguish, among pLoF variants, those that are likely to be truly loss-of-function from those that are likely misannotated. Each filter has numerical thresholds that we calibrated using the probability of neutrality described in the previous supplementary material section ($p_{neutral}$). For each candidate filter parameter, we swept a range of threshold values and, at each value, computed the geometric mean of the per-variant LoF probability, $GM_{LoF}$, across all retained variants in the gnomAD v4 dataset (chromosome 1, canonical transcripts). When two candidate thresholds produced comparable $GM_{LoF}$ values, we selected the less restrictive one (admitting more variants). Thresholds were placed at transition points in these sweep curves, where tightening the filter began to appreciably increase the mean deleteriousness of the retained set (Extended Data Fig. 6). Confidence intervals on $GM_{LoF}$ were obtained from a Normal approximation to the mean of log-probabilities via the central limit theorem (see Equations below).

We applied this procedure to three classes of pLoF variants:

#### Stop-gained variants in multi-exon genes

We applied two positional filters. First, following the established 50-nt rule, variants falling in the last exon — within 50 nt of the last exon–exon junction — are flagged, as premature stop codons downstream of this point are predicted to escape nonsense-mediated decay. The sweep confirmed that 50 nt is optimal in our data (Extended Data Fig. 6a). Second, following previous research[24], we evaluated a filter on distance from the coding sequence (CDS) start, motivated by the possibility of translation reinitiation following a premature stop[17,24]. The optimal CDS-start cutoff in our data was 50 nt (relaxed) to 70 nt (strict), lower than the 150 nt previously reported;

we did not observe a transition in the segregation-based metric around 150 nt (Extended Data Fig. 6b).

#### Stop-gained variants in single-exon genes

No established positional rule exists for single-exon genes. From the sweep procedure, we found that requiring sufficient distance from both the CDS start and the transcript end produced a segregation-based signal comparable to the multi-exon filters (Extended Data Fig. 6c–d). We adopted distance-from-CDS-start and distance-to-transcript-end filters calibrated independently.

#### Splice-affecting variants

For each splice-site variant, we summarized the predicted splice impact by taking the maximum of the four Pangolin class scores (donor gain, donor loss, acceptor gain, acceptor loss). A variant is retained only if this maximum score exceeds a calibrated threshold. We evaluated both Pangolin and SpliceAI using the sweep procedure; Pangolin provided better discrimination (Extended Data Fig. 6e–f), and we adopted it as the sole splice predictor. The resulting thresholds define two operating points (**Supplementary Table 12**).

| Metric | Relaxed Filter | Strict Filter |
|---|---|---|
| Pangolin max score (splice variants) | ≥ 0.7 | ≥ 0.9 |
| Distance from last exon (multi-exon) | > 50 bp | > 50 bp |
| Distance from CDS start (multi-exon) | > 50 bp | > 70 bp |
| Distance from CDS start (single-exon) | > 250 bp | > 350 bp |
| Distance to transcript end (single-exon) | > 200 bp | > 200 bp |

**Supplementary Table 12 | Strict versus relaxed filters for LOFTEE-2**

The relaxed mode prioritizes sensitivity, while the strict mode prioritizes specificity, allowing users to select the appropriate trade-off for their application.

#### Equations: threshold selection criterion

Given two sets of filtering criteria, $\Lambda_1$ and $\Lambda_2$, that admit variant sets $V_1$ and $V_2$, respectively, we summarize each set by the geometric mean of the segregation-based LoF probabilities

$GM_{LoF}(V_1)$ and $GM_{LoF}(V_2)$. If $GM_{LoF}(V_1) > GM_{LoF}(V_2)$, we regard $\Lambda_1$ as superior to $\Lambda_2$; conversely if the inequality is reversed.

Let $X_v = \log \hat{P}(\text{LoF} \mid d_v, \hat{\mu}_v, a_v, \hat{s}_{gv})$. Under approximate independence and similar dispersion of $\{X_v\}$ within each set, the mean log-scores

$$\bar{X}_V = \frac{1}{|V|} \sum_{v \in V} X_v$$

are approximately Normal by the central limit theorem. We then use the Normal approximation with mean and variance computed from the genome-wide distribution of $\hat{P}(\text{LoF} \mid d, \hat{\mu}, a, \hat{s}_g)$ to compute confidence intervals of $GM_{LoF}(V)$ (assuming $|V| \gg 1$).

If $\Lambda_1$ and $\Lambda_2$ yield $GM_{LoF}(V_{\Lambda 1}) \approx GM_{LoF}(V_{\Lambda 2})$, we select the less restrictive criterion (the one admitting more variants).

### Methods compared to LOFTEE-2

We compared LOFTEE-2 to both the original LOFTEE[2] and to $p_{neutral}$ used as a binary classifier.

#### LOFTEE

LOFTEE applies a set of rule-based filters to identify high-confidence loss-of-function variants among predicted pLoF calls. It flags variants near the end of transcripts, in non-canonical splice sites, or in poorly conserved exonic regions, and classifies each variant as high-confidence (HC) or low-confidence (LC). We used the HC classification as the positive call for LOFTEE.

#### $p_{neutral}$ thresholding

As a complementary approach, we directly used the posterior probability of neutrality as a binary classifier: variants with $p_{neutral}$ below 0.8 were classified as likely loss-of-function. Unlike the rule-based methods, this approach leverages allele frequency information across the cohort rather than sequence or positional features.

### Benchmarks of LoF prediction

We assembled a suite of benchmarks to evaluate loss-of-function prediction. The benchmarks include (i) expert-curated reference sets of LoF and benign variants and (ii) mechanism-specific biological signatures expected of LoF variants. In this work, we focus on signatures of nonsense-mediated decay (NMD) and splicing disruption; benchmarks related to protein instability or direct disruption of catalytic activity are intentionally excluded from scope.

#### Curated LoF variant set

To benchmark the different filtering methods, we used a set of manually curated variants compiled from two previous studies[2,17]. We restricted the analysis to curated variants with an unambiguous verdict ("LoF" or "not LoF"), excluding homozygous-only variants whose functional impact is harder to ascertain through curation. We further required all compared predictors to be defined for each variant, ensuring that precision and recall are computed on the same set of variants across all methods.

Each curated verdict was treated as a binary ground truth label (positive for "LoF", negative for "not LoF"). For each filtering method, we computed precision (TP / (TP + FP)), recall (TP / (TP + FN)), and the $F_{0.5}$ score, which places more weight on precision than on recall, reflecting the priority of avoiding false loss-of-function calls in downstream analyses. Confidence intervals for precision and recall were obtained from a Beta(TP + 1, FP + 1) and Beta(TP + 1, FN + 1), respectively.

#### Expression-based benchmark

Loss-of-function (LoF) variants that trigger nonsense-mediated decay (NMD) are expected to reduce the observed abundance of transcripts carrying the variant allele. However, there are several nuances:

- *(i)* this signature must be distinguished from monoallelic expression (MAE), in which only one allele is expressed independent of the presence of a specific variant; unlike MAE, NMD generally yields partial (not complete) depletion of the variant-bearing transcript, so the variant-allele fraction is expected to fall below 50% but not to zero;
- *(ii)* epigenetic and technical sources of allelic imbalance introduce dispersion around 50%, invalidating a simple Binomial test with $p = 0.5$; and

- *(iii)* splice-disrupting variants can alter isoform composition, complicating direct read-fraction comparisons.

To avoid (iii), we restrict this benchmark to stop-gained variants and construct a data-driven neutral from synonymous variants to address (i) and (ii).

##### Data

We used allele-specific expression (ASE) data from GTEx v10. For each variant, we obtained per-individual, per-tissue read counts (reference allele count, alternative allele count, and total read depth) from the GTEx ASE-by-subject VCF files. Variants were additionally annotated with genotype information (allele depth, total depth, genotype quality, and genotype call) from the GTEx WGS VCF. ASE observations were grouped by variant, individual, and tissue to produce a single ASE record per (variant, individual, tissue) triple.

Variants were restricted to those passing all VCF site filters (PASS), annotated by VEP on protein-coding Ensembl transcripts (BIOTYPE = protein_coding; transcript IDs beginning with ENST), and carrying a consequence of either synonymous_variant or a predicted stop-gained consequence. LOFTEE annotations (HC/LC classification, filter reason, flags, and auxiliary info) were carried over from the VEP annotation run.

##### Background model

For the background, we consider 155,382 synonymous variants in GTEx v10. As a measure of allelic symmetry, we use the fraction of RNA-seq reads carrying the derived allele. With total read depth $n$ and $k$ reads supporting the derived allele, the ideal symmetric case is $k \sim \text{Binom}(k \mid n, 0.5)$. However, beyond perfectly symmetric loci we expect two additional classes:

- **Monoallelic expression (MAE)/extreme imbalance:** $p \approx 0$ or $p \approx 1$, modeled via a U-shaped Beta–Binomial with parameters ($\alpha_U$, $\beta_U$).
- **Random imbalance (noise) around 0.5:** biological/technical variation near symmetry, modeled via a peaked Beta–Binomial with parameters ($\alpha_C$, $\beta_C$).

The resulting mixture for synonymous counts $k_S$ at depth $n$ is:

$$
\begin{aligned}
P(k_S \mid n, \pi_{\text{Binom}}, \pi_U, \alpha_U, \beta_U, \alpha_C, \beta_C) &= \pi_{\text{Binom}} \operatorname{Binom}(k_S \mid n, \tfrac{1}{2}) \\
&\quad + (1-\pi_{\text{Binom}})[\pi_U \operatorname{BetaBinom}(k_S \mid n, \alpha_U, \beta_U) \\
&\quad + (1-\pi_U) \operatorname{BetaBinom}(k_S \mid n, \alpha_C, \beta_C)], \\
\mathcal{L}(\pi_{\text{Binom}}, \pi_U, \alpha_U, \beta_U, \alpha_C, \beta_C \mid \{k_S^{(i)}, n^{(i)}\}) &= \prod_i P(k_S^{(i)} \mid n^{(i)}, \pi_{\text{Binom}}, \pi_U, \alpha_U, \beta_U, \alpha_C, \beta_C), \qquad (1) \\
\ell(\pi_{\text{Binom}}, \pi_U, \alpha_U, \beta_U, \alpha_C, \beta_C \mid \{k_S^{(i)}, n^{(i)}\}) &= \sum_i \log P(k_S^{(i)} \mid n^{(i)}, \pi_{\text{Binom}}, \pi_U, \alpha_U, \beta_U, \alpha_C, \beta_C). \qquad (2)
\end{aligned}
$$

where:

- $\pi_{\text{Binom}}$ is the fraction of perfectly symmetric cases conforming to Binom(*n*, 1/2);
- $\pi_U$ is the mixture weight on the U-shaped Beta–Binomial (MAE/extremes);
- BetaBinom(· | *n*, α, β) denotes the Beta–Binomial pmf at depth *n* with Beta prior Beta(α, β);
- each observation $k_S^{(i)}$ is paired with its own depth $n^{(i)}$.

To accommodate technical artifacts in allele calling and mapping, we do not constrain the Beta–Binomial means to 0.5; both ($\alpha_U$, $\beta_U$) and ($\alpha_C$, $\beta_C$) are freely estimated. In total we estimate six parameters {$\pi_{\text{Binom}}$, $\pi_U$, $\alpha_U$, $\beta_U$, $\alpha_C$, $\beta_C$}; the large sample size (155,382 variants) provides adequate power for stable maximum-likelihood estimation.

We fit the mixture by maximum likelihood using the log-likelihood in Eq. (2) and optimize with the L-BFGS-B algorithm as implemented in scipy.optimize.minimize, enforcing parameter bounds $\pi_{\text{Binom}}$, $\pi_U \in [0, 1]$, $1 > \alpha_U, \beta_U > 0$ and $\alpha_C, \beta_C > 1$.

##### Model of NMD

Let $f_S(k \mid n, \hat{\theta}_s)$ denote the fitted synonymous neutral, with

$$
\hat{\theta}_S = (\hat{\pi}_{\text{Binom}},\ \hat{\pi}_U,\ \hat{\alpha}_U,\ \hat{\beta}_U,\ \hat{\alpha}_C,\ \hat{\beta}_C)
$$

obtained by maximum likelihood on the synonymous set (see previous subsection). We model NMD as an additional Beta–Binomial component with mass shifted left of 1/2. Let $\pi_{\text{LoF}}$ be the fraction of stop-gained variants that trigger NMD and let $g(k \mid n, \varphi) = \text{BetaBinom}(k \mid n, \alpha_{\text{LoF}}, \beta_{\text{LoF}})$ with $\varphi = (\alpha_{\text{LoF}}, \beta_{\text{LoF}})$. The mixture for observed counts *k* at depth *n* is:

$$
P(k \mid n, \pi_{\text{LoF}}, \phi) = (1-\pi_{\text{LoF}})\, f_S(k \mid n, \hat{\theta}_S) \ + \ \pi_{\text{LoF}}\, g(k \mid n, \phi). \qquad (3)
$$

##### Inference

Given the already inferred $\hat{\theta}_s$, we estimate $\pi_{LoF}$ and φ by maximum likelihood on the set of 3,918 stop-gained variants observed in GTEx-v10. For a site with observed (*k*, *n*), the posterior probability that it triggers NMD is given by Bayes' rule:

$$P(\mathrm{LoF} \mid k, n, \pi_{\mathrm{LoF}}, \phi) = \frac{\pi_{\mathrm{LoF}}\, g(k \mid n, \phi)}{\pi_{\mathrm{LoF}}\, g(k \mid n, \phi) + (1 - \pi_{\mathrm{LoF}})\, f_S(k \mid n, \hat{\theta}_S)}. \tag{4}$$

As the shape of the right tails of g and f affects the inference, we additionally regularize the likelihood by polynomially suppressing the $k > n/2$ values at the tail of g.

##### Application of the benchmark

Given a stop-gained variant set *V*, we restrict to those with RNA-seq measurements in GTEx, $V_{GTEx} = \{v \in V : \text{covered in GTEx}\}$. For each $v \in V_{GTEx}$ with observed ($k_v$, $n_v$), we compute the site-level posterior $P_v = P(\mathrm{LoF} \mid k_v, n_v, \pi_{LoF}, \varphi)$ as defined above. For any subset $S \subseteq V_{GTEx}$, we summarize evidence for NMD-like depletion of transcripts by the geometric mean:

$$\mathrm{GM}(S) \;=\; \exp\left(\frac{1}{|S|}\sum_{v\in S}\log P_v\right).$$

To compare two LoF predictors *A* and *B*, we stratify $V_{GTEx}$ by their calls. Let $P_A = \{v : A \text{ calls LoF}\}$, $N_A = \{v : A \text{ calls non-LoF}\}$ (and analogously $P_B$, $N_B$).

For a single-method summary, we compute an enrichment score:

$$E_m \;=\; \frac{\mathrm{GM}(\mathcal{P}_m)}{\mathrm{GM}(\mathcal{N}_m)}, \qquad m \in \{A, B\},$$

and compare $E_A$ vs. $E_B$. Uncertainty was quantified via a 95% confidence interval on the log-enrichment score. The standard error of the difference in log-geometric means was computed as SE = sqrt($s^2P$ / $n_P$ + $s^2_N$ / $n_N$), where $s^2_P$ and $s^2_N$ are the sample variances of the log-posteriors in the LoF-called and non-LoF-called groups, respectively. The confidence interval on the enrichment ratio was then obtained by exponentiating $\log(E) \pm 1.96 \cdot \mathrm{SE}$.

##### Gene-level constraint benchmark (LOEUF)

As an additional benchmark, we evaluated how each LoF filtering method affects the accuracy of gene-level constraint estimation. The rationale is that better LoF filtering — by removing misannotated variants — should yield cleaner observed-to-expected LoF ratios and therefore more accurate identification of constrained genes.

For each filtering method, we computed the LOEUF (Loss-of-function Observed/Expected Upper bound Fraction) for every protein-coding gene in gnomAD v4 by counting, as observed LoF variants, only those variants that pass the given filter. This produces one LOEUF score per gene per method. We then assessed each method's ability to discriminate a curated set of known haploinsufficient (HI) genes (positives) from all remaining genes (negatives) using precision–recall curves, with LOEUF as the continuous score (lower LOEUF indicating stronger constraint). We restricted the analysis to genes for which LOEUF was defined under all compared methods, ensuring that all precision–recall curves are computed on the same gene set (Extended Data Fig. 5b).

We compared six approaches: (i) LOEUF with no filtering ("no LOFTEE"), counting all pLoF variants regardless of annotation quality; (ii) LOEUF with LOFTEE v1, counting only high-confidence variants from the original LOFTEE; (iii–iv) LOEUF with LOFTEE-2 Relaxed and LOFTEE-2 Strict; (v) LOEUF with $p_{neutral} < 0.8$, counting only variants whose posterior probability of neutrality falls below 0.8; and (vi) $s_{het}$, the selection coefficient against heterozygous loss-of-function estimated as described in the previous supplementary material section. For LOEUF-based methods, scores were negated so that higher values indicate stronger constraint; for $s_{het}$, higher values already indicate stronger selection and were used directly. Precision–recall curves and their area under the curve (AUC-PR) were computed using the PRROC package.

#### Guidance on selecting LOFTEE-2 strict versus relaxed mode

LOFTEE-2 provides two operating points — relaxed and strict — that offer different trade-offs between sensitivity and specificity (**Supplementary Table 12**). The choice between them depends primarily on whether the unit of analysis is the gene or the variant.

##### Gene-level analyses: relaxed mode preferred

When pLoF variants are aggregated across a gene to derive a summary statistic — such as LOEUF, burden test statistics, or selection coefficient estimates — individual false positives contribute noise but do not systematically bias results. In contrast, excluding true pLoF variants reduces the effective number of informative observations, decreasing statistical power. In the specific case of LOEUF, using the relaxed filter retains more variants, which reduces the number of genes for which no pLoF variant is expected, a situation that produces undefined constraint estimates. For these reasons, the relaxed mode is generally more appropriate for gene-level analyses, including constraint estimation, gene-level association testing (e.g., rare variant burden or SKAT tests in case-control studies), and calibration of mutation rate models. Accordingly, all analyses in this paper downstream of the LOFTEE-2 construction and benchmarking section — including LOEUF computation, constraint-based gene prioritization, LOEUF-MIS, OMELET, and DisPo — use the relaxed mode.

##### Variant-level interpretation: strict mode preferred

When a specific pLoF variant is being evaluated — for instance, in clinical diagnostics, return of secondary findings, or variant curation — the cost of a false positive is high, as it may trigger unnecessary clinical follow-up or misclassification. In this setting, strict mode is preferred because it maximizes precision, ensuring that variants passing the filter have a high probability of being truly loss-of-function. This is particularly important for variants in medically actionable genes, where the consequences of misannotation extend directly to patient care.

##### Using $p_{neutral}$ for genes under strong selection

For genes under strong purifying selection, $p_{neutral}$ offers a more granular, continuous measure of the probability that a given pLoF variant is misannotated, as opposed to the binary classification provided by the strict and relaxed filters. Because $p_{neutral}$ is estimated from the site frequency spectrum, it is only well-calibrated in genes where the contrast between the selected and neutral frequency distributions is sufficiently strong — that is, genes with low LOEUF values and enough expected pLoF variants to support reliable mixture model inference. In such genes, $p_{neutral}$ can be used to rank variants by their likelihood of being truly loss-of-function, enabling finer prioritization than either LOFTEE-2 mode alone. However, $p_{neutral}$ should be interpreted with caution: it is uninformative for genes under weak or no selection, where the selected and neutral components of the mixture model are poorly separated, and its accuracy depends on the correctness of the underlying mutation rate estimates and demographic model. In particular, $p_{neutral}$ is unreliable for genes affected by clonal expansion in blood or germline, where observed allele frequencies are inflated by somatic or pre-meiotic selective advantages rather than by neutrality, leading to artificially elevated $p_{neutral}$ values. Because the mixture model assumes that variant frequencies are governed by germline mutation rates and population-level selection, any process that distorts the site frequency spectrum — such as clonal hematopoiesis or positive spermatogonial selection — will violate these assumptions and produce misleading estimates. We therefore recommend $p_{neutral}$ as a complementary tool for variant-level assessment in strongly constrained genes, rather than as a genome-wide replacement for the LOFTEE-2 filters.

#### Inspecting discordant variants

In practice, users need not treat the two modes as a binary choice. Variants that pass the relaxed filter but are excluded by the strict filter represent an informative intermediate category. For these variants, examining the specific genomic feature responsible for the discordance — such as a Pangolin splice score between 0.7 and 0.9, or a distance from the CDS start between 50 and 70 bp in multi-exon genes — can inform case-by-case decisions. For example, a splice variant with a Pangolin score of 0.85 excluded only by the strict threshold may warrant retention if orthogonal evidence (e.g., RNA-seq data, functional assays, or allelic expression) supports splice disruption. Conversely, a variant near the CDS start that narrowly passes the relaxed threshold but lacks supporting evidence may be best excluded even in gene-level analyses. We therefore recommend that users examine both modes and use the discordant set as a starting point for further investigation when the application permits.

### Definition of gene lists

Here are defined the gene lists used in this paper, with their sources. One of the main sources is the github page from MacArthur lab (https://github.com/macarthur-lab/gene_lists) which we term here "MacArthur lab's github".

#### Haploinsufficient (HI) genes associated with severe phenotypes

HI genes were downloaded from MacArthur lab's github (severe list)[25]. We added to this list a number of genes more recently curated by Sanna Gudmundsson in Anne O'Donnell-Luria's group (available in the supplement of Gudmundsson *et al.*[26]). All HI genes used in this study can be found in **Supplementary Table 13**.

**Supplementary Table 13 | Haploinsufficient genes used in this study.**

*See attached files.*

#### Haploinsufficient (HI) genes associated with moderate and mild phenotypes

These HI genes were downloaded from MacArthur lab's github (mild and moderate lists; n mild = 46 genes, n moderate = 81 genes, n total = 127 genes)[25].

#### Neurodevelopmental disorder (NDD) associated genes

NDD associated genes were downloaded from: "Genetic modifiers of rare variants in monogenic developmental disorder loci"[27]. They used the clinically curated Developmental Disorders Genotype-to-Phenotype Database (DDG2P)[28] and selected all genes that were annotated as monoallelic (i.e., autosomal dominant) and with a "confirmed" or "probable" evidence level as of November 27, 2020 (n=599 genes).

#### Genes with autosomal recessive (AR) inheritance

AR genes were downloaded from MacArthur lab's github, which used one publications[29,30] (n=1183).

#### Olfactory receptor (OR) genes

OR genes were downloaded from MacArthur lab's github, which used one publication[31] (n=371).

#### Human transcription factors (TFs)

TFs were downloaded from https://humantfs.ccbr.utoronto.ca/download.php, and are based on the article: "The Human Transcription Factors"[32] (n=1554).

#### Kinases

Kinases were downloaded from MacArthur lab's github, curated by UniProt[33] using three publications[34–36] (n=347).

#### Dimer genes

Dimer genes were downloaded from the QuickGo[37–39] (accessed August 2025) using GO:0042803 (protein homodimerization activity, n = 799), GO:0046982 (protein heterodimerization activity, n = 272) and GO:0046983 (protein dimerization activity, n = 29). The gene lists were combined and deduplicated.

#### Helicase genes

Helicase genes were downloaded from QuickGO (accessed August 2025) using GO:0004386 (helicase activity) with descendants, filtered for Swiss-Prot reviewed entries in Homo sapiens (taxon 9606). The genes were deduplicated, yielding 148 unique genes.

#### Channel genes

Ion channel genes (n = 325) were obtained from the HGNC (HUGO Gene Nomenclature Committee) gene group resource at genenames.org[40] (accessed August 2025), encompassing 40 ion channel gene families including potassium voltage-gated channels, sodium voltage-gated channels, calcium voltage-gated channels, transient receptor potential cation channels, glutamate ionotropic receptors, chloride voltage-gated channels, and others.

#### Oncogenes / Tumor suppressor genes (TSG)

Oncogenes (n = 396) and tumor suppressor genes (n = 328) were obtained from the OncoKB cancer gene list[41–43] (accessed August 2025), which integrates cancer gene annotations from OncoKB, MSK-IMPACT, MSK-HEME, Foundation One, Foundation One Heme, Vogelstein et al., and COSMIC Cancer Gene Census v99. Gene aliases were included to maximize matching.

#### Gain of function (GoF) and Dominant Negative (DN) external list

Although we generated our own curated list of GoF and DN using the agentic LLM, we used external lists to validate the results from Figure 4b. The GoF/DN list was a merge of two lists: (1) https://itanlab.shinyapps.io/goflof/ based on one publication[44] and, according to the website, “generated using natural language processing (NLP) on the available abstracts in the Human Gene Mutation Database (HGMD) Professional version” (n=131 genes), and (2) G2P[28] curated list at https://www.ebi.ac.uk/gene2phenotype/panel/DD (downloaded on June 2025; 120 genes). This resulted in a list of 227 genes after deduplication.

#### Online Mendelian Inheritance in Man (OMIM) genes

OMIM genes were downloaded from MacArthur lab’s github which uses the OMIM database[45].

### Combining Deleterious Missense and Loss-of-Function Constraint (Figure 4)

Jeremy Guez, Ruchit Panchal, Julia Goodrich, Kaitlin Samocha, Konrad Karczewski

#### Overview

Figure 4 presents four analyses demonstrating that predicted deleterious missense variants provide constraint information complementary to pLoF variants, and that combining both improves clinical significant genes detection. This section describes the data processing, statistical methods and gene lists used for each panel.

#### Data Sources

All constraint analyses use variant counts from gnomAD v4.1. For each gene, observed and expected variant counts were computed per MANE Select transcript. Expected counts were adjusted for regional mutation rate variation using the gnomAD v3 model[3], as described above, which accounts for local sequence context effects on mutation rates.

Three deep learning-based variant effect predictors were used to score missense variants:

- **ESM1v**[46]: a protein language model trained on evolutionary sequence data that predicts the effect of amino acid substitutions. Because ESM1v was not trained on human population data, it serves as an unbiased predictor with respect to allele frequency information.
- **AlphaMissense**[47]: a structure-aware model that integrates protein language model features with AlphaFold-predicted structures and indirect tuning on population allele frequency data from gnomAD.
- **PopEVE**[48]: an extension of the EVE model[49] that incorporates human population frequency data to refine variant effect predictions.

For each predictor, every possible missense variant in the human exome was scored, and scores were converted to percentile ranks (0–99, where higher values indicate greater predicted pathogenicity). Two types of aggregation were used depending on the analysis. For the percentile-stratified analysis (Fig. 4a), observed and expected variant counts were computed for each specific percentile bin. For the per-gene analyses (Fig. 4b–d), observed and expected counts of variants in the 99th percentile were computed for each gene (i.e., the top 1% most deleterious predicted missense variants). All expected counts use gnomAD v3 mutation model correction for regional mutation rate variation.

### Observed/Expected Ratios by Predicted Pathogenicity Percentile

#### Data Preparation

Variant-level missense data were aggregated from MANE Select transcripts. For each of the three predictors (ESM1v, AlphaMissense, PopEVE), and for each percentile bin (0–99), we summed the observed variant counts and the expected variant counts (with adj_r correction) across all genes. The obs/exp ratio was then computed for each bin as total_obs / total_exp, and plotted as a function of percentile (Figure 4a).

#### Reference Lines

Two horizontal reference lines are shown:

- *Synonymous obs/exp ratio*: the global ratio of observed to expected synonymous variants across all genes, providing a near-neutral baseline (expected value ~1, actual value 1.015).
- *pLoF obs/exp ratio*: the global (genome-wide) ratio of observed to expected predicted loss-of-function variants across all genes (obs/exp = 0.55), representing the level of constraint on pLoF variants.

#### Interpretation

A declining obs/exp ratio with increasing predicted pathogenicity indicates that variants predicted to be more deleterious are depleted from the population, consistent with purifying selection. The observation that the most pathogenic missense variants (99th percentile) show stronger depletion than pLoF variants (obs/exp < 0.55) suggests that some of these variants may cause gain-of-function or dominant-negative effects that are more severely selected against than simple loss of function.

#### Analysis of other missense effect predictors

We can apply the same procedure to other missense effect predictors, such as MPC (Missense deleteriousness Prediction by Constraint[50]; updated in a companion paper), MisFit s[51], EVE[49] (evolutionary model of variant effect), and RaSP[52] (Rapid protein stability prediction).

- MPC is a constraint-aware missense score that combines local (regional or gene-level) missense depletion with amino-acid substitution severity (e.g., BLOSUM[53]) and PolyPhen-2[54] in a logistic regression model, thereby emphasizing mutations that occur in locally missense-intolerant segments of genes.
- MisFit s is a probabilistic graphical model that estimates the human fitness effect of missense variants by linking a learned molecular effect to a predicted selection coefficient (s), trained directly on observed allele counts in large population sequencing data.
- EVE is an unsupervised deep generative model that learns constraints from evolutionary sequence variation across species and predicts variant pathogenicity without relying on supervised clinical labels.
- RaSP is a rapid protein-stability predictor that combines self-supervised structural representations with supervised fine-tuning to estimate mutation-induced changes in protein stability.

In the new figure (**Supplementary Figure 15**), all four predictors still show a general decrease in observed/expected with increasing score percentile, indicating that higher-scoring variants are increasingly depleted from the population, consistent with purifying selection. However, the magnitude of this gradient differs substantially across methods: MPC and MisFit *s* show the strongest separation, with the highest-scoring bins falling to or below the pLoF baseline only at the extreme tail. This pattern suggests that these predictors may be capturing a more constrained subset of missense variants, although differences in calibration or potential overfitting could also contribute to the observed effect. EVE shows a smoother but more modest decline, with only the most extreme scores approaching pLoF-like depletion. RaSP shows the weakest overall gradient and does not reach the pLoF reference line, possibly as a result of protein destabilization capturing only one subset of deleterious missense mechanisms.

These data suggest that different predictors capture distinct biological axes of missense deleteriousness, with population data-aware models providing the clearest enrichment for strongly selected variants. This pattern is also consistent with the Figure 4 comparison between

ESM1v on one hand and AlphaMissense and PopEVE on the other, and is in part expected: predictors that are trained on, or indirectly tuned using, human population variation should more effectively separate variants by gnomAD observed/expected, because this metric itself is derived from population depletion.

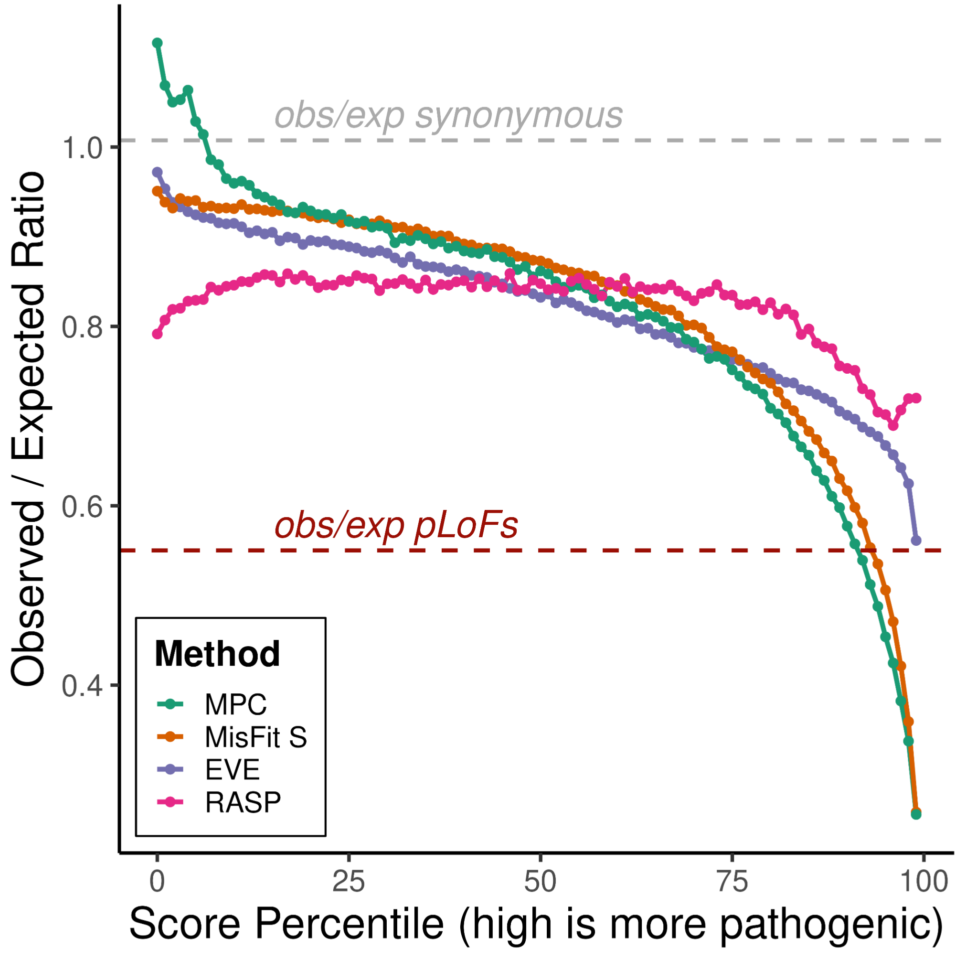


**Supplementary Figure 15 | Observed-to-expected (obs/exp) ratios across score percentiles for missense variants scored by missense effect predictors.**

Four variant effect predictors are shown: MPC, MisFit s, EVE, and RASP. Dashed grey line denotes synonymous variants (obs/exp ≈ 1), and dashed red line denotes pLoF variants (obs/exp = 0.55).

### Enrichment of Gene Categories for Missense-over-pLoF Constraint

Figure 4b tests whether specific gene categories are enriched among genes for which the most deleterious missense variants are more constrained than pLoF variants, relative to expectation.

#### Per-Gene p-Value Computation

For each gene, we tested whether missense constraint exceeds pLoF constraint after accounting for their respective expected mutation rates. The goal is to evaluate whether the depletion of highly deleterious missense variants is stronger than that observed for pLoF variants.

##### Statistical Model

Let O and E denote observed and expected variant counts, respectively. We assume a Poisson observation model:

$$O \mid r \sim \mathrm{Poisson}(rE),$$

where *r* is the true (unknown) observed/expected ratio for the gene. Using an uninformative Gamma prior on *r*,

$$r \sim \mathrm{Gamma}(1, 0),$$

the posterior distribution is conjugate and follows a Gamma distribution:

$$r \mid O, E \sim \mathrm{Gamma}(O + 1, E),$$

using the rate parameterization, where

$$\mathrm{Gamma}(\alpha, \lambda) \quad \text{has density} \quad f(x) = \frac{\lambda^{\alpha}}{\Gamma(\alpha)} x^{\alpha-1} e^{-\lambda x}.$$

##### Missense and pLoF Posteriors

For missense variants, we focus on the top 1% most deleterious variants according to the three predictors (AlphaMissense, ESM1v, PopEVE). Let

$$\overline{O}_{\text{mis}} = \text{average observed count across predictors,}$$

$$\overline{E}_{\text{mis}} = \text{average expected count across predictors.}$$

The posterior for the missense constraint parameter is

$$r_{\text{mis}} \sim \text{Gamma}(a, \lambda_1), \quad a = \overline{O}_{\text{mis}} + 1, \quad \lambda_1 = \overline{E}_{\text{mis}}.$$

For pLoF variants:

$$r_{\text{pLoF}} \sim \text{Gamma}(b, \lambda_2), \quad b = O_{\text{pLoF}} + 1, \quad \lambda_2 = E_{\text{pLoF}}.$$

##### Hypothesis and p-value

We compute the probability:

$$p = \mathbb{P}(r_{\text{mis}} < r_{\text{pLoF}}),$$

which quantifies the posterior probability that missense constraint is stronger than pLoF constraint (i.e., a smaller observed/expected ratio).

Because both posteriors are Gamma-distributed with integer shape parameters, this probability has a closed-form expression:

$$\mathbb{P}(r_{\text{mis}} < r_{\text{pLoF}}) = \sum_{k=0}^{a-1} \binom{b+k-1}{k} \left(\frac{\lambda_1}{\lambda_1 + \lambda_2}\right)^k \left(\frac{\lambda_2}{\lambda_1 + \lambda_2}\right)^b.$$

This expression is valid under the Gamma rate parameterization described above and when $a$ is a positive integer.

##### Interpretation

Small values of *p* indicate strong evidence that

$$r_{\text{mis}} < r_{\text{pLoF}},$$

meaning that the most deleterious missense variants in that gene are more depleted than pLoF variants, suggesting missense-specific selective constraint beyond simple loss-of-function intolerance. This is consistent with gain-of-function or dominant-negative selective pressures, or with other mechanisms (see discussion in the main text).

#### Synonymous Variant Filter

Genes with anomalous synonymous variant counts were excluded to mitigate biases in the mutation rate model. Specifically, only genes with a synonymous obs/exp ratio between 0.7 and 1.3 were retained (±30% tolerance around the expected neutral ratio of 1), removing genes where the expected counts are poorly calibrated (n=3,166 genes excluded).

#### LOF-Matched Controls

To control for the confounding effect of overall gene constraint (genes under strong pLoF constraint tend to also have low missense observed/expected ratios), we used LOEUF-matched controls. For each gene in a given category, we identified the three nearest-neighbour genes (by Euclidean distance on LOEUF scores) not belonging to any benchmark category, using the scikit-learn NearestNeighbors implementation with k = 3.

#### Enrichment Calculation

For each gene category, we tested whether genes from that category are over-represented among genes with $p < 0.1$ (i.e., genes where missense variants are significantly more constrained than pLoF variants). The enrichment ratio was computed as:

$$\text{enrichment} = (\text{proportion of category genes among significant genes}) \\ / \, (\text{proportion of category genes among all tested genes})$$

Statistical significance was assessed using Fisher's exact test (two-sided) on the 2×2 contingency table of category membership × significance status.

#### Background Cleaning

To avoid contamination between gene categories, all genes belonging to any benchmark category were removed from the background (non-category) set. Each category was therefore tested against a clean background of genes not belonging to any curated functional category.

#### Gene Categories Tested

The following gene categories were tested, as described in “Definition of gene lists” above: Oncogenes, TSG, GOF genes, Channels, HI, Kinases, OMIM, OMIM filtered (OMIM minus all enriched categories), Dimers, and Helicases. “OMIM filtered” removes Oncogenes, TSG, GOF, Channels, HI and Dimers from the OMIM set to test whether the OMIM enrichment is driven by known GoF/DN-prone categories.

#### Enrichment analysis using individual variant effect predictors

To assess the contribution of each variant effect predictor individually, we repeated the enrichment analysis from Figure 4b using per-gene p-values computed separately for ESM1v, AlphaMissense, and PopEVE, rather than the averaged missense counts used in the main analysis (**Supplementary Figure 16**). For the averaged score, we computed the mean across available predictor scores for each gene, using the remaining predictors when some scores were missing.

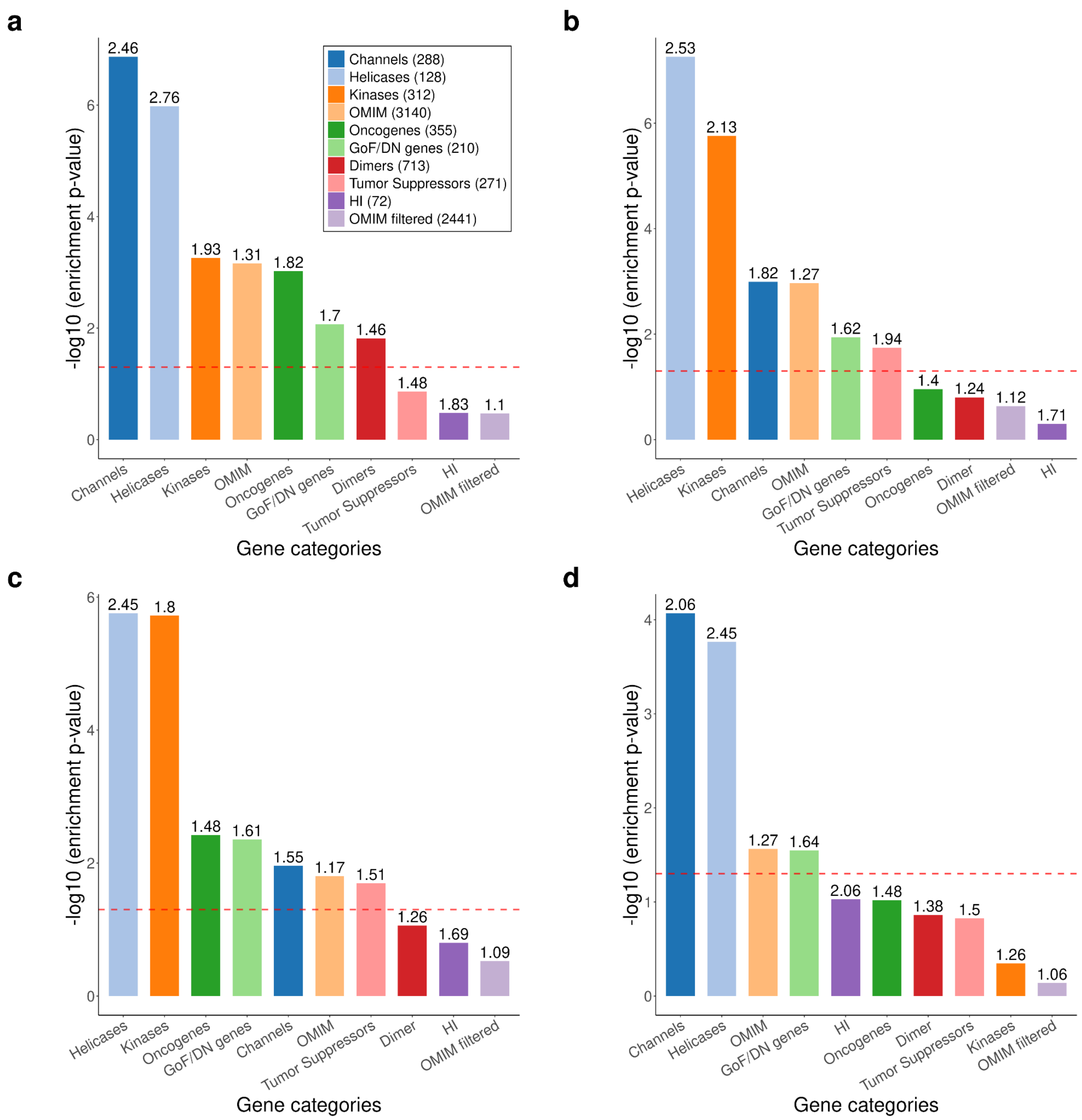


**Supplementary Figure 16 | Enrichment of gene categories for missense constraint higher than pLoF using individual and averaged variant effect predictors.**
Each panel shows the enrichment of ten gene categories among genes whose top 1% most deleterious missense variants are significantly more constrained than their pLoF variants (gamma p-value < 0.1, LOF-matched controls, clean background). Bar height indicates -log10(Fisher's exact test p-value); the dashed red line marks p = 0.05. The number above each bar is the enrichment ratio. (a) Averaged score across ESM1v, AlphaMissense, and PopEVE (as in main text Figure 4b; 11,552 testable genes). (b) ESM1v only (9,471 testable genes). (c) AlphaMissense only (7,710 testable genes). (d) PopEVE only (3,331 testable genes). A gene is testable when its expected count of 99th-percentile missense variants is non-zero. Averaging across predictors increases the number of testable genes and yields broader significance across gene categories compared to any individual predictor, supporting the use of a multi-predictor consensus in the main analysis.

All three predictors individually recapitulate the key enrichment patterns observed in Figure 4b: Helicases, Kinases, Channels, and GOF genes are consistently enriched among genes whose top 1% most deleterious missense variants are more constrained than their pLoF variants. However, the number of testable genes — those with non-zero expected counts for the 99th percentile — varies substantially across predictors: ESM1v yields 9,471 testable genes, AlphaMissense 7,710, and PopEVE only 3,331, compared to 11,552 for the averaged score. This is because many genes, particularly shorter ones, have zero expected variants in the top 1% for a given predictor; averaging across three predictors recovers these genes by pooling information across complementary scoring methods. As a result, PopEVE loses significance for several categories (e.g., Kinases, Oncogenes) due to its smaller testable gene set.

More broadly, the averaged score used in Figure 4b yields stronger and more broadly significant enrichment across gene categories than any individual predictor alone, both because averaging reduces noise in variant-level scores and because it substantially increases gene coverage. This supports the use of a multi-predictor consensus as the primary metric in the main analysis.

#### Observed and Expected Variant Counts in NDD Genes Versus All Genes

Figure 4c compares the mean number of highly deleterious variants per gene in neurodevelopmental disorder (NDD) associated genes versus all other genes, for four variant classes.

##### Variant Classes

For each gene, observed and expected counts were computed for:

- **pLoFs**: predicted loss-of-function variants passing LOEUF v4 quality filters (99.5th percentile annotation confidence threshold, with adj_r correction).
- **ESM1v**: missense variants in the top 1% (99th percentile) of predicted deleteriousness according to ESM1v.
- **PopEVE**: missense variants in the top 1% (99th percentile) of predicted deleteriousness according to PopEVE.

- **AM (AlphaMissense)**: missense variants in the top 1% (99th percentile) of predicted deleteriousness according to AlphaMissense.

##### Gene Filtering

To focus on genes with limited pLoF data (where missense information is most valuable), only genes with fewer than 50 expected pLoF variants were included (n=8,841). HI genes were excluded from the NDD gene set to isolate the contribution of non-haploinsufficiency mechanisms.

##### Computation

For each gene group (NDD or all genes) and each variant class, the mean observed and mean expected counts per gene were computed. The observed/expected ratio was calculated as mean(observed) / mean(expected) and is annotated above each bar pair.

#### Precision-Recall Curves for NDD Gene Classification

Figure 4d evaluates the ability of different constraint metrics to distinguish NDD genes from non-NDD genes using precision-recall analysis.

##### Constraint Metrics Compared

- **LOEUF v2**: the upper bound of the 90% Poisson confidence interval for the observed/expected pLoF ratio, computed from gnomAD v2 exome data as published previously[2].
- **LOEUF v4**: the 95% upper bound of the Gamma posterior of the pLoF mutation rate, divided by the expected count under neutrality, computed from gnomAD v4 exome data. As described in the "LOEUF" section of the Supplementary Methods, under a Poisson observation model with uninformative prior, the posterior of the expected number of segregating pLoF variants given the observed count O is Gamma(shape=O+1, scale=1), and LOEUF is defined as $Q_\gamma(0.975; O+1, 1) / E$, where $Q_\gamma$ denotes the gamma quantile function and E is the expected count under neutrality.
- **LOEUF-MIS**: a combined score that integrates pLoF and highly deleterious missense constraint. For each gene, the observed and expected counts of the top 1% most deleterious

missense variants (averaged across ESM1v, PopEVE and AlphaMissense) are added to the pLoF observed and expected counts, respectively:

$$\text{obs\_combined} = \text{obs\_pLoF} + \text{obs\_missense}$$

$$\text{exp\_combined} = \text{exp\_pLoF} + \text{exp\_missense}$$

where *obs_missense* and *exp_missense* are the row-wise means of the observed and expected counts across the three missense predictors. The LOEUF-mis score is then computed using the same Gamma posterior framework: $Q_{\gamma}$(0.975; obs_combined+1, 1) / exp_combined.

- **GeneBayes** (Zeng et al., 2023): the posterior mean of the selection coefficient ($s_{het}$) inferred by the GeneBayes Bayesian framework, which integrates constraint, conservation and functional annotations.

#### Evaluation Procedure

Precision-recall curves were computed using the PRROC R package[55]. For each metric, genes were ranked (lower scores indicating stronger constraint, except for GeneBayes where the score was used directly), and the area under the precision-recall curve (AUPRC) was computed via numerical integration (auc.integral).

#### Gene Set Definition

Positive genes were NDD genes present in the evaluation set; negative genes were all remaining genes. HI genes were excluded from the entire evaluation to isolate non-haploinsufficiency-mediated NDD genes. Only genes with fewer than 50 expected pLoF variants were included, focusing the evaluation on short genes where missense information provides the greatest added value.

#### Complete-Case Analysis

For each metric, only genes with non-missing and finite scores were included in the evaluation. The LOEUF v2 metric was not used for case filtering (i.e., genes missing LOEUF v2 scores but having valid scores for all other metrics were retained), since its gene coverage differs from gnomAD v4-based metrics.

#### Decomposition of LOEUF-MIS by individual variant effect predictor

To assess the relative contribution of each missense variant effect predictor (VEP) to the combined LOEUF-MIS score, we computed LOEUF-MIS separately using each of the three VEPs included in the main analysis (ESM1v, AlphaMissense, PopEVE), as well as MPC, which was not included in the main LOEUF-MIS definition. For each individual VEP, the LOEUF-MIS score is computed identically to the combined version, except that the missense observed and expected counts come from a single predictor rather than the average of three:

$$\text{LOEUF-MIS (VEP)} = Q_{\gamma}(0.975; \text{obs}_{\text{pLoF}} + \text{obs}_{\text{missense, VEP}} + 1, 1) \;/\; (\text{exp}_{\text{pLoF}} + \text{exp}_{\text{missense,VEP}})$$

where obs_missense_VEP and exp_missense_VEP are the observed and expected counts of variants scoring in the top 1% most deleterious according to that specific VEP. We also computed a four-predictor average (avg 4 VEPs) that includes MPC alongside ESM1v, AlphaMissense and PopEVE.

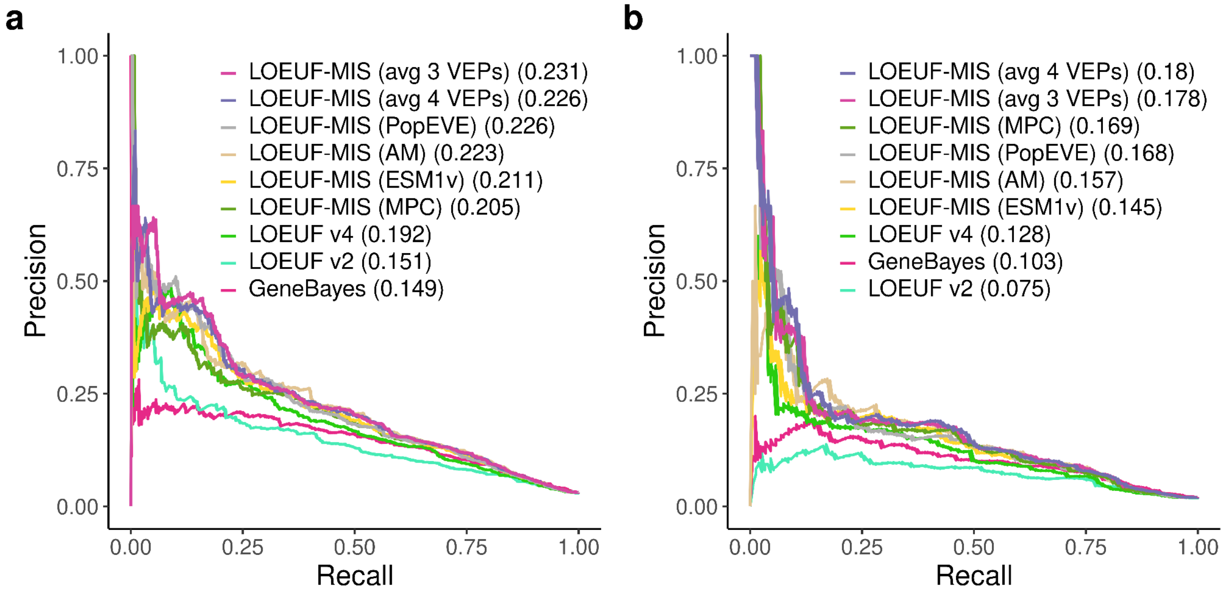


**Supplementary Figure 17 | Precision-recall analysis of LOEUF-MIS decomposed by individual variant effect predictor.**

Precision-recall curves for classifying neurodevelopmental disorder (NDD) genes, excluding haploinsufficient (HI) genes from the positive set. Seven LOEUF-MIS variants are compared—each using a single VEP (ESM1v, AlphaMissense, PopEVE, or MPC) or the average of three or four VEPs—alongside LOEUF v2, LOEUF v4 and GeneBayes. (a) All genes with complete scores (498 positive, 15,765 negative). (b) Genes with fewer than 50 expected pLoF variants (174 positive, 8,792 negative). AUPRC values are shown in parentheses in each legend.

We evaluate all genes for which the metrics can be computed (498 NDD-positive genes, 15,765 negative genes after excluding HI genes and requiring complete cases; **Supplementary Figure 17a**), as well as an evaluation restricted to genes with fewer than 50 expected pLoF variants (**Supplementary Figure 17b**), the regime in which missense information is most valuable because pLoF counts alone provide limited statistical power.

Across all genes (**Supplementary Figure 17a**), LOEUF-MIS averaged over the three main VEPs (AUPRC = 0.231) outperforms every individual VEP decomposition: PopEVE (0.226), AlphaMissense (0.223), ESM1v (0.211), and MPC (0.205). The four-predictor average (0.226) does not improve over the three-predictor average, suggesting that MPC provides partially redundant information relative to the other three predictors in this setting. All LOEUF-MIS variants substantially outperform LOEUF v4 (0.192), LOEUF v2 (0.151) and GeneBayes (0.149).

Among genes with few expected LoFs (**Supplementary Figure 17b**), the advantage of incorporating missense information becomes even more pronounced: LOEUF-MIS (avg 3 VEPs) achieves an AUPRC of 0.178 compared to 0.128 for LOEUF v4, a 39% relative improvement. In this regime, the four-predictor average (0.180) marginally outperforms the three-predictor average (0.178), and MPC (0.169) ranks second among individual VEPs, ahead of PopEVE (0.168) and AlphaMissense (0.157).

These results demonstrate that (i) each individual VEP contributes meaningful signal to missense constraint estimation, (ii) averaging across multiple predictors consistently outperforms any single predictor, and (iii) the benefit of the combined LOEUF-MIS score is greatest for genes where pLoF data alone is insufficient to assess constraint.

#### Software and Packages

All statistical analyses were performed in R (v4.3+) and Python (v3.10+). Precision-recall curves were computed with PRROC (v1.3.1). Nearest-neighbour matching used scikit-learn (v1.3+). Fisher's exact tests used SciPy (v1.11+). Plots were generated with ggplot2 (v3.4+) for the final panels.

### PEPPER: A clinical impact score derived from an LLM pipeline

Jeremy Guez, Stephanie DiTroia, Dmitry Biba, Mark Daly, Heidi Rehm, Anne O'Donnell-Luria, Kaitlin Samocha, Konrad Karczewski

#### Overview

We developed an agentic pipeline that systematically extracts and synthesizes information from the biomedical literature to generate PEPPER (Phenotype Evidence from Published Papers Extracted via Representation), a gene-level clinical impact score. The pipeline employs multiple specialized LLM agents, each focusing on a distinct clinical dimension, and aggregates their probabilistic outputs through Monte Carlo simulations to produce a continuous score ranging from 0 (no established clinical impact) to 1 (severe Mendelian phenotype with high penetrance and early onset). **Figure 5a** provides a high-level overview of the pipeline architecture.

#### Literature Retrieval

The pipeline takes a single gene symbol as input and automatically retrieves relevant literature.

##### Base PubMed Search

For the input gene, the pipeline queries the NCBI PubMed database using the Entrez E-utilities API. The base search combines the gene symbol with disease-related terms:

```
None
("{gene\_symbol}"\[Title/Abstract\]) AND ("variant" OR "mutation" OR "variants"
OR "mutations" OR "disorder" OR "disorders" OR "neurodevelopmental"
OR "developmental" OR "syndrome" OR "syndromes")
```

Articles are retrieved with the following parameters:
- Maximum articles per gene: configurable (default: 50)
- Fields retrieved: PMID, title, abstract, publication date

- Sorting: by relevance (PubMed default ranking)

These articles are passed to the Disease Association Agent (A1, described below), which identifies all diseases associated with the gene and cites supporting PMIDs from the retrieved literature.

#### Disease-Specific Searches

The diseases identified by Agent A1 are used to construct targeted PubMed searches for the downstream agents (A2, A3, A4). Each search combines the gene name, the A1-identified diseases, and agent-specific keywords:

```None
"{gene\_symbol}" AND ({disease\_1} OR {disease\_2} OR ...) AND ({agent\_keywords})
```

The agent-specific keywords are:

```None
A2 (Penetrance): penetrance, "complex trait", "complex disease", polygenic, mendelian

A3 (Inheritance): dominant, "dominant negative", recessive, "co-dominant", recessivity, haploinsufficient, haploinsufficiency

A4 (Mechanism): "gain-of-function", "loss-of-function", "dominant negative", gain, loss
```

Articles from each targeted search are merged with the articles cited by Agent A1 (from the base search), with deduplication by PMID. When more than 15 diseases are identified, only the first 15 (in order of appearance in the A1 response, which typically reflects evidence strength) are included in the query clause.

#### Agent Architecture

The pipeline employs five specialized agents implemented using Claude Haiku 4.5 (Anthropic) with temperature set to 0.0 to ensure reproducibility and minimize creative extrapolation in scientific assessment tasks. Agent A1 receives the gene name and retrieved literature; downstream agents (A2–A4, Onset/Severity) additionally receive the disease list produced by A1.

##### Disease Association Agent (A1)

Purpose: Identify all distinct diseases causally associated with the gene and assess the strength of evidence for each association.

Input: Gene symbol, formatted article list (titles and abstracts)

Output: For each identified disease:
- Disease name (standardized)
- Association score (1–4 scale), which relies on the number of supporting articles
- Justification text
- Supporting PMIDs

Prompt:

```None
You are a senior clinical geneticist. Using the provided PubMed titles and abstracts, identify all distinct diseases directly associated with {gene_name}.

### Input
- Gene name: {gene_name}
- Articles: {articles_text}

### Core Rules

1. **Gene Specificity**
   - Include only diseases linked to variants/mutations of {gene_name} itself
    - Exclude diseases associated with other genes (even if similar names, same family, or interacting partners)
      - Ignore articles where {gene_name} appears as a homonym, acronym, or unrelated context
```

```
2. **Disease Normalization**
   - Merge synonyms, subtypes, and umbrella terms into single entities
    - If one phenotype is the symptom of a more general disease, merge it with the main disease
   - Use the most clinically standard name
   - Do not list symptoms or sub-manifestations as separate diseases
   - Only separate conditions with distinct pathogenic mechanisms

3. **Evidence Requirements**
    - Count only articles providing causal/strong evidence linking {gene_name} to the disease in humans
   - Extract up to 5 supporting PMIDs per disease
   - Use only PMIDs present in the provided text

4. **Scoring System** (strict thresholds)
   - **1 (Definitive)**: ≥5 supporting human articles
   - **2 (Sufficient)**: ≥3 supporting human articles
   - **3 (Moderate)**: ≥2 supporting human articles
   - **4 (Weak)**: ≥1 supporting article
   - **Exclude diseases with 0 PMIDs**

### Output Format

Markdown table with columns: `Disease | AssociationScore | Justification | PMIDs`

- **Disease**: normalized name
- **AssociationScore**: 1-4 integer
- **Justification**: ≤2 sentences summarizing evidence strength
- **PMIDs**: comma-separated list (max 5)

### If No Phenotype Found

If no phenotype caused by a mutation in {gene_name} is found in the provided articles (no causal evidence linking {gene_name} variants to any human disease), return exactly this single row:

`None | NA | [3-sentence summary of articles] | NA`

In the Justification column, provide a 3-sentence summary of what the articles discuss, then state that no phenotype caused by {gene_name} mutations was identified.
```

```
Do NOT invent placeholder names like "No diseases identified" or similar.

Think step by step internally and your output will not break any of the above rules.
```

#### Penetrance Agent (A2) — Probabilistic

Purpose: Estimate the penetrance pattern for each gene-disease pair.

Input: Gene symbol, disease list, penetrance-focused article excerpts

Output: For each disease, a probability distribution over four penetrance categories:
1 – Fully Mendelian: Highly penetrant with clear segregation in families
2 – High penetrance: Strong familial aggregation but not absolute
3 – Moderate penetrance: Variable expressivity or incomplete penetrance
4 – Complex trait: Polygenic or clear departure from Mendelian expectations

The agent outputs probabilities P(mendelian), P(high), P(moderate), P(complex) that sum to 1.

Prompt:

```
None
You are an expert in penetrance and complex trait architecture. Using the diseases detected for {gene_name}, determine the penetrance category for each disease based on the supplied PubMed snippets. Rules:

- Use the same disease names provided, copying each one verbatim without adding suffixes/prefixes or reformatting.
- Output a Markdown table with columns `Disease | P_mendelian | P_high | P_moderate | P_complex | Justification | PMIDs`.
- The four probability columns represent your confidence in each penetrance category:
  * P_mendelian = fully Mendelian (highly penetrant, segregation in families).
  * P_high = high penetrance (strong familial aggregation but not absolute).
   * P_moderate = moderate penetrance (variable expressivity or incomplete penetrance).
   * P_complex = complex trait (polygenic, clear departure from Mendelian
```

```
expectations).
- Each probability must be an integer from 0 to 100.
- The four probabilities MUST sum to exactly 100 for each disease.
- If you are highly confident, concentrate probability on one category (e.g.,
90/5/3/2).
- If evidence is conflicting or ambiguous, spread probability across categories
to reflect uncertainty (e.g., 30/30/25/15).
- Keep justifications ≤2 sentences.
- Cite up to 5 PMIDs (comma-separated) that support the call; write `none` if
unavailable.

Input:
- Diseases: {disease_list}
- Articles: {articles_text}
```

#### Inheritance Agent (A3) — Probabilistic

Purpose: Determine the inheritance pattern for each gene-disease pair.

Input: Gene symbol, disease list, inheritance-focused article excerpts

Output: For each disease, a probability distribution over six inheritance categories:

1 – Dominance: Autosomal dominant (including haploinsufficiency and dominant-negative)

2 – Incomplete dominance (mostly dominant): Predominantly dominant expression

3 – Incomplete dominance: Variable expression

4 – Co-dominant: Co-dominant inheritance

5 – Incomplete dominance (mostly recessive): Predominantly recessive expression

6 – Recessive: Autosomal recessive

Prompt:

```None
You are an expert clinical geneticist specializing in inheritance patterns. For
{gene_name}, evaluate each disease listed and determine the best-supported
inheritance mode using the supplied literature excerpts. Instructions:

- Consider the diseases exactly as given, and reproduce each disease name
```

```
verbatim (no added descriptors or formatting changes).
- Output a Markdown table with columns `Disease | P_dominant | P_inc_dom |
P_incomplete | P_codominant | P_inc_rec | P_recessive | Justification | PMIDs`.
- The six probability columns represent your confidence in each inheritance
pattern:
      * P_dominant = dominance (including dominant negative and
haploinsufficiency).
  * P_inc_dom = incomplete dominance mostly dominant.
  * P_incomplete = incomplete dominance.
  * P_codominant = co-dominant.
  * P_inc_rec = incomplete dominance mostly recessive.
  * P_recessive = recessive.
- Each probability must be an integer from 0 to 100.
- The six probabilities MUST sum to exactly 100 for each disease.
- If you are highly confident, concentrate probability on one category (e.g.,
85/5/5/2/2/1).
- If evidence is contradictory or uncertain, spread probability across
categories to reflect uncertainty (e.g., 40/20/15/10/10/5).
- Keep justifications ≤2 sentences and cite up to 5 PMIDs (comma-separated).
Use `none` if unavailable.

Input:
- Diseases: {disease_list}
- Articles: {articles_text}
```

#### Mechanism Agent (A4)

Purpose: Classify the molecular mechanism underlying each gene-disease association.

Input: Gene symbol, disease list, mechanism-focused article excerpts

Output: For each disease:

- Mechanism classification: Loss-of-Function (LoF), Gain-of-Function (GoF), or Dominant-Negative (DN)
- Confidence score (1–5 scale)
- Justification text
- Supporting PMIDs

This agent enables mechanism-stratified scoring ($PEPPER_{LoF}$, $PEPPER_{GoF}$, $PEPPER_{DN}$) described below.

Prompt:

```
None
You are a molecular mechanism specialist.

Your task: For **{gene_name}** and each disease listed, determine the **best-supported pathogenic mechanism(s)** based on the supplied PubMed article snippets.

### Input
- Diseases: {disease_list}
- Articles: {articles_text}

### Output Format
Produce a **Markdown table** with the following columns:

`Disease | Mechanism | ConfidenceScore | Justification | PMIDs`

- Copy each disease name **exactly as provided** (no edits, abbreviations, or added qualifiers).

### Mechanism Rules
For each disease, assign the mechanism(s) supported by the literature:

- **GoF** — Gain-of-Function
- **LoF** — Loss-of-Function
- **DN** — Dominant-Negative
- **Unknown** — No mechanism is mentioned in the provided articles.
- **Conflicting** — The literature contains genuine contradictions about the mechanism, and the true mechanism is unresolved or debated.

**Multiple mechanisms:** If the literature provides clear evidence that *different mutations in the same gene* cause disease through *different mechanisms*, list all supported mechanisms separated by `/` (e.g., `DN/LoF`, `GoF/LoF`).
- Example: recessive LoF mutations and dominant DN mutations both cause the same disease → write `DN/LoF`.
- Only include mechanisms that are individually supported by evidence in the articles. Do not guess.

**When to use Conflicting:** Use `Conflicting` only when the articles
```

```
explicitly *disagree* about the mechanism (e.g., one study claims GoF while
another claims LoF for the same mutation or patient group), and the question is
genuinely unresolved. Do NOT use `Conflicting` when different mutations simply
act through different well-characterized mechanisms — use `/` notation instead.

**Important:**
- "Autosomal dominant" ≠ "dominant negative."
  *Dominant-negative implies AD inheritance, but AD inheritance alone does not
imply DN.*
- DN is a form of loss-of-function via a dominant mechanism. DN and LoF are
compatible (DN/LoF) — they are not conflicting.

### Confidence Score Rules
`ConfidenceScore` must be ONE of: **1, 2, 3, 4, or NA**

- **1** — Definitive evidence (≥4 independent studies).
- **2** — Strong evidence (clear but fewer studies).
- **3** — Moderate evidence.
- **4** — Weak evidence (very limited or indirect).
- **NA** — Use only for **Conflicting** or **Unknown** mechanisms.

When multiple mechanisms are listed (e.g., `DN/LoF`), the confidence score
reflects the overall confidence in the combined evidence.

Only output the **digit** (1/2/3/4) or **NA**—no text.

### Justification
- Maximum **2 sentences**.
- Cite **1–3 PMIDs** (comma-separated).
- Use **`none`** if no relevant article is available.
```

#### Onset/Severity Agent — Probabilistic

Purpose: Estimate the typical age of onset and clinical severity for each disease.

Input: Disease list, gene symbol

Note: Unlike other agents, this agent relies on the LLM's medical knowledge rather than retrieved literature, as onset and severity are well-established clinical characteristics for most diseases.

Output: For each disease, two probability distributions:
Onset Distribution (7 categories):
1 – Prenatal onset
2 – Neonatal onset
3 – Infancy onset
4 – Childhood onset
5 – Adolescence onset
6 – Adulthood onset
7 – Late-onset (usually > 50y)

Severity Distribution (5 categories):
1 – Profound: Lethal or profoundly disabling, including neurodevelopmental disorders
2 – Severe: Severe impact
3 – Moderate: Moderate impact
4 – Mild: Mild disease
5 – Very mild: Very mild or incidental

Severity assessment considers the disease course with appropriate treatment/management, not untreated natural history.

Prompt:

```None
You are an experienced medical doctor. Estimate the typical age of onset AND
the overall disease severity for each disease listed below. Base your answers
on core medical knowledge (no literature references are required). Follow these
instructions:

- Output a Markdown table with columns `Disease | P_prenatal | P_neonatal |
P_infancy | P_childhood | P_adolescence | P_adulthood | P_late | P_profound |
P_severe | P_moderate | P_mild | P_verymild | Justification`.
```

```
- Copy each disease name exactly as listed (verbatim, no added qualifiers or formatting changes).

- The first seven probability columns represent your confidence in each onset category:
  * P_prenatal = Prenatal onset.
  * P_neonatal = Neonatal onset.
  * P_infancy = Infancy onset.
  * P_childhood = Childhood onset.
  * P_adolescence = Adolescence onset.
  * P_adulthood = Adulthood onset.
  * P_late = Late-onset (usually >50y).

- The last five probability columns represent your confidence in each severity category:
   * P_profound = lethal or profoundly disabling, including neurodevelopmental disorders.
  * P_severe = severe impact.
  * P_moderate = moderate impact.
  * P_mild = mild disease.
  * P_verymild = very mild / incidental.

- Each probability must be an integer from 0 to 100.
- The onset probabilities (P_prenatal through P_late) MUST sum to exactly 100.
- The severity probabilities (P_lethal through P_verymild) MUST sum to exactly 100.
- If you are highly confident, concentrate probability on one category.
- If onset/severity is variable or uncertain, spread probability across categories to reflect that variability.

- **Important**: Base the severity probabilities on the disease's impact WITH appropriate
  treatment/management, not the untreated natural history. Consider:
  - Long-term quality of life under standard care
  - Whether treatment fully resolves symptoms or just manages them
  - Lifelong burden of treatment/monitoring
   - Diseases impacting cognitive abilities and autonomy of the individual should have a high probability of P_profound

- The `Justification` column should contain 1–2 sentences covering both onset and severity in a single explanation (e.g., "Typically manifests in adolescence with progressive muscle weakness leading to moderate lifelong disability.").

- Evaluate every disease in the list. Do not invent new diseases.
```

```
- For diseases that are well treated, do not concentrate probability on
profound or severe categories.

Diseases:
{disease_list}
```

#### PEPPER Calculation

PEPPER is computed for each gene-disease association identified by Agent A1. Genes associated with multiple diseases will have multiple PEPPER values, which are then aggregated at the gene level.

##### Continuous Score Framework (v2)

The raw outputs from agents A1–A4 and the Onset/Severity agent are integrated into a continuous PEPPER score ranging from 0 to 1, where higher scores indicate more severe clinical impact. The score is computed by multiplying individual variable scores, each mapped to a continuous scale:

Score Components:

1. Association Score (A1): Evidence strength from Agent A1
   - Score 1 (Definitive) → 1.0
   - Score 2 (Strong) → 0.5
   - Score 3 (Supportive) → 0.25
   - Score 4 (Limited) → 0.1

2. Penetrance Score (A2): Penetrance pattern from Agent A2
   - Fully Mendelian → 1.0
   - High penetrance → 0.75
   - Moderate penetrance → 0.5
   - Complex trait → 0.1

3. Inheritance Score (A3): Inheritance pattern from Agent A3

- Dominant → 1.0
- Incomplete dominance (mostly dominant) / Co-dominant → 0.75
- Incomplete dominance → 0.5
- Incomplete dominance (mostly recessive) → 0.25
- Recessive → 0.1

4. Onset Score: Age of onset from Onset/Severity Agent
   - Prenatal / Neonatal → 1.0
   - Infancy → 0.9
   - Childhood → 0.75
   - Adolescence → 0.5
   - Adulthood → 0.25
   - Late-onset → 0.1

5. Severity Score: Clinical severity from Onset/Severity Agent
   - Critical (including lethal) → 1.0
   - Severe → 0.75
   - Moderate → 0.5
   - Mild → 0.1
   - Very mild → 0.01

PEPPER is computed as the product of all five component scores:

PEPPER = $\text{score}_{A1} \times \text{score}_{A2} \times \text{score}_{A3} \times \text{score}_{onset} \times \text{score}_{severity}$

This multiplicative formulation ensures that all components must contribute positively for a high PEPPER, reflecting the principle that the most severe clinical impact score requires strong evidence, high penetrance, dominant inheritance, early onset, and high severity simultaneously.

Example:

- A1=1 (1.0), A2=Mendelian (1.0), A3=Dominant (1.0), Onset=Neonatal (1.0), Severity=Profound (1.0)

→ PEPPER = 1.0 × 1.0 × 1.0 × 1.0 × 1.0 = 1.0 (maximum severity)

- A1=2 (0.5), A2=High (0.75), A3=Incomplete dominant (0.5), Onset=Childhood (0.75), Severity=Severe (0.75)
→ PEPPER = 0.5 × 0.75 × 0.5 × 0.75 × 0.75 = 0.105 (moderate severity)

#### Monte Carlo Simulation

Four of the agents' scores are given as probability distributions (age of onset, severity, penetrance, mode of inheritance), which inherently capture agent uncertainty regarding the gene. Monte Carlo simulation propagates this uncertainty from the agent probability distributions through the continuous score calculation. This yields both an expected PEPPER value and a variance estimate that are subsequently used for Bayesian integration with constraint metrics and for quantifying disagreement between PEPPER and observed genetic constraint.

Algorithm:

```
None
Input: Disease record with probability distributions
       P_penetrance, P_inheritance, P_onset, P_severity
       Association score A1 (fixed, from Agent A1)

Parameters: N = 3,000 samples, seed = 42

For i = 1 to N:
    Sample penetrance_i ~ Categorical(P_penetrance)
    Sample inheritance_i ~ Categorical(P_inheritance)
    Sample onset_i ~ Categorical(P_onset)
    Sample severity_i ~ Categorical(P_severity)

    Map each sampled value to continuous score:
        score_A1 = MapAssociation(A1)
        score_A2 = MapPenetrance(penetrance_i)
        score_A3 = MapInheritance(inheritance_i)
        score_onset = MapOnset(onset_i)
        score_severity = MapSeverity(severity_i)

        Compute  PEPPER_i  =  score_A1  ×  score_A2  ×  score_A3  ×  score_onset  ×
score_severity

Output:
    - Expected PEPPER: E[PEPPER] = (1/N) × Σ_i PEPPER_i
    - Variance: Var[PEPPER] = (1/N) × Σ_i (PEPPER_i - E[PEPPER])²
    - Sample distribution: {PEPPER_1, PEPPER_2, ..., PEPPER_N}
```

##### PEPPER Interpretation

PEPPER is a continuous value between 0 and 1, directly computed from the multiplicative combination of component scores. No additional normalization is required:
- PEPPER = 1.0: Maximum clinical severity (definitive evidence, fully Mendelian, dominant inheritance, prenatal/neonatal onset, profound severity)
- PEPPER ≈ 0.0: Minimal or no established clinical impact
- PEPPER = 0.0: Genes with no disease associations identified by Agent A1 are assigned a PEPPER of 0.0, indicating no established clinical impact

The continuous nature of the framework provides finer granularity than discrete level-based approaches and naturally captures the multiplicative interactions between clinical dimensions.

#### Gene-Level Aggregation

##### Gene-level PEPPER

For genes associated with multiple diseases, the gene-level score is defined as the maximum PEPPER across all associated diseases:

$$\text{PEPPER} = \text{max over d in Diseases(g) of PEPPER(d)}$$

This reflects the principle that a gene's clinical relevance should be assessed by its most severe associated phenotype.

##### Variance and Bayesian Prior

The Monte Carlo simulation also produces a variance estimate for each disease-level PEPPER. This variance quantifies the uncertainty in the LLM agents' assessments and will be used to derive an informative prior strength (κ parameter) for downstream Bayesian inference integrating constraint metrics (see Supplementary Section: OMELET–Bayesian Integration of LLM Priors with LOEUF Constraint).

$$\kappa = f(\text{Var[PEPPER]})$$

where higher variance (greater uncertainty) yields lower κ (weaker prior influence). The κ parameter is computed from the PEPPER score (0-1) and its variance using a transformation that maps the continuous score to a probability scale for Bayesian integration.

##### Mechanism-Stratified Scores

Using the Mechanism Agent (A4) output, the pipeline computes mechanism-specific PEPPER values:
- $PEPPER_{LoF}$: Maximum PEPPER among diseases with Loss-of-Function mechanism
- $PEPPER_{GoF}$: Maximum PEPPER among diseases with Gain-of-Function mechanism
- $PEPPER_{DN}$: Maximum PEPPER among diseases with Dominant-Negative mechanism

These stratified scores enable analyses of mechanism-specific constraint patterns and can be integrated with mechanism-aware constraint metrics.

#### Implementation Details

##### Software and Dependencies

- LLM API: Anthropic Claude Haiku 4.5
- Literature retrieval: NCBI Entrez E-utilities (Biopython)
- Parallel processing: Python multiprocessing (ProcessPoolExecutor)
- Data handling: pandas, NumPy

##### Reproducibility Parameters

- LLM temperature: 0.0
- Monte Carlo samples: 3,000
- Random seed: 42
- Max articles per search: Configurable (default: 50)

##### Output Format

For each gene, the pipeline produces:
1. Per-disease records: Association score, penetrance/inheritance/onset/severity distributions, mechanism classification, PEPPER with confidence intervals

2. Gene-level summary: gene-level PEPPER, $PEPPER_{LoF}$, $PEPPER_{GoF}$, $PEPPER_{DN}$, variance estimates
3. Raw agent outputs: Full LLM responses for reproducibility and auditing, including citations and justifications

### Citation Validation (Hallucination rate)

To assess the reliability of literature citations generated by the LLM agents, we systematically verified that all PMIDs cited in agent outputs were present in the input article sets provided to each agent. This validation guards against PMID hallucination, where an LLM might fabricate plausible-looking but non-existent or irrelevant citations.

#### Methodology

For each of the 21,955 genes processed, we extracted:
1. Input PMIDs: All article PMIDs provided to each agent (A1–A4) as context
2. Cited PMIDs: All PMIDs referenced by agents in their disease association outputs

A citation was flagged as potentially hallucinated if it appeared in an agent's output but was not present in that agent's input article set.

#### Results

Citation validation across all agents demonstrated zero hallucination rate (**Supplementary Table 14**), with the note that A2–A4 receive both their targeted search results and articles cited by A1, hence the "unique" count excludes articles already provided to A1.

**Supplementary Table 14 | Hallucination rate for agentic agents.**

| Agent | Input PMIDs | Cited PMIDs | Hallucination Rate |
|---|---|---|---|
| A1 (Disease) | 436,401 | 71,961 | 0.00% |
| A2 (Penetrance) | 478,498 (405,581 unique) | 117,991 | 0.00% |
| A3 (Inheritance) | 114,019 (32,061 unique) | 74,803 | 0.00% |
| A4 (Mechanism) | 116,659 (36,475 unique) | 61,624 | 0.00% |
| **Total (deduplicated)** | **903,345** | **156,341** | **0.00%** |

##### Interpretation

The absence of PMID hallucinations indicates that the LLM agents reliably constrain their citations to the provided literature context. This grounding behavior is essential for scientific reproducibility, as it ensures that all evidence claims can be traced back to verifiable source articles. The use of explicit article context in prompts, combined with temperature=0.0 for deterministic outputs, effectively prevents citation fabrication.

#### External Validation Against GenCC

##### GenCC Database Overview

The Gene Curation Coalition (GenCC[56]) is a collaborative effort that harmonizes gene-disease validity assessments from multiple expert curation groups including ClinGen, G2P, Orphanet, GEL PanelApp, PanelApp AU, internal clinical laboratory databases, and others. Most submitters to GenCC (except Orphanet) provide standardized classifications of gene-disease relationships using an evidence-based framework with the following levels (in decreasing order of evidence strength): Definitive, Strong, Moderate, Limited, Disputed, and Refuted and No Known Disease Relationship. Orphanet records are not classified and are mapped to “Supportive” based on the intent of the database to capture valid disease relationships. All submissions are mapped to the Monarch Disease Ontology (MONDO).

The expert curated ClinGen dataset, enhanced by additional submissions to GenCC, serves as a gold standard for evaluating the accuracy of our agentic pipeline's disease association predictions.

##### Disease Matching Methodology

Comparing disease associations between the agentic pipeline output and GenCC entries requires semantic matching, as disease nomenclature varies significantly across sources. We implemented a hybrid algorithmic-LLM approach:

###### GenCC Disease Grouping

GenCC entries for a given gene often contain multiple submissions referring to the same disease under different names. To consolidate these entries:

1. Algorithmic grouping: Diseases are clustered using asymmetric token coverage. Two disease names are grouped if ≥ 30% of tokens from either name appear in the other (whichever direction yields higher coverage). This captures variants such as "hereditary spastic paraplegia 75" and "complex hereditary spastic paraplegia" as the same entity.

2. LLM semantic refinement: An LLM (Claude Haiku 4.5) reviews the algorithmic groups and assigns clinically meaningful group titles, merging semantically related entries that token-based methods may miss.

3. Classification aggregation: For each group, the strongest evidence classification among constituent entries is retained as the group's primary classification.

##### Pairwise Disease Comparison

For each disease identified by the agentic pipeline, we compare it against each GenCC disease group using a two-stage approach:

Stage 1 – Token coverage optimization:
If ≥ 30% of tokens from either disease name appear in the other, the pair is scored as identical (score = 1) without LLM invocation.

Stage 2 – LLM semantic comparison:
When token overlap is insufficient, an LLM evaluates the pair with the following prompt:
"Are "{pipeline_disease}" and "{gencc_disease}" the same? Reply: 1 (identical), 2 (similar), 3 (related), or 4 (completely different)."

##### Match Classification

Based on comparison scores, disease pairs are classified as:
- Matched: Score ∈ {1, 2, 3} — the agentic pipeline successfully identified a GenCC-curated association
- GenCC-only: GenCC entries with no matching pipeline disease
- Agentic pipeline-only: Pipeline diseases absent from GenCC

#### Validation Metrics

The primary validation metric is the concordance rate, defined as:

Matching Rate = (Matched / (Matched + GenCC-only)) × 100

This metric quantifies the proportion of GenCC-curated gene-disease associations successfully recovered by the agentic pipeline.

Additional metrics include:
- Concordance by GenCC evidence level: Stratified analysis across Definitive, Strong, Moderate, Supportive, and Limited classifications. If a disease had multiple submissions per disease association with different evidence levels, we selected the most confident association.
- Concordance by pipeline evidence level: Stratified by the agentic pipeline's association score (1–4) which was mapped to Definitive (1), Strong (2), Supportive (3), Limited (4).
- Novel discovery rate: Number of pipeline-only associations, representing potentially novel gene-disease relationships not yet curated in GenCC.

#### Interpretation Considerations

Several factors should be considered when interpreting validation results:
1. GenCC scope: GenCC submitters focus on genes for which a Mendelian gene-disease relationship has been claimed in the literature. Associations involving complex traits or modifier effects may be absent from GenCC but validly identified by the pipeline.

2. Temporal lag: Submissions to GenCC might follow publication with some delay. Recent literature discoveries may appear as pipeline-only associations pending curation by GenCC submitters and submission to GenCC, while the LLM pipeline can read and curate articles as soon as they are indexed in Pubmed.

3. Nomenclature heterogeneity: Despite our hybrid matching approach, some true matches may be missed due to substantial nomenclature differences between sources.

4. Evidence threshold differences: The agentic pipeline and GenCC submitters may apply different thresholds for what constitutes sufficient evidence, leading to classification discrepancies.

#### Results Summary

The validation analysis demonstrated high concordance between our agentic pipeline and GenCC, with 95.6% overall recovery of curated associations (Definitive, Strong Moderate, Supportive, Limited) and near-complete capture of Definitive (99.3%) and Strong (97.7%) classifications (**Supplementary Figure 18**). Beyond reproducing known associations, the pipeline identified over 7,000 gene-disease relationships not yet curated by GenCC submitters. Full results are presented in the main text (**Supplementary Figure 19**).

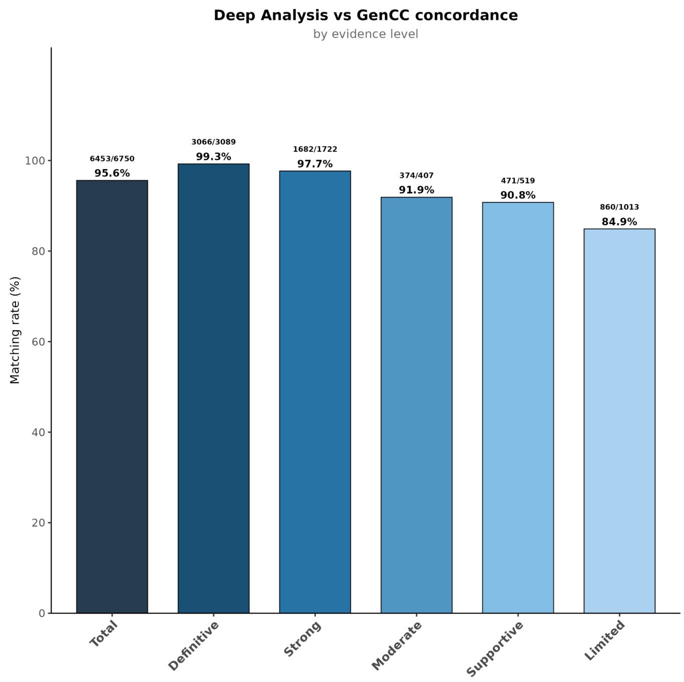


**Supplementary Figure 18 | Matching rate per GenCC confidence category.**

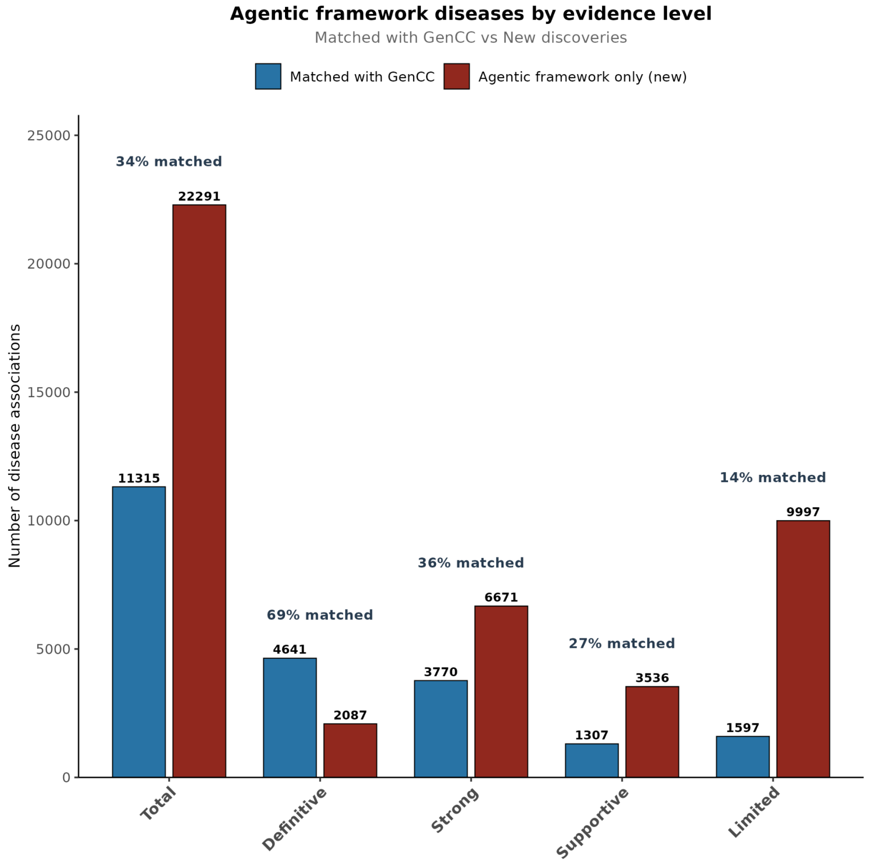


**Supplementary Figure 19 | Number of disease associations for each level of confidence in the agentic LLM framework.**

The diseases are split into those found in GenCC and those not present in GenCC.

### Agentic Framework Results

We examined baseline statistics of the agentic framework outputs across all identified gene-disease associations. Results are presented separately for genes and diseases.

#### Gene Statistics

A total of 21,944 genes were analyzed by the framework. Of these, 11,451 genes (52.2%) were associated with at least one disease, including 5,040 genes linked to at least one Mendelian disease with a minimum confidence level of "Supportive." The remaining 10,493 genes without disease associations comprised two subsets: 3,269 genes with no published literature available, and 7,224 genes for which agent A1 did not detect any disease associations in the available literature (**Supplementary Figure 20**).

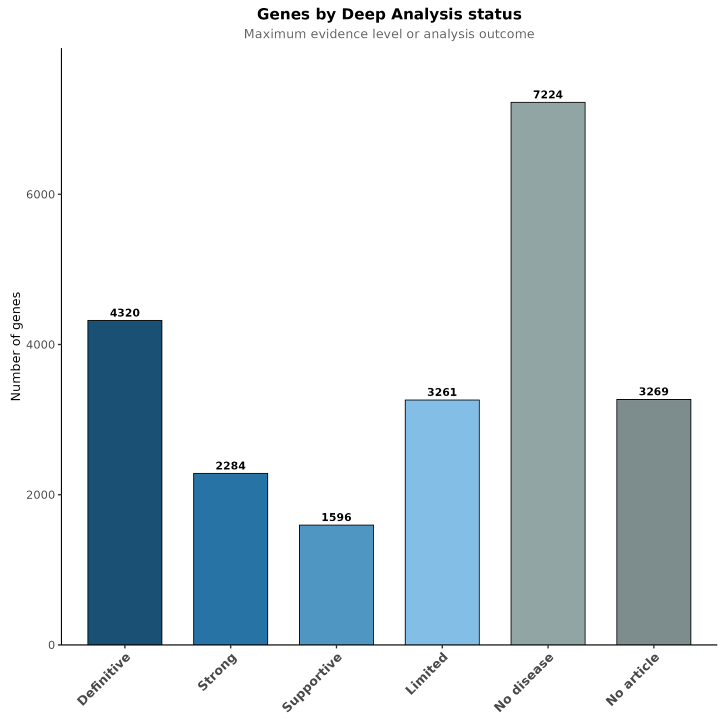


**Supplementary Figure 20 | Classification of genes by maximum evidence level.**
For each gene, we determined the highest evidence level among all its disease associations. Genes are grouped into four evidence categories (Definitive, Strong, Supportive, Limited) based on their best association score, or classified as having no disease identified after article analysis (no disease) or no articles retrieved (no article). This classification reflects the most confident disease association per gene.

We next examined the number of genes associated with a single disease mechanism versus those exhibiting two or all three mechanisms (LoF, GoF, and DN). For this analysis, we restricted our dataset to gene-disease associations with high confidence levels (Definitive or Strong evidence), and high mechanism confidence levels (≤ 2). Genes with LoF as the sole mechanism were the most prevalent (n=4,308), followed by genes exhibiting both LoF and GoF mechanisms (n=813) or LoF and DN (n=186). Notably, few genes were associated exclusively with a dominant-negative mechanism (n=142), while only 26 genes displayed both GoF and DN mechanisms. Interestingly, we identified 81 genes associated with all three pathogenic mechanisms simultaneously (**Supplementary Figure 21**, **Supplementary Table 15**).

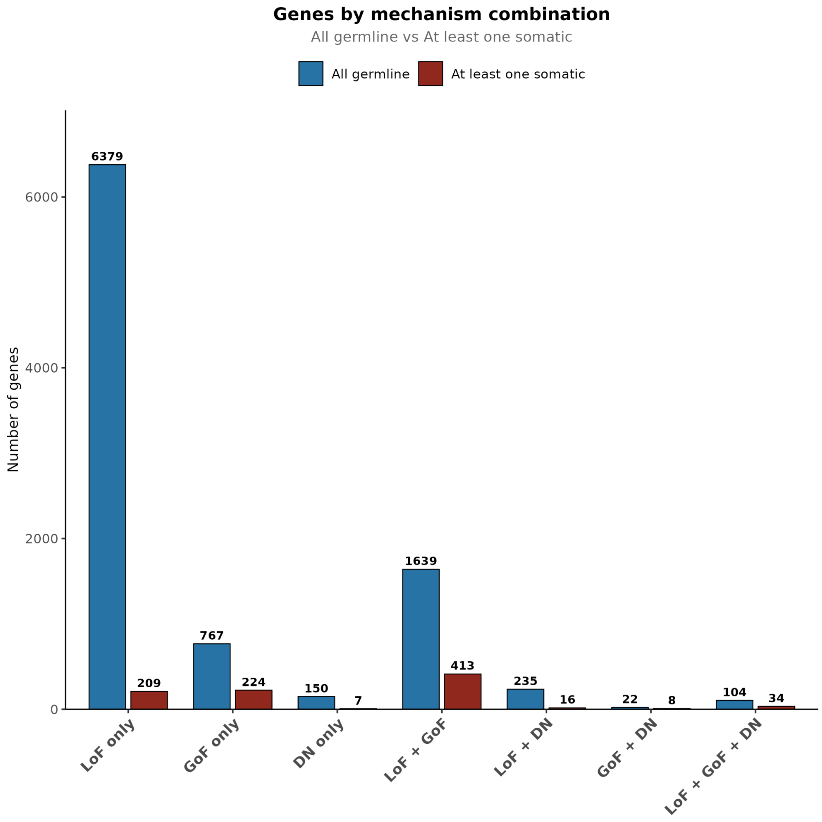


**Supplementary Figure 21 | Distribution of genes across disease mechanism combinations, stratified by mutation origin.**

Genes are classified by their associated pathogenic mechanisms: loss-of-function (LoF), gain-of-function (GoF), dominant-negative (DN), or combinations thereof. For each mechanism category, genes are further stratified into two groups based on mutation origin: "All germline" (blue bars) represents genes in which all associated disease mechanisms are linked to at least one germline mutation, while "At least one somatic" (red bars) represents genes in which at least one disease mechanism is exclusively associated with somatic mutations. Analysis is restricted to gene-disease associations with Definitive or Strong evidence classifications. Numbers above bars indicate gene counts for each category.

**Supplementary Table 15 | Representative examples of genes exhibiting single and multiple disease mechanisms.**

Genes are grouped by mechanism: loss-of-function (LoF) only, gain-of-function (GoF) only, dominant-negative (DN) only, and combinations thereof. For genes with multiple mechanisms, the specific mechanism underlying each associated disease is indicated in parentheses. All associations represent Definitive or Strong evidence classifications.

| Category | Gene Symbol | Disease Names |
|---|---|---|
| LoF only | *DIS3L2* | Perlman Syndrome |
| LoF only | *MED23* | Intellectual Disability |
| LoF only | *VLDLR* | Cerebellar Hypoplasia |
| GoF only | *CACNA1C* | CACNA1C-Related Neurodevelopmental Disorder |
| GoF only | *HRAS* | Costello syndrome |
| GoF only | *RAC3* | Neurodevelopmental disorder with structural brain anomalies and dysmorphic facies (NEDBAF) |
| DN only | *FZD2* | Omodysplasia Type 2 (OMOD2) |
| DN only | *P4HB* | Cole-Carpenter Syndrome |
| DN only | *WDR37* | Neurooculocardiogenitourinary syndrome (NOCGUS) |
| LoF + GoF | *CCND2* | Megalencephaly-Polymicrogyria-Polydactyly-Hydrocephalus (MPPH) Syndrome (GoF) \| Microcephaly with Developmental Delay (LoF) |
| LoF + GoF | *GPC3* | Wilms Tumor (GoF) \| Simpson-Golabi-Behmel Syndrome (LoF) |
| LoF + GoF | *ANTXR1* | Infantile Hemangioma (GoF) \| GAPO Syndrome (LoF) |
| LoF + DN | *SMC3* | Cornelia de Lange Syndrome (DN) \| Developmental Delay/Intellectual Disability (LoF) |
| LoF + DN | *FOXN1* | Transient T-Cell Lymphopenia (DN) \| Nude Severe Combined Immunodeficiency (Nude/SCID) (LoF) |
| LoF + DN | *WDR62* | Premature Ovarian Insufficiency (POI) (DN) \| Autosomal Recessive Primary Microcephaly (MCPH2) (LoF) |
| GoF + DN | *DPF2* | Coffin-Siris Syndrome (DN) \| Cervical Cancer (GoF, somatic) |
| GoF + DN | *CTBP1* | Hypotonia, Ataxia, Developmental Delay, and Tooth Enamel Defects Syndrome (HADDTS) (DN) \| Mitochondrial Respiratory Chain Dysfunction (GoF) |
| GoF + DN | *RAC2* | Neutrophil Dysfunction/Deficiency (DN) \| Primary Immunodeficiency - Severe Combined Immunodeficiency (SCID) (GoF) |
| LoF + GoF + DN | *ZSWIM6* | Severe Intellectual Disability/Neurodevelopmental Disorder (DN) \| Acromelic Frontonasal Dysostosis (GoF) \| Autism Spectrum Disorder (LoF) |
| LoF + GoF + DN | *CDH11* | Teebi Hypertelorism Syndrome (DN) \| Aneurysmal Bone Cyst (GoF, somatic) \| Elsahy-Waters Syndrome (LoF) |
| LoF + GoF + DN | *EVC2* | Weyers Acrofacial Dysostosis (DN) \| Acute Myeloid Leukemia (GoF, somatic) \| Ellis-van Creveld Syndrome (LoF) |

#### Gene-disease associations-level statistics

##### Distribution of gene-disease associations

Among the 33,641 gene-disease associations identified, 17,169 (51.0%) were classified with high confidence levels: 6,728 as Definitive and 10,441 as Strong evidence. We subsequently analyzed the inheritance patterns of these high-confidence associations.

Autosomal dominant inheritance was the most prevalent mode, accounting for 7,062 disease associations, followed closely by autosomal recessive inheritance with 6,209 associations, while other inheritance patterns were less frequent (**Supplementary Figure 22**).

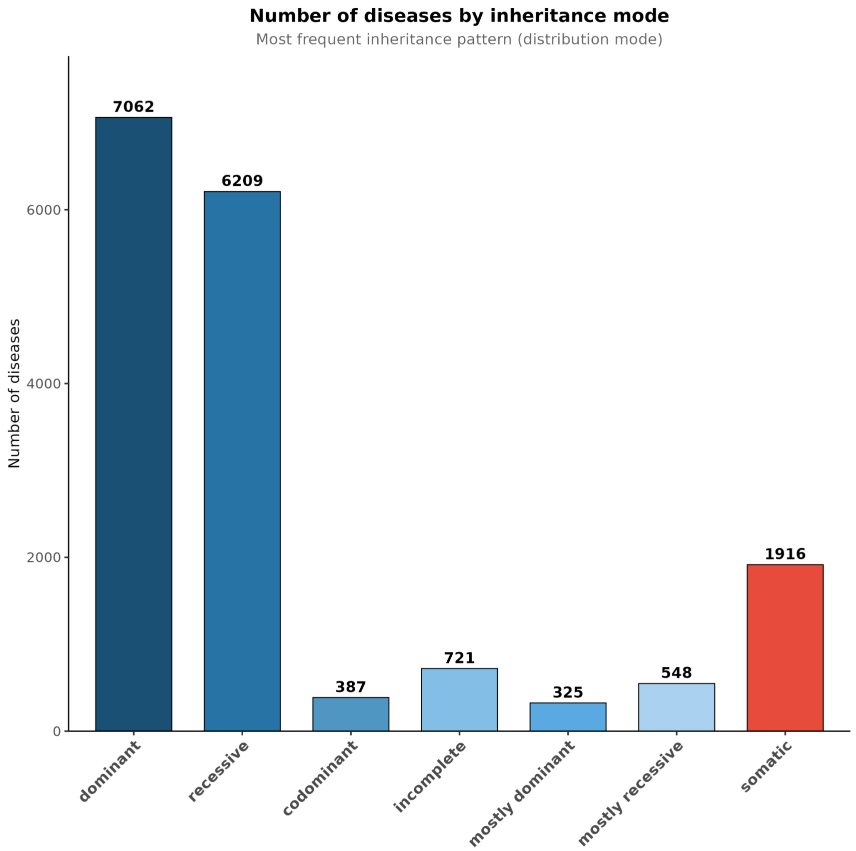


**Supplementary Figure 22 | Distribution of inheritance modes among high-confidence gene-disease associations (Definitive and Strong).**

We further characterized high-confidence gene–disease associations according to penetrance, age of onset, and disease severity. Penetrance analysis showed that most associations were classified as fully penetrant (n=7,122), followed by complex disease associations (n=4,424), moderately penetrant associations (n=3,371), and highly penetrant associations (n=2,250) (**Supplementary Figure 23**). It should be noted that the proposed agentic pipeline is optimized for the identification of high-penetrance gene–disease associations. Consequently, complex disease associations are underrepresented in our results, as many association studies—such as genome-wide association studies (GWAS)—primarily report their findings in supplementary tables and figures rather than in abstracts or titles, which constitute the input of our analysis. Complementary retrieval of complex disease associations could be achieved by integrating curated association aggregation resources (e.g., Open Targets), which is beyond the scope of this study, as our focus is on high-penetrance genes.

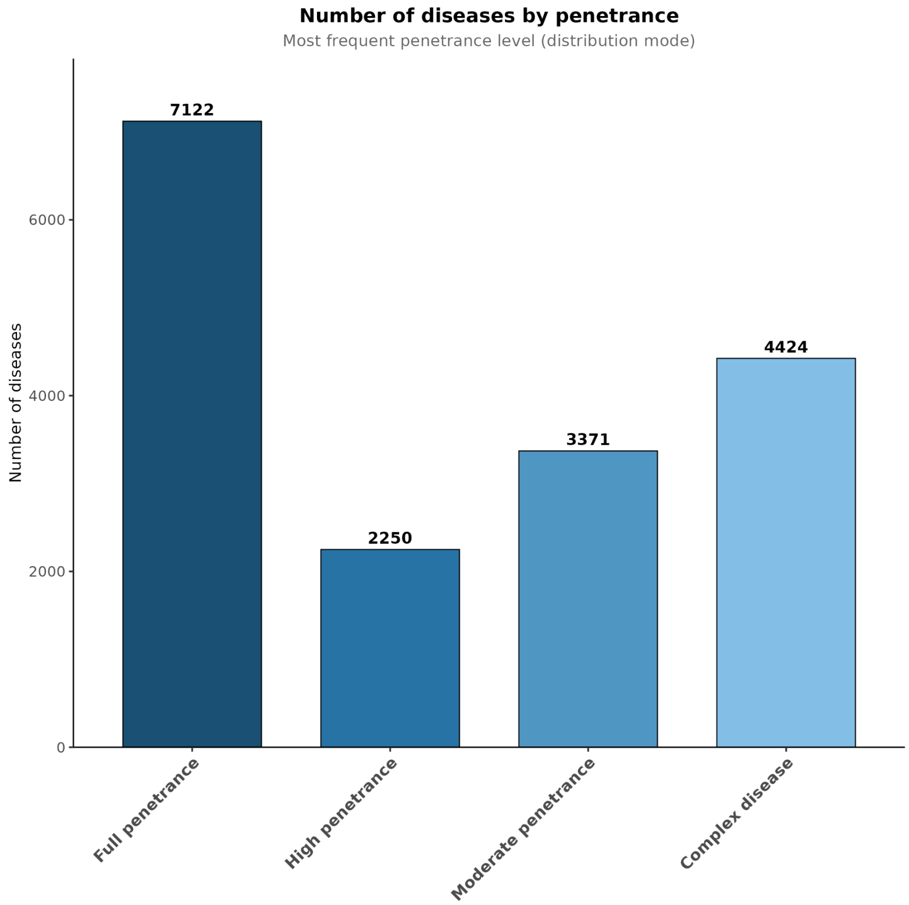


**Supplementary Figure 23 | Distribution of penetrance categories among high-confidence gene–disease associations (Definitive and Strong).**

Regarding age of onset, adulthood onset was most prevalent (n=4,800), followed by late-onset diseases (n=3,009), infancy (n=2,843), and childhood (n=2,624) onset. Prenatal onset accounted for 1,876 associations, while neonatal (n=1,045) and adolescence (n=921) onset were less common (**Supplementary Figure 24**).

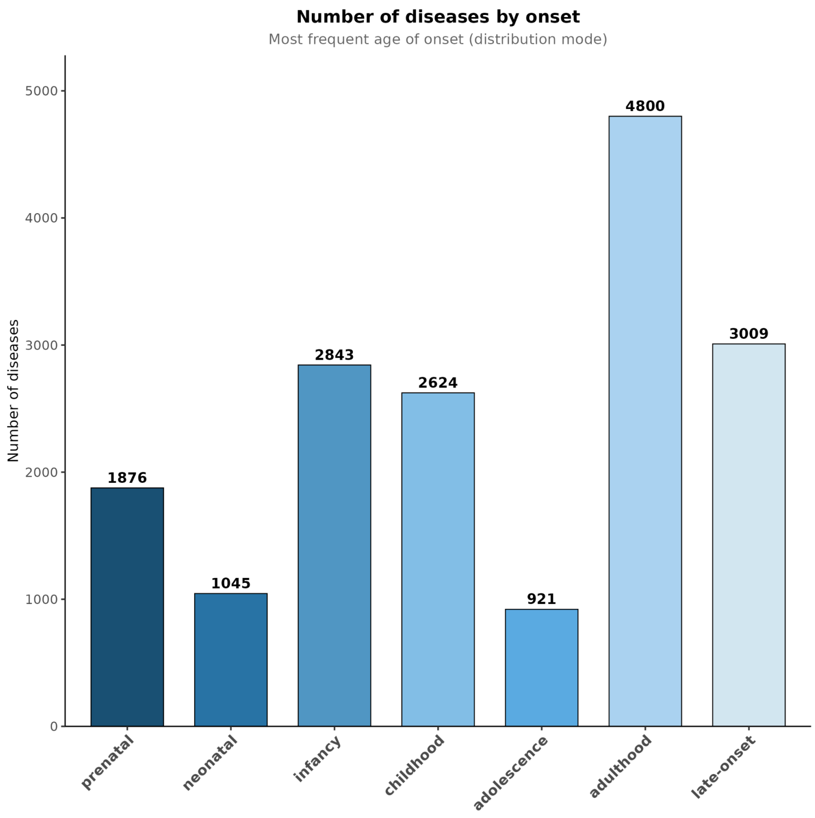


**Supplementary Figure 24 | Distribution of age-of-onset categories among high-confidence gene–disease associations (Definitive and Strong).**

Disease severity distribution showed that moderate severity was most frequent (n=5,227), followed by critical severity (n=4,984). Severe and mild phenotypes were observed in 3,471 and 3,076 associations, respectively, while very mild presentations were rare (n=360) (**Supplementary Figure 25**).

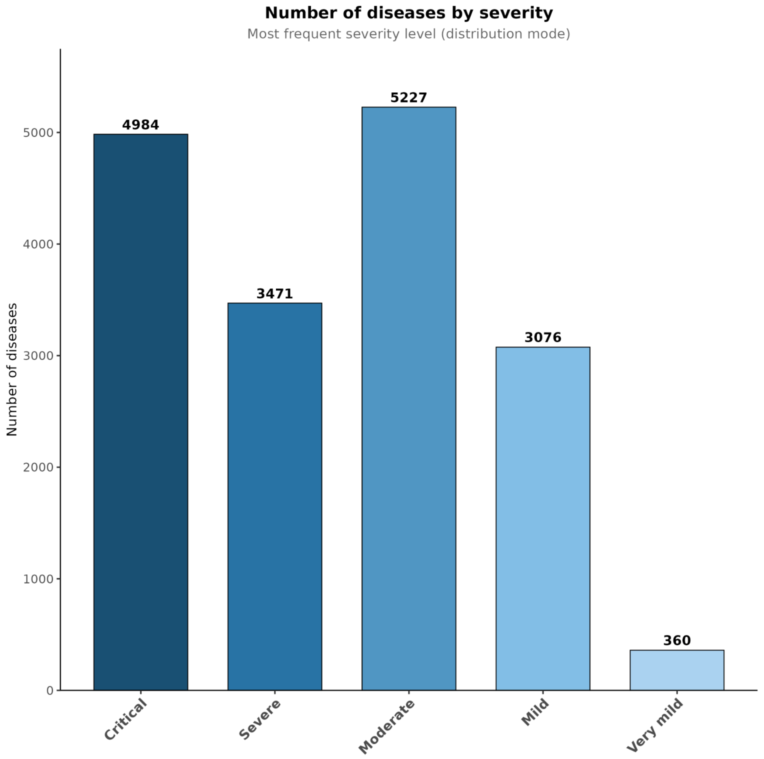


**Supplementary Figure 25 | Distribution of disease severity among high-confidence gene–disease associations (Definitive and Strong).**

#### Joint analysis of gene-diseases association categories

We then performed a contingency analysis to explore combinations across the different categories.

Examination of the contingency table relating inheritance mode and molecular mechanism (**Supplementary Figure 26**) shows that LoF associations are distributed across dominant and recessive inheritance with comparable proportions, with a slight predominance of recessive inheritance. Specifically, 50.1% of LoF associations are recessive, 35.3% are dominant, and the remaining 14.6% fall under other inheritance modes. In contrast, GoF and DN mechanisms are predominantly associated with dominant inheritance, accounting for 46.4% of GoF and 90.0% of DN associations, respectively.

The presence of a small number of recessive DN associations (n=15) is potentially unexpected, as this combination appears mechanistically contradictory. While a genuine biological explanation cannot be entirely excluded, these cases are more likely attributable to misclassification by the agentic framework. Importantly, the agent outputs a confidence distribution for each prediction, allowing post hoc assessment of result reliability. Consistent with this interpretation, recessive DN associations exhibit a markedly lower confidence than dominant DN associations, as reflected by a significantly smaller mean distance between the modal prediction and the second-highest probability peak (38.07% for recessive DN versus 71.45% for dominant DN, t-test p: $8.07 \times 10^{-5}$). This pronounced difference indicates reduced model confidence for recessive DN assignments, supporting the conclusion that these cases should be interpreted with caution.

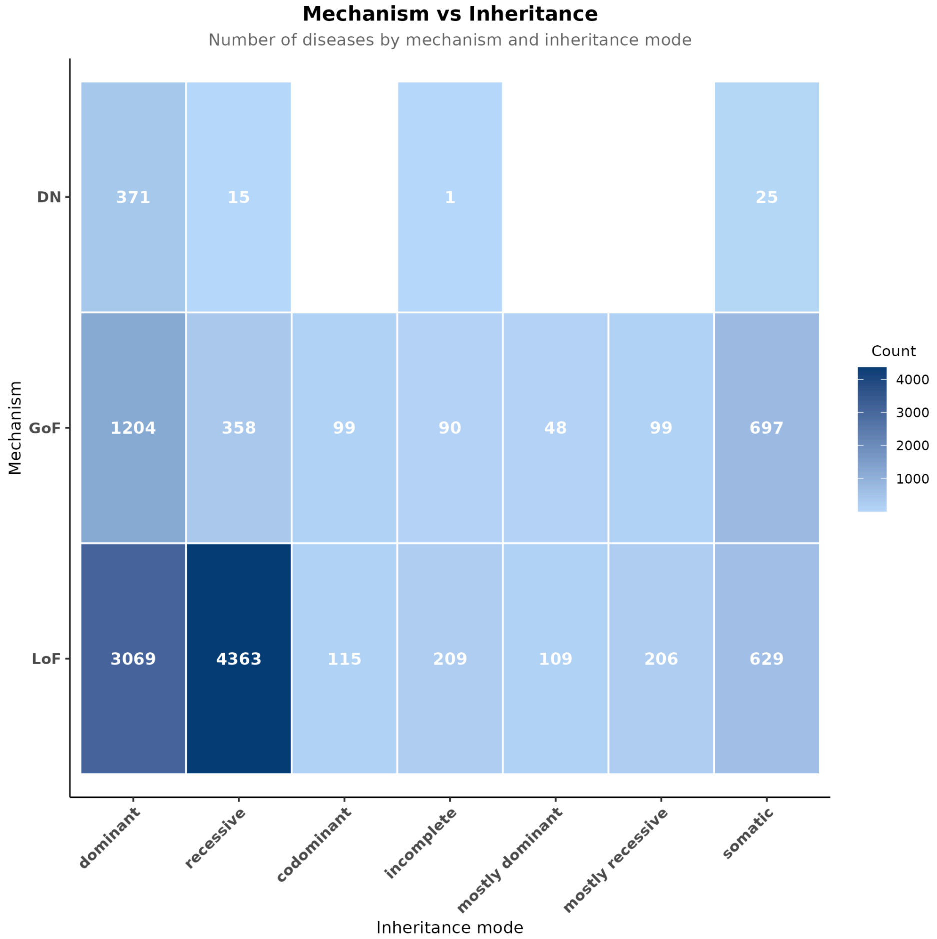


**Supplementary Figure 26 | Contingency matrix of inheritance mode and molecular mechanism among high-confidence gene–disease associations.**

The contingency table between severity and age of onset shows two peaks, one at adult onset and moderate diseases (1,837 associations) and one at infancy onset and critical disease

(1,568 associations) (**Supplementary Figure 27**). Early onset diseases are also on average more penetrant than late onset diseases (**Supplementary Figure 28**).

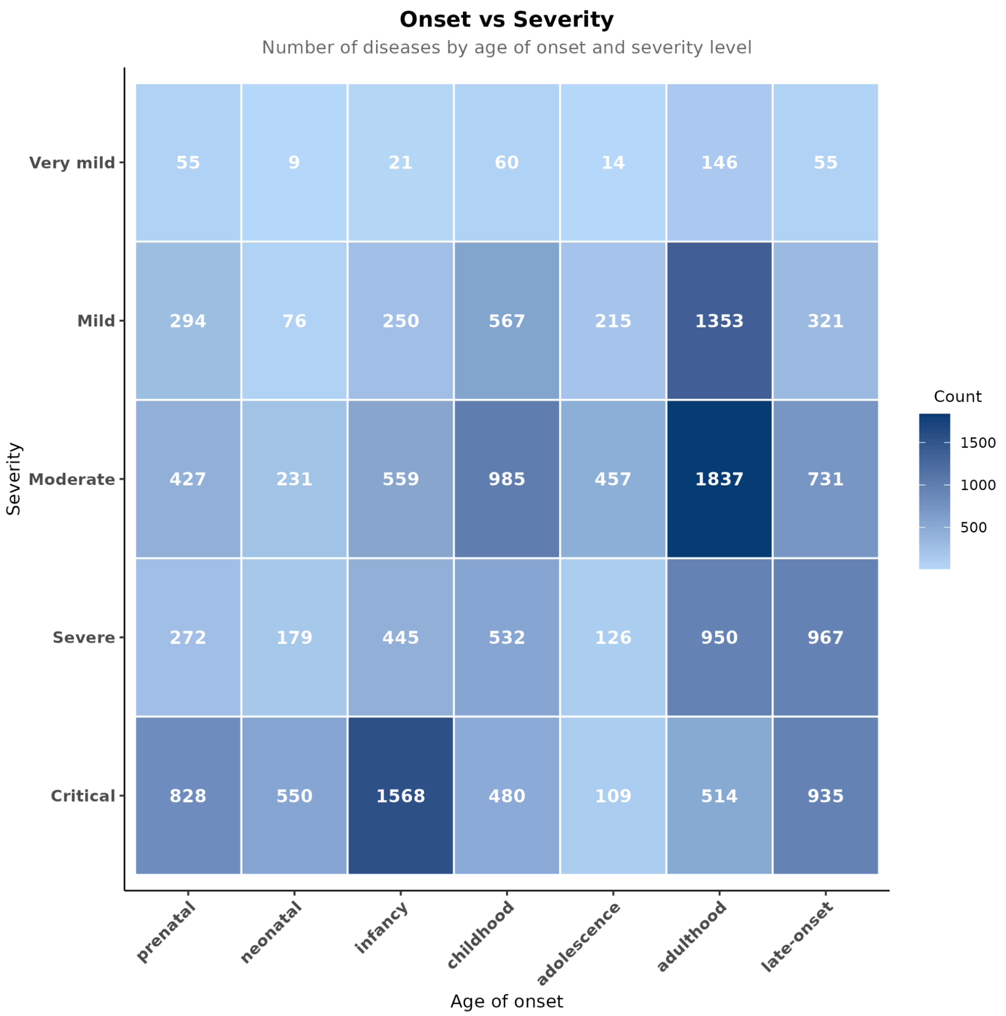


**Supplementary Figure 27 | Contingency matrix of age of onset and disease severity among high-confidence gene–disease associations.**

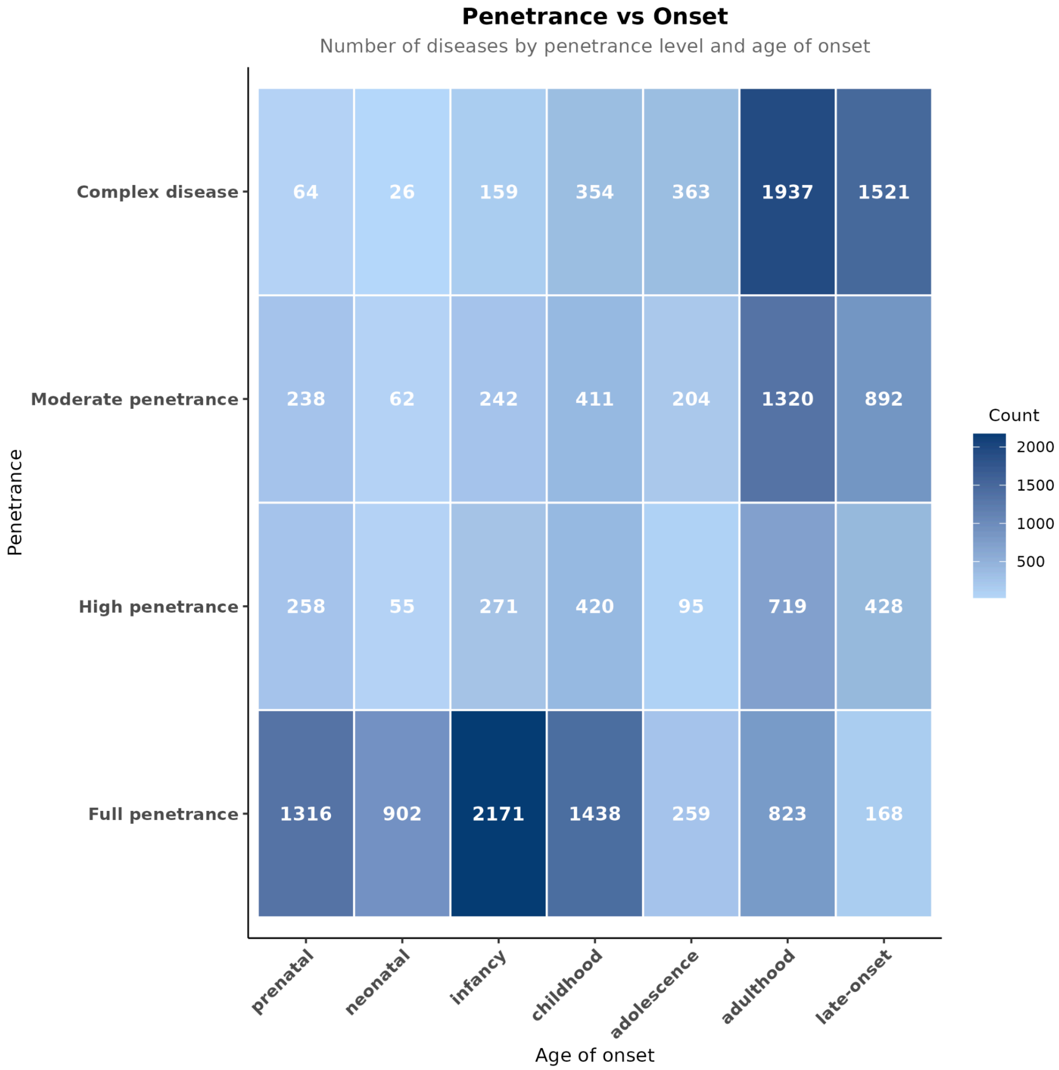


**Supplementary Figure 28 | Contingency matrix of penetrance level and age of onset among high-confidence gene–disease associations.**

### XGBoost Feature-Based Prediction of Clinical Impact

Jeremy Guez, Heidi Rehm, Kaitlin Samocha, Konrad Karczewski

#### Rationale for XGBoost-Based PEPPER Prediction

A direct comparison between literature-derived clinical scores (e.g., PEPPER) and population-based constraint metrics (e.g., LOEUF) on disease gene benchmarks presents a fundamental methodological challenge: benchmark gene lists were curated by experts using the same literature that informs our LLM-based scoring. This circularity means that any observed improvement may reflect accurate literature curation rather than true predictive power. Importantly, faithful literature curation is itself valuable, demonstrating that LLMs can help experts systematically extract clinical knowledge from publications. However, this shared information source prevents fair benchmarking against constraint-based metrics that operate independently of the literature.

To address this limitation, we developed a machine learning approach that predicts PEPPER from gene-level biological features entirely independent of literature content. By training an XGBoost model[57] on features derived from protein sequence, expression patterns, protein-protein interactions, and evolutionary conservation, we effectively distill the clinical knowledge encoded in publications into a generalizable predictive model. We termed the resulting out-of-fold predicted score $PEPPER_{XGB}$. This approach offers two key advantages:

1. Unbiased benchmarking: For any given gene's prediction, the model was never trained on that gene's literature-derived score (through cross-validation), enabling fair comparison with constraint-based metrics.

2. Generalization to understudied genes: The model can predict clinical impact for genes with limited or no literature coverage by leveraging their biological properties.

### XGBoost Model Architecture and Training

#### Feature Set

We used 1,118 gene-level features previously derived[15], organized into the categories listed in **Supplementary Table 16**.

**Supplementary Table 16 | Categories for gene-level features used in the XGBoost model**

| Category | Feature Count | Description |
|---|---|---|
| Gene Expression | 818 | Features derived from 77 bulk and single-cell RNA-seq datasets across 24 tissue/cell type categories, including PCA/ICA loadings and differential expression statistics |
| Protein Embedding | 204 | Learned representations from ProtT5, a protein language model trained on protein sequences, capturing biophysical and functional properties |
| Coexpression | 51 | Gene connectivity in coexpression networks derived from GTEx, representing correlation patterns with highly variable genes |
| Gene Regulatory | 16 | Enhancer-gene links predicted by Activity-By-Contact (ABC) method, promoter counts from FANTOM, and Roadmap Epigenomics features |
| Conservation | 14 | PhastCons scores across vertebrate alignments (7-100 species) and constraint metrics from the Zoonomia project (240 mammals, 43 primates) |
| Gene Structure | 7 | Transcript count, exon count, CDS length, and UTR properties derived from GENCODE annotations |
| PPI | 3 | Connectedness measures in protein-protein interaction networks, weighted by interaction confidence scores |
| Missense Constraint | 1 | UNEECON-G score measuring gene-level intolerance to missense variants[58] |

The full feature set (1,247 features) additionally includes 129 GO term and pathway annotations, which we excluded from the primary analysis to avoid reliance on manually curated functional knowledge. Generally, the GO configuration slightly outperforms the "No GO" configuration.

#### Model Configuration

We used XGBoost (eXtreme Gradient Boosting) regression with the hyperparameters listed in **Supplementary Table 17**.

**Supplementary Table 17 | Hyperparameters used in the XGBoost regression.**

| Parameter | Value | Rationale |
|---|---|---|
| n_estimators | 80 | Sufficient for convergence with early stopping |
| learning_rate | 0.05 | Conservative learning rate to prevent overfitting |
| max_depth | 3 | Shallow trees to encourage generalization |
| subsample | 0.85 | Row subsampling to reduce variance |
| colsample_bytree | 1.0 | Use all features at each tree |
| early_stopping_rounds | 50 | Halt training if validation loss plateaus |

#### Cross-Validation Strategy

We employed 5-fold cross-validation with random fold assignment. For each fold:
1. The model was trained on 4 folds (80% of genes)
2. Predictions were generated for the held-out fold (20% of genes)
3. SHAP (SHapley Additive exPlanations) values were computed for all test genes

This procedure generates out-of-fold predictions for every gene in the dataset, ensuring that each gene's predicted $PEPPER_{XGB}$ was generated by a model that never observed its true literature-derived score during training.

### Feature Importance Analysis

#### Global Category Importance

SHAP value analysis reveals the relative contribution of each feature category to clinical impact prediction (**Supplementary Table 18**). Missense constraint (UNEECON-G[58]) emerges as the single strongest predictor category (23.1%) despite being a single feature, underscoring the critical importance of selection pressure on protein-altering variants. Protein sequence embeddings (22.8%) and tissue-specific gene expression patterns (21.0%) contribute comparably, followed by gene structure features such as transcript count and CDS length (16.1%).

**Supplementary Table 18 | XGB features SHAP analysis.**

| Category | Total SHAP | Percentage |
|---|---|---|
| Missense constraint | 0.0208 | 23.1% |
| Protein embeddings | 0.0205 | 22.8% |
| Gene expression | 0.0188 | 21.0% |
| Gene structure | 0.0144 | 16.1% |
| PPI | 0.0083 | 9.3% |
| Coexpression | 0.0031 | 3.5% |
| Gene regulatory | 0.0030 | 3.3% |
| Conservation | 0.0007 | 0.7% |
| Pathways (non-GO) | 0.0001 | 0.1% |
| Subcellular localization | 0.0001 | 0.1% |

#### Top Individual Features

The most predictive individual features are listed in **Supplementary Table 19**. UNEECON-G, which captures missense variant intolerance, ranks as the single most informative feature, reflecting the strong link between protein-level selection pressure and disease relevance. PPI degree (the number of protein interaction partners) ranks second, consistent with the biological principle that hub proteins in interaction networks tend to be functionally important. Notably, five of the top ten individual features are protein sequence embedding dimensions, suggesting that learned protein representations capture disease-relevant structural and functional properties not fully encoded by other feature categories.

**Supplementary Table 19 | Predictive features from the XGBoost.**

| Rank | Feature | Mean absolute SHAP | Category |
|---|---|---|---|
| 1 | UNEECON-G | 0.0208 | Missense constraint |
| 2 | PPI_degree_decile | 0.0083 | PPI |
| 3 | CDS_length | 0.0075 | Gene structure |
| 4 | Transcript_count | 0.0064 | Gene structure |
| 5 | protdim_429 | 0.0040 | Protein embedding |
| 6 | protdim_444 | 0.0025 | Protein embedding |
| 7 | EDS | 0.0025 | Gene regulatory |
| 8 | protdim_23 | 0.0015 | Protein embedding |
| 9 | protdim_392 | 0.0014 | Protein embedding |
| 10 | protdim_591 | 0.0010 | Protein embedding |

#### Integration with Bayesian Scoring

$PEPPER_{XGB}$ values are integrated with LOEUF-MIS through a Bayesian framework, yielding the composite score $OMELET_{XGB}$. Briefly, $PEPPER_{LLM}$ or $PEPPER_{XGB}$ can both serve as the prior probability of clinical impact, while LOEUF-MIS provides the likelihood based on observed constraint.

$OMELET_{XGB}$ substantially outperforms LOEUF-MIS alone on the NDD benchmark (PRAUC 0.395 vs 0.265), demonstrating that biological features encode clinically relevant information that complements population-level constraint estimates.

**Supplementary Figure 29** shows the relationship between LOEUF-MIS and $OMELET_{XGB}$. Genes above the diagonal represent cases where $PEPPER_{XGB}$ elevates the final score beyond what LOEUF-MIS alone would suggest—these are genes whose biological properties indicate clinical importance despite moderate constraint levels.

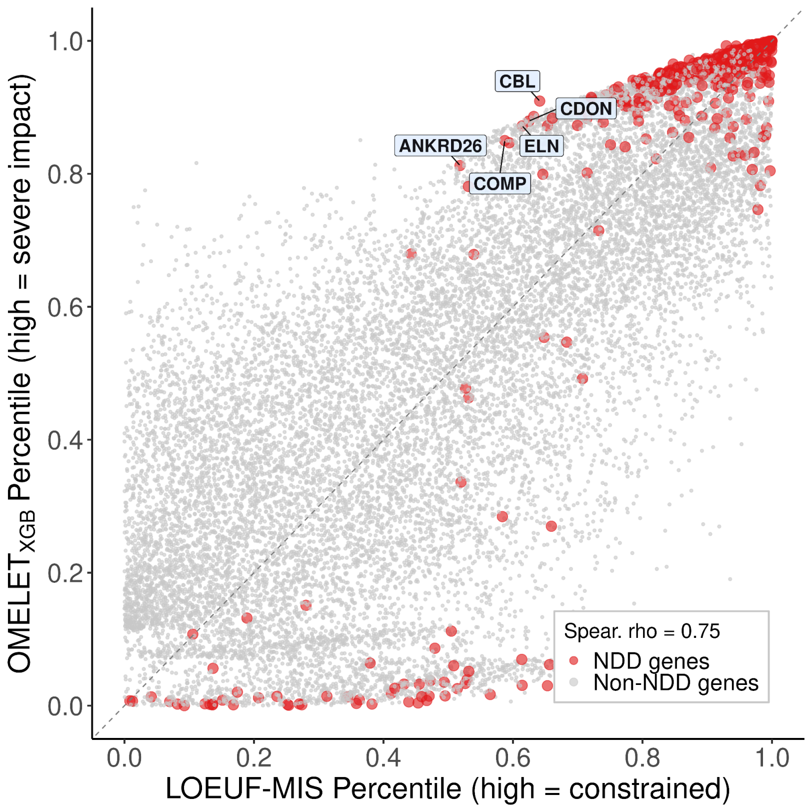


**Supplementary Figure 29 | OMELET$_{XGB}$ percentile vs LOEUF-MIS percentile.**
Labeled genes are predicted to have a high clinical impact by the prior PEPPER$_{XGB}$, but are not flagged as constrained by LOEUF-MIS.

### Case Study: *ACVR1*

#### Clinical Context

*ACVR1* (Activin A Receptor Type 1, also known as *ALK2*) exemplifies a gene where constraint-based metrics fail to capture clinical severity, while PEPPER$_{LLM}$ and PEPPER$_{XGB}$ succeed. *ACVR1* mutations cause Fibrodysplasia Ossificans Progressiva (FOP), an autosomal dominant disorder characterized by progressive heterotopic ossification [59].

FOP is caused exclusively by gain-of-function mutations, predominantly the R206H variant. The pathogenic mechanism involves dysregulated BMP signaling: in normal conditions, Activin A does not activate ACVR1 for SMAD1/5 signaling and may even form a non-signaling complex that buffers the signal[60]. With the FOP mutation, Activin A aberrantly activates ACVR1, triggering SMAD1/5 phosphorylation (a "BMP-like" signal) that drives heterotopic ossification[61].

#### Not Detected by Constraint Metrics

Because FOP results from GoF rather than LoF mutations, standard constraint metrics fail to detect *ACVR1*'s pathogenicity (**Supplementary Table 20**). LOEUF measures intolerance to loss-of-function variants, which is not relevant for a gain-of-function disease. LOEUF-MIS captures missense constraint but depends on the accuracy of the underlying missense effect predictors (PopEVE, AlphaMissense, ESM1v) and dilutes with LoFs the constraint present at the GoF critical residues, such as R206.

**Supplementary Table 20 | Constraint scores for *ACVR1.***

| Metric | Value | Interpretation |
|---|---|---|
| LOEUF v4 (LoF constraint) | 0.92 | Not constrained |
| LOEUF-MIS (LoF and missense constraint) | 0.8 | Weakly constrained |
| LLM-derived PEPPER (percentile) | 99.3 | High clinical impact (correctly identified) |
| XGBoost-predicted PEPPER (percentile) | 90.6 | High clinical impact (recovered from features) |

#### XGBoost Feature Attribution

SHAP analysis reveals how XGBoost correctly identifies *ACVR1* as clinically important despite weak constraint signals. The top contributing features for *ACVR1* are listed in **Supplementary Table 21**.

**Supplementary Table 21 | Top contributing XGBoost features for *ACVR1.***

Note: Positive SHAP values indicate features that increase predicted clinical impact.

| Feature | Weight | Interpretation |
|---|---|---|
| UNEECON-G | +0.037 | Strong missense constraint at critical residues, consistent with gain-of-function pathogenicity |
| Transcript_count | +0.024 | High transcript diversity indicates regulatory complexity |
| protdim_392 | +0.006 | Protein embedding dimension |
| protdim_429 | -0.006 | Protein embedding dimension |

| | | |
|---|---|---|
| PPI_degree_decile | +0.006 | High connectivity in protein interaction network, indicating functional importance |

#### Mechanistic Insights

In the case of *ACVR1*, XGBoost identifies clinical importance through missense constraint and PPI connectivity, capturing the gene's role in a signaling complex where gain-of-function mutations cause disease. More broadly, this suggests that XGBoost can highlight the most relevant biological features even for less well-characterized genes by leveraging biological features when classical constraint metrics are weak. Systematic evaluation across other gain-of-function disorders could therefore establish XGBoost-based prediction as both a complement to constraint metrics and a hypothesis-generating tool for uncovering disease mechanisms in understudied genes.

### Case Study: DENND2B — Prospective Gene Discovery

#### Clinical Context

*DENND2B* (DENN Domain Containing 2B) exemplifies a scenario where XGBoost-based prediction identifies a disease gene before sufficient literature evidence accumulates. At the time of our analysis (January 2026), *DENND2B* had only 3 articles retrieved by PubMed search, no entries in GenCC, and one article explicitly stated: *"DENND2B is a DENN domain-containing protein that has important roles in regulating the cell cycle, cell division and ciliogenesis, but to date has not been associated with any human disease"*[62].The same 2026 publication reports the first disease association for *DENND2B* based on 11 individuals with monoallelic variants:

> *"Features shared among these patients include developmental delay, intellectual disability and psychiatric/behavioral concerns, and episodes of psychosis and/or catatonia. Additional features common to our cohort include epilepsy, muscle weakness/hypotonia, and a wide range of congenital anomalies across different organ systems. [...] Altogether, these findings suggest that monoallelic loss of function variants in DENND2B cause a novel autosomal dominant neurodevelopmental disorder with variable vulnerability for psychosis and/or catatonia."*[62]

#### Pipeline Performance

Our LLM pipeline correctly identified "Neurodevelopmental impairment" as a disease association from this single article. However, the limited evidence resulted in a conservative score (**Supplementary Table 22**).

**Supplementary Table 22 | PEPPER scores for *DENND2B.***

| Metric | Value | Interpretation |
|---|---|---|
| $PEPPER_{LLM}$ Percentile | 67.3% | Below median pathogenicity |
| **$PEPPER_{XGB}$ Percentile** | **95.9%** | **Top 5% of genes** |

While the LLM correctly identified the disease association, it appropriately assigned a lower confidence due to a single publication. In contrast, the XGBoost—trained on biological features from thousands of well-characterized disease genes—recognized *DENND2B*'s biological profile as highly consistent with pathogenicity.

#### XGBoost Feature Attribution

SHAP analysis reveals why XGBoost predicts high clinical impact for *DENND2B* (**Supplementary Table 23**).

**Supplementary Table 23 | XGBoost Features of *DENND2B.***
Note: Positive SHAP values indicate features that increase predicted clinical impact.

| Feature | SHAP | Interpretation |
|---|---|---|
| Transcript_count | +0.053 | High transcript diversity signals regulatory complexity |
| UNEECON-G | +0.027 | Missense constraint detected at critical residues |
| CDS_length | +0.024 | Long coding sequence typical of large developmental genes |
| protdim_429 | +0.023 | Protein embedding capturing disease-relevant structural properties |
| mouse_development_icaloadings.20 | +0.011 | Mouse developmental expression pattern consistent with developmental role |

*DENND2B*'s gene structure and missense effects dominate the prediction: high transcript count (+0.053), missense constraint, and substantial coding sequence length. These features are characteristic of developmental regulators. Additionally, mouse development ICA loadings feature further support its role in neurodevelopment, the phenotype previously reported[62].

##### Implications for Gene Discovery

This case illustrates a powerful application of XGBoost-based prediction: prospective identification of disease genes before sufficient literature evidence accumulates. The model, trained on biological features from established disease genes, effectively generalizes patterns of pathogenicity to understudied genes.

In the case of *DENND2B*:
- Above median $PEPPER_{LLM}$: The LLM pipeline correctly detected the nascent disease association but appropriately assigned low confidence.
- Above 95th percentile $PEPPER_{XGB}$: XGBoost independently predicted high pathogenicity based on biological features alone.
- A 2026 publication recently identified *DENND2B* as a novel disease gene[62].

This suggests that genes with high $PEPPER_{XGB}$ but lower $PEPPER_{LLM}$ may represent candidate disease genes awaiting discovery. Systematic identification of such genes could prioritize targets for clinical sequencing studies and functional characterization, potentially accelerating the discovery of novel Mendelian disorders.

#### Candidate Disease Gene List

##### Rationale

Building on the *DENND2B* case study, we systematically identified genes that XGBoost predicts as highly pathogenic but which lack established disease associations in curated databases (high $PEPPER_{XGB}$, low $PEPPER_{LLM}$). These genes represent candidates for novel disease gene discovery—their biological features closely resemble known disease genes, yet they remain understudied in the clinical genetics literature.

##### Selection Criteria

We applied stringent filters to identify high-confidence candidates (**Supplementary Table 24**).

**Supplementary Table 24 | Selection criteria to define candidate disease genes using GenCC and PEPPER scores.**

| Criterion | Threshold | Rationale |
|---|---|---|
| No GenCC entry | Supportive or higher | Excludes genes with established disease associations |
| $PEPPER_{XGB}$ percentile | > 95% | Top 5% predicted clinical impact |
| $PEPPER_{XGB}$ – $PEPPER_{LLM}$ | ≥ 25 percentile points | Large discrepancy between predicted and literature-derived scores |

#### Candidate Gene Characteristics

This filtering yielded 220 candidate genes (**Supplementary Dataset 1**). These candidates exhibit significantly lower LOEUF compared to other genes (median LOEUF 0.31 vs. 0.92; Wilcoxon rank-sum test, $p = 2.4 \times 10^{-87}$; **Supplementary Table 25**).

**Supplementary Table 25 | LOEUF scores for candidate genes versus other genes.** Candidate genes had significantly lower LOEUF scores than other genes (Wilcoxon rank-sum test, $p = 2.4 \times 10^{-87}$).

| Group | Median LOEUF | N with a LOEUF |
|---|---|---|
| Candidate genes | 0.31 | 219 |
| Other genes | 0.92 | 16,944 |

The striking constraint enrichment (LOEUF about 3-fold lower than background) provides independent validation that these genes are under strong purifying selection. Combined with their biological profiles captured by XGBoost, these candidates merit prioritization for:

1. Clinical sequencing studies: Variants in these genes should be carefully evaluated in unsolved rare disease cases
2. Functional characterization: Model organism studies or cellular assays to establish gene function
3. Literature monitoring: Emerging case reports may establish disease associations

The complete list is available in **Supplementary Dataset 1**.

### OMELET: A Bayesian Integration of Constraint and Clinical Significance Scores

Jeremy Guez, Hilary Finucane, Heidi Rehm, Kaitlin Samocha, Konrad Karczewski

#### Overview

We introduce OMELET (Omnibus Mutation Effects with LOEUF and Embedded Texts), a Bayesian framework that integrates two orthogonal sources of evidence for gene-level pathogenicity: (i) a prior derived from a clinical impact score (PEPPER), obtained either from LLM-based literature analysis ($PEPPER_{LLM}$) or from a supervised XGBoost trained to predict $PEPPER_{LLM}$ using gene-level features ($PEPPER_{XGB}$), and (ii) a likelihood derived from population-level constraint data (LOEUF-MIS) obtained from gnomAD v4. The resulting posterior distribution provides a principled, unified score that combines text-derived or model-derived clinical knowledge with empirical mutational constraint.

We define two variants:

- $OMELET_{LLM}$: uses $PEPPER_{LLM}$ as the prior (from LLM agent literature analysis)
- $OMELET_{XGB}$: uses $PEPPER_{XGB}$ as the prior (from XGBoost out-of-fold predictions)

Both variants share the same Bayesian inference machinery described below.

#### Model Formulation

For each gene g, let $\theta_g$ in (0, 1) denote the latent "intolerance parameter", where low values of theta indicate strong constraint (i.e., depletion of observed loss-of-function variants relative to expectation).

##### Prior: Beta distribution from PEPPER

The PEPPER for gene g (denoted $s_g$) is a score in [0, 1], where higher values indicate greater predicted clinical severity. We map $s_g$ to a probability parameter $p_g$ as follows:

$$p_g = p_{min} + (1 - s_g) \times (p_{max} - p_{min})$$

where $p_{min}$ = 0.05 and $p_{max}$ = 0.95. This mapping ensures that genes with high clinical severity ($s_g$ close to 1) are assigned a low $p_g$ (strong constraint expected), while genes with low severity ($s_g$ close to 0) receive a high $p_g$ (little constraint expected).

The prior is a Beta distribution parameterized by a concentration parameter $\kappa_g$:

$$\theta_g \sim \mathrm{Beta}(\alpha_g, \beta_g)$$

where:

- $\alpha_g = \kappa_g \times p_g$
- $\beta_g = \kappa_g \times (1 - p_g)$

The mean of this prior is $p_g$, and $\kappa_g$ controls the precision (inverse variance). A larger $\kappa_g$ concentrates the prior more tightly around $p_g$, reflecting greater confidence in the PEPPER estimate.

#### Adaptive kappa from Monte Carlo variance

Since three of the five LLM agents (penetrance, inheritance, and onset/severity) output probability distributions over their respective parameters, we use Monte Carlo sampling to propagate this uncertainty through the scoring function, yielding a distribution of the PEPPER score. The concentration parameter $\kappa_g$ is computed adaptively from the variance of this resulting distribution. Given the variance $\sigma_g^2$ of the Monte Carlo PEPPER samples for gene g:

$$\kappa_g = [\ p_g \times (1 - p_g)\ ] \ /\ [\ \sigma_g^2 \times b^2\ ] - 1$$

where b = 0.90 (the range of the probability mapping, i.e., $p_{max} - p_{min}$). This formula derives from the variance of a Beta distribution:

$$\mathrm{Var}[\theta] = \alpha \times \beta \ /\ [\ (\alpha + \beta)^2 \times (\alpha + \beta + 1)\ ] = p\ (1 - p) \ /\ (\kappa + 1)$$

where $p = \alpha / (\alpha + \beta)$ is the mean and $\kappa = \alpha + \beta$ is the concentration. Combined with the chain-rule propagation of the score variance through the affine mapping $p_g = p_{min} + (1 - s_g) \times b$, solving for κ yields the expression above. Thus, genes for which the LLM agents show high agreement (low $\sigma_g^2$) receive a large $\kappa_g$, producing a concentrated prior, while genes with uncertain scores receive a diffuse prior that defers to the gnomAD likelihood.

#### Scaling parameter ζ

The raw $\kappa_g$ reflects the internal confidence of the LLM agents but does not encode our external confidence in the LLM framework relative to population constraint data. We therefore introduce a scaling parameter ζ such that the effective concentration becomes $\kappa'_g = \zeta \times (\kappa_g + 1) - 1$, which is mathematically equivalent to dividing the Monte Carlo variance by ζ before computing $\kappa_g$.

- For the curation task ($OMELET_{LLM}$), we expect the literature-derived PEPPER score to be more informative than the gnomAD constraint alone, motivating $\zeta > 1$. This is because $PEPPER_{LLM}$ parses the literature, and hence has direct access to the knowledge necessary for curation. We explored ζ over a range of values and found that PRAUC on the NDD curation benchmark remains stable for $\zeta \in [20, 50]$ (range: 0.683–0.686), indicating that the model is robust to the precise choice of this parameter. We selected $\zeta = 30$ for the main analysis.

- For disease-gene prediction ($OMELET_{XGB}$), since $PEPPER_{XGB}$ is trained to approximate the LLM-derived scores, the LLM Monte Carlo variance remains the relevant measure of epistemic uncertainty about each gene's clinical characterization in the literature. We therefore use the raw LLM-derived $\kappa_g$, with $\zeta = 1$, as the XGBoost predictions do not benefit from the same direct access to literature evidence that would justify upweighting the prior relative to gnomAD data. Future work could derive a specific κ for $OMELET_{XGB}$, combining model-specific prediction uncertainty from the XGBoost ensemble with the LLM uncertainty.

In both cases ($OMELET_{LLM}$ and $OMELET_{XGB}$), outlier values exceeding $\kappa_g = 1000$ are clipped to prevent any single gene's prior from dominating the posterior.

#### Likelihood: Poisson model from LOEUF-MIS

The likelihood is derived from the observed and expected counts of missense variants from gnomAD v4. For each gene g, let:

- $O_g$: observed number of pLoFs and worst missense variants (average of $O_g$ at 99 percentile for PopEVE, AlphaMissense and ESM1v)
- $E_g$: expected number of pLoFs and worst missense variants under a neutral mutation model (average of $E_g$ at 99 percentile for PopEVE, AlphaMissense and ESM1v)

We model the observed count as:

$$O_g \mid \theta_g \sim \text{Poisson}(E_g \times \theta_g)$$

When $\theta_g < 1$, fewer variants are observed than expected (constraint); when $\theta_g > 1$, more are observed (possible positive selection or relaxed constraint). This likelihood naturally captures the ratio O/E (which is the LOEUF-MIS score) in a probabilistic framework that properly accounts for gene size through $E_g$.

#### Posterior

By Bayes' theorem, the posterior distribution of $\theta_g$ is:

$$P(\theta_g \mid O_g) \sim P(O_g \mid \theta_g) \times P(\theta_g)$$

Since neither the Beta-Poisson conjugacy admits a closed-form posterior, we compute it numerically on a discrete grid of N = 50 points spanning (ε, 1 - ε) where $\varepsilon = 10^{-6}$:

$$\log P(\theta_j \mid O_g) = \log P(O_g \mid \theta_j) + \log P(\theta_j) + \text{const}$$

for j = 1, ..., N. We then normalize the posterior by exponentiating (with log-sum-exp for numerical stability) and summing to unity.

#### Summary statistic

The final OMELET score for gene g is the 95th percentile (q95) of the posterior distribution:

$$\text{OMELET}_g = Q_{0.95}(\theta_g \mid O_g)$$

where $Q_{0.95}$ denotes the 95th percentile, computed via linear interpolation on the cumulative distribution function (CDF) of the discretized posterior. The q95 was chosen as the summary statistic because it provides a conservative upper bound on the intolerance parameter. A low q95 value indicates that, even under the most permissive credible interpretation of the data, the gene remains clinically relevant.

### Inputs and Data Sources

#### $PEPPER_{LLM}$ (Clinical impact score from LLM agents)

The $PEPPER_{LLM}$ is derived from a multi-agent LLM pipeline that systematically analyzes the biomedical literature for each gene. Multiple specialized agents assess disease association, inheritance patterns, penetrance, severity, and age of onset. The resulting score (scale 0-1, where 1 = most severe) is obtained via Monte Carlo sampling over multiple independent LLM runs, yielding both a mean score and a variance estimate that informs the adaptive κ.

#### $PEPPER_{XGB}$ (Clinical Impact Score from XGBoost)

The $PEPPER_{XGB}$ is the out-of-fold prediction from a supervised XGBoost model trained to predict $PEPPER_{LLM}$. Features include gene-level attributes excluding LOEUF to ensure independence from the likelihood term. The same κ derived from $PEPPER_{LLM}$ uncertainty is used.

#### LOEUF-MIS

The LOEUF-MIS (Loss-of-function Observed/Expected Upper bound Fraction for missense variants) is computed from gnomAD v4.1. Specifically, the observed (O) and expected (E) are the sum of LoFs and worst missenses O and E (average of 99 percentile for PopEVE, AlphaMissense and ESM1v). The 95th upper bound of O/E constitutes the LOEUF-MIS metric, but in our Bayesian framework we use O and E separately to properly model uncertainty.

### Implementation Details

- Grid resolution: N = 50 points for score computation.
- Numerical stability: All computations are performed in log-space. The log-sum-exp trick is used for normalization. Non-finite values are handled gracefully with fallback to a uniform posterior.
- CDF quantiles: Computed via linear interpolation between adjacent grid points for smooth quantile estimation.

### Discovery Potential (DisPo) Score: Definition, Computation, and Validation Analyses

Jeremy Guez, Hilary Finucane, Heidi Rehm, Kaitlin Samocha, Konrad Karczewski

#### Overview

The Discovery Potential (DisPo) score quantifies the discordance between two independent lines of evidence for gene-level pathogenicity: (i) population-level predicted loss-of-function (pLoF) constraint from gnomAD v4 (LOEUF or LOEUF-MIS), and (ii) literature-derived predicted LoF clinical impact from the LLM agentic framework ($PEPPER_{LLM\text{-}pLoF}$). A high DisPo score identifies genes for which gnomAD constraint substantially exceeds documented clinical severity; i.e., genes that are strongly constrained in the population but underrepresented in the clinical literature, suggesting that significant disease-relevant biology remains to be discovered.

#### Mathematical Definition of the DisPo Score

The DisPo score is computed within the same Bayesian framework used for OMELET (see the prior section on OMELET). However, we use $PEPPER_{LLM\text{-}pLoF}$ — the loss-of-function–specific component of the PEPPER score — rather than the full $PEPPER_{LLM}$, and compare it to LOEUF (not LOEUF-MIS), so that both the prior and the likelihood reflect the same biological signal: predicted loss-of-function intolerance. Rather than combining the prior and likelihood into a posterior distribution as in OMELET, DisPo quantifies the standardized discordance between the two

##### Prior and Likelihood as Distributions over θ

For each gene g, the Bayesian model defines a latent intolerance parameter $\theta_g$ in (0, 1). We compute:

- The prior distribution $P_{prior}(\theta)$, a Beta distribution derived from $PEPPER_{LLM\text{-}LoF}$, reflecting literature-based clinical severity.
- The likelihood function $L(\theta \mid O_g, E_g)$, a Poisson model derived from gnomAD observed/expected variant counts.

Both distributions are evaluated on a discrete grid of N = 50 points and normalized to sum to unity, yielding two probability mass functions over the same theta grid.

#### Centers of Mass

We compute the mean (center of mass) of each normalized distribution:

```
None
mu_prior = sum_j [ theta_j * P_prior(theta_j) ]
mu_lik   = sum_j [ theta_j * L_norm(theta_j) ]
```

where L_norm is the normalized likelihood and the sum runs over all grid points j = 1, ..., N.

A low $\mu_{prior}$ indicates that the literature predicts strong pathogenic constraint (theta expected to be low), while a low $\mu_{lik}$ indicates that gnomAD data shows strong observed constraint.

#### Variances

The variances of each distribution are:

```
None
sigma²_prior = sum_j [ (theta_j - mu_prior)² * P_prior(theta_j) ]
sigma²_lik   = sum_j [ (theta_j - mu_lik)² * L_norm(theta_j) ]
```

These variances capture the uncertainty (spread) of each source of evidence.

#### Signed Disagreement (DisPo)

The signed disagreement is defined as the standardized difference between the prior and likelihood centers of mass:

```
None
DisPo = (mu_prior - mu_lik) / sqrt(sigma²_prior + sigma²_lik +
epsilon)
```

where epsilon = $10^{-12}$ is a small constant for numerical stability.

**Interpretation:**

- DisPo > 0: The prior mean exceeds the likelihood mean, meaning the gnomAD data indicates stronger constraint (lower theta) than the literature predicts. The gene is more constrained in the population than its clinical profile suggests—high discovery potential.
- DisPo < 0: The literature predicts stronger pathogenicity than what gnomAD constraint alone would suggest.
- DisPo ≈ 0: Agreement between the two sources.

##### DisPo Percentile

For visualization and cross-gene comparisons, the raw DisPo score can be converted to a percentile rank across all genes with valid scores:

```None
DisPo_percentile = percentile_rank(DisPo) * 100
```

where percentile_rank assigns each gene its rank divided by the total number of genes (percent_rank function in R). This yields a score in [0, 100] where higher values indicate greater discovery potential.

#### Relationship to OMELET

The DisPo score and the OMELET score are computed within a similar Bayesian framework but capture different information:

- **OMELET:** the 95th percentile (q95) of the posterior distribution $P(\theta \mid O_g)$, which integrates both sources of evidence into a single pathogenicity score.
- **DisPo:** the standardized signed difference between the prior and likelihood means, which measures the discordance between the two sources.

The difference is that the DisPo score specifically uses the $PEPPER_{LLM\text{-}LoF}$ variant of the prior, because LoF constraint (LOEUF) is the gnomAD metric being compared, while $OMELET_{LLM}$ uses the overall $PEPPER_{LLM}$.

### Validation Analyses

#### Temporal Trend in GenCC Submissions

To assess whether the DisPo score reflects the evolving discovery of gene–disease associations, we examined its relationship with the timing of submissions to GenCC. For each gene with both a valid DisPo score and an annotated GenCC submission date, we evaluated how DisPo percentiles vary over time.

We first curated GenCC submissions by retaining the earliest reported submission date for each gene, thereby capturing the initial point of gene–disease association. DisPo percentiles were computed across all 17,112 genes with valid DisPo values. These data were then integrated with the GenCC dataset and aggregated by year of first submission to obtain the mean DisPo percentile per year.

To quantify the relationship between time and DisPo score, we applied Spearman's rank correlation between the year of submission and the corresponding mean DisPo percentile. This analysis revealed a strong positive correlation (Spearman's $\rho = 0.96$, $p = 1.9 \times 10^{-6}$), indicating that genes associated with disease more recently tend to have higher DisPo scores.

This temporal trend is consistent with the interpretation that high-DisPo genes are enriched for disease associations that are being uncovered progressively, reflecting ongoing discovery of genes with subtler or more complex pathogenic mechanisms.

#### Mouse Phenotype Validation

We compared DisPo percentile distributions across three gene sets:

- **Mouse infertility genes:** 622 genes with mouse infertility phenotypes (MP:0001922, MP:0001923, MP:0001924 from the Mouse Genome Informatics database)
- **Mouse embryonic lethal genes:** 2,710 genes with embryonic lethality phenotype from MGI

- **GenCC disease genes:** 2,660 genes with Definitive and Strong evidence in GenCC, excluding fertility-only genes and embryonic lethal genes to avoid overlap

The DisPo percentile for each gene was computed as described above. Distributions were compared using the Wilcoxon rank-sum test (one-sided, testing whether mouse phenotype genes have higher DisPo than disease genes).

**Results:**

- Mouse infertility median DisPo percentile: 56.8
- Mouse embryonic lethal median DisPo percentile: 61.9
- GenCC disease gene median DisPo percentile: 41.7
- Mouse infertility vs. disease genes: $p = 4.19 \times 10^{-16}$
- Mouse embryonic lethal vs. disease genes: $p = 7.07 \times 10^{-58}$

#### Tissue-Specific Expression Enrichment

To investigate the biological basis of high DisPo scores, we performed enrichment analyses using a LOEUF-matched case-control design.

##### LOEUF-Matched Design

Because LOEUF and DisPo are correlated (high-DisPo genes tend to be constrained), naive enrichment tests would be confounded by LOEUF. To address this, we used a 1:1 matching design:

- **Cases (TOP):** Genes with DisPo >= 6
- **Controls:** Genes with DisPo in [-6, 6], matched 1:1 to cases on LOEUF (tolerance: +/- 0.01)

The matching algorithm proceeds as follows:

1. Sort case genes by decreasing |DisPo|.
2. For each case gene, find all unmatched control genes within LOEUF tolerance (± 0.01).
3. Among candidates, select the control with DisPo closest to zero.
4. Mark the selected control as used (without replacement).

This ensures that any enrichment observed in cases vs. controls is attributable to the DisPo score itself, not to differences in constraint level. We excluded genes with synonymous constraint upper bound < 0.75.

##### GTEx Tissue Enrichment

For each of the 54 GTEx tissues, we tested whether high-DisPo genes were enriched among the top 10% most expressed genes. To focus on tissue-specific effects, a gene was classified as tissue-specifically expressed if its expression in the target tissue was in the top 10% but its median expression across all other tissues was below the 90th percentile.

Enrichment was assessed using Fisher's exact test, comparing the proportion of tissue-specifically expressed genes in the TOP group vs. the LOEUF-matched control group.

Among all 54 tissues, only testis showed significant enrichment (OR = 1.83, $p = 9.6 \times 10^{-5}$), indicating that fertility-related biology is a primary driver of high DisPo scores.

##### Fetal Expression Enrichment

To assess fetal-specific enrichment while removing fertility-related confounders, we excluded genes with high testis or broadly elevated adult expression (median across tissues) before LOEUF matching. We then tested whether high-DisPo genes were enriched for broad fetal expression, defined as above-median expression across all 15 fetal tissues from the Developmental Transcriptome dataset.

#### Fetal Tissue Expression Boxplots

We compared the median fetal expression (TPM) between LOEUF-matched high-DisPo genes and controls across all 15 fetal tissues (Thymus, Adrenal, Cerebellum, Cerebrum, Eye, Heart, Intestine, Kidney, Liver, Lung, Muscle, Pancreas, Placenta, Spleen, Stomach), downloaded from Cao et al 2020[63]. For each tissue, the boxplot shows the distribution of TPM values in the high-DisPo group vs. the matched control group. Significance was assessed using Wilcoxon rank-sum tests per tissue. Genes with synonymous constraint (upper bound < 0.75), high testis-specific expression, and high median adult tissues expression were excluded, to isolate the fetal expression signal from reproductive and adult confounders.

### Definitions and Applications of Key Metrics

Jeremy Guez, Katherine Chao, Matteo Tranchero, Hilary Finucane, Heidi Rehm, Kaitlin Samocha, Konrad Karczewski

The gnomAD v2 release[2] introduced LOEUF as the primary metric for quantifying gene-level intolerance to loss-of-function variation. Building on the fivefold increase in sample size afforded by gnomAD v4 and the methodological advances described in this study, we have refined LOEUF and developed a suite of new metrics that extend constraint estimation, integrate clinical knowledge from the biomedical literature, and identify genes with high discovery potential. This section first provides a concise summary of each metric's definition, then concludes with practical guidance on when to use which metric (see "Practical guidance: selecting the appropriate metric").

#### Score Summaries

##### 1. LOEUF (Loss-of-Function Observed/Expected Upper Bound Fraction)

LOEUF quantifies purifying selection against predicted loss-of-function (pLoF) variants in a given gene. The observed count of segregating pLoF variants is modeled as Poisson-distributed, and with an uninformative prior, the posterior is Gamma(shape = O + 1, scale = 1). LOEUF is defined as the 95th percentile of this posterior divided by the expected number of segregating variants under neutrality (see the LOEUF section in this Supplementary Material for full derivation). Lower values indicate stronger intolerance to loss-of-function variation; values near 1 indicate neutrality. In this paper, we showed that LOEUF is influenced by sample size and mutation rate due to saturation effects.

##### 2. LOEUF-MIS (Loss-of-Function Observed/Expected Upper Bound Fraction incorporating Missense variants)

LOEUF-MIS extends LOEUF by combining LOFTEE-2–filtered pLoF variants with the top 1% most deleterious missense variants—as scored by AlphaMissense, ESM1v, and PopEVE—to compute a joint observed/expected ratio using the same Bayesian procedure as LOEUF. This captures constraint signals missed by pLoF-only metrics, particularly in short genes and in genes where gain-of-function (GoF) or dominant-negative (DN) mechanisms place missense variants under stronger selection than pLoF variants.

#### 3. $\text{PEPPER}_{\text{LLM}}$ (Phenotype Evidence from Published Papers Extracted via Representation with Large Language Models)

$\text{PEPPER}_{\text{LLM}}$ is a gene-level clinical significance score derived from automated extraction of gene–disease associations from PubMed abstracts using five specialized LLM agents that assess disease association, penetrance, inheritance, and severity/onset. Each gene is assigned a score equal to the maximum across all its associated diseases. $\text{PEPPER}_{\text{LLM}}$ serves as a standalone measure of literature-derived clinical characterization and as a prior in the Bayesian framework used to compute OMELET scores.

#### 4. $\text{PEPPER}_{\text{XGB}}$ (XGBoost-predicted PEPPER)

$\text{PEPPER}_{\text{XGB}}$ is a literature-independent prediction of clinical significance, generated by training an XGBoost model to predict $\text{PEPPER}_{\text{LLM}}$ from gene-level biological features (transcript diversity, expression patterns, protein domains, paralogy, constraint metrics) without using curated disease labels. Fivefold cross-validation ensures independence of each gene's prediction from its own literature-derived score. $\text{PEPPER}_{\text{XGB}}$ enables prospective identification of disease genes before sufficient literature accrues and is used as a prior in the Bayesian framework to compute $\text{OMELET}_{\text{XGB}}$.

#### 5. $\text{OMELET}_{\text{LLM}}$ (Omnibus Mutation Effects with LOEUF and Embedded Texts, literature-based)

$\text{OMELET}_{\text{LLM}}$ integrates $\text{PEPPER}_{\text{LLM}}$ as a prior with LOEUF-MIS as the likelihood in a Bayesian framework. The score is defined as the 95% lower bound (5th percentile) of the resulting posterior distribution, providing a conservative estimate of gene-level clinical impact that combines literature evidence with population-level constraint.

#### 6. $\text{OMELET}_{\text{XGB}}$ (Omnibus Mutation Effects with LOEUF and Embedded Texts, XGBoost-based)

$\text{OMELET}_{\text{XGB}}$ applies the same Bayesian framework as $\text{OMELET}_{\text{LLM}}$ but uses $\text{PEPPER}_{\text{XGB}}$ as the prior instead of $\text{PEPPER}_{\text{LLM}}$, providing an estimate of clinical impact that is not confounded by publication history. This makes it suitable for unbiased benchmarking against curated gene lists and for prospective gene discovery.

#### 7. DisPo (Discovery Potential)

The Discovery Potential (DisPo) score quantifies gene-level discordance between population-based loss-of-function constraint and literature-derived clinical evidence. It is computed as the distance between $\text{PEPPER}_{\text{LLM-LoF}}$ (constructed from LoF disease associations) and gnomAD loss-of-function constraint (from observed and expected pLoF). Positive values indicate constraint exceeding documented clinical significance, suggesting unexplored disease-relevant biology; values near zero indicate concordance; negative values indicate literature evidence exceeding constraint. DisPo is designed to prioritize genes for functional characterization, particularly in under-ascertained phenotypic categories such as embryonic lethality and infertility.

#### Summary of Metrics

**Supplementary Table 26 | LOEUF scores for candidate genes versus other genes.**

| Metric | Inputs | Measures | Primary Application |
|---|---|---|---|
| **LOEUF** | gnomAD pLoF variant counts, mutation model | LoF intolerance (purifying selection) | Disease gene prioritization, variant classification |
| **LOEUF-MIS** | gnomAD pLoF + top 1% deleterious missense (average AM, PopEVE, ESM1v) | Combined LoF and missense constraint | Disease gene prioritization, variant classification, with increased power for short genes and GoF/DN disease genes |
| **$\text{PEPPER}_{\text{LLM}}$** | PubMed abstracts via agentic LLM framework | Literature-derived clinical significance | Bayesian prior; curation support |
| **$\text{PEPPER}_{\text{XGB}}$** | Gene-level biological features (XGBoost model) | Predicted clinical significance (literature-independent) | Bayesian prior; Prospective gene discovery when compared with $\text{PEPPER}_{\text{LLM}}$ |
| **$\text{OMELET}_{\text{LLM}}$** | $\text{PEPPER}_{\text{LLM}}$ (prior) + LOEUF-MIS (likelihood) | Bayesian posterior clinical impact | Curation support |

| | | | |
|---|---|---|---|
| **$OMELET_{XGB}$** | $PEPPER_{XGB}$ (prior) + LOEUF-MIS (likelihood) | Bayesian posterior clinical impact (unbiased) | Disease gene prioritization, variant classification |
| **DisPo** | $PEPPER_{LLM\text{-}LoF}$ vs. gnomAD LoF constraint | Constraint–literature discordance | Prospective gene discovery, Identifying under-characterized disease genes |

### Practical Guidance: Selecting the Appropriate Metric

The metrics described above address distinct but complementary questions. Below, we provide guidance on which metric to use depending on the analytical goal.

#### Assessing loss-of-function constraint

When the objective is specifically to quantify a gene's intolerance to loss-of-function variation—for instance, to evaluate whether haploinsufficiency is likely deleterious—LOEUF should be used. Because LOEUF is computed exclusively from pLoF variants, it provides a direct measure of selective constraint against gene disruption without conflating loss-of-function and missense effects.

#### Assessing broad constraint against high-impact coding variants

When a more general measure of constraint is desired—one that captures the overall intolerance of a gene to all high-impact coding variants regardless of mechanism, including both pLoF and highly deleterious missense variants—LOEUF-MIS is preferable. LOEUF-MIS is particularly advantageous for short genes with few expected pLoF variants, and for genes in which diseases operate through gain-of-function (GoF) or dominant-negative (DN) mechanisms, where missense variants may be under stronger purifying selection than pLoF variants. However, users should be aware that LOEUF-MIS conflates LoF and missense constraint signals and should not be used when the goal is to isolate LoF-specific effects.

#### Assessing the clinical characterization of a gene

To determine the extent to which a gene's role in disease has been documented in the biomedical literature, $PEPPER_{LLM}$ provides a comprehensive, automated summary of published clinical evidence. It is useful for identifying well-characterized genes and for curating gene–disease associations at scale.

#### Predicting high clinical significance genes

When the goal is to predict which genes are likely to be clinically significant independently of existing literature, $PEPPER_{XGB}$ and $OMELET_{XGB}$ are recommended. Both scores are substantially less susceptible to literature bias than scores derived directly from published evidence, as demonstrated in the main text, and avoid simply recapitulating the publication history of well-studied genes. $PEPPER_{XGB}$ provides a standalone prediction based on biological features, while $OMELET_{XGB}$ further integrates population-level constraint from LOEUF-MIS into a Bayesian framework. $OMELET_{XGB}$ is the single best-performing score for unbiased disease gene prediction (AUPRC = 0.504 on the NDD benchmark, significantly outperforming either component alone), making it the recommended metric when the objective is to identify genes of high clinical significance without relying on prior literature characterization. For applications focused specifically on loss-of-function–mediated disease, an analogous score combining LOEUF with a LoF-specific version of $PEPPER_{XGB}$ could be constructed to restrict the framework to LoF signals only.

#### Identifying understudied disease gene candidates

This study presents two independent and complementary approaches for identifying genes whose disease relevance is likely under-characterized: (i) the discordance between $PEPPER_{XGB}$ and $PEPPER_{LLM}$ (percentile $PEPPER_{XGB}$ - percentile $PEPPER_{LLM}$, termed in this section $\Delta_{PEPPER}$), and (ii) the DisPo score. Although both aim to highlight understudied disease gene candidates, they answer different questions and may therefore flag different sets of genes.

The first approach leverages $PEPPER_{XGB}$, which predicts clinical significance from biological features independently of the literature. Genes for which $PEPPER_{XGB}$ substantially exceeds $PEPPER_{LLM}$ are likely under-characterized relative to their biological properties, making this discordance particularly suited for prospective gene discovery and for nominating candidates for functional follow-up. A practical advantage of using an XGBoost model is that SHAP (SHapley Additive exPlanations) analysis can be applied to identify which biological features drive a gene's high predicted clinical significance—for example, high expression in the brain or strong missense constraint. These feature-level explanations can provide mechanistic hints about the nature of the undiscovered gene–disease association and guide the design of targeted functional studies.

The second approach, DisPo, addresses a related but distinct question: which genes show strong population constraint against loss-of-function variants that is not accounted for by known clinical significance? While $\Delta_{PEPPER}$ highlights genes whose biological properties suggest clinical importance despite a sparse literature, DisPo specifically measures the distance between observed purifying selection in gnomAD and documented clinical impact. High-DisPo genes are candidates for harbouring undiscovered disease associations in phenotypic categories that are under-ascertained in current clinical databases, such as embryonic lethality, infertility, and other conditions with reduced reproductive fitness.

Despite being constructed from different signals—biological feature predictions versus population-level constraint—the two approaches are strongly correlated (Pearson $r = 0.500$, Spearman $\rho = 0.501$, both $p < 2.2 \times 10^{-308}$; $R^2 = 0.250$; Supplementary Figure 29a). This correlation reflects the fact that both metrics ultimately capture aspects of the same underlying biology: genes that are predicted to be clinically important by their biological features also tend to be under strong purifying selection relative to their literature characterization. The density heatmap (Supplementary Figure 29b) confirms that the highest concentration of genes falls along the positive diagonal, with a particularly dense cluster in the upper-right quadrant corresponding to genes ranked highly by both approaches. At the top 5% threshold for both metrics, 239 genes are jointly flagged; at the top 10%, 610 genes overlap (Supplementary Figure 29a).

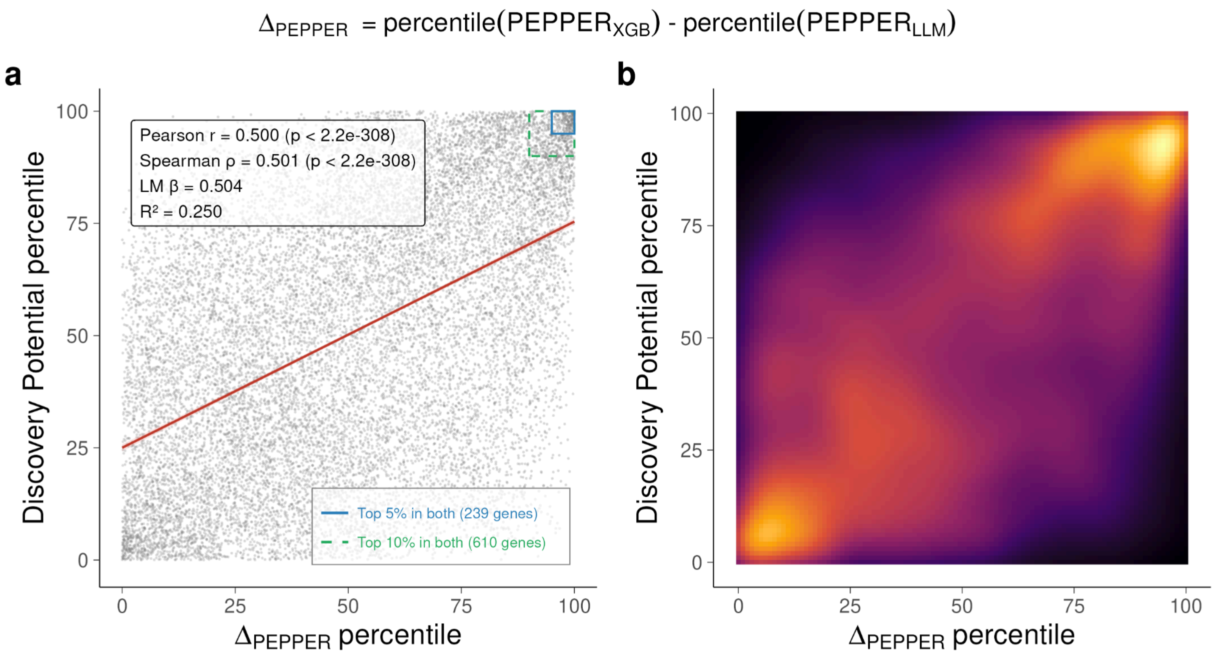


**Supplementary Figure 29 | DisPo compared to $\Delta_{PEPPER}$**

Nevertheless, substantial dispersion around the regression line ($R^2 = 0.250$) indicates that each approach captures complementary information that the other misses. Importantly, DisPo is focused exclusively on LoF constraint and therefore cannot detect undiscovered GoF or DN disease mechanisms, whereas $\Delta_{PEPPER}$ should capture under-characterization regardless of the mechanism. Users interested in identifying understudied disease gene candidates may therefore benefit from considering both approaches jointly: genes flagged by both methods represent especially strong candidates, while those flagged by only one may point to distinct biological scenarios worthy of investigation.

It should be noted that DisPo rests on a more principled statistical foundation, as it accounts for the variance of the underlying distributions and compares literature evidence against LOEUF, a well-established and extensively characterized constraint metric. In contrast, $\Delta_{PEPPER}$ compares literature to $PEPPER_{XGB}$, whose predictions are inherently dependent on the choice and availability of training features, introducing an additional layer of model-specific assumptions.

### References

1. Bycroft, C. *et al.* The UK Biobank resource with deep phenotyping and genomic data. *Nature* **562**, 203–209 (2018).
2. Karczewski, K. J. *et al.* The mutational constraint spectrum quantified from variation in 141,456 humans. *Nature* **581**, 434–443 (2020).
3. Chen, S. *et al.* A genomic mutational constraint map using variation in 76,156 human genomes. *Nature* (2023) doi:10.1038/s41586-023-06045-0.
4. Poterba, T. *et al.* The scalable variant call representation: enabling genetic analysis beyond one million genomes. *Bioinformatics* **41**, (2024).
5. Lu, W. *et al.* CHARR efficiently estimates contamination from DNA sequencing data. *Am. J. Hum. Genet.* **110**, 2068–2076 (2023).
6. McInnes, L., Healy, J. & Astels, S. hdbscan: Hierarchical density based clustering. *J. Open Source Softw.* **2**, 205 (2017).
7. Karczewski, K. J. *et al.* Systematic single-variant and gene-based association testing of thousands of phenotypes in 394,841 UK Biobank exomes. *Cell Genom.* **2**, 100168 (2022).
8. Bergström, A. *et al.* Insights into human genetic variation and population history from 929 diverse genomes. *Science* **367**, eaay5012 (2020).
9. 1000 Genomes Project Consortium *et al.* A global reference for human genetic variation. *Nature* **526**, 68–74 (2015).
10. Kaplanis, J. *et al.* Evidence for 28 genetic disorders discovered by combining healthcare and research data. *Nature* **586**, 757–762 (2020).
11. Cummings, B. B. *et al.* Transcript expression-aware annotation improves rare variant interpretation. *Nature* **581**, 452–458 (2020).
12. GTEx Consortium *et al.* Genetic effects on gene expression across human tissues. *Nature* **550**, 204–213 (2017).

13. Schraiber, J. G., Spence, J. P. & Edge, M. D. Estimation of demography and mutation rates from one million haploid genomes. *Am. J. Hum. Genet.* **112**, 2152–2166 (2025).
14. Weghorn, D. *et al.* Applicability of the mutation-selection balance model to population genetics of heterozygous protein-truncating variants in humans. *Mol. Biol. Evol.* **36**, 1701–1710 (2019).
15. Zeng, T., Spence, J. P., Mostafavi, H. & Pritchard, J. K. Bayesian estimation of gene constraint from an evolutionary model with gene features. *Nat. Genet.* **56**, 1632–1643 (2024).
16. Nei, M. The frequency distribution of lethal chromosomes in finite populations. *Proc. Natl. Acad. Sci. U. S. A.* **60**, 517–524 (1968).
17. Singer-Berk, M. *et al.* Advanced variant classification framework reduces the false positive rate of predicted loss-of-function variants in population sequencing data. *Am. J. Hum. Genet.* **110**, 1496–1508 (2023).
18. Seplyarskiy, V. *et al.* A mutation rate model at the basepair resolution identifies the mutagenic effect of polymerase III transcription. *Nat. Genet.* **55**, 2235–2242 (2023).
19. Nagy, E. & Maquat, L. E. A rule for termination-codon position within intron-containing genes: when nonsense affects RNA abundance. *Trends Biochem. Sci.* **23**, 198–199 (1998).
20. Abramowicz, A. & Gos, M. Splicing mutations in human genetic disorders: examples, detection, and confirmation. *J. Appl. Genet.* **59**, 253–268 (2018).
21. Vembar, S. S. & Brodsky, J. L. One step at a time: endoplasmic reticulum-associated degradation. *Nat. Rev. Mol. Cell Biol.* **9**, 944–957 (2008).
22. Boettcher, S. *et al.* A dominant-negative effect drives selection of TP53 missense mutations in myeloid malignancies. *Science* **365**, 599–604 (2019).
23. Basel, D. & Steiner, R. D. Osteogenesis imperfecta: recent findings shed new light on this once well-understood condition. *Genet. Med.* **11**, 375–385 (2009).
24. Lindeboom, R. G. H., Vermeulen, M., Lehner, B. & Supek, F. The impact of

nonsense-mediated mRNA decay on genetic disease, gene editing and cancer immunotherapy. *Nat. Genet.* **51**, 1645–1651 (2019).

25. Rehm, H. L. *et al.* ClinGen--the clinical genome resource. *N. Engl. J. Med.* **372**, 2235–2242 (2015).
26. Gudmundsson, S. *et al.* Exploring penetrance of clinically relevant variants in over 800,000 humans from the Genome Aggregation Database. *Nature Communications* **16**, 9623 (2025).
27. Kingdom, R., Beaumont, R. N., Wood, A. R., Weedon, M. N. & Wright, C. F. Genetic modifiers of rare variants in monogenic developmental disorder loci. *Nat. Genet.* **56**, 861–868 (2024).
28. Thormann, A. *et al.* Flexible and scalable diagnostic filtering of genomic variants using G2P with Ensembl VEP. *Nat. Commun.* **10**, 2373 (2019).
29. Berg, J. S. *et al.* An informatics approach to analyzing the incidentalome. *Genet. Med.* **15**, 36–44 (2013).
30. Blekhman, R. *et al.* Natural selection on genes that underlie human disease susceptibility. *Curr. Biol.* **18**, 883–889 (2008).
31. Mainland, J. D., Li, Y. R., Zhou, T., Liu, W. L. L. & Matsunami, H. Human olfactory receptor responses to odorants. *Sci. Data* **2**, 150002 (2015).
32. Lambert, S. A. *et al.* The human transcription factors. *Cell* **172**, 650–665 (2018).
33. UniProt Consortium, T. UniProt: the universal protein knowledgebase. *Nucleic Acids Res.* **46**, 2699 (2018).
34. Manning, G., Whyte, D. B., Martinez, R., Hunter, T. & Sudarsanam, S. The protein kinase complement of the human genome. *Science* **298**, 1912–1934 (2002).
35. Miranda-Saavedra, D. & Barton, G. J. Classification and functional annotation of eukaryotic protein kinases. *Proteins* **68**, 893–914 (2007).
36. Hunter, T. Signaling--2000 and beyond. *Cell* **100**, 113–127 (2000).

37. Binns, D. *et al.* QuickGO: a web-based tool for Gene Ontology searching. *Bioinformatics* **25**, 3045–3046 (2009).

38. Ashburner, M. *et al.* Gene ontology: tool for the unification of biology. The Gene Ontology Consortium. *Nat. Genet.* **25**, 25–29 (2000).

39. Gene Ontology Consortium. The Gene Ontology knowledgebase in 2026. *Nucleic Acids Res.* **54**, D1779–D1792 (2026).

40. Seal, R. L. *et al.* Genenames.Org: The HGNC and PGNC resources in 2026. *Nucleic Acids Res.* **54**, D1098–D1107 (2026).

41. Chakravarty, D. *et al.* OncoKB: A precision oncology knowledge base. *JCO Precis. Oncol.* **2017**, (2017).

42. Suehnholz, S. P. *et al.* Quantifying the expanding landscape of clinical actionability for patients with cancer. *Cancer Discov.* **14**, 49–65 (2024).

43. Vogelstein, B. *et al.* Cancer genome landscapes. *Science* **339**, 1546–1558 (2013).

44. Sevim Bayrak, C. *et al.* Identification of discriminative gene-level and protein-level features associated with pathogenic gain-of-function and loss-of-function variants. *Am. J. Hum. Genet.* **108**, 2301–2318 (2021).

45. Amberger, J. S., Bocchini, C. A., Schiettecatte, F., Scott, A. F. & Hamosh, A. OMIM.org: Online Mendelian Inheritance in Man (OMIM®), an online catalog of human genes and genetic disorders. *Nucleic Acids Res.* **43**, D789–98 (2015).

46. Meier, J. *et al.* Language models enable zero-shot prediction of the effects of mutations on protein function. *bioRxiv* 29287–29303 (2021) doi:10.1101/2021.07.09.450648.

47. Cheng, J. *et al.* Accurate proteome-wide missense variant effect prediction with AlphaMissense. *Science* **381**, eadg7492 (2023).

48. Orenbuch, R. *et al.* Proteome-wide model for human disease genetics. *Nat. Genet.* **57**, 3165–3174 (2025).

49. Frazer, J. *et al.* Disease variant prediction with deep generative models of evolutionary

data. *Nature* **599**, 91–95 (2021).

50. Chao, K. R. *et al.* The landscape of regional missense mutational intolerance quantified from 125,748 exomes. *bioRxiv* (2024) doi:10.1101/2024.04.11.588920.
51. Zhao, Y. *et al.* A probabilistic graphical model for estimating selection coefficients of nonsynonymous variants from human population sequence data. *Nat. Commun.* **16**, 4670 (2025).
52. Blaabjerg, L. M. *et al.* Rapid protein stability prediction using deep learning representations. *Elife* **12**, e82593 (2023).
53. Henikoff, S. & Henikoff, J. G. Amino acid substitution matrices from protein blocks. *Proc Natl Acad Sci U S A* **89**, 10915–10919 (1992).
54. Adzhubei, I. A. *et al.* A method and server for predicting damaging missense mutations. *Nat Methods* **7**, 248–249 (2010).
55. Grau, J., Grosse, I. & Keilwagen, J. PRROC: computing and visualizing precision-recall and receiver operating characteristic curves in R. *Bioinformatics* **31**, 2595–2597 (2015).
56. DiStefano, M. T. *et al.* The Gene Curation Coalition: A global effort to harmonize gene-disease evidence resources. *Genet Med* **24**, 1732–1742 (2022).
57. Chen, T. & Guestrin, C. XGBoost: A Scalable Tree Boosting System. *arXiv [cs.LG]* (2016).
58. Huang, Y.-F. Unified inference of missense variant effects and gene constraints in the human genome. *PLoS Genet.* **16**, e1008922 (2020).
59. Qi, Z., Luan, J., Zhou, X., Cui, Y. & Han, J. Fibrodysplasia ossificans progressiva: Basic understanding and experimental models. *Intractable Rare Dis. Res.* **6**, 242–248 (2017).
60. Aykul, S. *et al.* Activin A forms a non-signaling complex with ACVR1 and type II Activin/BMP receptors via its finger 2 tip loop. *Elife* **9**, (2020).
61. Hatsell, S. J. *et al.* ACVR1R206H receptor mutation causes fibrodysplasia ossificans progressiva by imparting responsiveness to activin A. *Sci. Transl. Med.* **7**, 303ra137 (2015).
62. Murthy, H. *et al.* Variants in DENND2B are associated with vulnerability for

neurodevelopmental impairment, psychosis and catatonia. *Brain* **149**, 252–261 (2026).

63. Cao, J. *et al.* A human cell atlas of fetal gene expression. *Science* **370**, eaba7721 (2020).
